# Global multi-ancestry genetic study elucidates genes and biological pathways associated with thyroid cancer and benign thyroid diseases

**DOI:** 10.1101/2025.05.15.25327513

**Authors:** Samantha L. White, Maizy S. Brasher, Jack Pattee, Wei Zhou, Sinéad Chapman, Yon Ho Jee, Caitlin C. Bell, Taylor L. Jamil, Martin Barrio, Jibril Hirbo, Nancy J. Cox, Peter Straub, Shinichi Namba, Emily Bertucci-Richter, Lindsay Guare, Ahmed EdrisMohammed, Sam Morris, Ashley J Mulford, Haoyu Zhang, Brian Fennessy, Martin D Tobin, Jing Chen, Alexander T Williams, Catherine John, David A van Heel, Rohini Mathur, Sarah Finer, Marta Riise Moksnes, Ben Brumpton, Bjørn Olav Åsvold, Raitis Peculis, Vita Rovite, Ilze Konrade, Ying Wang, Kristy Crooks, Sameer Chavan, Matthew J. Fisher, Nicholas Rafaels, Meng Lin, Jonathan Shortt, Alan R Sanders, David Whiteman, Stuart MacGregor, Sarah Medland, Unnur Thorsteinsdóttir, Kári Stefánsson, Tugce Karaderi, Kathleen M. Egan, Therese Bocklage, Hilary C McCrary, Greg Riedlingeer, Bodour Salhia, Craig Shriver, Minh D Phan, Janice L. Farlow, Stephen Edge, Varinder Kaur, Michelle Churchman, Robert J. Rounbehler, Pamela L. Brock, Matthew D. Ringel, Milton Pividori, Rebecca Schweppe, Christopher D. Raeburn, Robin Walters, Zhengming Chen, Liming Li, Koichi Matsuda, Yukinori Okada, Sebastian Zoellner, Anurag Verma, Michael Preuss, Eimear Kenny, Audrey Hendricks, Lauren Fishbein, Peter Kraft, Mark Daly, Benjamin Neale, The biobank at the Colorado Center for Personalized Medicine, Genes & Health Research Team, The BioBank Japan Project, Alicia Martin, Joanne B. Cole, Bryan R. Haugen, Christopher R. Gignoux, Nikita Pozdeyev

**Affiliations:** Department of Biomedical Informatics, University of Colorado Anschutz Medical Campus, Aurora, CO, USA; Center for Innovative Design & Analysis, Colorado School of Public Health, University of Colorado Anschutz Medical Campus, Aurora, CO, USA; Program in Medical and Population Genetics, Broad Institute of Harvard and MIT, Cambridge, MA, USA, Stanley Center for Psychiatric Research, Broad Institute of Harvard and MIT, Cambridge, MA, USA, Analytic and Translational Genetics Unit, Massachusetts General Hospital, Boston, MA, USA; The Broad Institute, Cambridge, MA, USA; Department of Epidemiology, Harvard T.H. Chan School of Public Health, Boston, MA, USA; Division of Endocrinology, Diabetes and Metabolism, University of Colorado Anschutz Medical Campus, Aurora, CO, USA; Department of Otolaryngology, Head and Neck Surgery, University of Colorado Anschutz Medical Campus, Aurora, Colorado, USA; Division of GI, Trauma, and Endocrine Surgery, Department of Surgery, University of Colorado School of Medicine, Aurora, CO, USA; Division of Genetic Medicine, Department of Medicine, Vanderbilt University Medical Center, Nashville, TN, USA, Vanderbilt Genetic Institute, Vanderbilt University Medical Center, Nashville, TN, USA; Department of Genome Informatics, Graduate School of Medicine, The University of Tokyo, Tokyo, Japan, Department of Statistical Genetics, Osaka University Graduate School of Medicine, Suita, Japan, Laboratory for Systems Genetics, RIKEN Center for Integrative Medical Sciences, Yokohama, Japan; Department of Biostatistics, University of Michigan, Ann Arbor, MI, USA; Department of Medicine, Perelman School of Medicine, University of Pennsylvania, Philadelphia, PA, USA; Nuffield Department of Population Health, University of Oxford, Oxford, UK; Genomic Health Initiative, Endeavor Health Research Institute, Evanston, IL, USA; The Charles Bronfman Institute for Personalized Medicine, Icahn School of Medicine at Mount Sinai, New York, NY, USA; Department of Population Health Sciences, University of Leicester, Leicester, UK University Hospitals of Leicester NHS Trust, Leicester, LE15WW, UK; Department of Population Health Sciences, University of Leicester, Leicester, UK; Blizard Institute, Queen Mary University of London, London E1 2AB, UK; Wolfson Institute of Population Health, Queen Mary University of London, London E1 2AB, UK; HUNT Research Centre, Department of Public Health and Nursing, NTNU, Norwegian University of Science and Technology, Levanger, Norway; Latvian Biomedical Research and Study Centre, Ratsupites 1-1, Riga, LV-1067, Latvia; Department of Internal Medicine, Riga Stradins University, Dzirciema Str 16, LV-1007 Riga, Latvia; Program in Medical and Population Genetics, Broad Institute of MIT and Harvard, Cambridge, MA, USA; Colorado Center for Personalized Medicine, University of Colorado Anschutz Medical Campus, Aurora, CO, USA; Department of Biomedical Informatics, University of Colorado Anschutz Medical Campus, Aurora, CO, USA, Colorado Center for Personalized Medicine, University of Colorado Anschutz Medical Campus, Aurora, CO, USA; Genomic Health Initiative, Endeavor Health Research Institute, Evanston, IL, USA, Department of Psychiatry and Behavioral Neuroscience, University of Chicago, Chicago, IL, USA; QIMR Berghofer Medical Research Institute: Herston, QLD, AU; deCODE genetics/Amgen, Inc., Reykjavik, Iceland, Faculty of Medicine, University of Iceland, Reykjavik, Iceland; deCODE genetics/Amgen, Inc., Reykjavik, Iceland; Faculty of Medicine, University of Iceland, Reykjavik, Iceland; Center for Health Data Science, Section for Health Data Science and Artificial Intelligence, Department of Public Health, Faculty of Health and Medical Sciences, University of Copenhagen, Copenhagen, Denmark; Moffitt Cancer Center, Department of Cancer Epidemiology, H. Lee Moffitt Cancer Center & Research Institute, Tampa, FL, USA; University of Kentucky Markey Cancer Center, University of Kentucky-Department of Pathology and Laboratory Medicine, University of Kentucky-Chandler Medical Center, Lexington, KY, USA; University of Utah Huntsman Cancer Institute, Department of Otolaryngology-Head and Neck Surgery, School of Medicine, Huntsman Cancer Institute, University of Utah, Salt Lake City, UT, USA; Rutgers Cancer Institute of New Jersey Department of Medicine, Division of Medical Oncology, Rutgers Cancer Institute of New Jersey, New Brunswick, NJ, USA; Norris Comprehensive Cancer Center and Keck School of Medicine of University of Southern California, Department of Translational Genomics, Keck School of Medicine, University of Southern California, Los Angeles, California, USA; Muthra Cancer Center, Murtha Cancer Center, Uniformed Services University/Walter Reed National Military Medical Center, Bethesda, MD, USA; University of Oklahoma Stephenson Cancer Center, Medical Oncology/Head and Neck Oncology, Stephenson Cancer Center, University of Oklahoma, Oklahoma City, Oklahoma, USA; Indiana University School of Medicine, Indianapolis, Indiana; Roswell Park Comprehensive Cancer Center, Departments of Surgical Oncology and Cancer Prevention and Control, Roswell Park Comprehensive Cancer Center, Buffalo, NY, USA; University of Virginia Cancer Center, Department of Internal Medicine, Division of Hematology & Oncology, University of Virginia Health, Charlottesville, VA, USA; Aster Insights, Hudson, FL, USA; The Ohio State University Comprehensive Cancer Center and the Ohio State University College of Medicine, Columbus, OH, USA; Division of Endocrinology, Diabetes and Metabolism, University of Colorado Anschutz Medical Campus, Aurora, CO, USA, University of Colorado Cancer Center, University of Colorado Anschutz Medical Campus, Aurora, CO, USA; Department of Epidemiology and Biostatistics, School of Public Health, Peking University, Beijing, China, Peking University Center for Public Health and Epidemic Preparedness and Response, Beijing, China, Key Laboratory of Epidemiology of Major Diseases (Peking University), Ministry of Education, Beijing 100191, China; Laboratory of Genome Technology, Human Genome Center, Institute of Medical Science, The University of Tokyo, Tokyo, Japan, Laboratory of Clinical Genome Sequencing, Graduate School of Frontier Sciences, The University of Tokyo, Tokyo, Japan; Division of Endocrinology, Diabetes and Metabolism, University of Colorado Anschutz Medical Campus, Aurora, CO, USA, Department of Biomedical Informatics, University of Colorado Anschutz Medical Campus, Aurora, CO, USA, University of Colorado Cancer Center, University of Colorado Anschutz Medical Campus, Aurora, CO, USA, Research Service, Rocky Mountain Regional VA Medical Center, Aurora, CO, USA; Department of Epidemiology, Harvard T.H. Chan School of Public Health, Boston, MA, USA, Transdivisional Research Program, Division of Cancer Epidemiology and Genetics, National Cancer Institute, National Institutes of Health, MD, USA; Program in Medical and Population Genetics, Broad Institute of MIT and Harvard, Cambridge, MA, USA, Analytical and Translational Genetics Unit, Department of Medicine, Massachusetts General Hospital and Harvard Medical School, Boston, MA, USA, Stanley Center for Psychiatric Research, Broad Institute of MIT and Harvard, Cambridge, MA, USA; Program in Medical and Population Genetics, Broad Institute of MIT and Harvard, Cambridge, MA, USA, Analytical and Translational Genetics Unit, Department of Medicine, Massachusetts General Hospital and Harvard Medical School, Boston, MA, USA, Stanley Center for Psychiatric Research, Broad Institute of MIT and Harvard, Cambridge, MA, USA, University of Colorado Anschutz Medical Campus, Aurora, CO, USA; Analytic and Translational Genetics Unit, Massachusetts General Hospital, Boston, MA, USA, Stanley Center for Psychiatric Research and Program in Medical and Population Genetics, Broad Institute of MIT and Harvard, Cambridge, MA, USA, Department of Medicine, Harvard Medical School, Boston, MA, USA; Department of Biomedical Informatics, University of Colorado Anschutz Medical Campus, Aurora, CO, USA, Human Medical Genetics and Genomics Program, University of Colorado Anschutz Medical Campus, Aurora, CO, USA; Human Medical Genetics and Genomics Program, University of Colorado Anschutz Medical Campus, Aurora, CO, USA, Department of Biomedical Informatics, University of Colorado Anschutz Medical Campus, Aurora, CO, USA, Colorado Center for Personalized Medicine, University of Colorado Anschutz Medical Campus, Aurora, CO, USA, University of Colorado Cancer Center, University of Colorado Anschutz Medical Campus, Aurora, CO, USA; Department of Biomedical Informatics, University of Colorado Anschutz Medical Campus, Aurora, CO, USA, Division of Endocrinology, Diabetes and Metabolism, University of Colorado Anschutz Medical Campus, Aurora, CO, USA, University of Colorado Cancer Center, University of Colorado Anschutz Medical Campus, Aurora, CO, USA, Colorado Center for Personalized Medicine, University of Colorado Anschutz Medical Campus, Aurora, CO, USA

## Abstract

Thyroid diseases are common and highly heritable. Under the Global Biobank Meta-analysis Initiative, we performed a meta-analysis of genome-wide association studies from 19 biobanks for five thyroid diseases: thyroid cancer, benign nodular goiter, Graves’ disease, lymphocytic thyroiditis, and primary hypothyroidism. We analyzed genetic association data from ∼2.9 million genomes and identified 235 known and 501 novel independent variants significantly linked to thyroid diseases. We discovered genetic correlations between thyroid cancer, benign nodular goiter, and autoimmune thyroid diseases (*r*^2^=0.21-0.97). Telomere maintenance genes contribute to benign and malignant thyroid nodular disease risk, whereas cell cycle, DNA repair, and DNA damage response genes are predominantly associated with thyroid cancer. We proposed a paradigm explaining genetic predisposition to benign and malignant thyroid nodules. We evaluated thyroid cancer polygenic risk scores (PRS) for clinical applications in thyroid cancer diagnosis. We found PRS associations with thyroid cancer risk features: multifocality, lymph node metastases, and extranodal extension.

---

Thyroid diseases are highly prevalent. According to the American Thyroid Association, over 12 percent of the US population will develop a thyroid condition during their lifetime (https://www.thyroid.org/media-main/press-room/). Thyroid cancer is the most common endocrine malignancy, with 44,020 new cases and 2,170 deaths in the United States in 2024 ^1^. Thyroid function diseases, hypo- and hyperthyroidism, negatively affect most organ systems and are associated with major excess in cardiovascular mortality ^2^. It is not well understood why some individuals develop thyroid disease, although genetic ^3,4^ and environmental factors (radiation exposure^5^) play a role.

Genetic effects are estimated to contribute up to 53% to thyroid cancer susceptibility in family studies ^3,4^, making thyroid cancer one of the most heritable common cancers^3,6^. For autoimmune thyroid diseases, genetic factors account for approximately 75% of the total phenotypic variance ^7^.

Ruling out thyroid malignancy is a common clinical task due to the high prevalence of thyroid nodules. Thyroid ultrasound reveals nodules in up to 65% of the general population, with prevalence increasing in women, with increasing age, and after radiation exposure ^8,9^. Clinical providers assess thyroid nodule sonographic characteristics ^10^ to decide if fine needle aspiration (FNA) biopsy is indicated. Over 600,000 fine needle aspiration thyroid nodule biopsies (FNA) are performed annually in the United States to rule out cancer ^11^, and most (∼92%) produce benign, inadequate, or indeterminate results ^12,13^. Genetic thyroid cancer risk assessment with polygenic risk score (PRS) provides an opportunity to improve the diagnostic yield of FNA and reduce unnecessary procedures, molecular tests and diagnostic surgeries ^14^.

In addition to the clinical applications, discovering genetic variants predisposing to thyroid cancer and benign thyroid conditions helps in understanding the biological processes leading to disease. Several genome-wide association studies (GWAS) have been conducted on thyroid cancer ^15–20^. More recently, the Global Biobank Meta-analysis (GBMI) consortium combined data from 6,699 individuals with thyroid cancer and ∼2.2 million controls and identified 27 significant genetic associations^21^. GWAS for benign thyroid diseases and related traits, such as thyroid-stimulating hormone (TSH) levels, have been performed in large biobanks: UK Biobank^22,23^, FinnGen ^24^, and Million Veteran Program ^25^. However, a systematic analysis of underlying genes, pathways, and clinical relevance is missing.

Platforms such as the Global Biobank Meta-analysis initiative (GBMI, https://www.globalbiobankmeta.org/ ^21^) enable global collaborations among dozens of participating biobanks, resulting in unmatched GWAS discovery power and data diversity, particularly relevant to cross-phenotype investigations. In this study, we report results from a GBMI project dedicated to thyroid diseases. We confirmed many previously reported variants and discovered hundreds of novel genetic associations with thyroid diseases. We found genes and pathways unique to malignant and benign nodular disease. We defined potential clinical applications for polygenic risk scores (PRS) for thyroid cancer to better distinguish thyroid cancer from benign nodules before FNA. We found a genetic predisposition to multifocal and metastatic thyroid cancer.

## Results

The study had three phases (**Figure 1**): 1) Variant Discovery: GWAS, meta-analysis, and quality control procedures; 2) Functional Inference: genetic correlations, transcriptome-wide association studies (TWAS), pathway and gene expression analyses; and 3) Clinical Studies: PRS development, testing on the clinical use case of distinguishing benign from malignant thyroid nodules, and associations with cancer aggressiveness.

**Figure 1.**
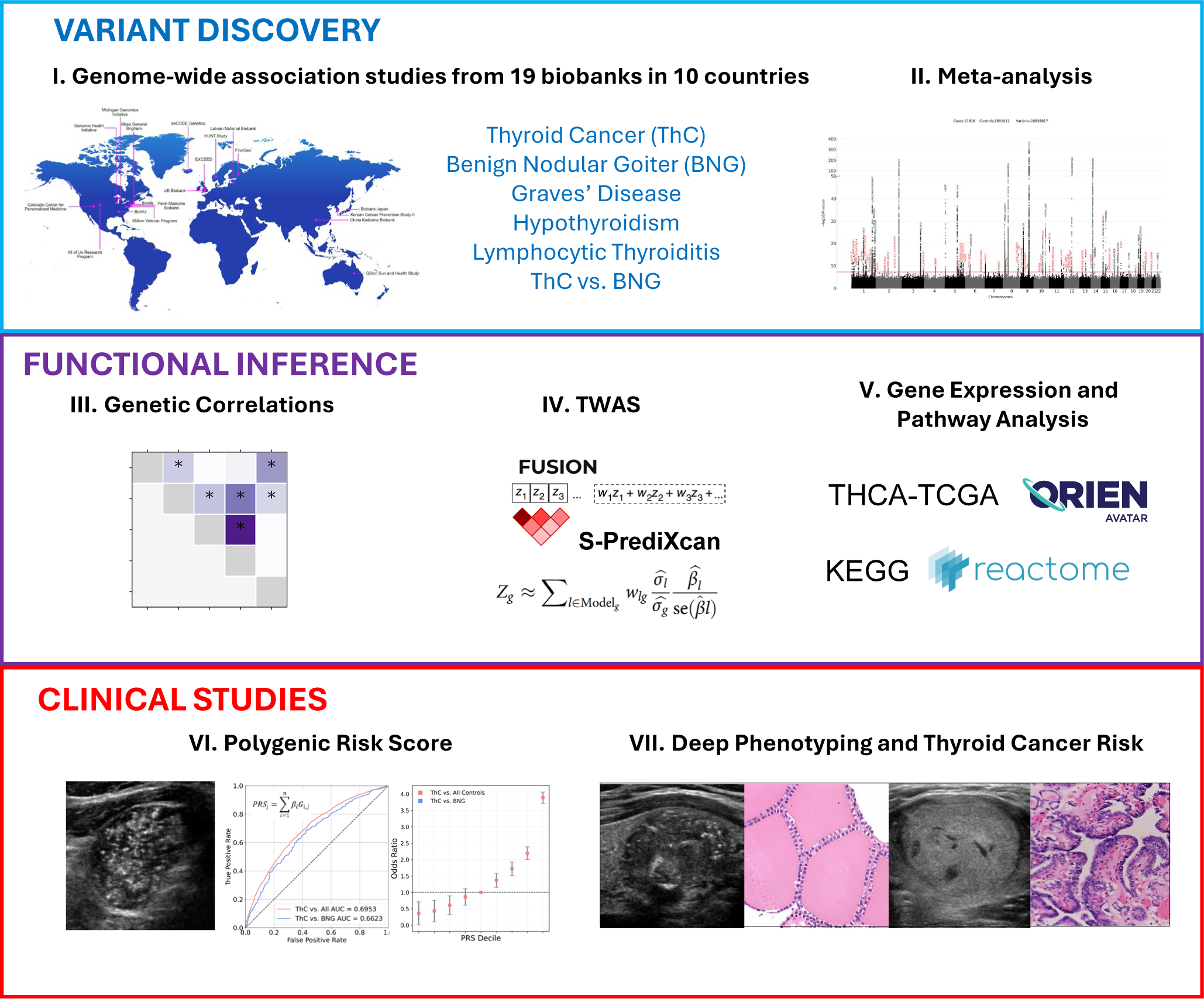
The study design. **I**. The Virtual Thyroid Biopsy Consortium was formed under the Global Biobank Meta-analysis Initiative. Participating biobanks performed genome-wide association studies (GWAS) for five thyroid diseases and GWAS of thyroid cancer vs. benign nodule goiters (ThC vs. BNG). **II.** Inverse variance-weighted meta-analysis was done after quality-control procedures. Previously known and novel independent genetic associations were identified. Functional inference studies included: **III**. Genetic correlation analysis with covariate-adjusted LD score regression. **IV. T**ranscriptome-wide association studies (FUSION and S-PrediXcan), **V.** Pathway (KEGG and Reactome), and gene-expression (The Cancer Genome Atlas and ORIEN Avatar) analyses. **VI.** Polygenic risk scores were developed for thyroid cancer, benign thyroid diseases, and to distinguish malignant and benign thyroid nodules. **VII.** Polygenic risk scores were tested for association with thyroid diseases and aggressive thyroid cancer features extracted from clinical charts and surgical histopathology reports.

### Virtual Thyroid Biopsy Consortium

We founded the Virtual Thyroid Biopsy Consortium (**Extended Data Figure 1**) under the Global Biobank Meta-analysis initiative (https://www.globalbiobankmeta.org/) ^21^ to study the genetic architecture of thyroid diseases at a global multi-ancestry scale. The Consortium aggregates data from 19 biobanks in 10 countries and four continents. (**Supplementary Table 1**). Biobanks performed multi-ancestry and/or ancestry-stratified GWAS for five thyroid diseases: thyroid cancer, benign nodular goiter, Graves’ disease, lymphocytic thyroiditis and primary hypothyroidism. In addition, a GWAS of thyroid cancer vs. benign nodular goiter was performed, focusing on the common clinical task of determining malignancy in thyroid nodules. Phenotype and GWAS definitions are listed in **Supplementary Tables 2 and 3**.

### Meta-analysis of genome-wide association studies for thyroid diseases

Our meta-analysis aggregated data from 198 GWAS summary data files (**Supplementary Table 4**). Individual GWAS runs were well controlled for confounding (covariate-adjusted LD score regression [cov-LDSC]^26^ Y-axis intercept 1.00 ± 0.05 [mean ± SD]). Healthcare system-based biobanks had a higher disease prevalence than population-based biobanks (**Extended Data Figure 2**), as reported previously ^21^. Bio*Me,* the All of Us Research Program (AoU), and the Million Veteran Program Biobanks had the most diverse participant pools as measured by Summix2 ^27^.

The meta-analysis included 21,816 cases of thyroid cancer, 68,987 cases of benign nodular goiter, 18,719 cases of Graves’ disease, 18,331 cases of lymphocytic thyroiditis, 257,365 cases of primary hypothyroidism, and ∼ 2.9 million controls (**Supplementary Table 3**). Population structure was determined with Summix2 ^27^ via mixture modeling of study-based allele frequencies compared to the gnomAD reference panel ^28^. Seventeen percent of genotypes were from individuals of African (AFR-like), 4.4 % from Admixed American (AMR-like), 8.1% from East Asian (EAS-like), and 70.5% from European (EUR-like) ancestry.

We found 736 independent (loci separated by at least 500 kb) variants significantly (p-value ≤ 5e-8) associated with thyroid diseases, including mixed-ancestry and ancestry-stratified genetic associations (**Table 1, Supplementary Tables 5.1-5.6**). Of these, 235 variants were reported to the NHGRI-EBI Catalog^29^ for thyroid traits (as of April 2024), and 501 genetic associations were novel. Most lead associations are in the introns of protein-coding genes (n = 356), followed by intergenic variants (n = 215 variants). Among 46 significant exonic variants, 43 are non-synonymous, potentially altering protein function.

**Table 1.** Independent genetic loci associated with thyroid diseases.

| Phenotype | Variant type, N |  | Variant location, N |  |  |  |  |  |  |  |  |  |
| --- | --- | --- | --- | --- | --- | --- | --- | --- | --- | --- | --- | --- |
|  | Known | Novel | Exonic | 3'UTR | 5'UTR | Upstream | Downstream | ncRNA<br>exonic | Intronic | ncRNA<br>intronic | Intergenic | Total |
| Thyroid cancer | 26 | 61 | 6 | 3 |  | 2 | 2 |  | 34 | 11 | 29 | 87 |
| Benign nodular goiter | 16 | 84 | 4 | 1 |  | 0 | 1 | 1 | 52 | 10 | 31 | 100 |
| Graves' disease | 30 | 70 | 6 | 1 |  |  | 4 |  | 52 | 6 | 31 | 100 |
| Hypothyroidism | 120 | 271 | 26 | 9 | 7 | 10 | 5 | 3 | 194 | 30 | 107 | 391 |
| Lymphocytic thyroiditis | 43 | 15 | 4 | 3 |  | 2 | 3 |  | 24 | 5 | 17 | 58 |
| Total | 235 | 501 | 46 | 17 | 7 | 14 | 15 | 4 | 356 | 62 | 215 | 736 |

Counts of variants (N) by type and location are shown. Variant type was determined based on genetic associations reported in the NHGRI- EBI GWAS catalog (April 2024, version 1.0.2). The variant location was extracted from the ANNOVAR annotation.

Ancestry-stratified GWAS (**Supplementary Tables 5.1-5.6**) replicated many associations from the combined mixed-ancestry meta-analysis with some exceptions. For example, a rare (minor allele frequency [MAF] = 0.0007) non-synonymous exonic variant in the shelterin complex gene *TERF1* (8:73046129:G:A, β=1.33, p-value = 8.8e-10) was significantly associated with thyroid cancer in the EUR-like meta-analysis only (mixed ancestry GWAS β=1.16, p-value = 5e-4).

### SNP heritability (h^2^_SNP_) and genetic correlation

Cov-LDSC estimated ℎ^2^ ranged from 0.067 (SE = 0.01) for benign nodular goiter in mixed-ancestry meta-analysis to 0.11 (0.026) for EUR-like thyroid cancer meta-analysis (**Supplementary Table 6**).

Genetic correlation was very high for lymphocytic thyroiditis and hypothyroidism (mixed-ancestry, *r^2^* = 0.97 [0.04], p-value = 7.9e-105, **Figure 2, Supplementary Table 7**). We found significant (Benjamini-Hochberg false discovery rate (FDR) <0.05) genetic correlations between lymphocytic thyroiditis and Graves’ disease (*r^2^* = 0.62 [0.07]), lymphocytic thyroiditis and benign nodular goiter (*r^2^* = 0.15 [0.07]), thyroid cancer and benign nodular goiter (*r^2^* = 0.40 [0.16]), Graves’ disease and hypothyroidism (*r^2^* = 0.36 [0.07]), Graves’ disease and benign nodular goiter (*r^2^* = 0.31 [0.07]), and Graves’ disease and thyroid cancer (*r^2^* = 0.21 [0.05]). Genetic correlation analysis in EUR-like meta-analysis showed similar results. (**Extended Data Figure 3, Supplementary Table 7**).

**Figure 2.**
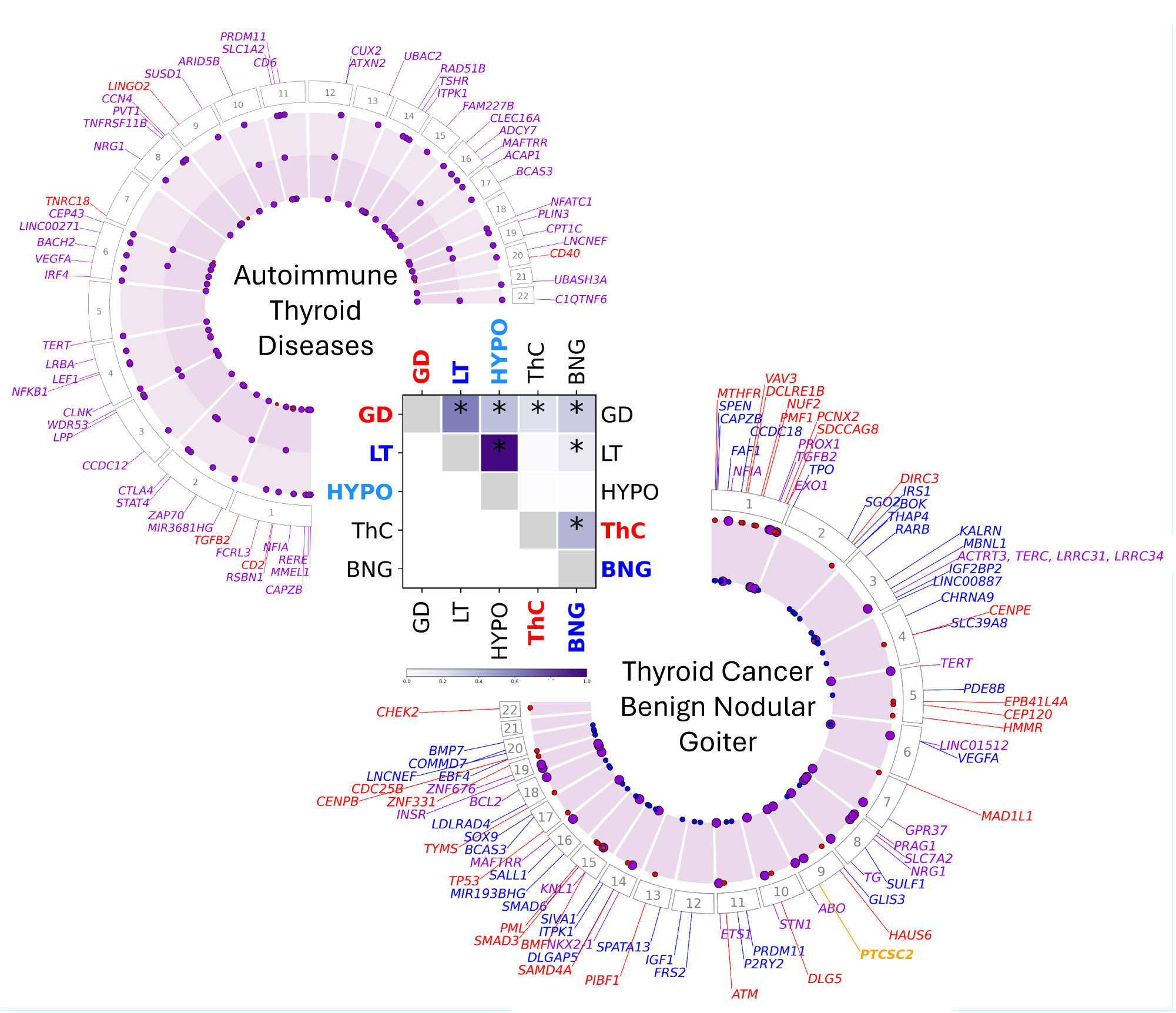
Pleiotropic and phenotype-specific loci associated with thyroid diseases in multi-ancestry GWAS meta-analysis. The heatmap illustrates the genetic correlation (*r^2^*) between thyroid phenotypes, which was estimated using covariate-adjusted LD score regression. The asterisks denote Benjamini-Hochberg false discovery rate (FDR) <0.05. Circular plots highlight loci significantly associated with thyroid cancer and benign nodular goiter (right), and autoimmune thyroid diseases (left). Pleiotropic loci are marked by large purple dots. In the circular plot on the right small dots show loci predominantly associated with thyroid cancer or benign nodular goiter (red or blue respectively). In the circular plot on the left the small red dots depict loci which are significantly associated with Graves’ disease, but not lymphocytic thyroiditis or primary hypothyroidism. For clarity, only loci significantly associated with Graves’ disease are shown on the autoimmune thyroid diseases circular plot. The *PTCSC2* (right, yellow) is the only locus inversely associated with thyroid cancer and benign nodular goiter.

### Transcriptome-wide association study (TWAS)

We performed *cis-*acting expression quantitative trait loci (*cis-*eQTLs) TWAS using two methods, FUSION^30^ and S-PrediXcan^31,32^ and GTEx v8 thyroid tissue expression models ^33^, to identify potential causal variants affecting gene expression and assign intergenic and non-coding RNA variants to protein-coding genes.

FUSION TWAS, as applied to the mixed-ancestry and EUR-like thyroid cancer GWAS meta-analysis, identified expression of 55 unique protein-coding genes (**Supplementary Tables 8 and 9**). FUSION also identified 47 and 45 significant (after Bonferroni adjustment) lead *cis*-eQTL variants from mixed ancestry and EUR-like GWAS, respectively. TWAS attributed many significant intergenic and non-coding variants to protein-coding genes based on reported eQTL status. For example, GWAS intergenic variant 1:218515813:T:C (mixed-ancestry thyroid cancer GWAS meta-analysis p-value 4.07e-39) was also a lead *cis*-eQTL variant in FUSION TWAS for *TGFB2* (p-value = 3.59e-61). Most significant genes found by FUSION TWAS were also replicated by S-PrediXcan, indicating the analytic rigor of our analyses (**Supplementary Tables 5.1-5.6**).

TWAS found additional significant genes where meta-analysis failed to identify genome-wide significant associations, e.g., *VEGFC* (p-value = 1.30e-06), *CDCA7L* (p-value = 6.02e-07), and *NBR1* (p-value = 1.02e-06), further expanding our knowledge of genes associated with thyroid cancer risk beyond those with variants with marginal significance.

### Gene expression analysis

We evaluated mRNA expression of genes discovered in the thyroid cancer GWAS meta-analysis and TWAS in normal and malignant thyroid tissues (**Extended Data Figure 4**, **Supplementary Table 10**). Of the twenty total evaluated tissue types ^34^, normal thyroid tissue was among the top three highest-expressing tissues for twenty-two genes. Two genes, *TG* and *PTCSC2*, are expressed only in the thyroid.

The expression of four genes (*VAV3*, *PCNX2*, *ETS1*, and *PIBF1*) correlated with younger age at thyroid cancer diagnosis in The Cancer Genome Atlas study for papillary thyroid cancer (THCA-TCGA)^35^ and/or the Oncology Research Information Exchange Network (ORIEN) AVATAR study (https://www.oriencancer.org/) (*r^2^* = −0.14 to −0.22, p-value ≤ 4.2e−05). *TERT* expression correlated with older age at diagnosis (*r^2^* = −0.17 to −0.21, p-value ≤ 7.4e−05), matching a similar association with somatic *TERT* promoter mutations ^36^. Overall, the expression of most significant genes (47 out of 54 with RNA sequencing data in THCA-TCGA) was correlated with at least one clinical or molecular thyroid cancer risk feature: higher stage, presence of extrathyroidal extension, *BRAF* V600E mutation, lower BRAF/RAS score (indicating BRAF-like expression profile^35^), higher ERK score (measuring RAS/MAPK pathway activity), and lower thyroid cancer differentiation (estimated with thyroid differentiation score ^35^) (**Extended Data Figure 4**, **Supplementary Table 10,** Bonferroni-corrected p-value ≤ 9.1e−05).

### Pleiotropic and disease-specific associations with thyroid cancer and benign nodular goiter

We do not know why some patients develop benign thyroid nodules, while others get thyroid cancer. To understand cellular functions and pathways leading to benign or malignant thyroid nodular disease, we explored pleiotropic and thyroid cancer- and benign nodular goiter-specific loci (**Figure 2**).

We generated locus plots for genes with significant GWAS meta-analysis associations and genes with significant *cis*-eQTL in TWAS (**Supplementary Figures 1.1-1.3**). We categorized loci as those significantly associated with: 1) thyroid cancer but not benign nodular goiter (these loci may contribute to malignant transformation of follicular cells, **Supplementary Table 11, Supplementary Figure 1.1**); 2) benign nodular goiter but not thyroid cancer (these loci may lead to non-neoplastic thyroid nodules and thyroid neoplasms with low malignant potential, **Supplementary Figure 1.2**); and 3) both benign and malignant thyroid nodules (**Supplementary Figure 1.3**). In addition, we identified loci that were strongly associated with thyroid cancer (p-value < 1e-20) but relatively weakly (p-value >1e-10) associated with benign nodular goiter (e.g., *HAUS6* and *SDCCAG*) despite greater discovery power of benign nodular goiter GWAS meta-analysis.

Among 28 genes associated predominantly with thyroid cancer, 12 encode components of cell cycle checkpoints and proteins regulating centrosome and kinetochore function, microtubule attachment, and chromosome segregation (*ATM, CDC25B, CENPB, CENPE, CEP120, CHEK2, HAUS6, HMMR, MAD1L1, NUF1, PMF1, SDCCAG8*). Five genes play a role in DNA repair and cellular response to DNA damage (*ATM, DCLRE1B, PCNX2, PML, TP53*).

Benign nodular goiter-specific genes (n = 38) participate in insulin-like growth factor 1 (*IGF1, IGF2BP2, and IRS1*) and fibroblast growth factor (*FGF7, FRS2*) signaling pathways. Genes participating in thyroid gland development and thyroid hormone synthesis were linked to benign nodules (*GLIS3, TPO*), but some are also associated with thyroid cancer (*NKX2-1, TG*).

Notably, significant loci in telomere maintenance genes (*ACTRT3, TERC, LRRC34, ETS1, EXO1, STN1, TERT*) were associated with both thyroid cancer and benign nodular goiter. Genes participating in apoptosis, senescence, and transforming growth factor β (TGFβ) signaling are present in all three gene categories (**Supplementary Table 11**) and contribute to the development of both benign and malignant thyroid nodules. Variants in some of these overlapping genes (e.g., *TERT,* 5:1280013:T:C, β = 0.11 [0.02], p-value = 1.71e-10; NRG1, 8:32571946:G:A, β = −0.18 [0.02], p-value = 9.89e-25) were also significant in our meta-analysis of thyroid cancer vs. benign nodular goiter GWAS (**Supplementary Table 5.6**), indicating differential contribution to these diseases. Of particular interest is the *PTCSC2* locus because its significant variants have the opposite direction of effect with thyroid cancer and benign nodular goiter (**Extended Data Figure 5**, ρ = −0.77, p-value = 1.2e-24).

KEGG^37^ and Reactome^38^ pathway analysis identified cell cycle, senescence and apoptosis as key biological processes contributing to thyroid cancer risk (**Supplementary Tables 12 and 13**). IGF1 and PI3K/AKT signaling pathways were significantly associated with benign nodular goiter. Consistent with our analysis of individual genes (**Supplementary Table 11, Supplementary Figure 1.3**) telomere-related pathways were found in both thyroid cancer and benign nodular goiter analyses.

### Pleiotropic and disease-specific associations with autoimmune thyroid diseases

Graves’ disease and lymphocytic thyroiditis/primary hypothyroidism are related autoimmune endocrine diseases with opposite clinical manifestations, causing hyperthyroidism and hypothyroidism, respectively ^39^.

Plausibly, most genes and KEGG and Reactome pathways associated with autoimmune thyroid diseases (**Supplementary Tables 12 and 13**) are related to the immune system. Almost all loci significantly associated with Graves’ disease (**Supplementary Figure 2.1**) are also linked to primary hypothyroidism. Four genes (*CD2, CD40, LINGO2, and TNRC18*) were discovered in Graves’ disease (p-value < 5e-8) but not hypothyroidism GWAS meta-analysis (**Supplementary Figure 2.2**). Genetic associations with lymphocytic thyroiditis (**Supplementary Table 5.4**) replicated those with primary hypothyroidism (**Figure 2**).

### Polygenic risk score (PRS) for thyroid cancer diagnosis

PRS quantifies an individual’s risk for developing a specific trait or disease based on their genetics. We explored the ability of PRS to identify people at high risk for thyroid cancer (PRS_ThC vs. All_) in the Colorado Center for Personalized Medicine (CCPM) Biobank population (n = 94,651). PRS_ThC vs. All_ was calculated from the independent significant variants from the multi-ancestry thyroid cancer meta-analysis, leaving CCPM out of training to avoid overfitting.

Population screening for thyroid cancer is controversial; therefore, we also assessed the utility of PRS for the clinically relevant task of distinguishing benign from malignant thyroid nodules (PRS_ThC vs. BNG_). PRS_ThC vs. BNG_ was defined as the difference between PRS_ThC vs. All_ and PRS for benign nodular goiter (PRS_BNG vs. All_): PRS_ThC vs. BNG_ = PRS _ThC vs. All_ - PRS_BNG vs. All_.

PRS_ThC vs. All_ achieved an area under the receiver operating characteristic curve (AUC) of 0.686 (95% CI, 0.669,0.703, **Supplementary Table 14**). Genome-wide PRS_ThC vs. All_ calculated using PRS-CS^40^ showed an AUC of 0.695 (0.669,0.721, **Figure 3A**). Individuals with PRS_ThC vs. All_ in the top decile have ∼10.7 times the odds to develop thyroid cancer than those in the first decile (**Figure 3B**).

**Figure 3.**
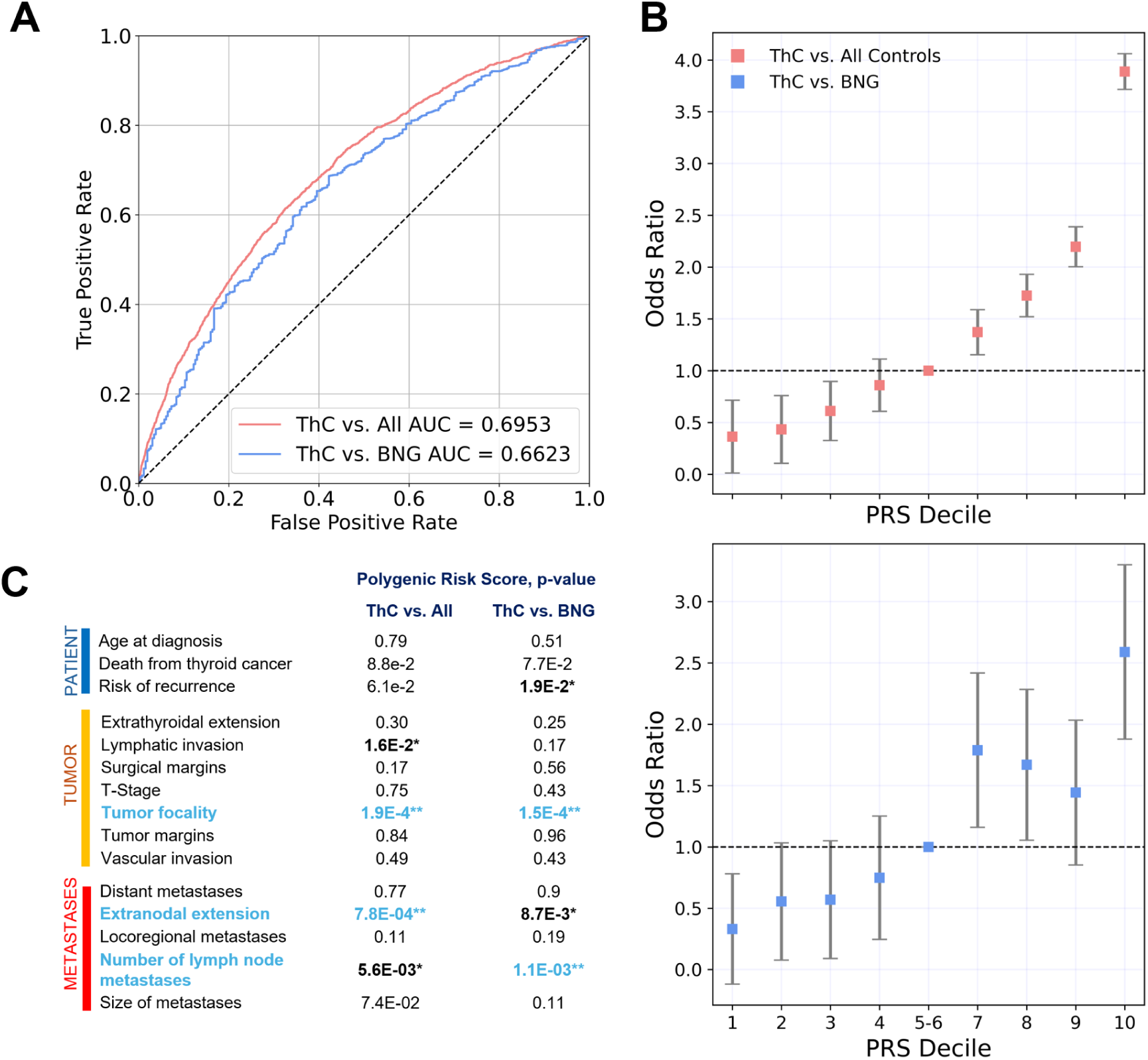
Thyroid cancer polygenic risk scores. Two thyroid cancer polygenic risk scores (PRS) were developed: PRS_ThC vs. All_ to identify individuals at risk in a population and PRS_ThC vs. BNG_ for clinically relevant task of discriminating malignant and benign thyroid nodules. PRS were tested in the Colorado Center for Personalized Medicine Biobank (CCPM) population (n = 94,651) and CCPM data was not used for PRS development. PRS_ThC vs. All_ was derived using PRS-CS method. PRS_ThC vs. BNG_ was calculated using independent significant associations with thyroid cancer and benign nodular goiter. **A.** Receiver operating characteristic curves. **B.** Thyroid cancer risk by PRS decile. **C.** PRS association with features of aggressive thyroid cancer. * - p-value ≤ 0.05. ** - p-value ≤ 1.7e-3 (Bonferroni-corrected significance threshold).

Our PRS_ThC vs. All_ significantly outperformed the thyroid cancer PRS derived from the largest previous GWAS meta-analysis from the GBMI phase I project^21^ (AUC 0.651 [0.632, 0.671], DeLong test p-value = 1.13e−07). This improvement highlights the greater discovery power of a large meta-analysis.

To test PRS performance on a clinically relevant use case of discriminating benign and malignant thyroid nodules (ThC vs. BNG), three clinicians (CB, TJ and NP) performed clinical chart reviews. We confirmed the diagnosis of non-medullary thyroid cancer in 1,344 patients and the diagnosis of benign nodular goiter in 281 (out-of-sample test set). All benign cases were supported by surgical histopathology to avoid contamination due to small thyroid cancers not eligible for biopsy.

PRS_ThC vs. All_ performed worse for the clinical ThC vs. BNG task (AUC 0.618 (0.561,0.675), which is expected due to the genetic associations shared between thyroid cancer and benign nodular goiter. PRS_ThC vs. BNG_, leveraging genetic associations with both thyroid cancer and benign nodular goiter, demonstrated an improved AUC for the ThC vs. BNG clinical task (0.664 [0.586,0.742], DeLong test p-value = 5e-4).

Thyroid nodules in individuals with PRS_ThC vs. BNG_ in the top decile had ∼7.8 times the odds to be malignant than in individuals with PRS_ThC vs. BNG_ in the first decile (**Figure 3B**).

For benign thyroid diseases, PRS AUCs ranged from 0.585 (0.572,0.599) for benign nodular goiter to 0.664 (0.638,0.69) for Graves’ disease. PRS analyses in the European population showed results similar to those from the mixed-ancestry GWAS meta-analysis (**Supplementary Table 14**).

Incorporating demographic and genetic ancestry covariates improved predictions for thyroid cancer (PRS_ThC vs. All_ AUC 0.721 [0.707,0.735]) and other thyroid diseases (AUC ranging from 0.686 (0.655,0.718) for benign nodular goiter to 0.727 (0.709,0.745) for hypothyroidism). We expected this improvement because of the higher incidence of thyroid diseases in women^41^ and the increased risk of developing thyroid nodules and hypothyroidism with age^8,42^. However, no significant improvement in clinical PRS_ThC vs. BNG_ performance was observed (**Supplementary Table 14**).

We did not find a significant drop in PRS performance measured with AUC in EUR-like, AMR-like, and AFR-like strata (De-Long test, p-value > 0.05) except for hypothyroidism PRS in AFR-like individuals (**Extended Data Figure 6**).

### Polygenic risk score and thyroid cancer aggressiveness

We evaluated associations between thyroid cancer PRS and aggressive features of thyroid cancer in three domains (patient, tumor, and metastatic disease), abstracted from surgical histopathology reports and clinical notes (**Figure 3C, Supplementary Table 15**). PRS_ThC vs. All_ was significantly associated with tumor focality and extranodal extension. PRS_ThC vs. BNG_ was significantly associated with tumor focality and the number of neck lymph node metastases (p-value ≤ 1.7e-3, Bonferroni-adjusted p-value threshold). In addition, PRS_ThC vs. All_ was associated with lymphatic invasion, and PRS_ThC vs. BNG_ was related to the risk of structural disease recurrence (defined per the American Thyroid Association guidelines ^43^) at a nominal p-value threshold of ≤0.05.

## Discussion

We completed the largest GWAS meta-analysis for five thyroid diseases, leveraging a global collaboration involving 19 Biobanks from 10 countries. The Consortium replicated 235 genetic associations deposited in the NHGRI-EBI GWAS Catalog as of April 2024 (v.1.0.2) and discovered 501 new independent associations (**Table 1**).

Genetic correlation analysis (**Figure 2**) identified shared genetic architecture between thyroid diseases that is physiologically plausible and clinically meaningful. Chronic lymphocytic thyroiditis is a leading cause of primary hypothyroidism^44^, explaining the near-perfect genetic correlation between these two diseases. The shared genetic basis for lymphocytic thyroiditis and Graves’ disease is also expected because both conditions are autoimmune diseases with highly concordant familial risk ^45^. The genetic correlation between Graves’ disease and thyroid nodular disease (both benign nodules and thyroid cancer) may be mechanistically explained by enhanced thyroid-stimulating hormone (TSH) receptor signaling, which promotes thyroid epithelial growth and protects thyroid cells from apoptosis ^46^. Previous population-based studies found an increased risk of thyroid (hazard ratio 10-15) and other cancers in patients with Graves’ disease ^47^, consistent with our findings.

Shared (genetic correlation *r^2^* = 0.4−0.5) and unique genetic associations with thyroid cancer and benign nodular goiter allowed insights into genes and pathways that lead to malignant and benign thyroid nodules. Our hypothesis explaining why some individuals are susceptible to thyroid nodules while others develop thyroid cancer is shown in **Figure 4**. We propose that two biological processes with distinct genetic architecture cause thyroid nodules: 1) hyperplasia, a polyclonal follicular cell proliferation with no malignant potential, and 2) neoplasia, a clonal growth driven by somatic genetic alterations. Neoplastic nodules can be benign or malignant, resulting in the mismatch between biological mechanisms (hyperplasia vs. neoplasia) and GWAS phenotype definitions (benign and malignant thyroid nodules), resulting in partial overlap in genetic associations and significant genetic correlation between thyroid cancer and benign nodular goiter phenotypes.

**Figure 4.**
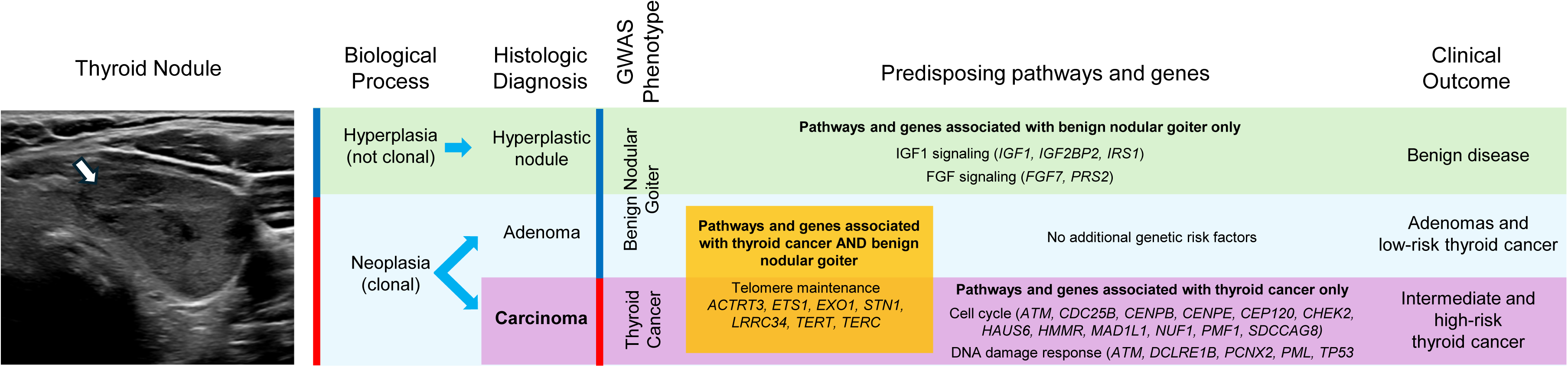
Germline genetic susceptibility to thyroid cancer and benign nodular goiter. We hypothesize that two biological processes with distinct genetic architecture cause thyroid nodules: 1) hyperplasia, a polyclonal follicular cell proliferation with no malignant potential, and 2) neoplasia, a clonal growth driven by somatic genetic alterations. Neoplastic nodules can be benign or malignant, and the mismatch between biological mechanisms (hyperplasia vs. neoplasia) and GWAS phenotype definitions (benign and malignant thyroid nodules) caused apparent genetic pleiotropy. Pathway and genes associated with benign nodular goiter but not thyroid cancer in GWAS meta-analysis (e.g., insulin-like growth factor 1 [IGF1] and fibroblast growth factor [FGF] signaling pathways) predispose to benign nodules that have no malignant potential (green). Pathways and genes associated with both benign nodular goiter and thyroid cancer (e.g., telomere maintenance) predispose to neoplastic thyroid nodules, benign or malignant (orange). In the absence of other genetic risk factors, patients are more likely to develop benign adenomas or low risk thyroid cancers (blue). Alternatively, genetic alterations in cell cycle and DNA damage response genes (purple, associated predominantly with thyroid cancer but nor benign nodular goiter in GWAS meta-analysis) predispose to high-risk thyroid cancer.

We found that genes participating in the cell cycle, DNA repair and cellular response to DNA damage are predominantly associated with thyroid cancer but not benign nodules, highlighting the importance of these biological processes for malignant transformation of thyroid follicular cells. These variants and genes can lead to more aggressive multifocal and metastatic thyroid cancer (**Figure 3C**). On the contrary, genes in fibroblast growth factor and insulin-like growth factor 1 signaling pathways were uniquely associated with benign nodular goiter and may lead to hyperplastic benign thyroid nodules without malignant potential. Variants in genes participating in telomere maintenance increase the risk of thyroid cancer and benign neoplastic thyroid nodules (adenomas). It was demonstrated previously that telomere-lengthening germline variants predispose to papillary thyroid cancer. ^48^

Our finding that autoimmune thyroid disorders share most genetic associations (**Figure 2**) indicates that similar fundamental mechanisms lead to Graves’ disease and lymphocytic thyroiditis/primary hypothyroidism despite opposite clinical manifestations.

Of special interest are genes that were only found in Graves’ disease meta-analysis despite the much greater discovery power of hypothyroidism GWAS. These unique associations (*CD2, CD40, etc.*) may be involved in the immune system processes that define the type of autoantibodies produced: TSHR antibodies in Graves’ disease or TPO/TG antibodies in lymphocytic thyroiditis and primary hypothyroidism.

Consistently, variants in *TSHR* was strongly associated with Graves’ disease (e.g., 14:80990913:A:C, β=0.27 [0.01], p-value = 2.52e-137), while *TPO* and *TG* associations were only seen in hypothyroidism meta-analysis.

Thyroid cancer caused 2,170 deaths in the United States in 2024^1^. PRS derived from thyroid cancer GWAS can identify individuals at thyroid cancer risk in the population (**Figure 4A** and **B**, and ^49^). Thyroid cancer screening is not currently recommended by the US Preventive Services Task Force (USPSTF) ^50^ due to concerns about overtreatment and lack of mortality benefit. However, we found that PRS is associated with high-risk thyroid cancer features (**Figure 4C**). Genetically-informed screening, when thyroid ultrasound is recommended only for a subset of individuals with high PRS, will not only increase diagnostic yield but may help identify patients who will benefit from early diagnosis and screening. Clinical testing of such an approach is needed.

Another clinically meaningful application for thyroid cancer PRS is for aiding in the diagnosis of thyroid cancer in patients with thyroid nodules ^14^. Despite the widespread use of clinical ultrasound-based algorithms ^8,9^, 72% of FNAs produce benign results, 20% are inadequate or indeterminate ^12,13,51,52^. PRS provides cancer risk assessment that is complementary and synergistic to ultrasound-based nodule evaluation ^14^.

We found that PRS derived from thyroid cancer GWAS meta-analysis alone is inferior for distinguishing benign and malignant thyroid nodules (because many genetic associations are shared between these two phenotypes, **Supplementary Table 14**).

Incorporating variants from both thyroid cancer and benign nodular goiter meta-analysis (PRS_ThC vs. BNG_) improved PRS performance. Active surveillance of thyroid nodules with low-risk sonographic appearance in patients with reassuring PRS could reduce the need for invasive procedures. Other putative clinical applications for PRS that have not yet been explored are for molecular diagnostics of thyroid nodules with indeterminate FNA cytology and to personalize decisions to proceed with hemi-or total thyroidectomy for thyroid cancer.

PRSs for autoimmune thyroid diseases have significant predictive power (e.g., covariate-adjusted hypothyroidism PRS AUC of 0.721 [0.701, 0.742]). However, both hypothyroidism and Graves’ disease are reliably diagnosed with biochemical and antibody testing, and the utility of genetic risk evaluation is uncertain.

We recognize that, due to the demographics of participants in the Virtual Thyroid Biopsy Consortium, we are underpowered in our ability to study individuals of non-EUR-like ancestry. As our consortium grows, we look forward to conducting more ancestry-specific analyses to ensure we find the results relevant to all individuals^53^, and improve our insights into rare variation across groups.

In summary, we conducted the largest meta-analysis of GWAS for five thyroid diseases. We found many previously known and novel mechanistically plausible variants, genes, and pathways contributing to the risk of thyroid cancer, benign nodular goiter, and autoimmune thyroid diseases. We explained why some individuals are prone to developing benign thyroid nodules while others are at risk of multifocal metastatic thyroid cancer. We derived and tested polygenic risk scores for thyroid cancer population screening and for a more clinically relevant task of distinguishing benign and malignant thyroid nodules. This study will serve as a foundation for future clinical applications leveraging the germline genetics of thyroid diseases.

## Supporting information

Banner Authorships

## Data Availability

The meta-analysis GWAS summary statistics will be deposited in the GWAS Catalog after the acceptance of this manuscript for peer-reviewed publication. The PRS weights will be deposited in the PGS Catalog (https://www.pgscatalog.org/). All original code is publicly available at GitHub at https://github.com/pozdeyevlab/gwas-analysis.

Further information and requests for resources should be directed to and will be fulfilled by the lead contact, Nikita Pozdeyev.

## Acknowledgments.

We want to acknowledge the participants and investigators of all participating biobanks. We thank Regeneron Genetics Center for genotyping part of the Colorado Center for Personalised Medicine data.

We gratefully acknowledge All of Us participants for their contributions, without whom this research would not have been possible. We also thank the National Institutes of Health’s All of Us Research Program for making available the participant data examined in this study.

The authors would like to acknowledge the Oncology Research Information Exchange Network (ORIEN) Institution Members for their commitment to data sharing. Thyroid cancer gene expression data included in this work was obtained through the Avatar Project.

Genes & Health thanks Social Action for Health, Centre of The Cell, members of our Community Advisory Group, and staff who have recruited and collected data from volunteers. We thank the NIHR National Biosample Centre (UK Biocentre), the Social Genetic & Developmental Psychiatry Centre (King’s College London), Wellcome Sanger Institute, and Broad Institute for sample processing, genotyping, sequencing and variant annotation. This work uses data provided by patients and collected by the NHS as part of their care and support. This research utilised Queen Mary University of London’s Apocrita HPC facility, supported by QMUL Research-IT, http://doi.org/10.5281/zenodo.438045

Genes & Health thanks Barts Health NHS Trust, NHS Clinical Commissioning Groups (City and Hackney, Waltham Forest, Tower Hamlets, Newham, Redbridge, Havering, Barking and Dagenham), East London NHS Foundation Trust, Bradford Teaching Hospitals NHS Foundation Trust, Public Health England (especially David Wyllie), Discovery Data Service/Endeavour Health Charitable Trust (especially David Stables), Voror Health Technologies Ltd (especially Sophie Don), NHS England (for what was NHS Digital) - for GDPR-compliant data sharing backed by individual written informed consent.

## Contributions

Nikita Pozdeyev and Christopher R. Gignoux conceived, designed and supervised the project. Samantha L. White and Nikita Pozdeyev conducted the main analyses in this study. Nikita Pozdeyev and Samantha L. White wrote the initial draft of the paper. Maizy S. Brasher and Joanne B. Cole contributed data and analyses from the UK Biobank. Jibril Hirbo, Nancy J. Cox, and Peter Straub contributed data and analyses from the BioVU. Shinichi Namba, Koichi Matsuda and Yukinori Okada contributed data and analyses from the BioBank Japan Project. Emily Bertucci-Richter and Sebastian Zoellner contributed data and analyses from the Michigan Genomics Initiative. Lindsay Guare and Anurag Verma contributed data and analyses from the Penn Medicine BioBank. Ahmed EdrisMohammed, Sam Morris, Robin Walters and Zhengming Chen contributed data and analyses from the China Kadoorie Biobank. Ashley J Mulford and Alan R Sanders contributed data and analyses from the Genomics Health Initiative.

Brian Fennessy, Michael Preuss and Eimear Kenny contributed data and analyses from the BioMe. Jack Pattee, Sameer Chavan, Matthew J. Fisher, Nicholas Rafaels, Meng Lin, Jonathan Shortt and Kristy Crooks contributed data and analyses from the biobank at the Colorado Center for Personalized Medicine. Yon Ho Jee, Peter Kraft and Alicia Martin contributed data and analyses from the Korean Cancer Prevention Study-II. Martin D Tobin, Jing Chen, Alexander T, Williams and Catherine John contributed data and analyses from the EXCEED study. David A. van Heel, Rohini Mathur and Sarah Finer contributed data and analyses from the Genes and Health study. Marta Riise Moksnes, Ben Brumpton, and Bjørn Olav Åsvold contributed data and analyses from the Trøndelag Health Study. Raitis Peculis, Vita Rovite, and Ilze Konrade contributed data and analyses from the Latvian Biobank. Ying Wang contributed data and analyses from the Mass General Brigham Biobank. David Whiteman, Stuart MacGregor and Sarah Medland contributed data and analyses from the QSkin Sun and Health Study.

Unnur Thorsteinsdóttir and Kári Stefánsson contributed data and analyses from deCODE Genetics. Lauren Fishbein contributed data and analyses from the Million Veteran Program. Kathleen M. Egan, Therese Bocklage, Hilary C McCrary, Greg Riedlingeer, Bodour Salhia, Craig Shriver, Minh D Phan, Janice L. Farlow, Stephen Edge, Varinder Kaur, Michelle Churchman, Robert J. Rounbehler, Pamela L. Brock, and Matthew D. Ringel contributed the Oncology Research Information Exchange Network (ORIEN) AVATAR study data. Haoyu Zhang and Milton Pividori performed S-PrediXcan analyses. Caitlin C. Bell, Taylor L. Jamil, Martin Barrio, Christopher D. Raeburn and Bryan R. Haugen collected and interpreted thyroid cancer clinical data. Tugce Karaderi contributed to the interpretation of the hypothyroidism meta-analysis data. Rebecca Schweppe assisted with the interpretation of pathway analysis data. Audrey Hendricks advised on the use of Summix2 for population structure estimates. Wei Zhou, Sinéad Chapman, Mark Daly and Benjamin Neale led the Global Biobank Meta-analysis Initiative. All authors contributed and approved the final version of the paper.

## Methods

### Ethical approval

Colorado Multiple Institutional Review Board for the University of Colorado Denver Anschutz Medical Campus, Aurora, Colorado, USA, waived ethical approval for this work (COMIRB #20-2315).

This research has been conducted using the UK Biobank Resource under Application Number 95339.

### Virtual Thyroid Biopsy Consortium

We founded the Virtual Thyroid Biopsy (VTB) Consortium under the umbrella of the Global Biobank Meta-analysis Initiative (GBMI) ^21^. Nineteen biobanks from ten countries and four continents contributed GWAS results to the meta-analysis (**Extended Data Figure 1**). **Supplementary Table 1** lists biobank size, available ancestry strata, phenotyping, genotyping and imputation methods, and software for GWAS.

### Phenotype definitions

We defined thyroid phenotypes using International Classification of Diseases ICD-9-CM and ICD-10-CM billing codes for United States biobanks, ICD-9 and ICD-10 billing codes for international biobanks, and SNOMED codes and survey codes for the All of Us Research Program biobank (**Supplementary Table 2**). These phenotype definitions were shared with teams participating in the Consortium.

To evaluate polygenic risk score performance and study its association with thyroid cancer risk phenotypes, we performed clinical chart reviews for genotyped participants in the Colorado Center of Personalized Medicine (CCPM) Biobank.

Histopathologic and cytologic diagnosis, patient characteristics (age at thyroid cancer diagnosis, death from thyroid cancer and risk of structural disease recurrence), tumor characteristics (tumor size, tumor focality, presence of extrathyroidal extension, lymphatic and angioinvasion, surgical margins positivity) and metastatic disease characteristics (presence of locoregional and distant metastases, extranodal extension, size and number of lymph node metastases) were extracted from surgical histopathology reports, thyroid nodule fine-needle aspirations reports, and endocrinology notes.

The risk of structural disease recurrence was estimated on a continuous scale (1-55% risk) as described in the American Thyroid Association thyroid cancer guidelines.^43^ For patients with multiple surgeries, the highest stage/risk was used (e.g., if the first surgery histopathology evaluation reported Nx stage but lateral neck metastases were found later, N1b stage was used for the association analysis). Thyroid cancer risk phenotypes are summarized in **Supplementary Table 14**. Benign cases for PRS evaluation in CCPM were defined based on surgical histopathology reports.

### Genome-wide association studies (GWAS)

Case and control definitions for GWAS are listed in **Supplementary Table 3.**

Phenotype exclusions were used only if clinically or biologically justified. We excluded:

1) patients diagnosed with medullary thyroid cancer from thyroid cancer GWAS (if medullary thyroid cancer data was available; because rare medullary thyroid cancers are genetically distinct from the common follicular cell-derived thyroid cancers); 2) thyroid cancer cases from benign nodular goiter GWAS (because all thyroid cancers are initially diagnosed as thyroid nodules to avoid contamination of benign nodular goiter cases with malignant tumors), and 3) patients diagnosed with hypothyroidism other than primary (iatrogenic, congenital, central) from hypothyroidism GWAS.

Each biobank conducted genotyping, imputation, quality control, and genetic ancestry analysis independently (**Supplementary Table 1**), except for the All of Us Research Program Biobank, where a custom pipeline was designed to use whole genome sequencing data and maximize variant overlap with other Biobanks.

GWAS analyses were run using either linear mixed models (SAIGE) ^54^ or whole genome regression (REGENIE)^55^, adjusted for case-control imbalances using saddlepoint approximation or Firth’s logistic regression. The biobanks were instructed to use age, sex, up to 20 first principal components, and biobank-specific variables (such as genotyping batches and recruiting centers) as covariates.

In addition to multi-ancestry analyses, GWAS stratified by genetic ancestry were performed when the case counts permitted. **Supplementary Table 4** lists case and control counts, Summix2 ^56^ population structure estimates, and quality control metrics calculated with the covariate-adjusted LD-score regression^26^ for 198 GWAS analyses integrated into the meta-analyses.

### Genome-wide association studies in the All of Us (All of Us) Research Program Biobank

We used All of Us whole genome sequencing v7 (WGS) data (245,388 WGS) to produce a genetic dataset that maximizes variant overlap with the analyses performed in the other biobanks (**Extended Data Figures 7 and 8**). An inclusive list of single-nucleotide polymorphisms (SNPs) and indels from GWAS analyses was compiled and supplemented with variants from the Polygenic Score Catalog (reported as of February 2024). This list contained ∼ 147 million SNPs and indels.

WGS variant level QC was performed by All of Us as reported in the All of Us Research Program Genomic Research Data Quality Report ^59^. In addition, we filtered the dataset to a maximal set of unrelated samples estimated from kinship scores and only included individuals with electronic health records and/or survey data for phenotype definitions (193,429 WGS).

We developed a Hail Python pipeline that extracts variants of interest from the AoU variant dataset (VDS) (https://hail.is/docs/0.2/vds/index.html). The code is publicly available in the GitHub repository for this study (https://github.com/pozdeyevlab/vds-filter/tree/main). The resulting BGEN dataset contained ∼118 million directly genotyped variants (a significant decrease from 972 million variants in VDS), permitting GWAS using REGENIE version 3.2.4.

### Post-GWAS quality control

The post-GWAS quality control workflow diagram is shown in **Extended Data Figure 9**. All GWAS summary data were harmonized to gnomAD (v4.1.0) (GRCh38 human genome reference)^28^.

Each GWAS summary data set (Supplementary Table 4) was processed using the following steps.

1. *<u>Variant-level quality control.</u>* The following variants were removed from the GWAS summary data:

a. Variants containing alleles with characters other than A,T,C, or G.
b. Variants with a p-value equal to zero, effect size (β) or standard error ≥ 1e6 or ≤-1e6, and variants with an imputation score < 0.3.
c. Variants with AF < 0.0005 or > 0.9995.
d. Variants with allele count < 20.
e. The variants were aligned to gnomAD (v4.1.0) reference. Ancestry-specific gnomAD allele frequencies (AFs) were used for single-ancestry GWAS. Both palindromic and non-palindromic variants were tested for exact and inverse alignments. Palindromic variants were removed due to potential strand flip if they met any of the following criteria: the fold difference between gnomAD AF and GWAS AF > 2; or GWAS AF > 0.4 and < 0.6; or GWAS AF < 0.4 and gnomAD AF > 0.6; or GWAS AF >0.4 and gnomAD AF < 0.6.
f. Variants flagged as low-quality by gnomAD.
g. Variants with Mahalanobis distance between gnomAD AF and harmonized GWAS AF of > 3 standard deviations from the mean.
2. *GWAS summary level data quality control*

a. *Population structure.* Summix2^56^ was used to estimate the population structure from the GWAS summary data. We used a random set of 10,000 variants from chromosome 21 and reference allele frequencies for AFR, AMR, EAS, NFE, MID and SAS genetic ancestry groups from gnomAD (v4.1.0). The results from five Summix2 runs, each using a different random set of reference variants, were averaged. We compared GWAS-derived Summix2 population proportion estimates to those published by MVP, CCPM and AoU and found almost perfect agreement (**Extended Data Figure 10**, *r^2^* = 0.999, p-value = 1.96e-22). Single-ancestry GWAS summary data analysis showed good agreement between the ancestry reported by the biobank and Summix2 estimate (median fraction of target ancestry is 0.88−0.97).
b. *Covariate-adjusted LD score regression* (cov-LDSC) ^26^ was used to evaluate for confounding in GWAS summary data, calculate the heritability of phenotypes, and estimate the genetic correlation between thyroid diseases. For each major continental ancestry, we generated a custom reference panel of 5,000 WGS from the AoU Biobank. For multi-ancestry GWAS, we used ancestry proportions calculated with Summix2 (**Supplementary Table 4**). Samples, regions and variants that met at least one of the following criteria were removed: 1) missingness of > 0.1; 2) closely related individuals (*plink king* cutoff of 0.0884); 3) Hardy-Weinberg equilibrium exact test p-value < 1e-6; 4) minor allele frequency of < 0.01, and 4) genomic regions with high linkage disequilibrium (LD). Genetic principal components were calculated using plink2. Ten principal components and a window of 20 cM were used to calculate covariate-adjusted LD scores and estimate LD score regression intercept (**Supplementary Table 4**), heritability (**Supplementary Table 6**), and genetic correlations (**Supplementary Table 7**) ^26^.

### GWAS meta-analysis

Fixed inverse variance weighted meta-analysis was run using METAL software.^62^ Individual GWAS summary data with cov-LDSC y-axis intercepts significantly deviating from one were adjusted before meta-analysis.

### Post-meta-analysis quality control, variant annotation and classification

To minimize false positive hits introduced by confounding within a single large biobank, only variants present in at least four input GWAS datasets were considered in the downstream analysis. If three or fewer datasets were available for the ancestry-stratified meta-analysis, then the threshold was set to two. Cochran’s Q p-values were calculated to assess heterogeneity across datasets.

We used hg38 human genome reference throughout the study. A genomic position-based algorithm similar to the one published in the GBMI flagship paper^21^ was used to define a set of non-overlapping genome-wide significant (p-value ≤ 5e-8) variants. The algorithm ensures that there is at least 500 kb between significant variants in two neighboring loci. Lead variants were mapped to the nearest gene and annotated using ANNOVAR (version date June 7, 2020)^63^. A locus was considered novel if no variants for the corresponding phenotype were reported within ±500 kb in the GWAS catalog (as of April 2024, v.1.0.2) ^29^. Otherwise, the variant was labeled as previously discovered.

### Heritability estimation and genetic correlation analysis

We used cov-LDSC (V. 1.0.0) ^26^ with a custom population structure-matched linkage disequilibrium (LD) reference panel (see the section on GWAS summary level-data quality control) to calculate SNP-based heritability (*h^2^_SNP_*). Observed-scale heritability estimates and the corresponding SEs were converted to liability scale using phenotype population prevalence calculated in the All of Us v7 dataset (**Supplementary Table 6)**. Similarly, pairwise genetic correlations between the five thyroid phenotypes (**Supplementary Table 7**) were calculated using cov-LDSC with a custom LD-score reference panel.

### Polygenic Risk Score Calculation and Evaluation

To calculate and evaluate polygenic risk scores (PRS), we performed a leave-CCPM-biobank-out GWAS meta-analysis. We used similar data derived from the GBMI phase I thyroid cancer meta-analysis ^21^ in comparison. All PRSs in this study were tested on the current out-of-sample CCPM dataset (n = 94,651). This approach minimizes inflation of PRS performance due to overfitting. Adjusted PRS (covariates of age, sex and ten genetic principal components) were cross-validated (5-fold).

Our PRS was calculated as a weighted sum of independent genome-wide significant risk alleles. For the clinically relevant use case of distinguishing thyroid cancer from benign nodular goiter, we defined PRS_ThC vs. BNG_ as the difference between PRS for thyroid cancer (PRS_ThC vs. All_) and PRS for benign nodular goiter (PRS_BNG vs. All_): PRS_ThC vs. BNG_ = PRS_ThC vs. All_ - PRS_BNG vs. All_. To compare with modern penalized methods, genome-wide PRSs were derived using the Bayesian regression framework within PRS-CS (version date May 14, 2024) ^40^. PRS-CS was run with a global shrinkage parameter of 1e-4, and all other fields were set as default.

PRS performance predicting binary phenotypes was assessed using the area under the receiver operating characteristic curve (AUC). AUCs were compared with DeLong’s test for significant differences.

### Transcriptome-wide association study (TWAS)

We performed *cis*-acting expression quantitative trait loci (*cis*-eQTL) TWAS using FUSION ^30^. FUSION was run on multi-ancestry and European meta-analysis summary data, 1000 genomes LD reference data, and all samples thyroid expression reference weights precomputed from GTEx v8 ^33^ (http://gusevlab.org/projects/fusion/).

To replicate our findings in FUSION, we also employed the Summary-PrediXcan (S-PrediXcan) ^32^ to derive gene-level association results from GWAS summary statistics and GTEx v8 ^33^ as the reference set. GWAS meta-analysis summary data were harmonized and imputed as described previously (https://github.com/hakyimlab/summary-gwas-imputation). An imputed GWAS was used to generate gene-trait associations in thyroid gland tissue.

### Candidate gene expression and pathway analysis

We studied gene expression of candidate genes linked to significant genetic associations discovered in the GWAS meta-analysis by ANNOVAR. In addition, we used significant (at a Bonferroni-corrected p-value threshold) protein-coding genes identified by *cis*-eQTLs TWAS. Intergenic variants that could not be unambiguously attributed to the expressed gene were not included.

mRNA expression of thyroid cancer-associated genes was compared in 20 human tissues using the National Center for Biotechnology Information Gene database (https://www.ncbi.nlm.nih.gov/gene ^34^). mRNA expression was analyzed in thyroid cancers from the Cancer Genome Atlas study ^35^ and the ORIEN AVATAR Program (https://www.oriencancer.org/research-programs). Gene expression was compared by age at diagnosis, cancer stage, somatic mutation status and other tumor features (**Supplementary Table 10**).

Reactome and KEGG pathway analyses were performed on all significant genes combined from FUSION and ANNOVAR using *ReactomePA* (v.1.16.2) ^38^ and *clusterProfiler* packages in R 4.4 with the default Benjamini-Hochberg adjustment for multiple hypothesis testing.

---

**Extended Data Figure 1.**
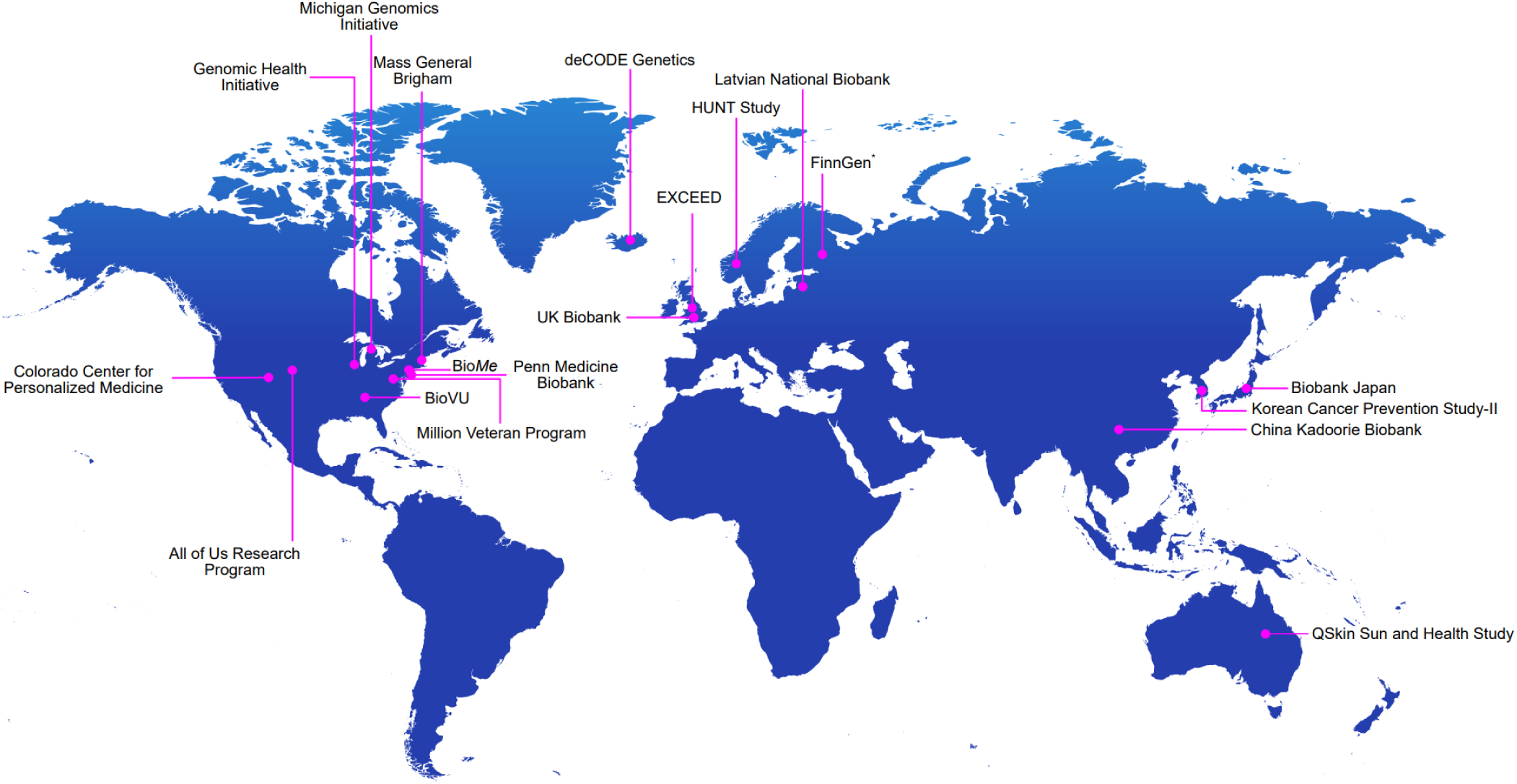
Virtual Thyroid Biopsy Consortium. The Consortium aggregated data from 19 biobanks, 10 countries, four continents and ∼2.9 million participants.

**Extended Data Figure 2.**
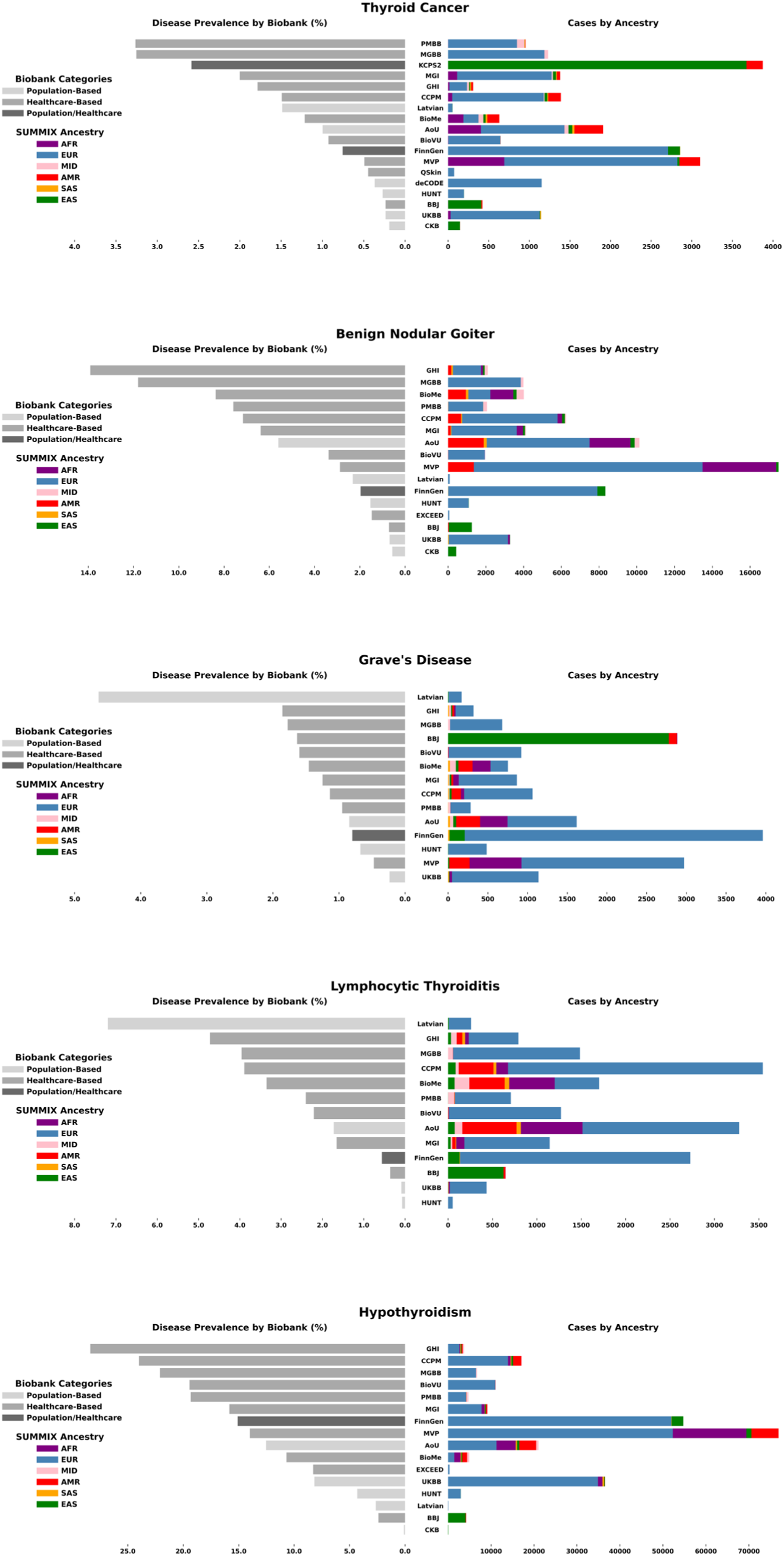
Thyroid disease meta-analysis case counts and prevalence across biobanks.

**Extended Data Figure 3.**
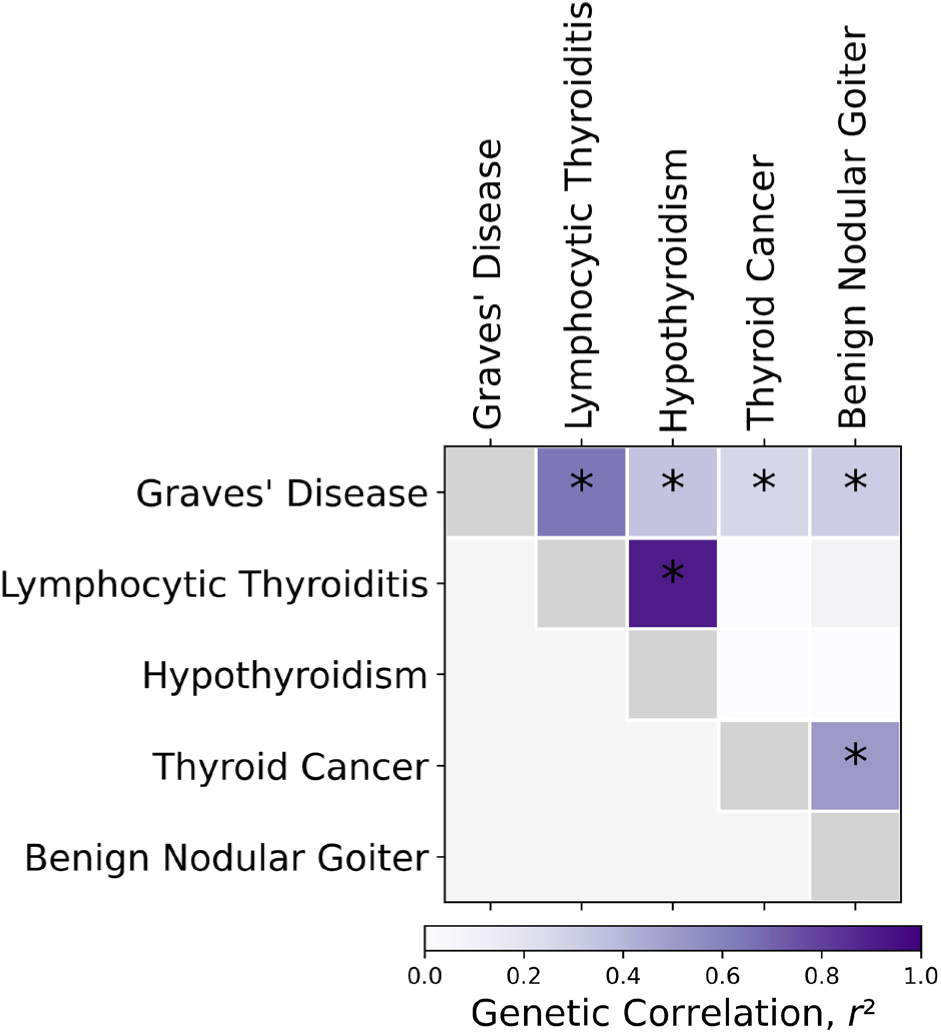
Genetic correlation analysis for thyroid diseases in EUR-like GWAS meta-analysis. Genetic correlations were estimated using covariate-adjusted LD score regression. The asterisks denote Benjamini-Hochberg false discovery rate (FDR) <0.05.

**Extended Data Figure 4.**
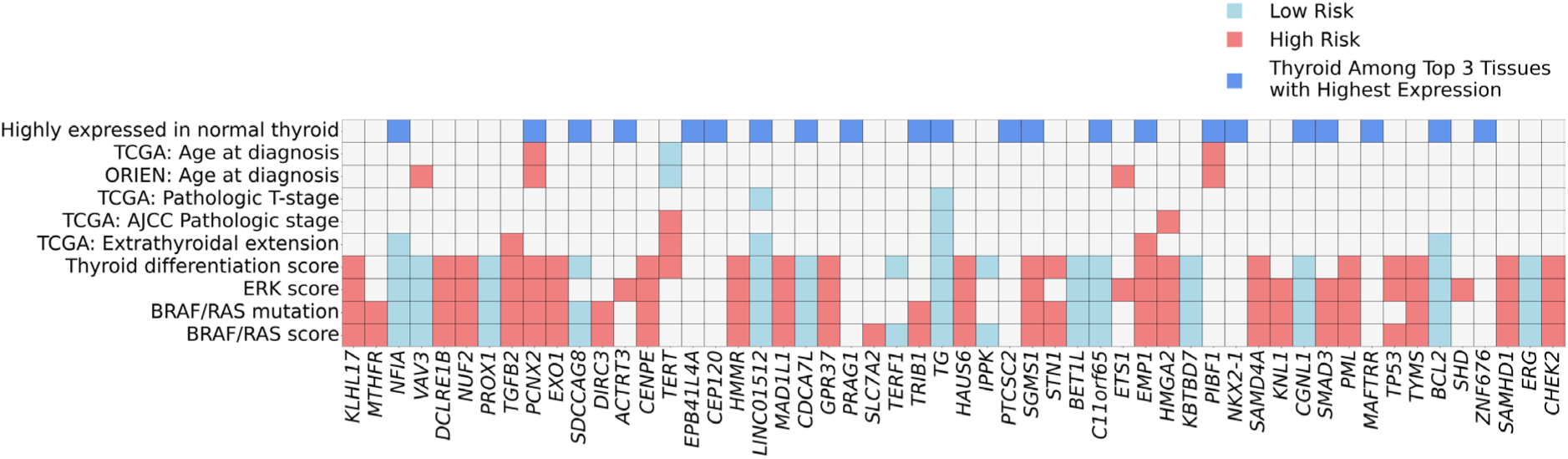
mRNA expression of thyroid cancer-associated genes in normal thyroid tissue and thyroid cancer. Genes were identified from ANNOVAR annotations of genome-wide significant variants in thyroid cancer GWAS meta-analysis and FUSION TWAS *cis*-eQTL analysis. Dark blue color shows genes with high expression in normal thyroid tissue (thyroid is among the top three highest expressing tissues in pan-tissue transcriptome analysis from NCBI Gene database https://www.ncbi.nlm.nih.gov/gene). Significant associations (at Bonferroni-corrected p-value ≤ 9.1e−05, red = positive; light blue = negative) of mRNA expression with high-risk thyroid cancer features such as presence of somatic *BRAF* V600E mutation, higher ERK score and lower thyroid differentiation score.

**Extended Data Figure 5.**
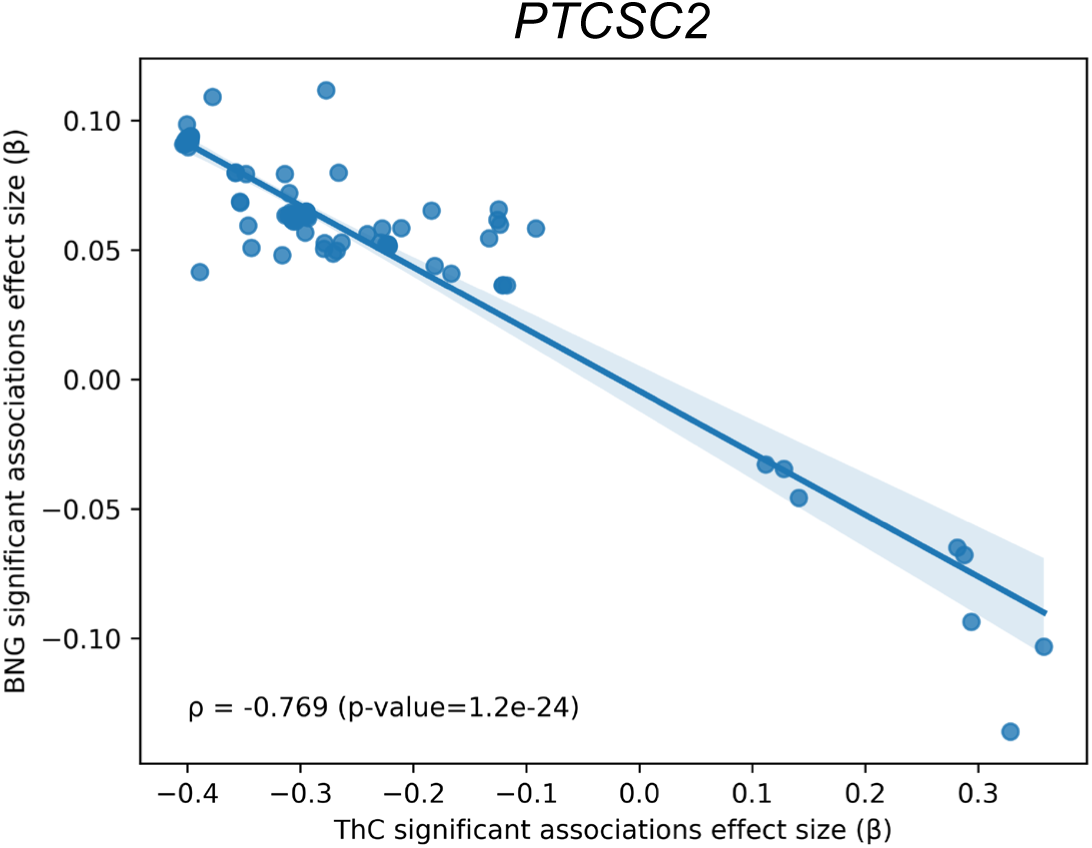
Scatterplot of effect sizes of the variants in *PTCSC2* locus significantly (p-value < 5e-8) associated with thyroid cancer and benign nodular goiter. ThC – thyroid cancer. BNG – benign nodular goiter. ρ - Spearman correlation.

**Extended Data Figure 6.**
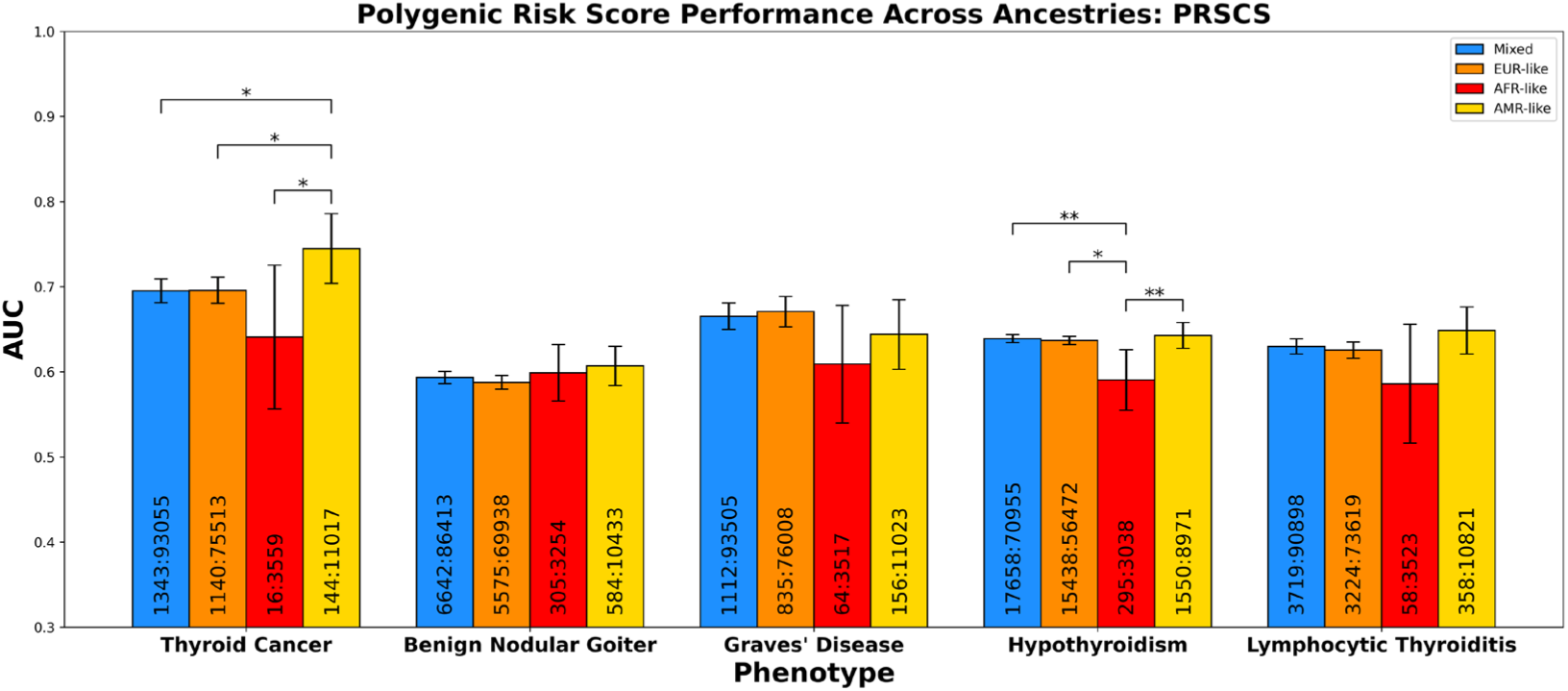
Thyroid cancer polygenic risk score (PRS_ThC vs. BNG_) performance by ancestry. Area under the receiver operating characteristic curve (AUC) and 95% confidence interval are shown for PRS_ThC vs. BNG_ developed from the leave-CCPM biobank-out thyroid cancer meta-analysis and tested on CCPM biobank participants of various ancestries. Case and control counts are shown within bars. * - p-value ≤ 0.05; ** - p ≤ 0.01.

**Extended Data Figure 7.**
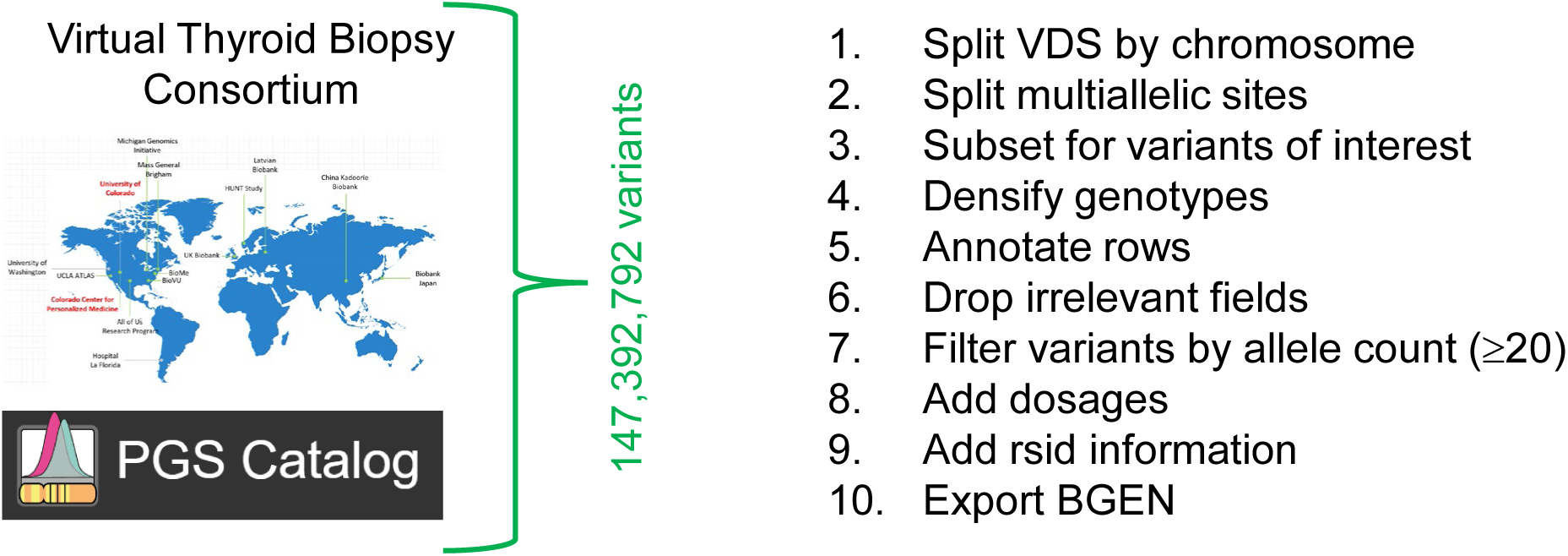
Whole genome sequencing data pipeline for the All of Us Research Program data. Variants (SNPs and indels) from participating biobanks’ GWAS summary data and PGS Catalog were extracted from the Hail variant dataset v7 object.

**Extended Data Figure 8.**
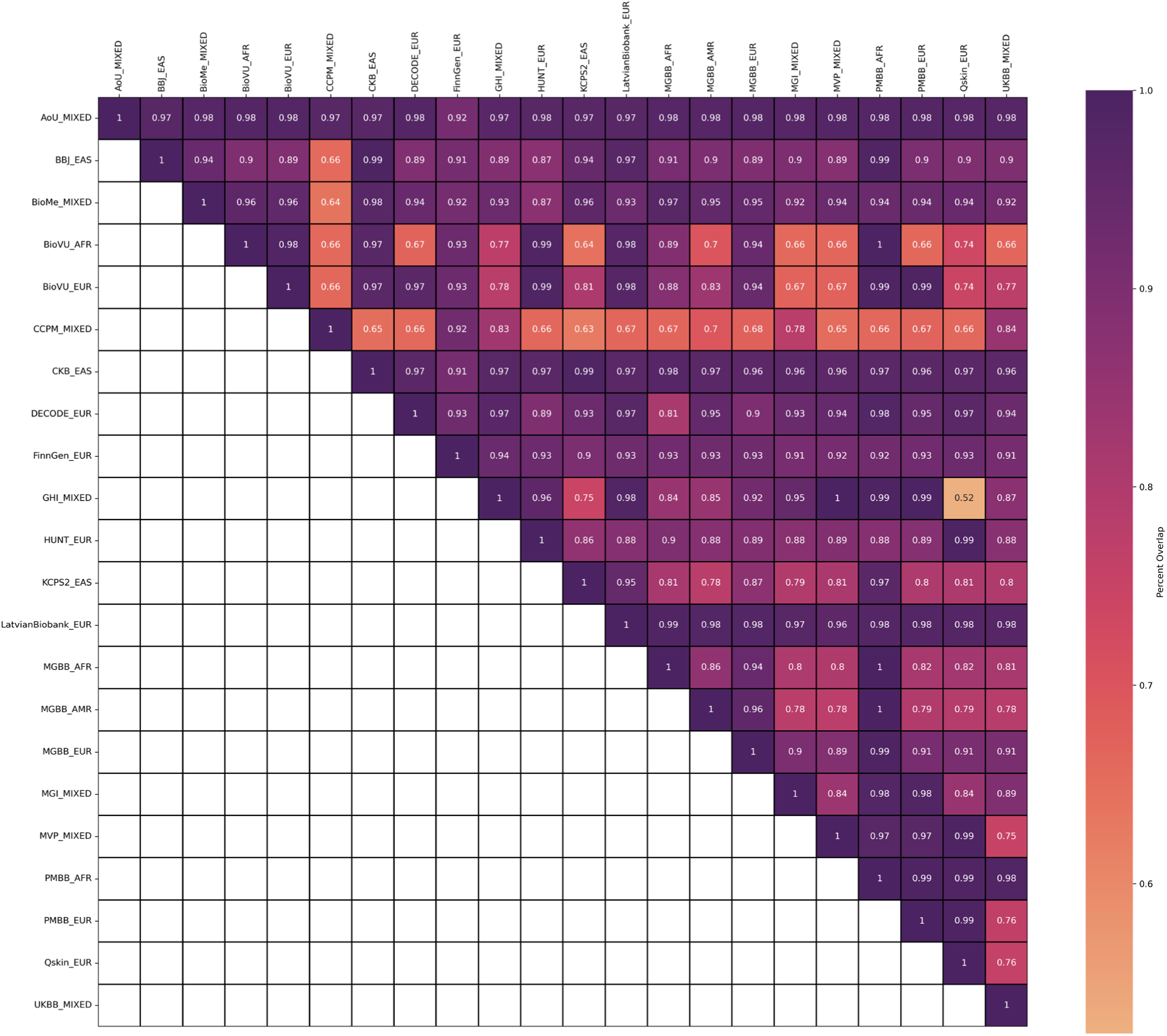
Variant overlap between GWAS from participating Biobanks. Data is shown as a fraction of variants that are identical by chromosome, position, reference and alternate allele in harmonized GWAS summary data from two biobanks. The All of Us Research Program GWAS (top row) was performed on whole genome sequencing data and was designed to maximize variant overlap with other biobanks.

**Extended Data Figure 9.**
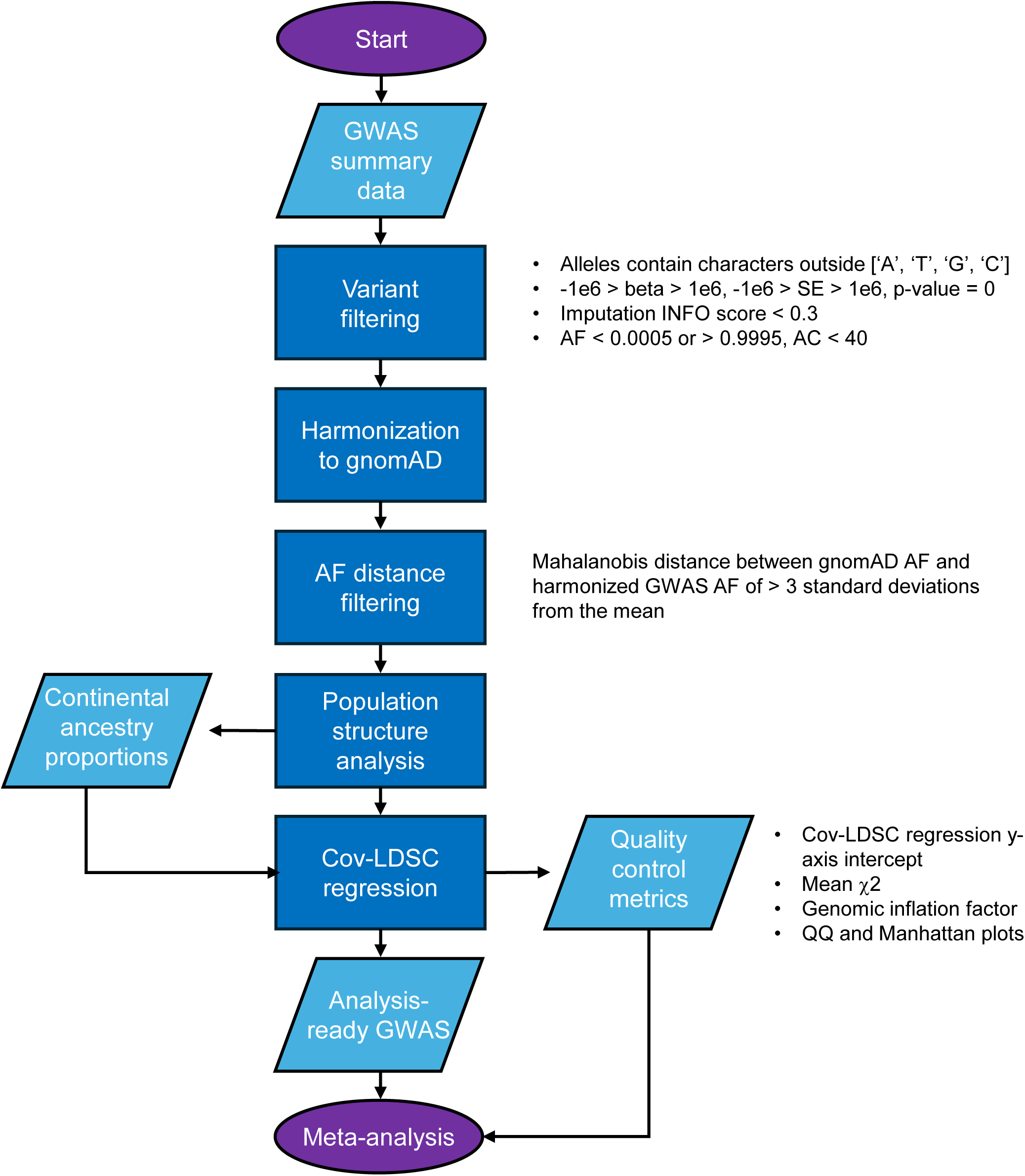
Post-GWAS quality control workflow diagram. AF – allele frequency. AC – allele count, cov-LDSC – covariate-adjusted LD score regression. QQ plot – quantile-quantile plot.

**Extended Data Figure 10.**
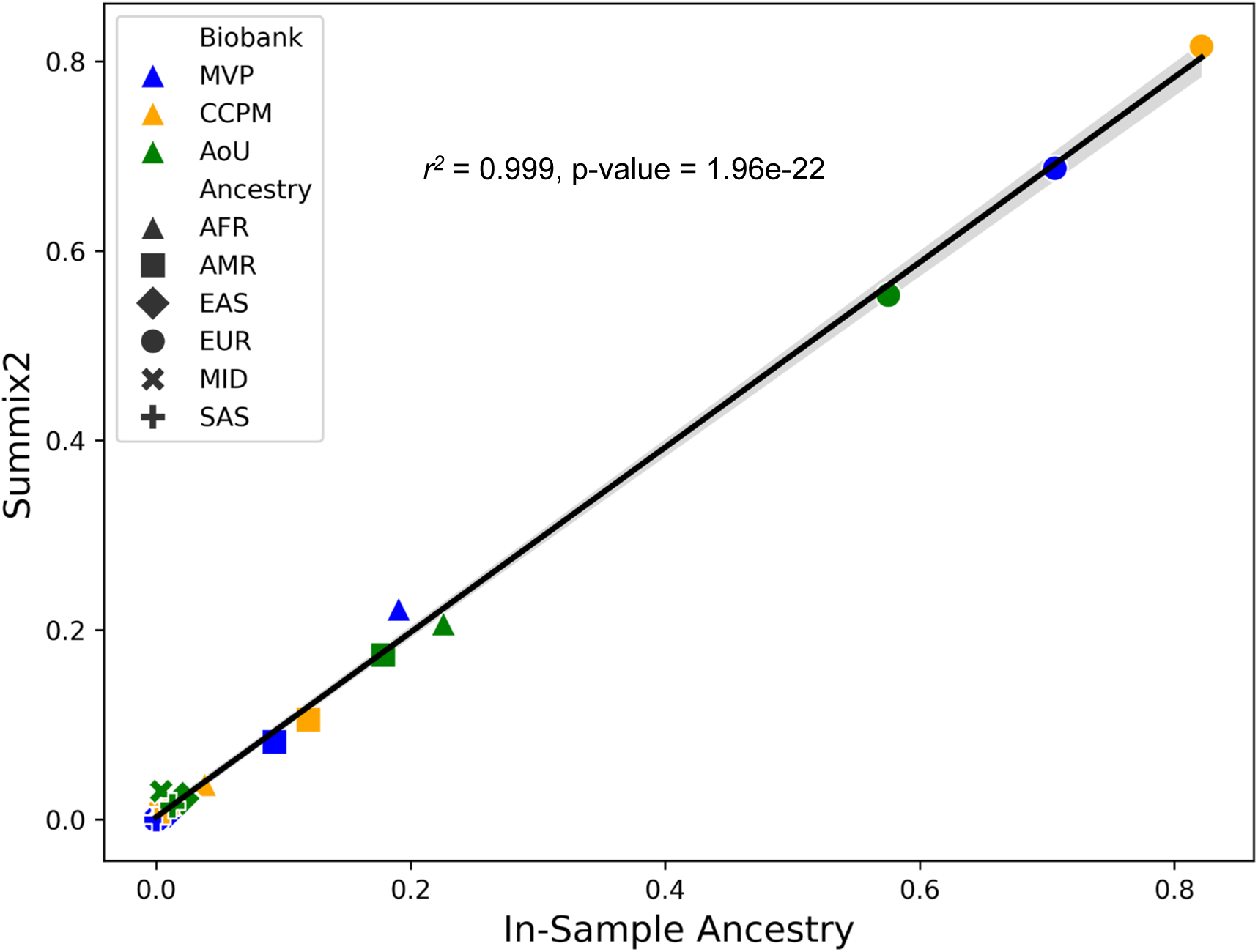
Correlation of major continental ancestry fractions estimated by Summix2 (y-axis) and published by the Million Veteran Program, Colorado Center for Personalized Medicine and All of Us Research Program Biobanks (x-axis). Multi-ancestry GWAS summary data was used for this analysis.

