## Supplementary Figure 1.1 for "Global multi-ancestry genetic study elucidates genes and biological pathways associated with thyroid cancer and benign thyroid diseases"

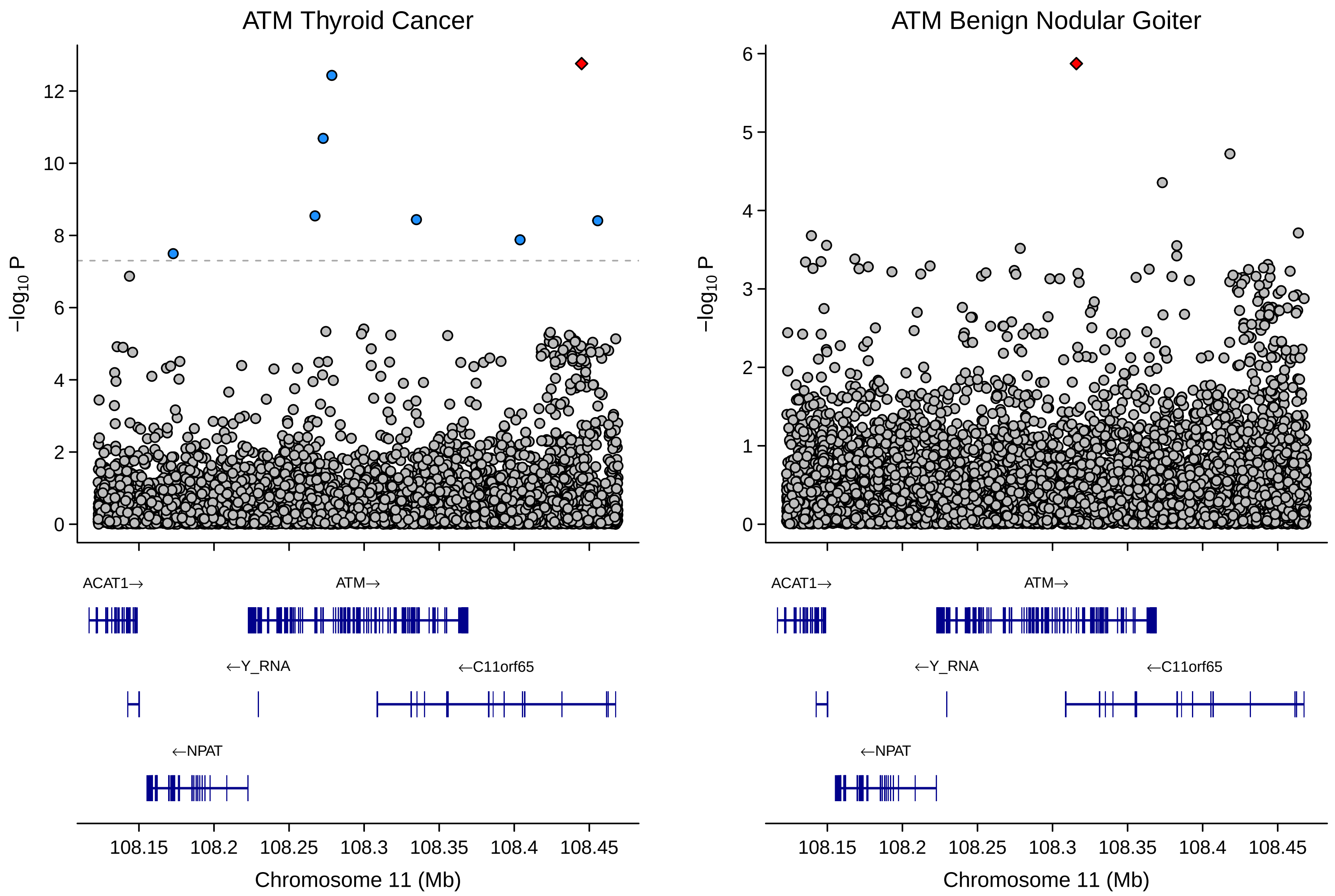

ATM Thyroid Cancer
ATM Benign Nodular Goiter
−log10 P
0
2
4
6
8
10
12
1
3
5
ACAT1→
ATM→
←Y_RNA
←C11orf65
←NPAT
108.15
108.2
108.25
108.3
108.35
108.4
108.45
Chromosome 11 (Mb)

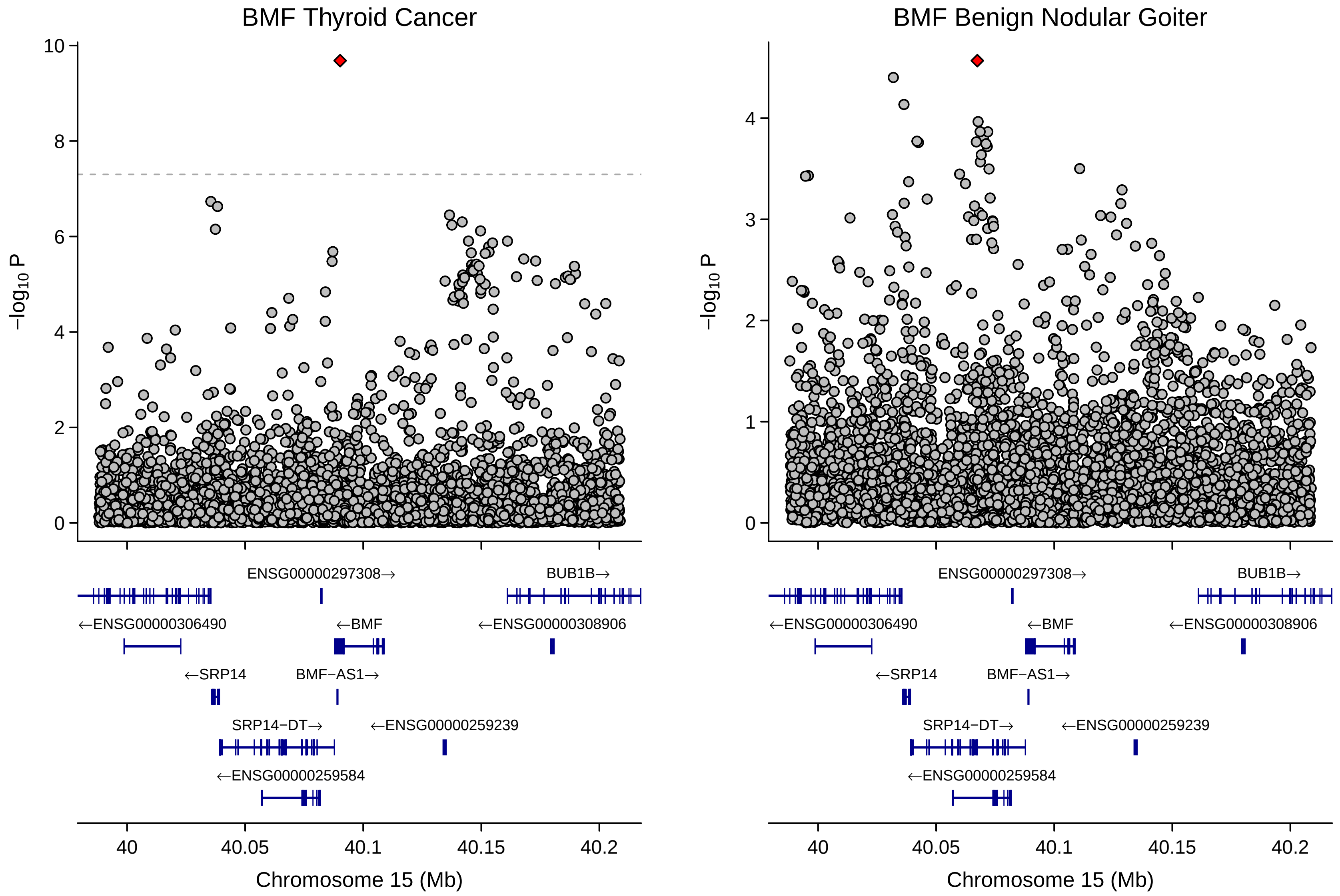
BMF Thyroid Cancer
BMF Benign Nodular Goiter
−log10 P
Chromosome 15 (Mb)
40
40.05
40.1
40.15
40.2
ENSG00000297308→
BUB1B→
←ENSG00000306490
←BMF
←ENSG00000308906
←SRP14
BMF−AS1→
SRP14−DT→
←ENSG00000259239
←ENSG00000259584

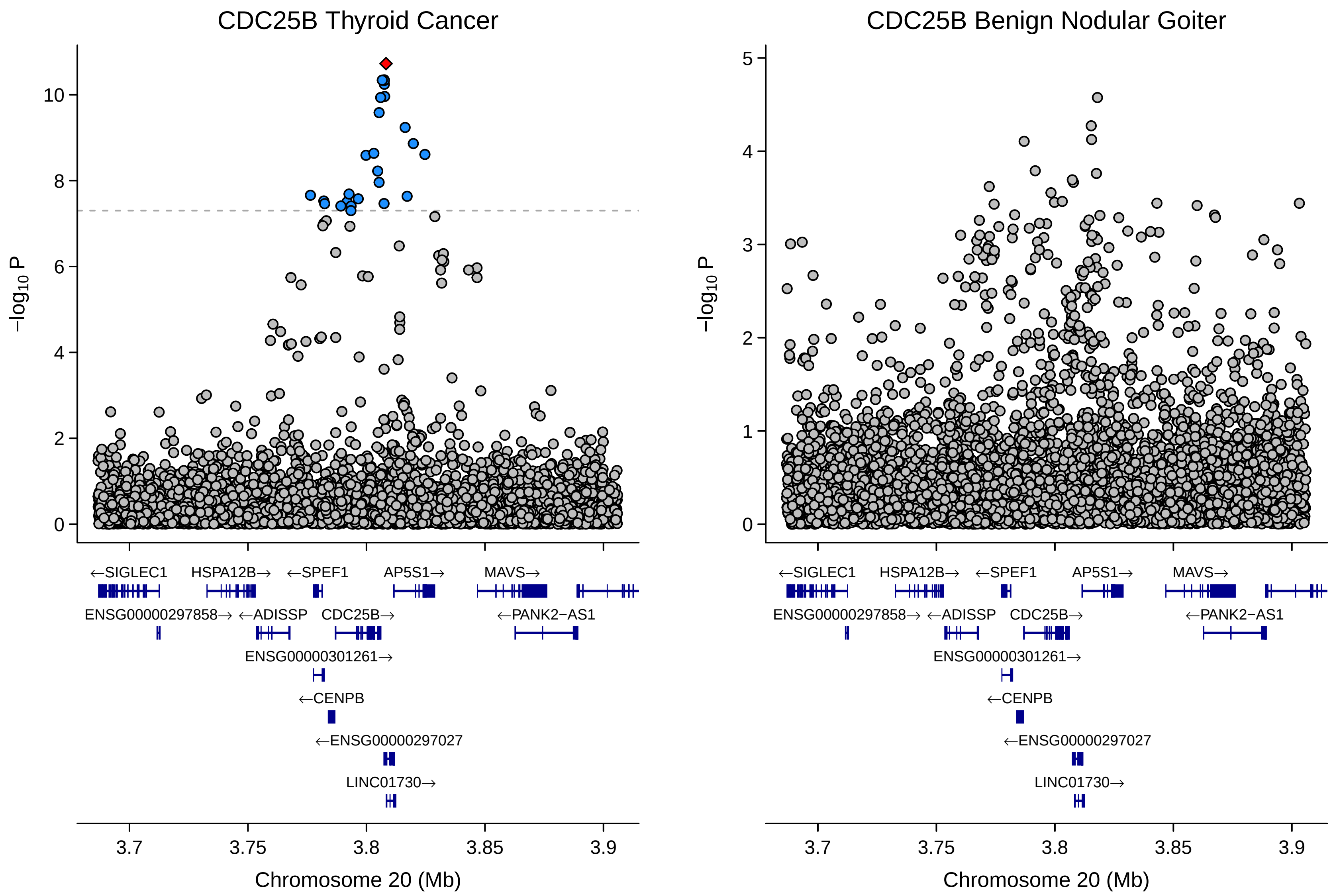

CDC25B Thyroid Cancer
CDC25B Benign Nodular Goiter
−log10 P
Chromosome 20 (Mb)
←SIGLEC1
HSPA12B→
←SPEF1
AP5S1→
MAVS→
ENSG00000297858→
←ADISSP
CDC25B→
←PANK2−AS1
ENSG00000301261→
←CENPB
←ENSG00000297027
LINC01730→

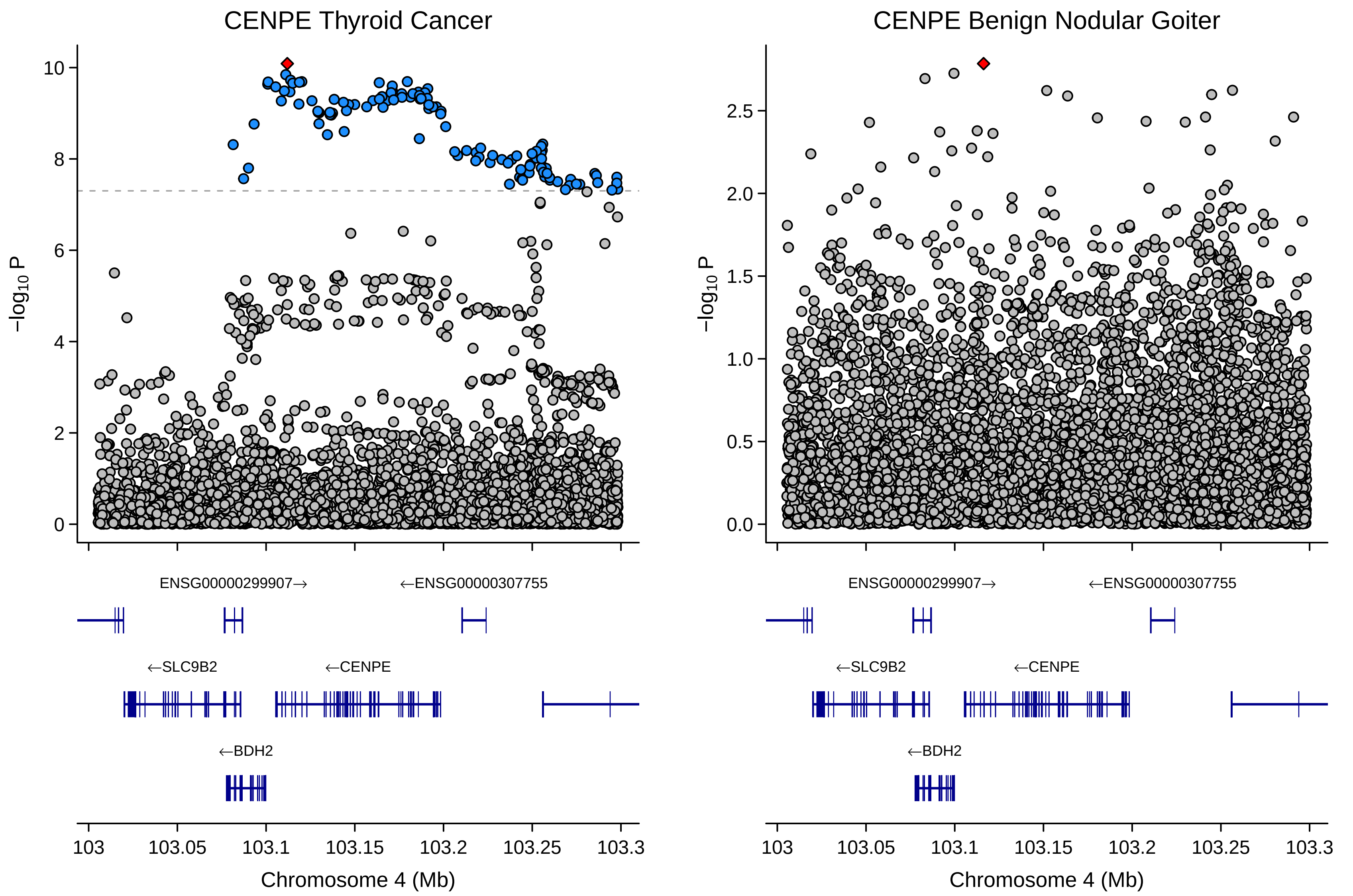

CENPE Thyroid Cancer
CENPE Benign Nodular Goiter
−log10 P
10
8
6
4
2
0
2.5
2.0
1.5
1.0
0.5
0.0
ENSG00000299907→
←ENSG00000307755
←SLC9B2
←CENPE
←BDH2
103
103.05
103.1
103.15
103.2
103.25
103.3
Chromosome 4 (Mb)

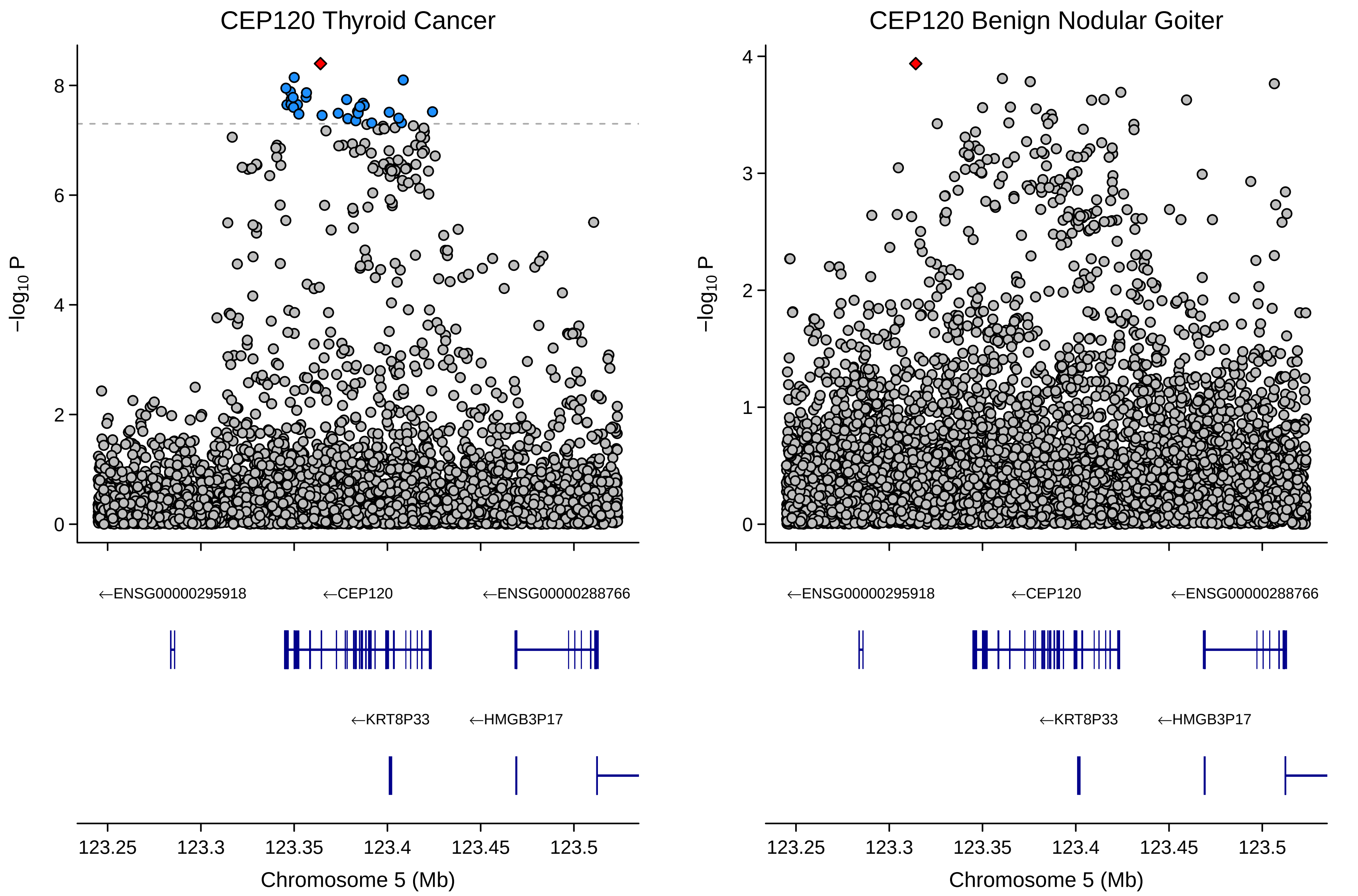

CEP120 Thyroid Cancer
CEP120 Benign Nodular Goiter
−log10 P
0
2
4
6
8
1
3
←ENSG00000295918
←CEP120
←ENSG00000288766
←KRT8P33
←HMGB3P17
123.25
123.3
123.35
123.4
123.45
123.5
Chromosome 5 (Mb)

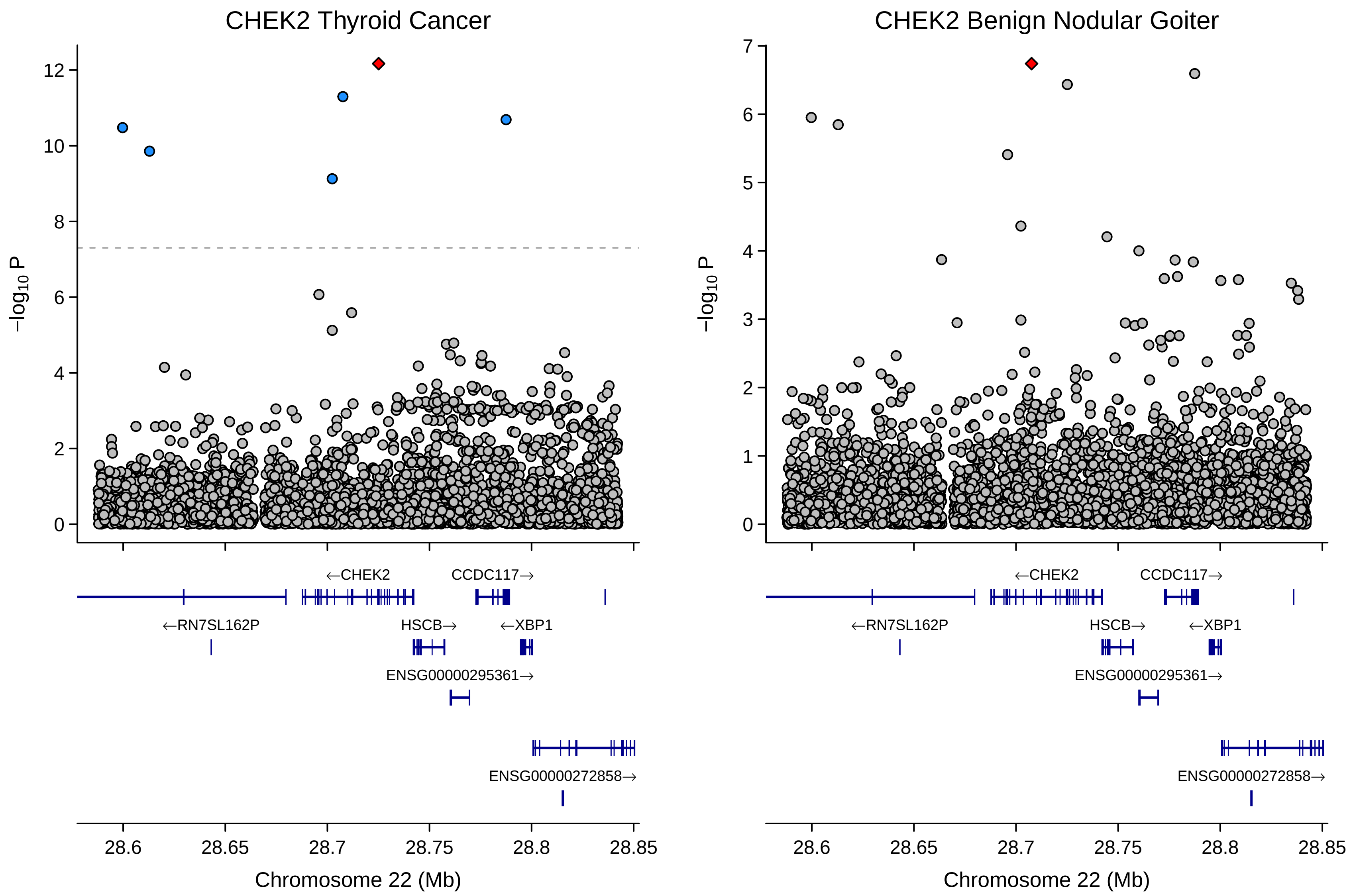

CHEK2 Thyroid Cancer
CHEK2 Benign Nodular Goiter
−log10 P
Chromosome 22 (Mb)
←CHEK2
CCDC117→
←RN7SL162P
HSCB→
←XBP1
ENSG00000295361→
ENSG00000272858→
28.6
28.65
28.7
28.75
28.8
28.85

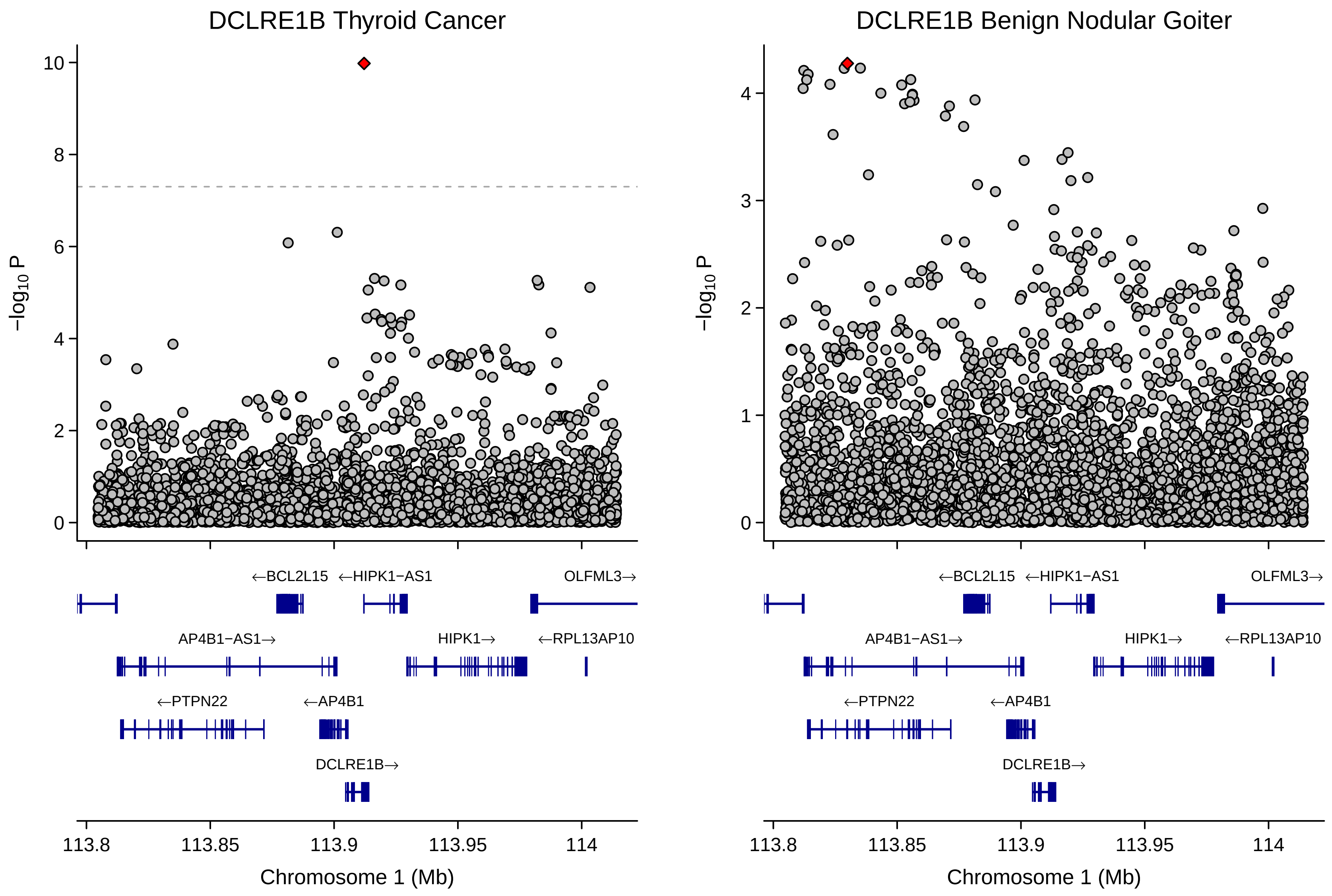
DCLRE1B Thyroid Cancer
DCLRE1B Benign Nodular Goiter
−log10 P
10
8
6
4
2
0
3
1
←BCL2L15
←HIPK1−AS1
OLFML3→
AP4B1−AS1→
HIPK1→
←RPL13AP10
←PTPN22
←AP4B1
DCLRE1B→
113.8
113.85
113.9
113.95
114
Chromosome 1 (Mb)

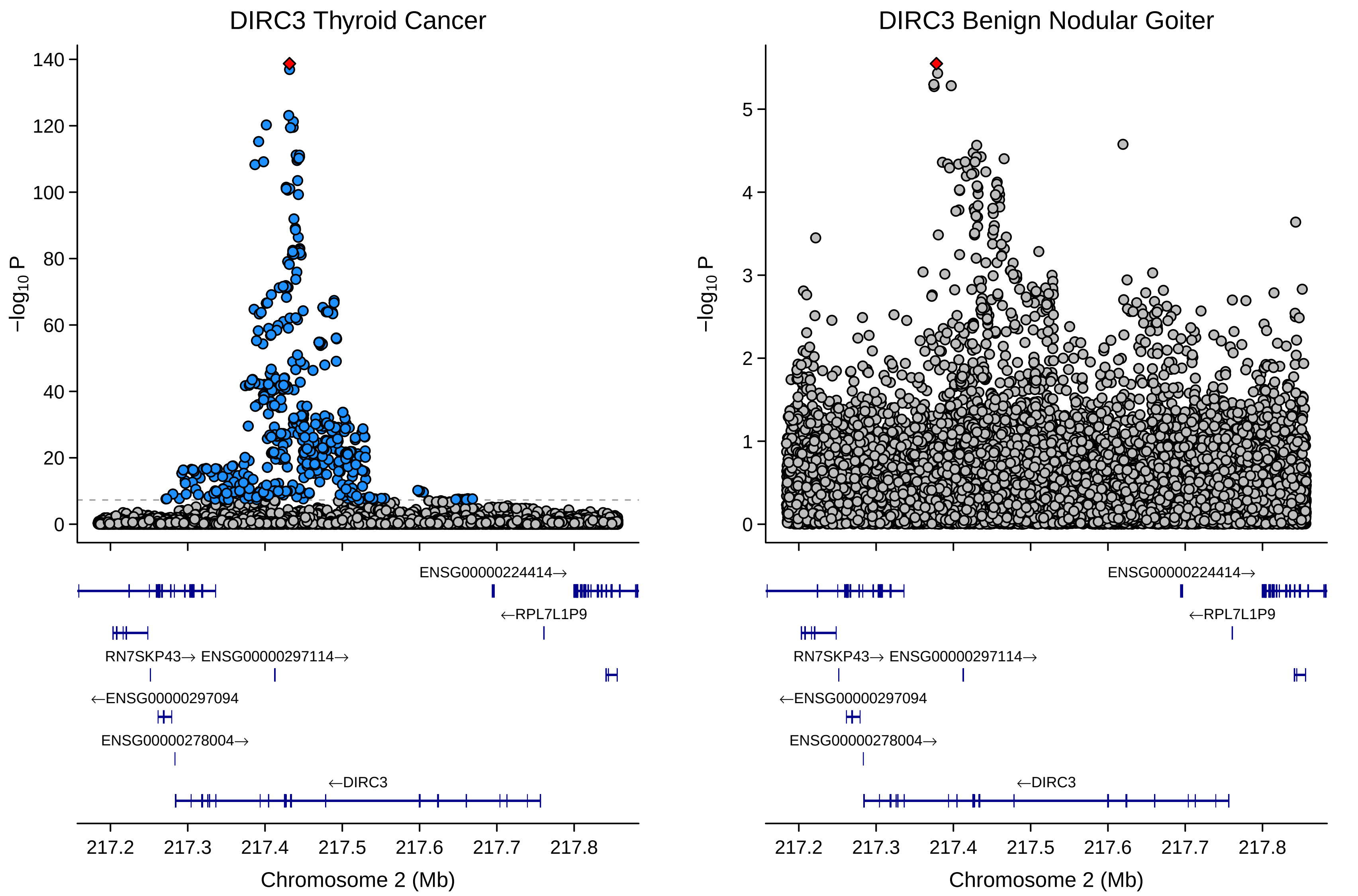
DIRC3 Thyroid Cancer
DIRC3 Benign Nodular Goiter
−log10 P
ENSG00000224414→
←RPL7L1P9
RN7SKP43→ ENSG00000297114→
←ENSG00000297094
ENSG00000278004→
←DIRC3
217.2 217.3 217.4 217.5 217.6 217.7 217.8
Chromosome 2 (Mb)

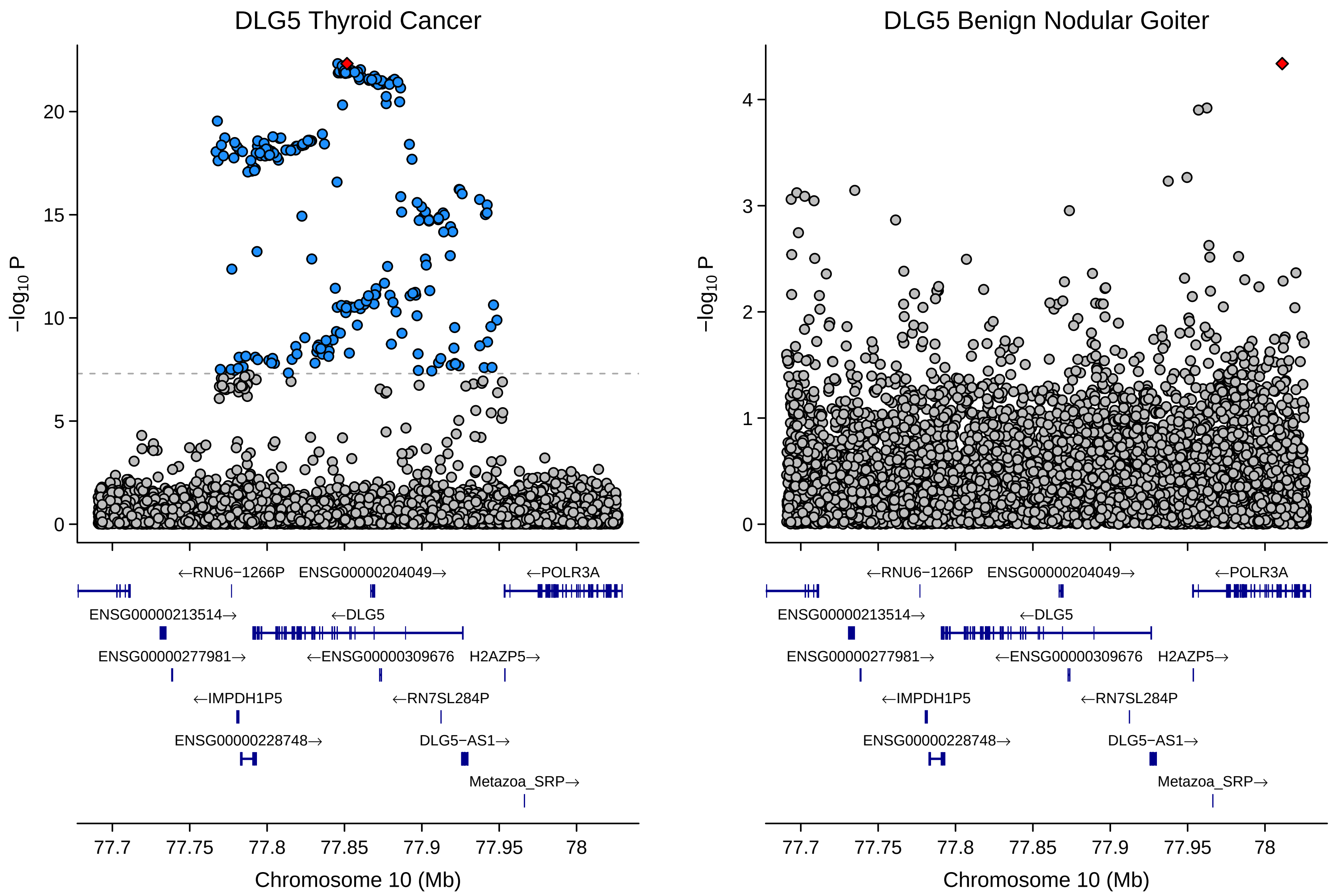
DLG5 Thyroid Cancer
DLG5 Benign Nodular Goiter
−log10 P
Chromosome 10 (Mb)
←RNU6−1266P
ENSG00000204049→
←POLR3A
ENSG00000213514→
←DLG5
ENSG00000277981→
←ENSG00000309676
H2AZP5→
←IMPDH1P5
←RN7SL284P
ENSG00000228748→
DLG5−AS1→
Metazoa_SRP→

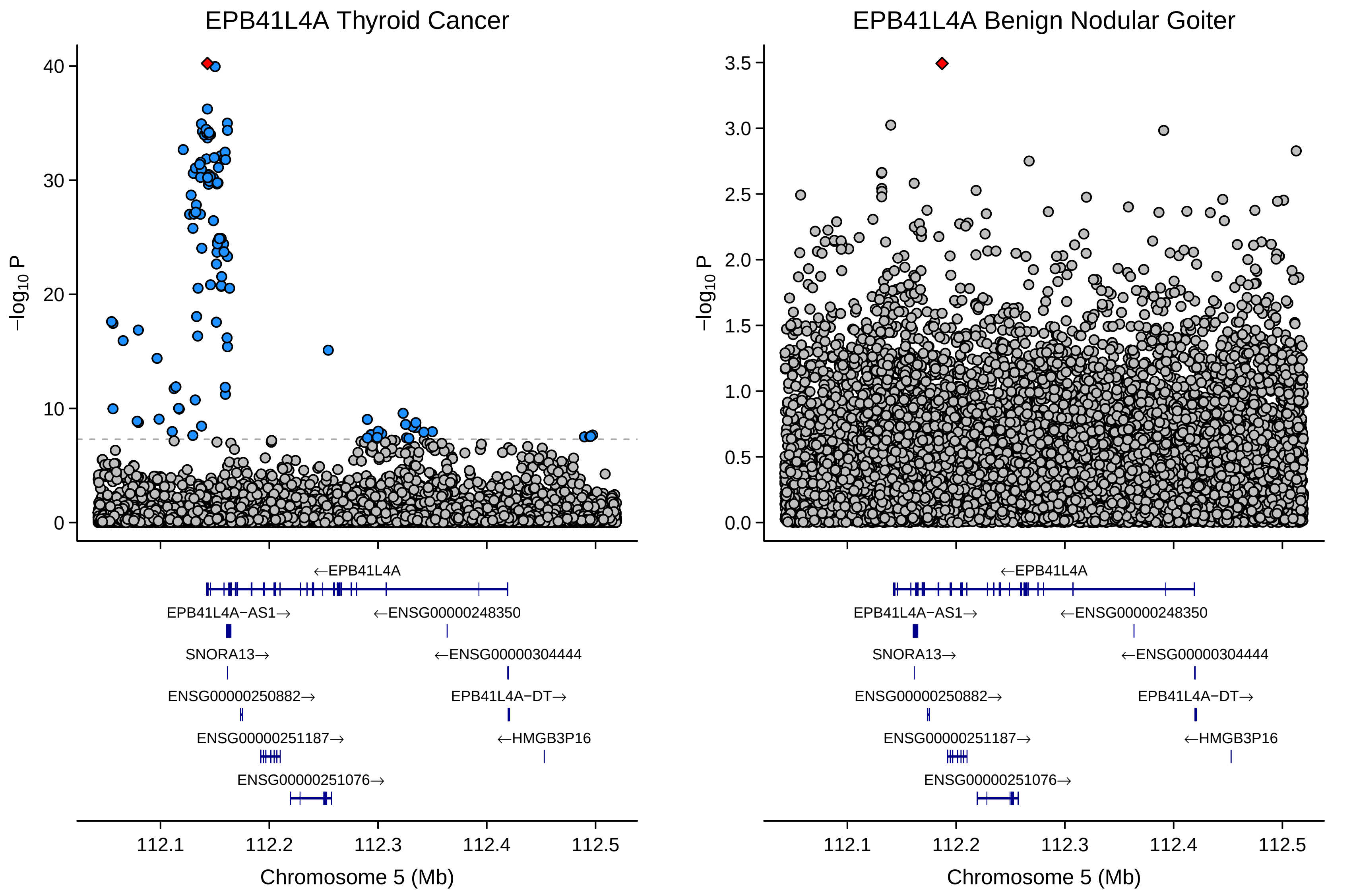
EPB41L4A Thyroid Cancer
EPB41L4A Benign Nodular Goiter
−log10 P
Chromosome 5 (Mb)
112.1
112.2
112.3
112.4
112.5
←EPB41L4A
EPB41L4A−AS1→
←ENSG00000248350
SNORA13→
←ENSG00000304444
ENSG00000250882→
EPB41L4A−DT→
ENSG00000251187→
←HMGB3P16
ENSG00000251076→

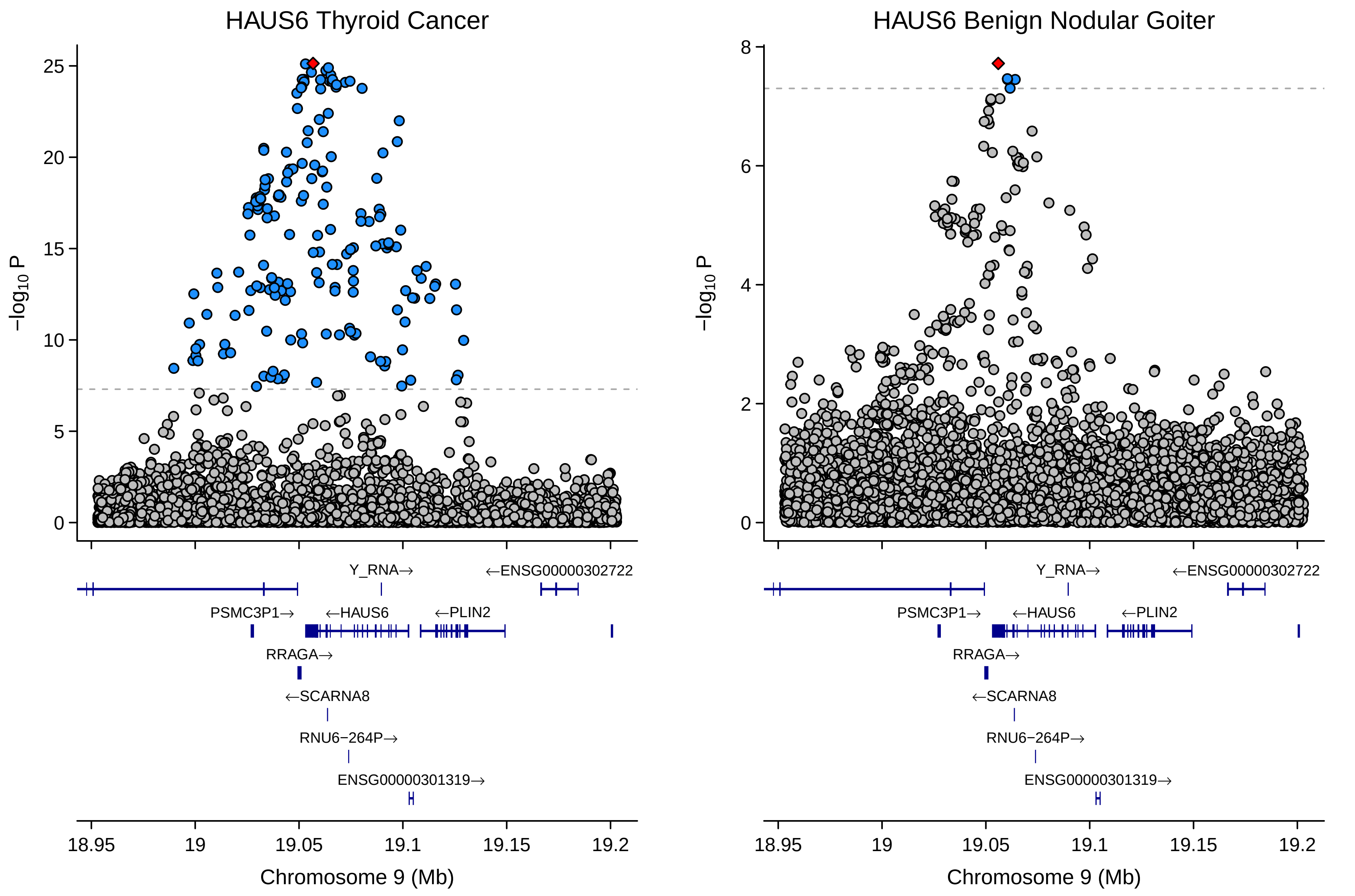

HAUS6 Thyroid Cancer
HAUS6 Benign Nodular Goiter
−log10 P
25
20
15
10
5
0
8
6
4
2
Y_RNA→
←ENSG00000302722
PSMC3P1→
←HAUS6
←PLIN2
RRAGA→
←SCARNA8
RNU6−264P→
ENSG00000301319→
18.95
19
19.05
19.1
19.15
19.2
Chromosome 9 (Mb)

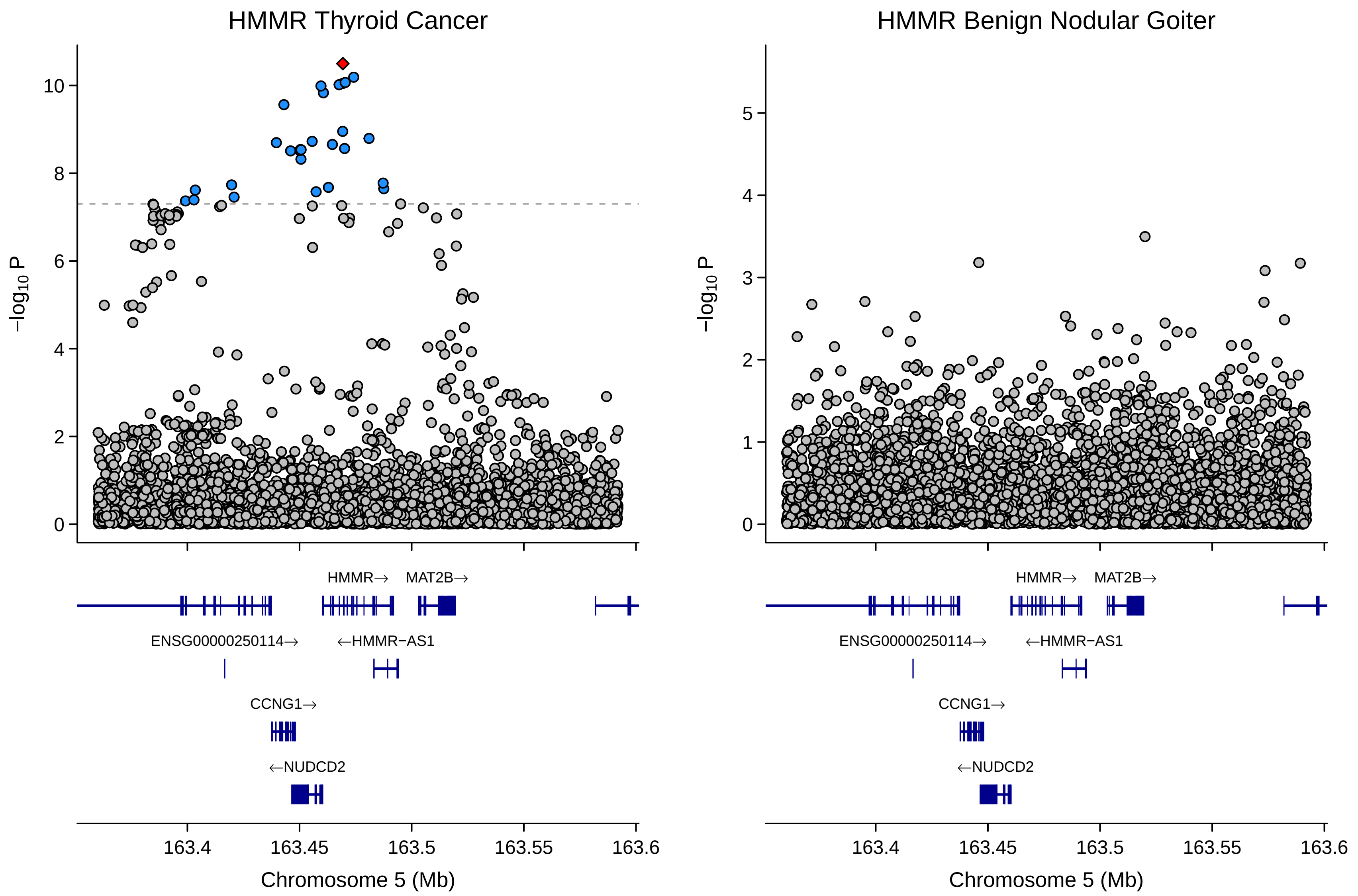
HMMR Thyroid Cancer
HMMR Benign Nodular Goiter
−log10 P
10
8
6
4
2
0
5
3
1
HMMR→
MAT2B→
ENSG00000250114→
←HMMR−AS1
CCNG1→
←NUDCD2
163.4
163.45
163.5
163.55
163.6
Chromosome 5 (Mb)

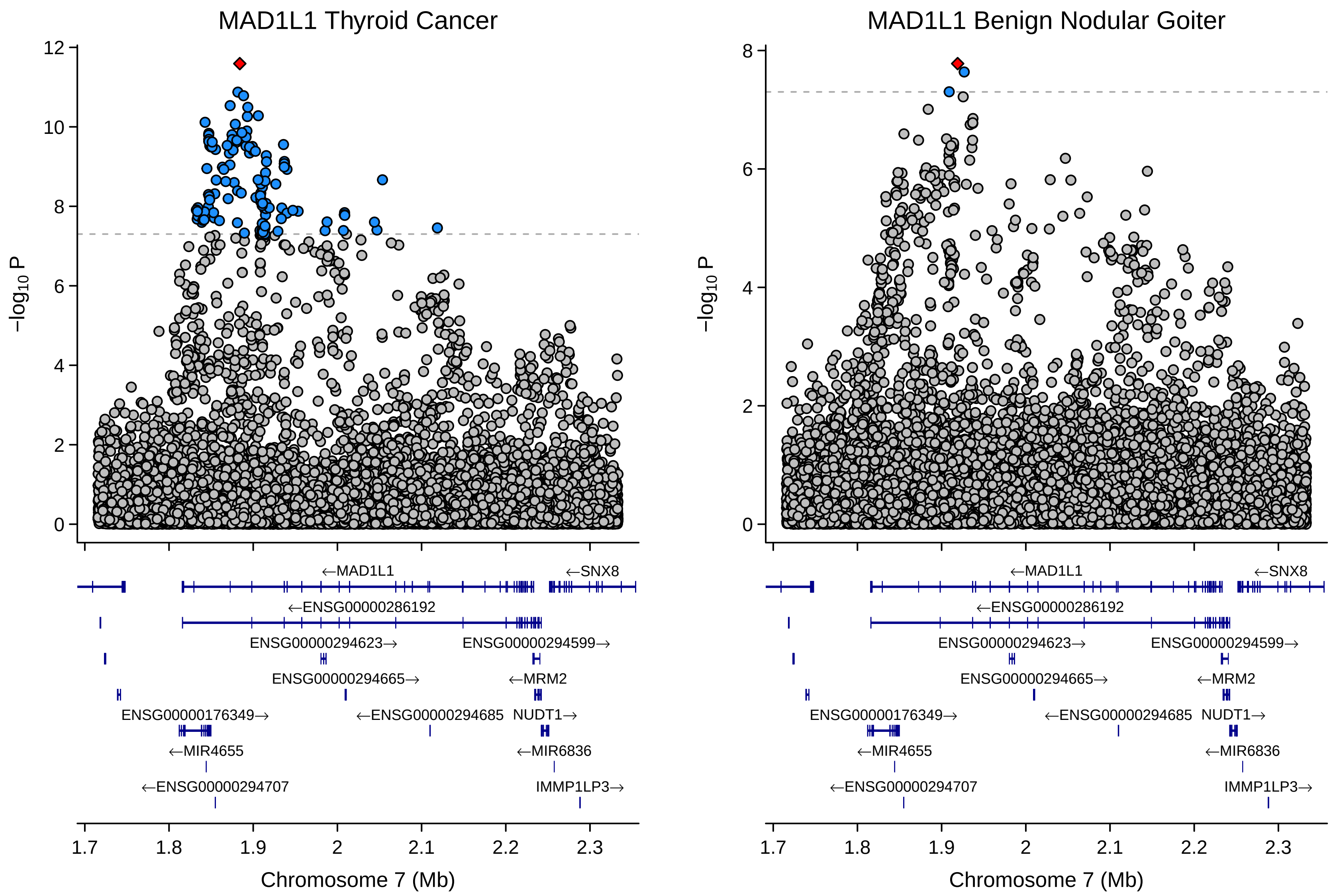

MAD1L1 Thyroid Cancer
MAD1L1 Benign Nodular Goiter
$-\log_{10}$ P
Chromosome 7 (Mb)
←MAD1L1
←SNX8
←ENSG00000286192
ENSG00000294623→
ENSG00000294599→
ENSG00000294665→
←MRM2
ENSG00000176349→
←ENSG00000294685
NUDT1→
←MIR4655
←MIR6836
←ENSG00000294707
IMMP1LP3→

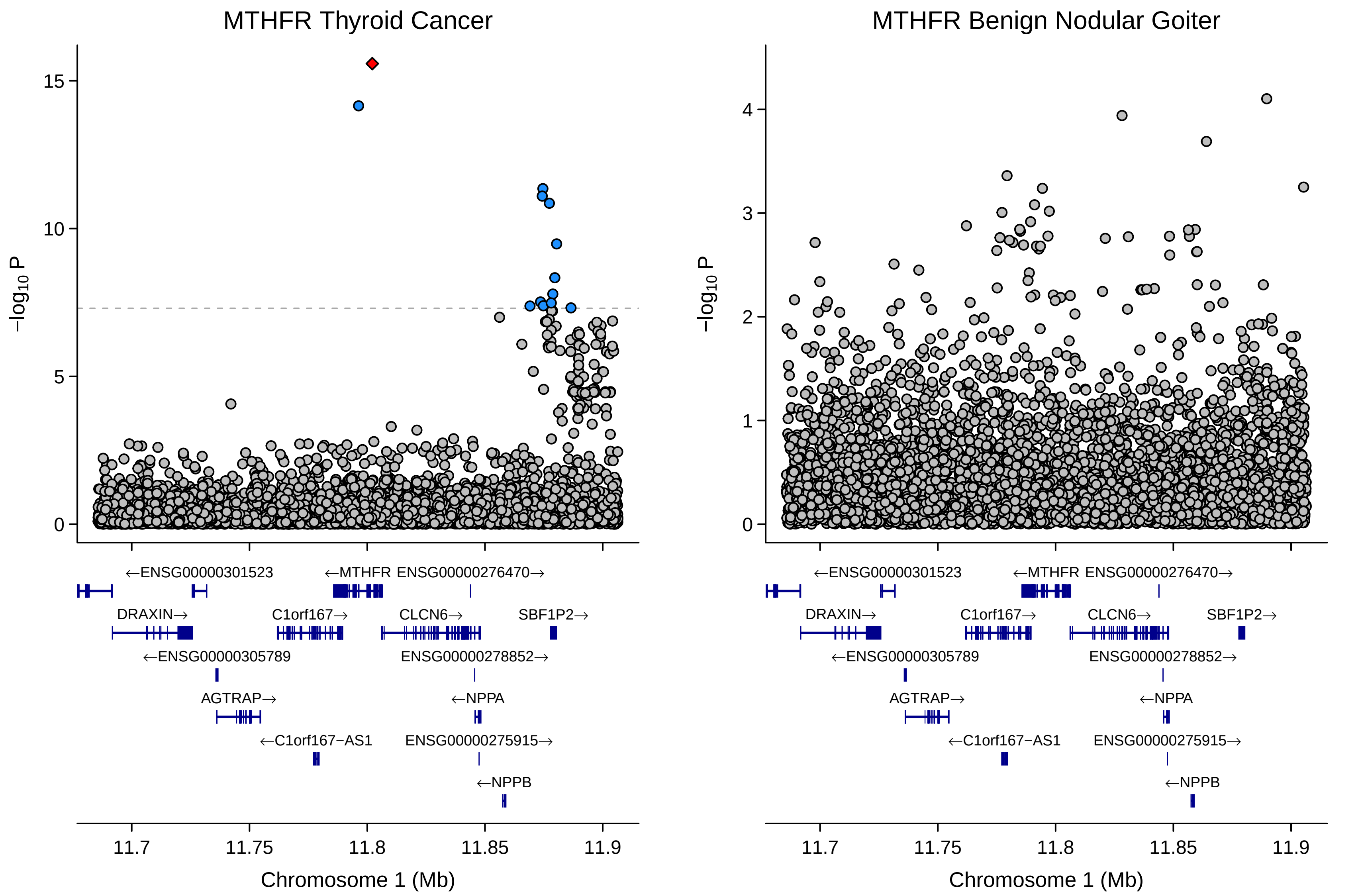
MTHFR Thyroid Cancer
MTHFR Benign Nodular Goiter
−log10 P
15
10
5
0
4
3
2
1
←ENSG00000301523
←MTHFR ENSG00000276470→
DRAXIN→
C1orf167→
CLCN6→
SBF1P2→
←ENSG00000305789
ENSG00000278852→
AGTRAP→
←NPPA
←C1orf167−AS1
ENSG00000275915→
←NPPB
11.7
11.75
11.8
11.85
11.9
Chromosome 1 (Mb)

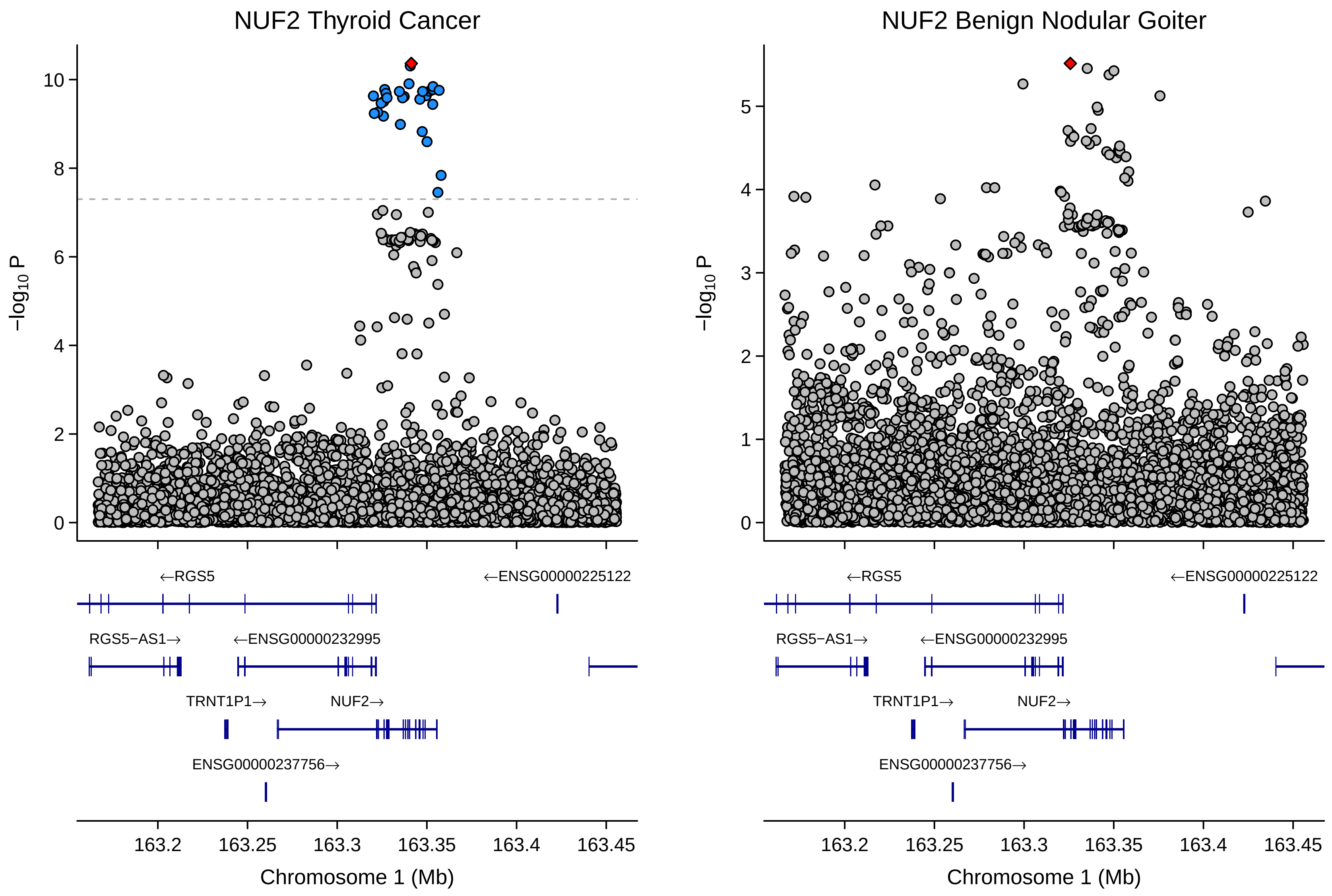

NUF2 Thyroid Cancer
NUF2 Benign Nodular Goiter
−log10 P
←RGS5
←ENSG00000225122
RGS5−AS1→
←ENSG00000232995
TRNT1P1→
NUF2→
ENSG00000237756→
163.2
163.25
163.3
163.35
163.4
163.45
Chromosome 1 (Mb)

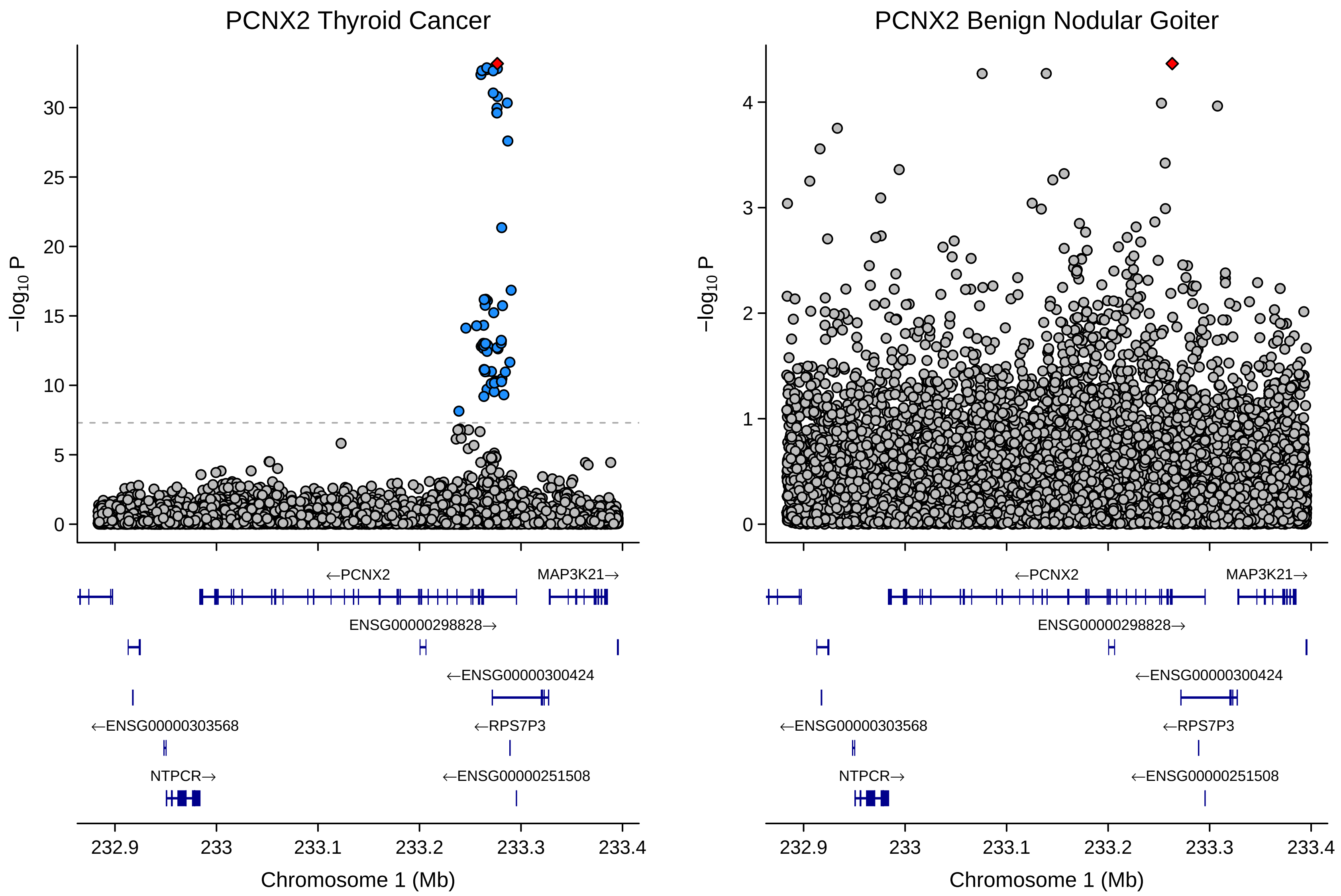
PCNX2 Thyroid Cancer
PCNX2 Benign Nodular Goiter
−log10 P
Chromosome 1 (Mb)
←PCNX2
MAP3K21→
ENSG00000298828→
←ENSG00000300424
←ENSG00000303568
←RPS7P3
NTPCR→
←ENSG00000251508
232.9
233
233.1
233.2
233.3
233.4

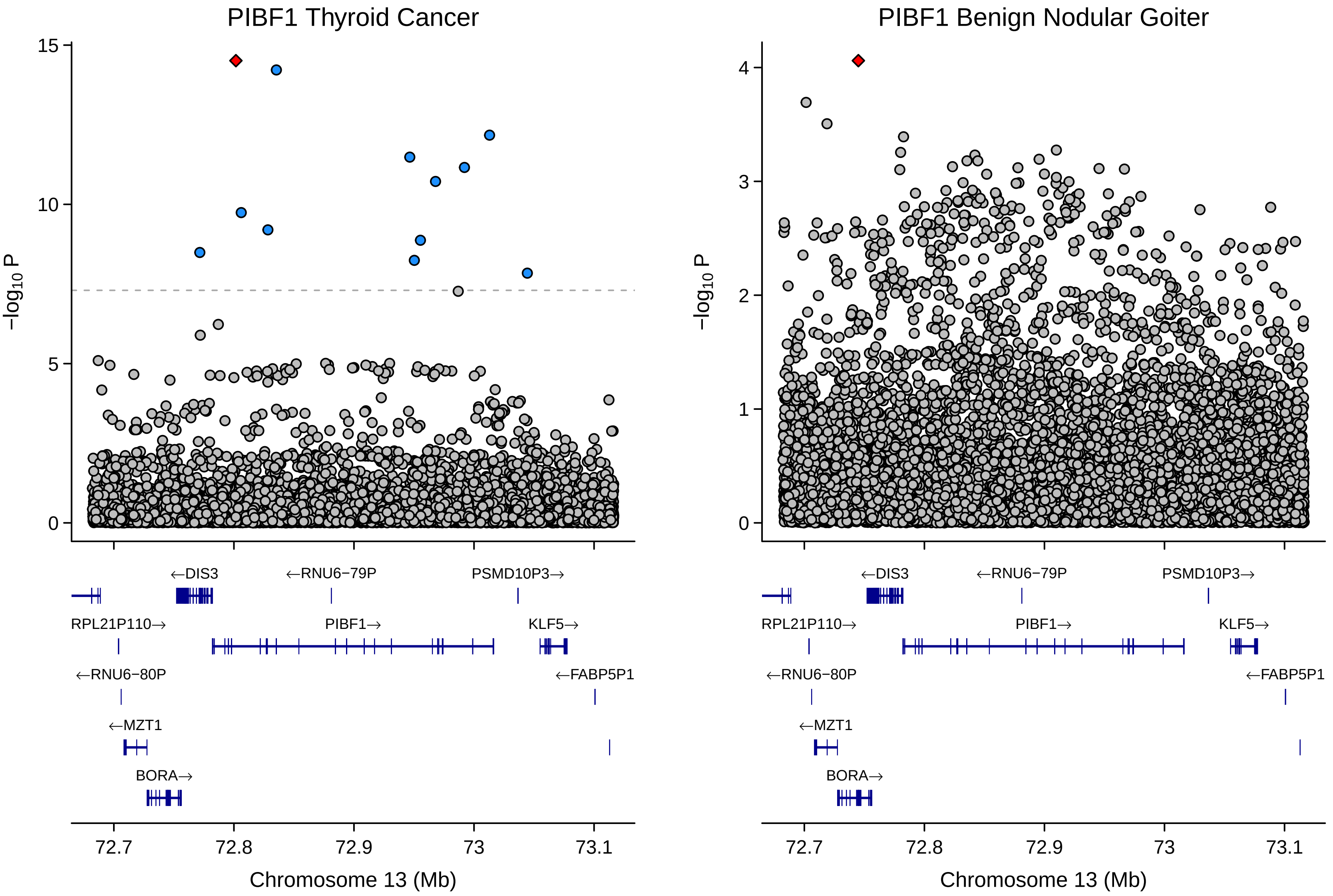
PIBF1 Thyroid Cancer
PIBF1 Benign Nodular Goiter
−log10 P
15
10
5
0
4
3
2
1
←DIS3
←RNU6−79P
PSMD10P3→
RPL21P110→
PIBF1→
KLF5→
←RNU6−80P
←FABP5P1
←MZT1
BORA→
72.7
72.8
72.9
73
73.1
Chromosome 13 (Mb)

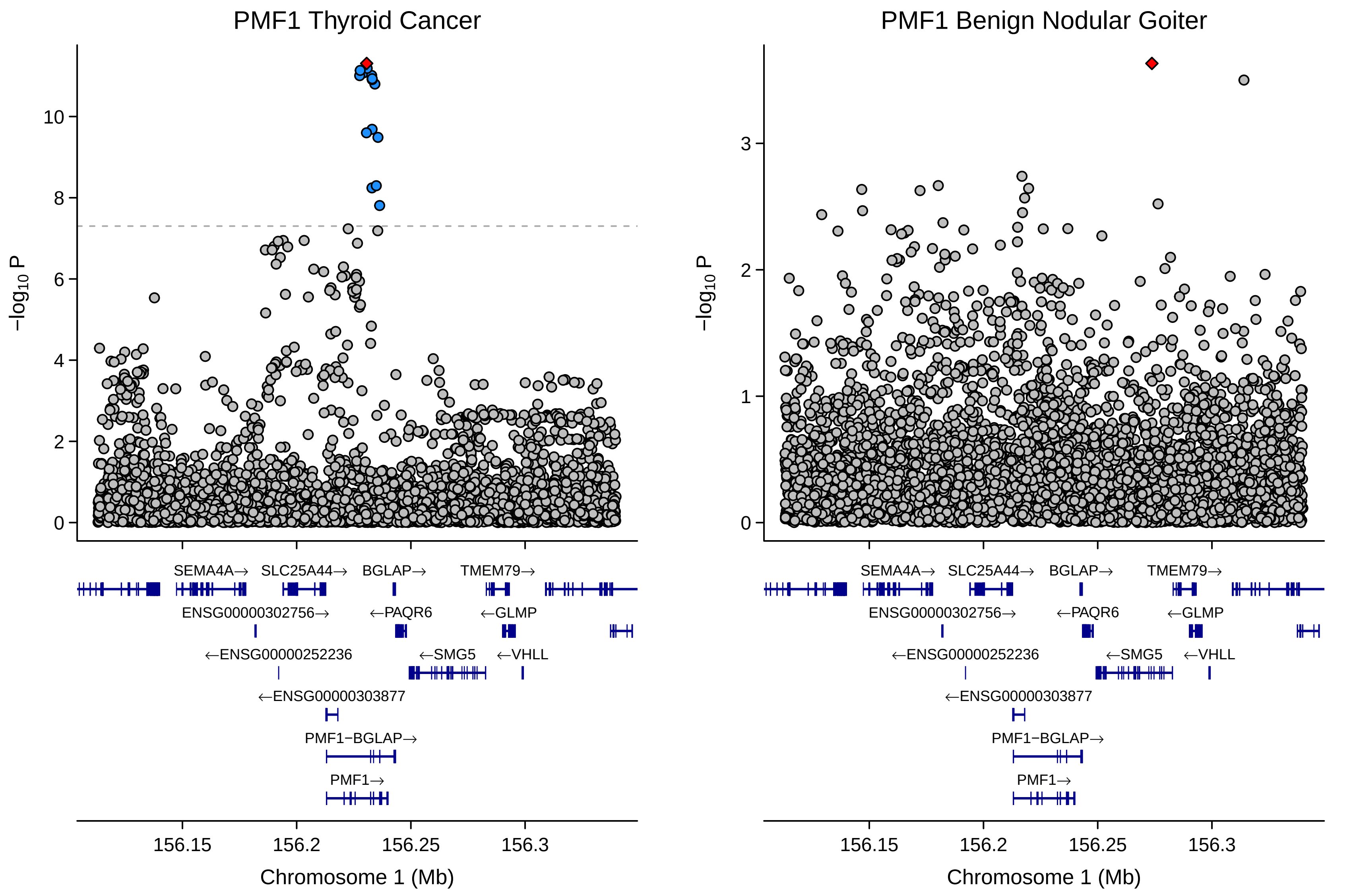
PMF1 Thyroid Cancer
PMF1 Benign Nodular Goiter
−log10 P
Chromosome 1 (Mb)
156.15
156.2
156.25
156.3
SEMA4A→
SLC25A44→
BGLAP→
TMEM79→
ENSG00000302756→
←PAQR6
←GLMP
←ENSG00000252236
←SMG5
←VHLL
←ENSG00000303877
PMF1−BGLAP→
PMF1→

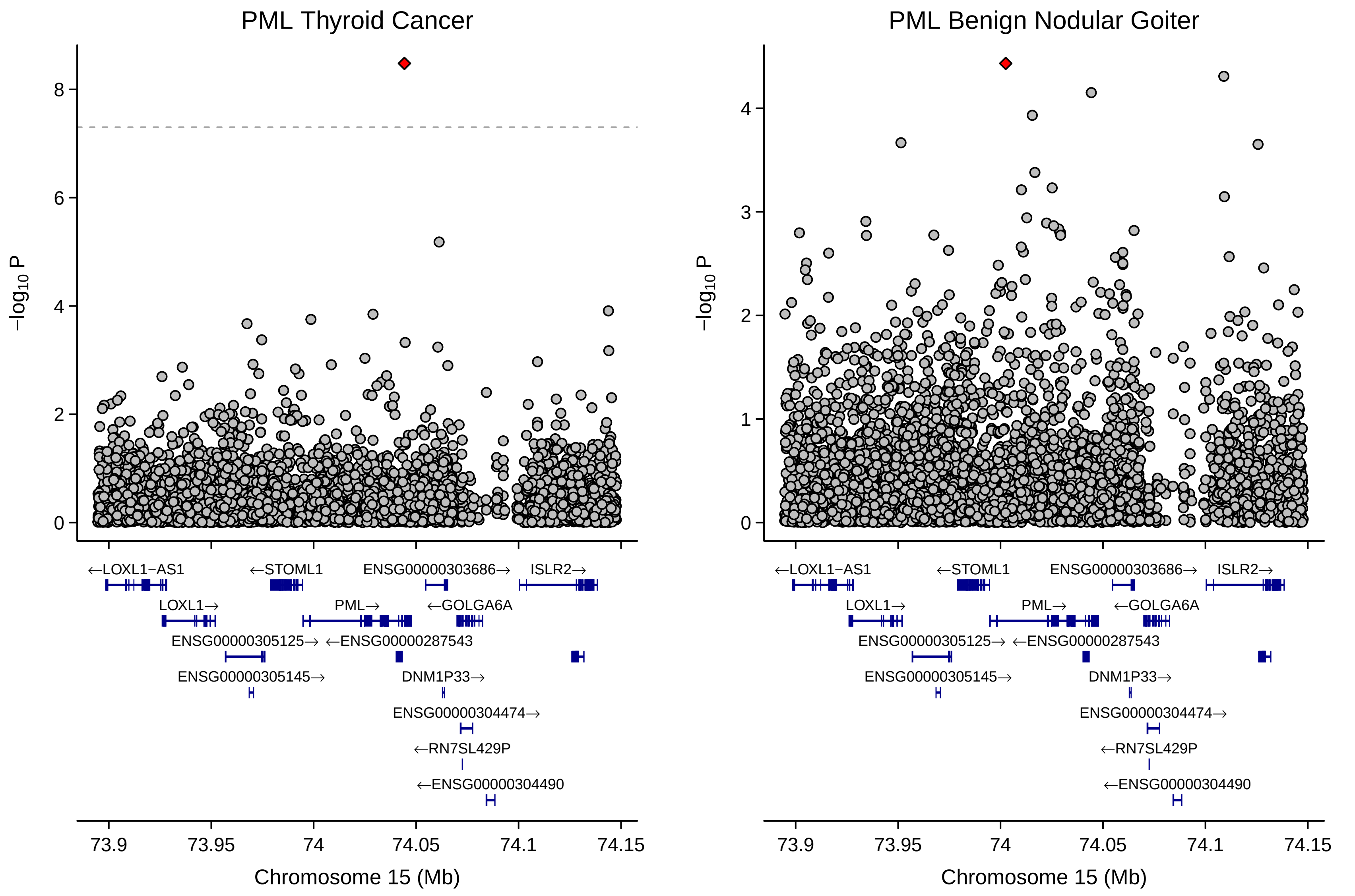
PML Thyroid Cancer
PML Benign Nodular Goiter
−log10 P
←LOXL1−AS1
←STOML1
ENSG00000303686→
ISLR2→
LOXL1→
PML→
←GOLGA6A
ENSG00000305125→
←ENSG00000287543
ENSG00000305145→
DNM1P33→
ENSG00000304474→
←RN7SL429P
←ENSG00000304490
73.9
73.95
74
74.05
74.1
74.15
Chromosome 15 (Mb)

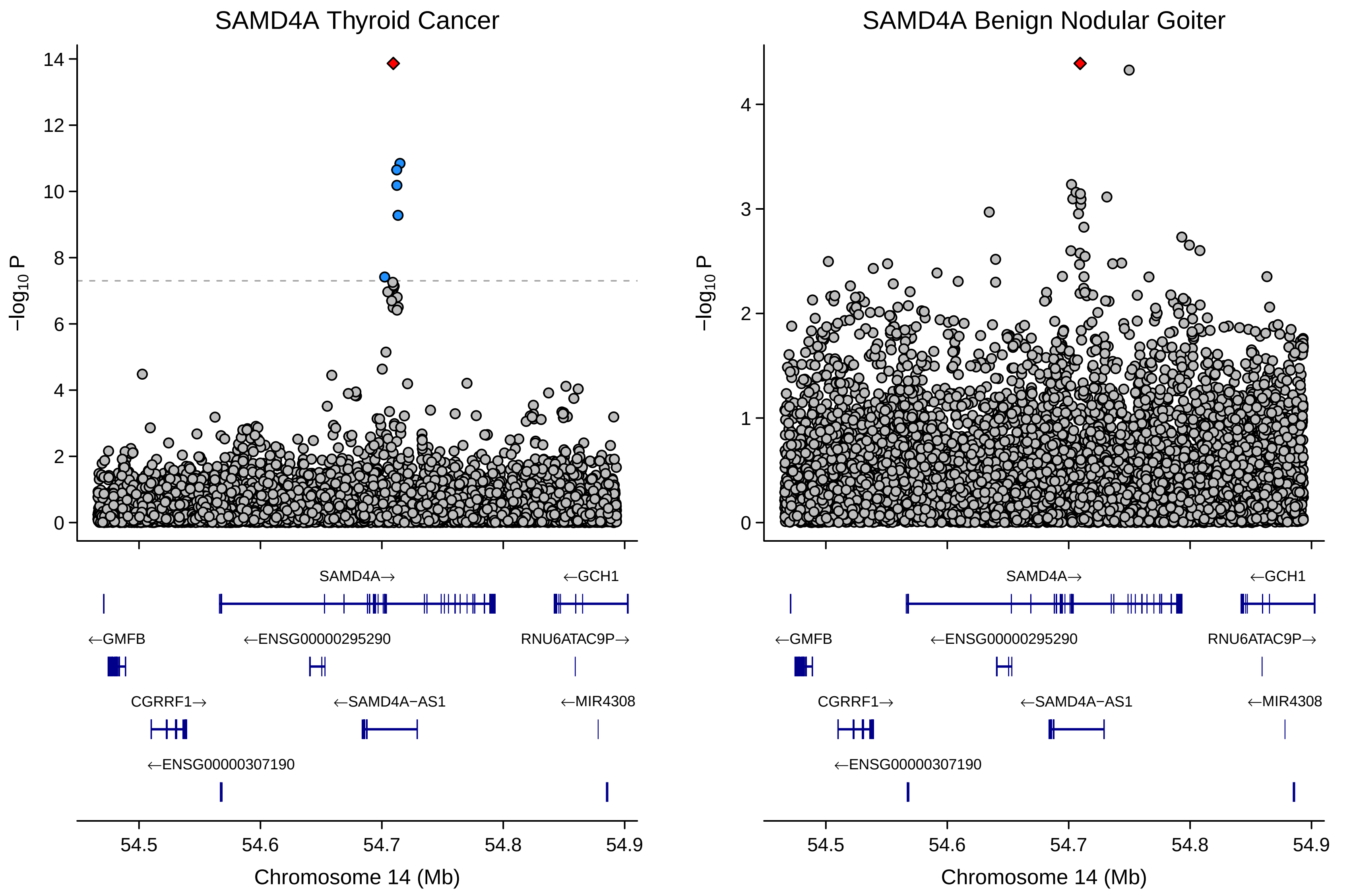

SAMD4A Thyroid Cancer
SAMD4A Benign Nodular Goiter
−log10 P
0
2
4
6
8
10
12
14
1
3
SAMD4A→
←GCH1
←GMFB
←ENSG00000295290
RNU6ATAC9P→
CGRRF1→
←SAMD4A−AS1
←MIR4308
←ENSG00000307190
54.5
54.6
54.7
54.8
54.9
Chromosome 14 (Mb)

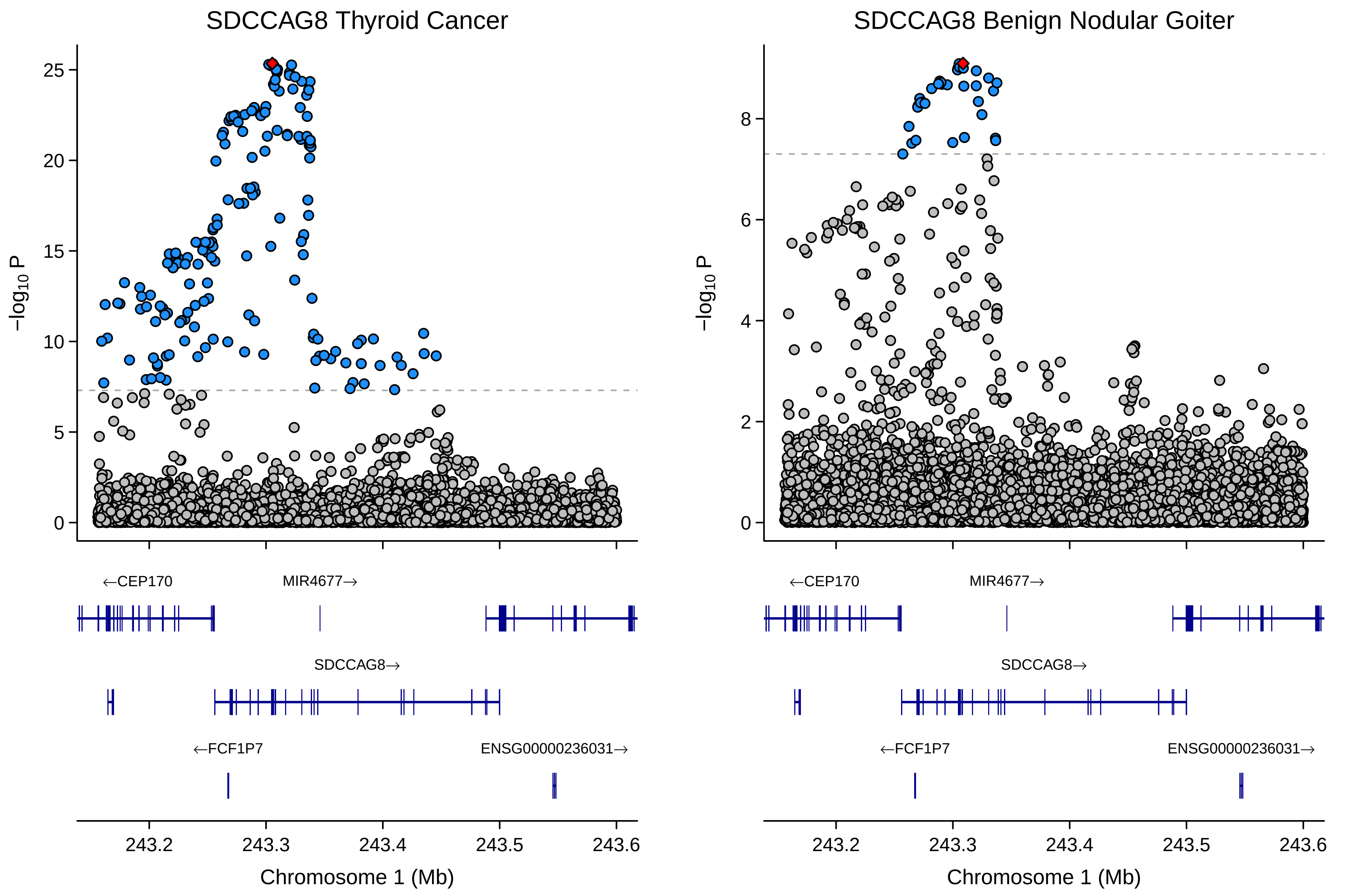
SDCCAG8 Thyroid Cancer
SDCCAG8 Benign Nodular Goiter
−log10 P
25
20
15
10
5
0
8
6
4
2
←CEP170
MIR4677→
SDCCAG8→
←FCF1P7
ENSG00000236031→
243.2
243.3
243.4
243.5
243.6
Chromosome 1 (Mb)

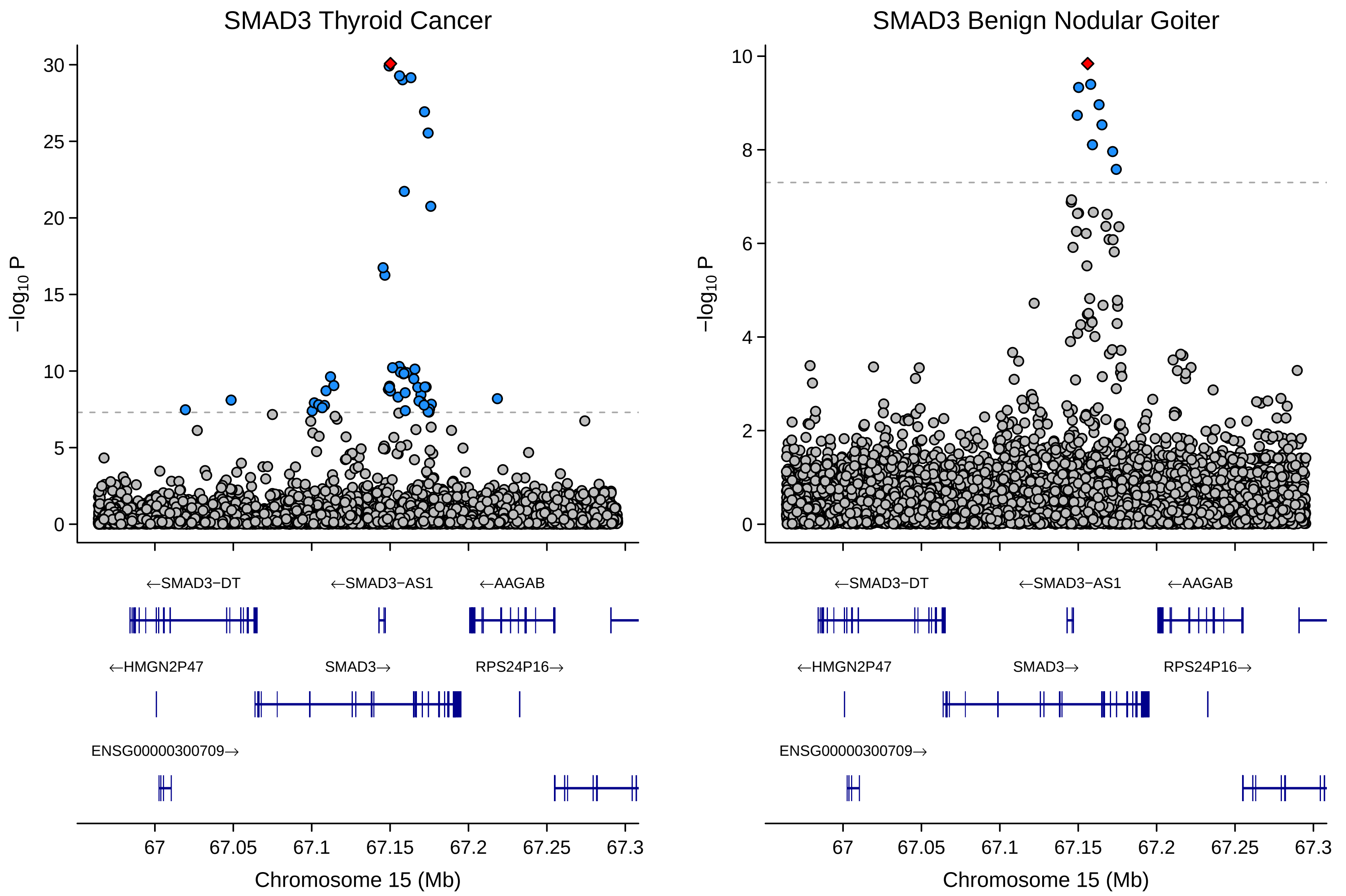
SMAD3 Thyroid Cancer
SMAD3 Benign Nodular Goiter
−log10 P
30
25
20
15
10
5
0
8
6
4
2
←SMAD3−DT
←SMAD3−AS1
←AAGAB
←HMGN2P47
SMAD3→
RPS24P16→
ENSG00000300709→
67
67.05
67.1
67.15
67.2
67.25
67.3
Chromosome 15 (Mb)

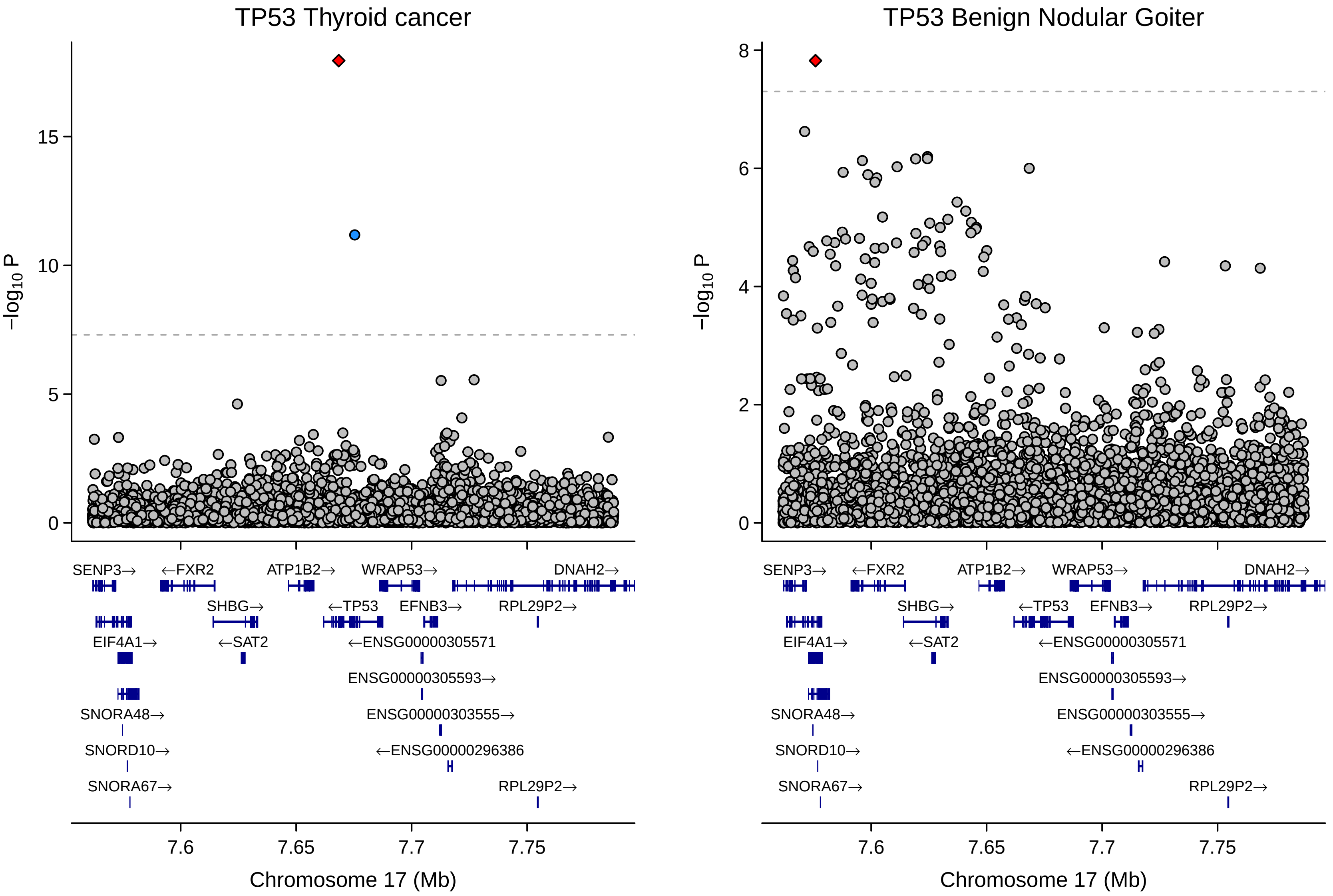
TP53 Thyroid cancer
TP53 Benign Nodular Goiter
−log10 P
0
5
10
15
2
4
6
8
SENP3→
←FXR2
ATP1B2→
WRAP53→
DNAH2→
SHBG→
←TP53
EFNB3→
RPL29P2→
EIF4A1→
←SAT2
←ENSG00000305571
ENSG00000305593→
SNORA48→
ENSG00000303555→
SNORD10→
←ENSG00000296386
SNORA67→
7.6
7.65
7.7
7.75
Chromosome 17 (Mb)

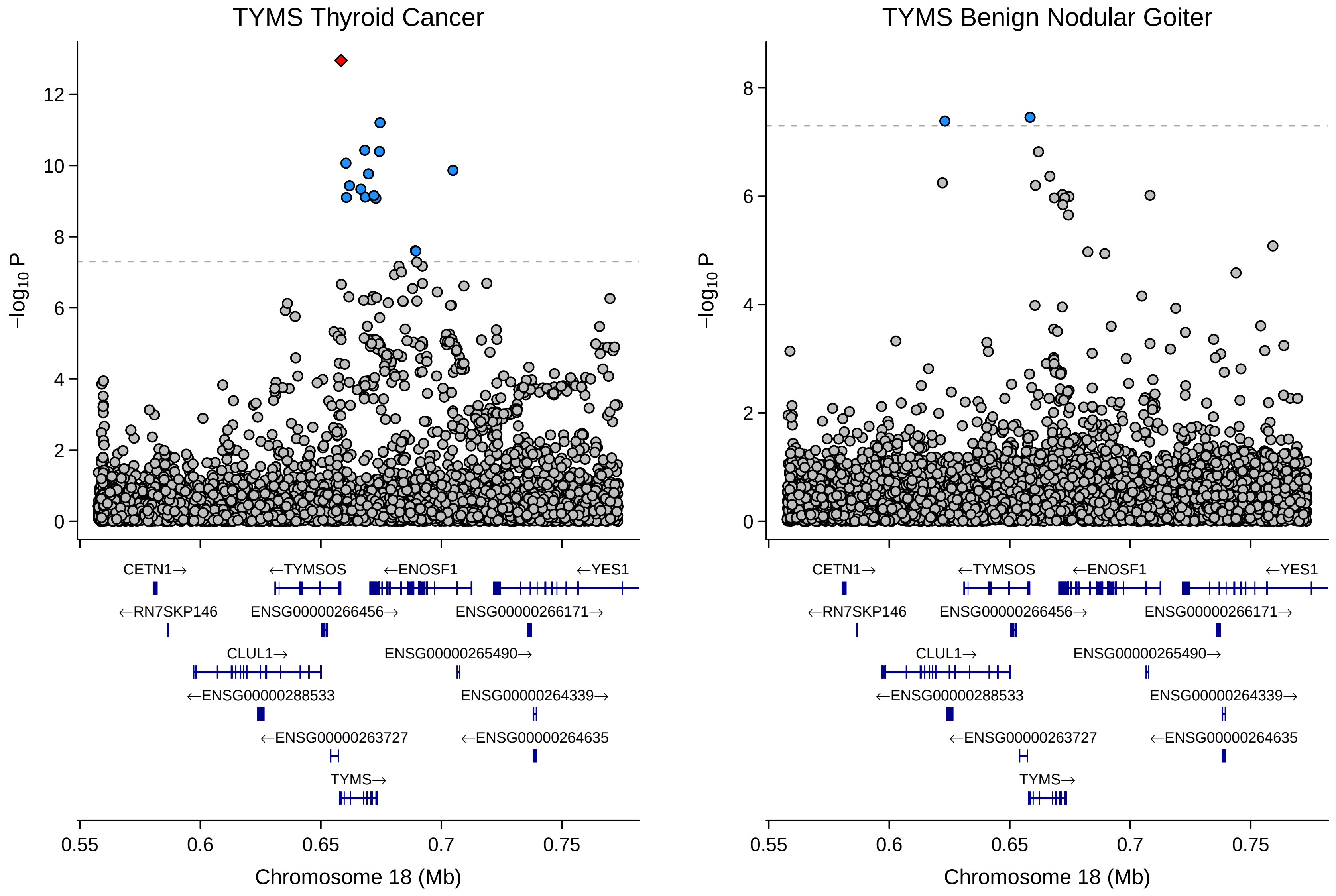
TYMS Thyroid Cancer
TYMS Benign Nodular Goiter
−log10 P
Chromosome 18 (Mb)
0.55
0.6
0.65
0.7
0.75
CETN1→
←TYMSOS
←ENOSF1
←YES1
←RN7SKP146
ENSG00000266456→
ENSG00000266171→
CLUL1→
ENSG00000265490→
←ENSG00000288533
ENSG00000264339→
←ENSG00000263727
←ENSG00000264635
TYMS→

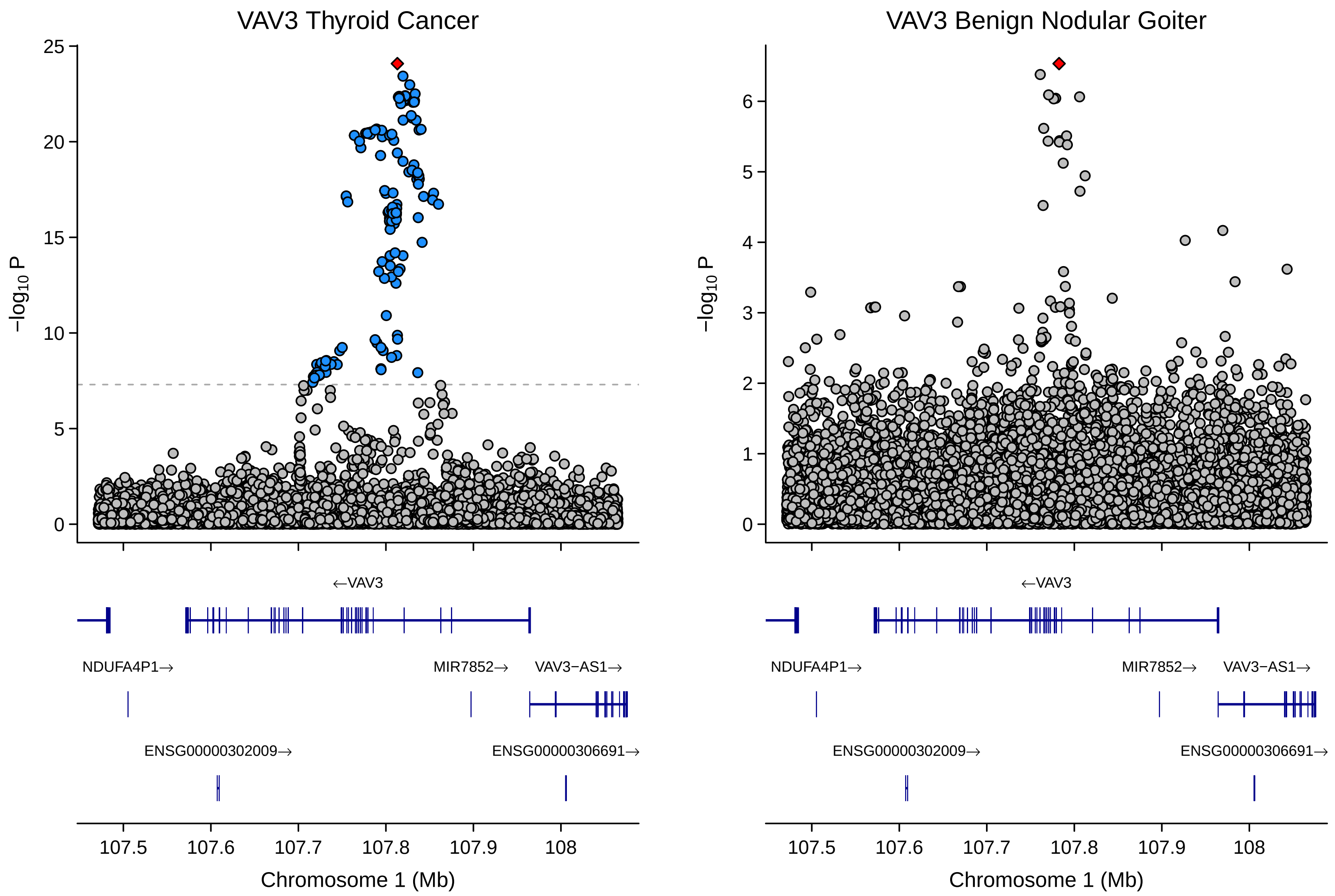
VAV3 Thyroid Cancer
VAV3 Benign Nodular Goiter
−log10 P
←VAV3
NDUFA4P1→
MIR7852→
VAV3−AS1→
ENSG00000302009→
ENSG00000306691→
107.5
107.6
107.7
107.8
107.9
108
Chromosome 1 (Mb)

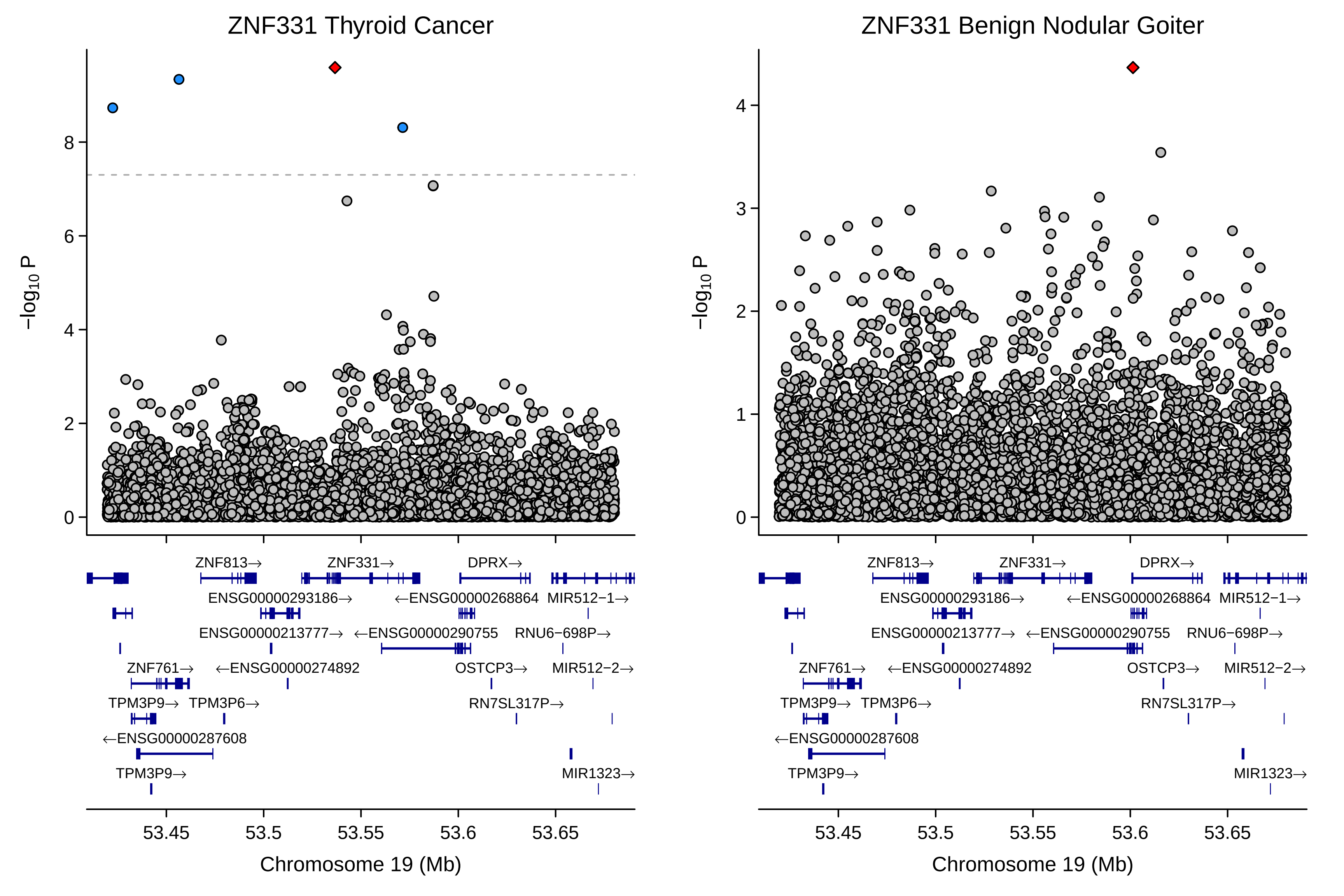
ZNF331 Thyroid Cancer
ZNF331 Benign Nodular Goiter
−log10 P
Chromosome 19 (Mb)
53.45
53.5
53.55
53.6
53.65
ZNF813→
ZNF331→
DPRX→
ENSG00000293186→
←ENSG00000268864
MIR512−1→
ENSG00000213777→
←ENSG00000290755
RNU6−698P→
ZNF761→
←ENSG00000274892
OSTCP3→
MIR512−2→
TPM3P9→
TPM3P6→
RN7SL317P→
←ENSG00000287608
MIR1323→

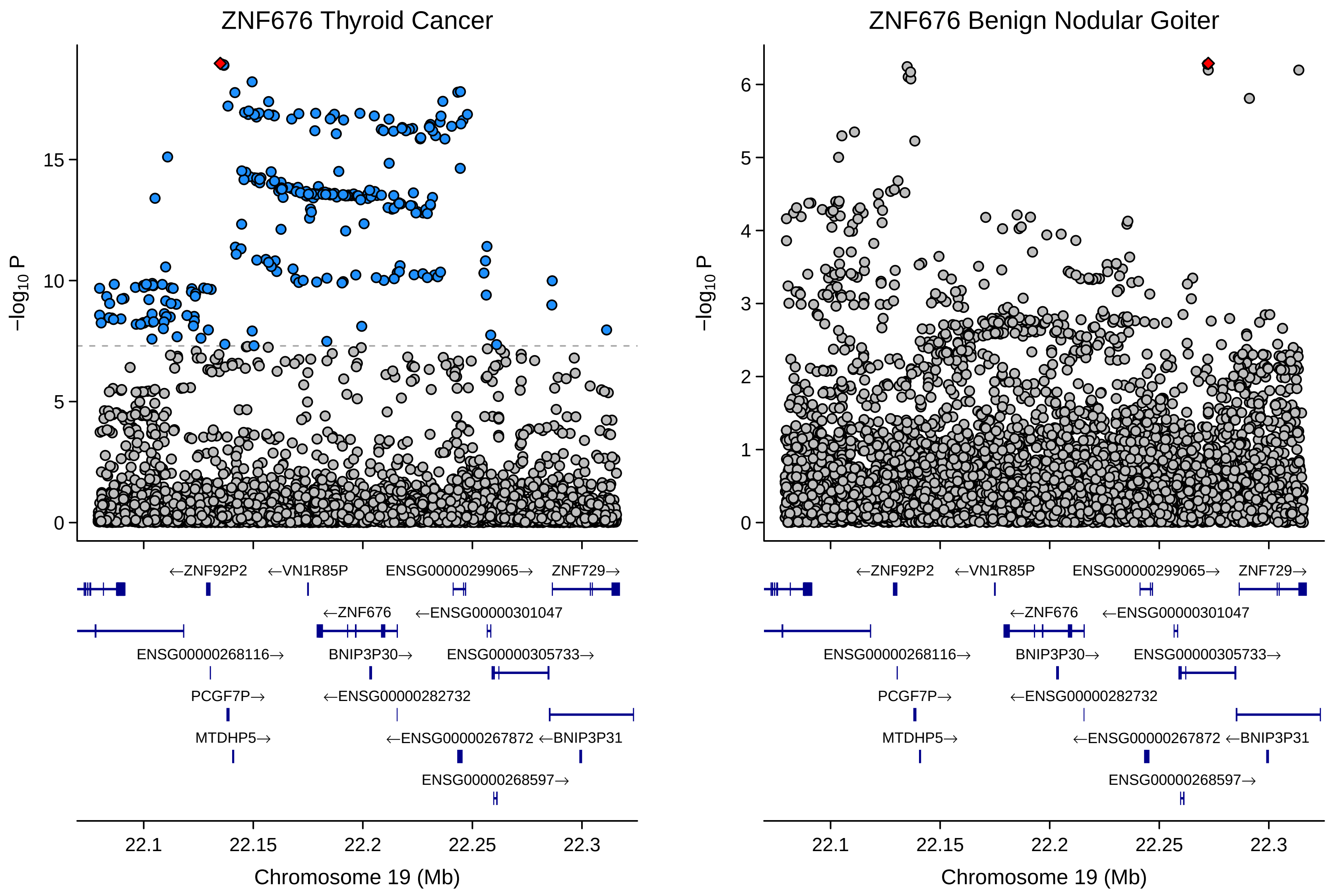
ZNF676 Thyroid Cancer
ZNF676 Benign Nodular Goiter
$-log_{10}$ P
0
5
10
15
0
1
2
3
4
5
6
←ZNF92P2
←VN1R85P
ENSG00000299065→
ZNF729→
←ZNF676
←ENSG00000301047
ENSG00000268116→
BNIP3P30→
ENSG00000305733→
PCGF7P→
←ENSG00000282732
MTDHP5→
←ENSG00000267872
←BNIP3P31
ENSG00000268597→
22.1
22.15
22.2
22.25
22.3
Chromosome 19 (Mb)
