## Supplementary Figure 1.2 for "Global multi-ancestry genetic study elucidates genes and biological pathways associated with thyroid cancer and benign thyroid diseases"

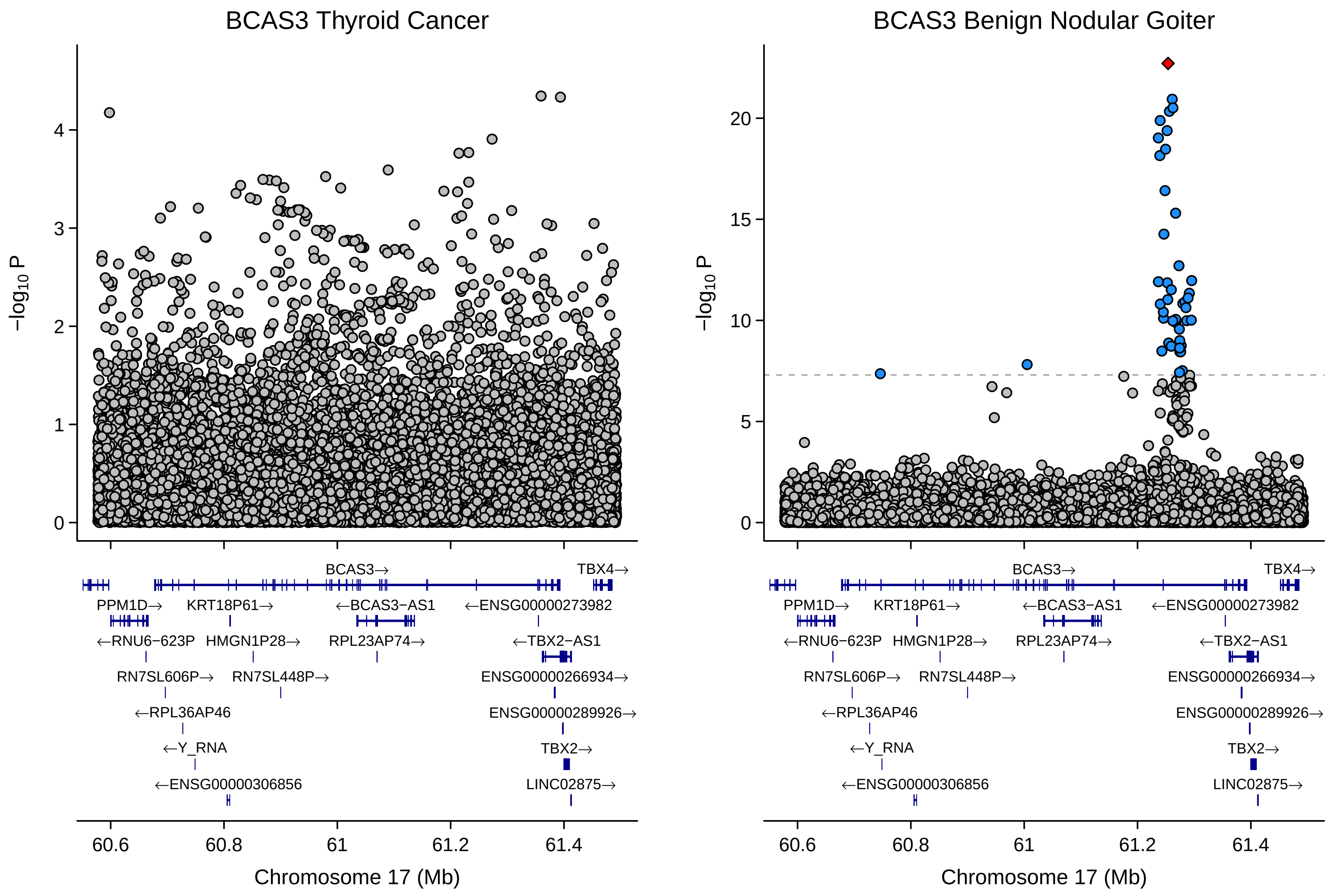

BCAS3 Thyroid Cancer
BCAS3 Benign Nodular Goiter
−log10 P
0
1
2
3
4
5
10
15
20
BCAS3→
TBX4→
PPM1D→
KRT18P61→
←BCAS3−AS1
←ENSG00000273982
←RNU6−623P
HMGN1P28→
RPL23AP74→
←TBX2−AS1
RN7SL606P→
RN7SL448P→
ENSG00000266934→
←RPL36AP46
ENSG00000289926→
←Y_RNA
TBX2→
←ENSG00000306856
LINC02875→
60.6
60.8
61
61.2
61.4
Chromosome 17 (Mb)

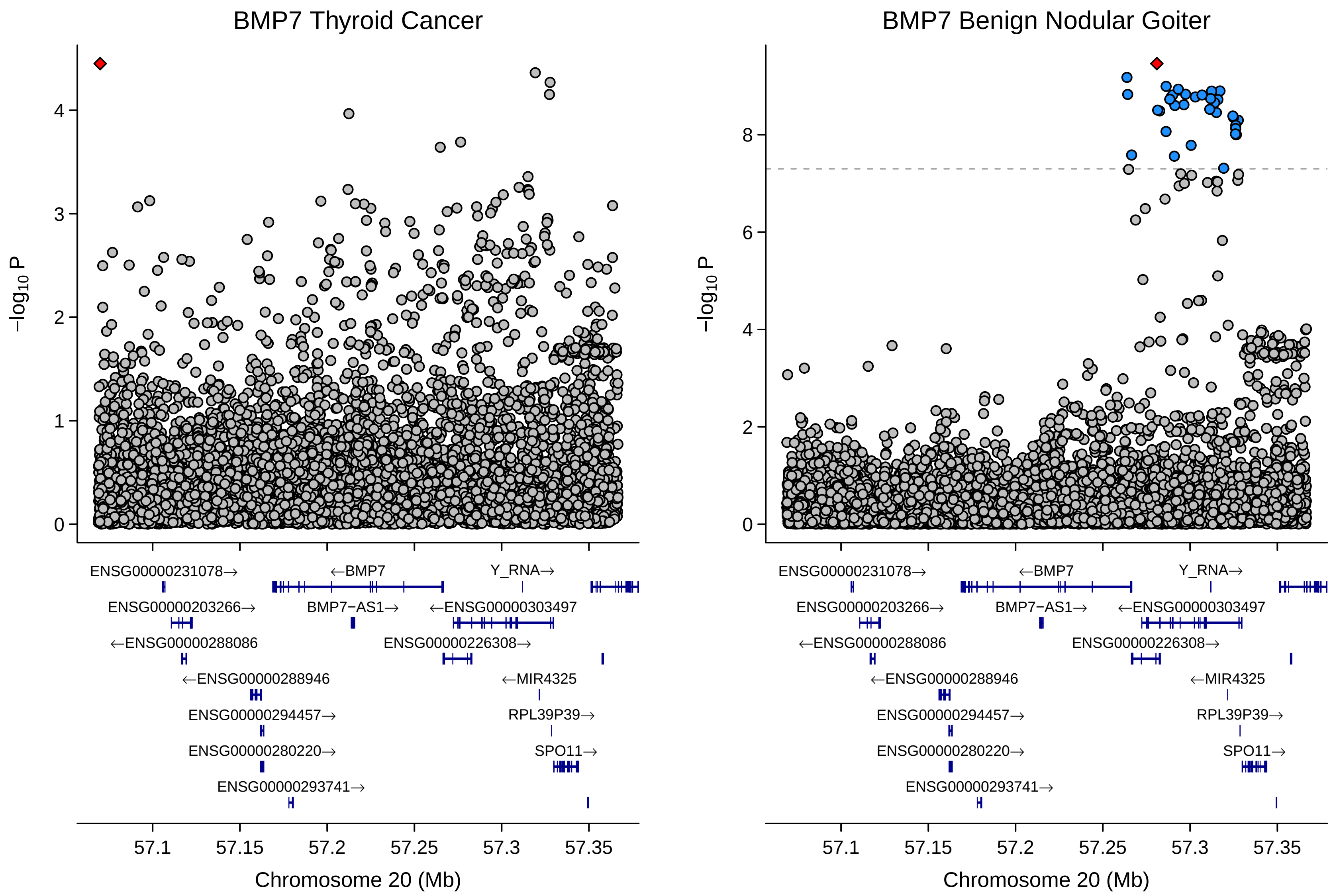
BMP7 Thyroid Cancer
BMP7 Benign Nodular Goiter
−log10 P
0
1
2
3
4
6
8
ENSG00000231078→
←BMP7
Y_RNA→
ENSG00000203266→
BMP7−AS1→
←ENSG00000303497
←ENSG00000288086
ENSG00000226308→
←ENSG00000288946
←MIR4325
ENSG00000294457→
RPL39P39→
ENSG00000280220→
SPO11→
ENSG00000293741→
57.1
57.15
57.2
57.25
57.3
57.35
Chromosome 20 (Mb)

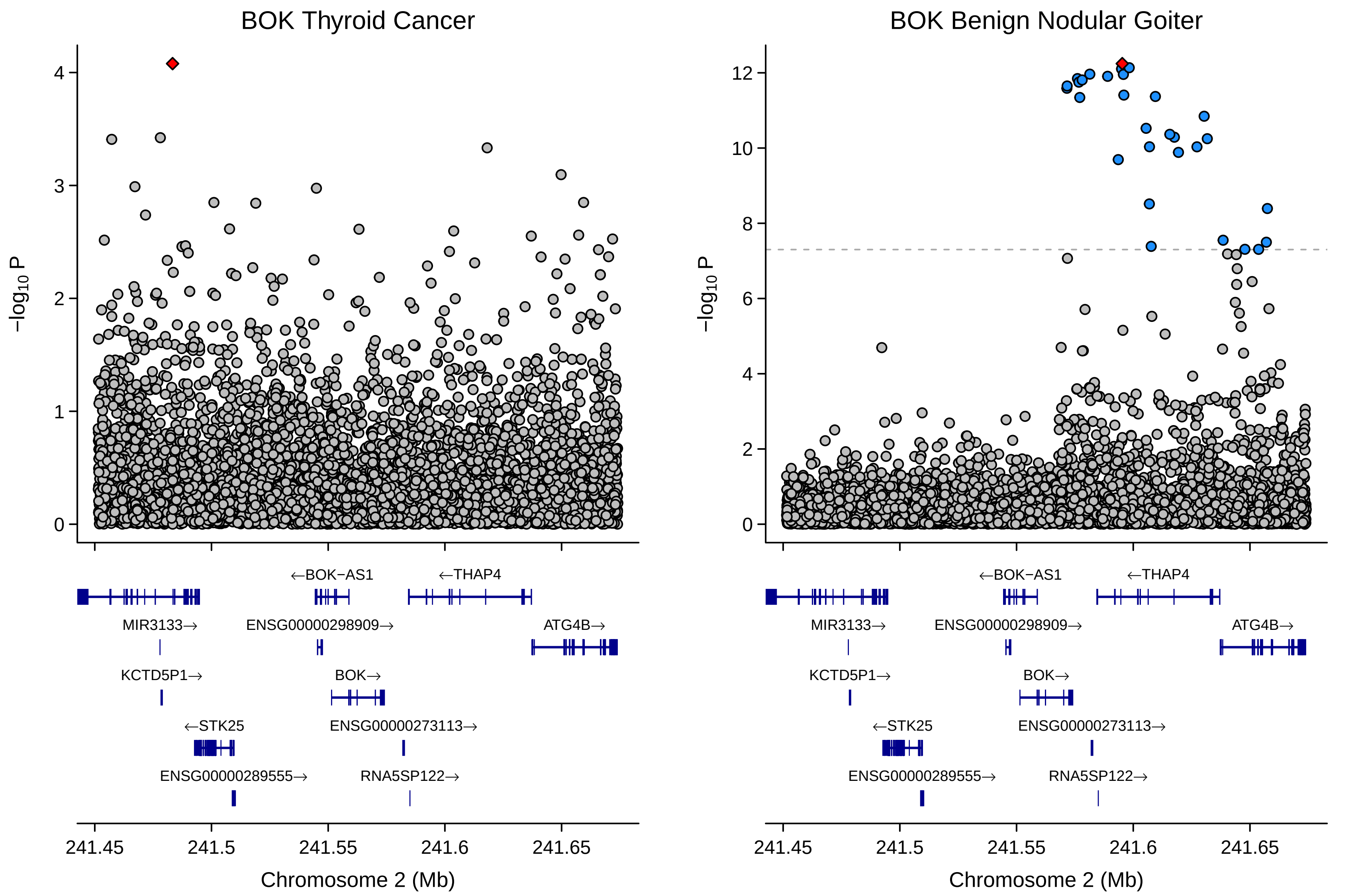
BOK Thyroid Cancer
BOK Benign Nodular Goiter
−log10 P
Chromosome 2 (Mb)
241.45
241.5
241.55
241.6
241.65
←BOK−AS1
←THAP4
MIR3133→
ENSG00000298909→
ATG4B→
KCTD5P1→
BOK→
←STK25
ENSG00000273113→
ENSG00000289555→
RNA5SP122→

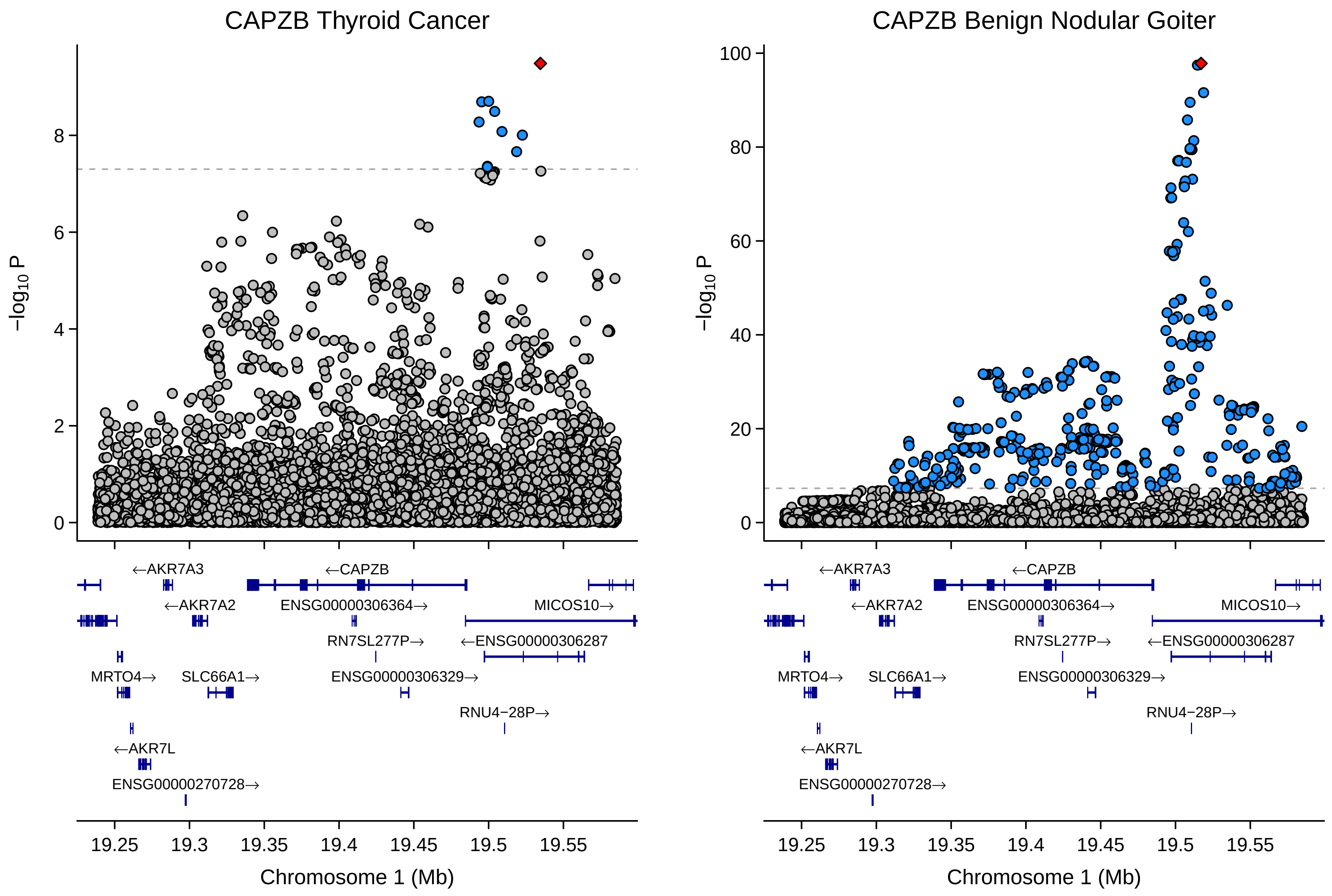
CAPZB Thyroid Cancer
CAPZB Benign Nodular Goiter
−log10 P
0
2
4
6
8
20
40
60
80
100
←AKR7A3
←CAPZB
←AKR7A2
ENSG00000306364→
MICOS10→
RN7SL277P→
←ENSG00000306287
MRTO4→
SLC66A1→
ENSG00000306329→
RNU4−28P→
←AKR7L
ENSG00000270728→
19.25
19.3
19.35
19.4
19.45
19.5
19.55
Chromosome 1 (Mb)

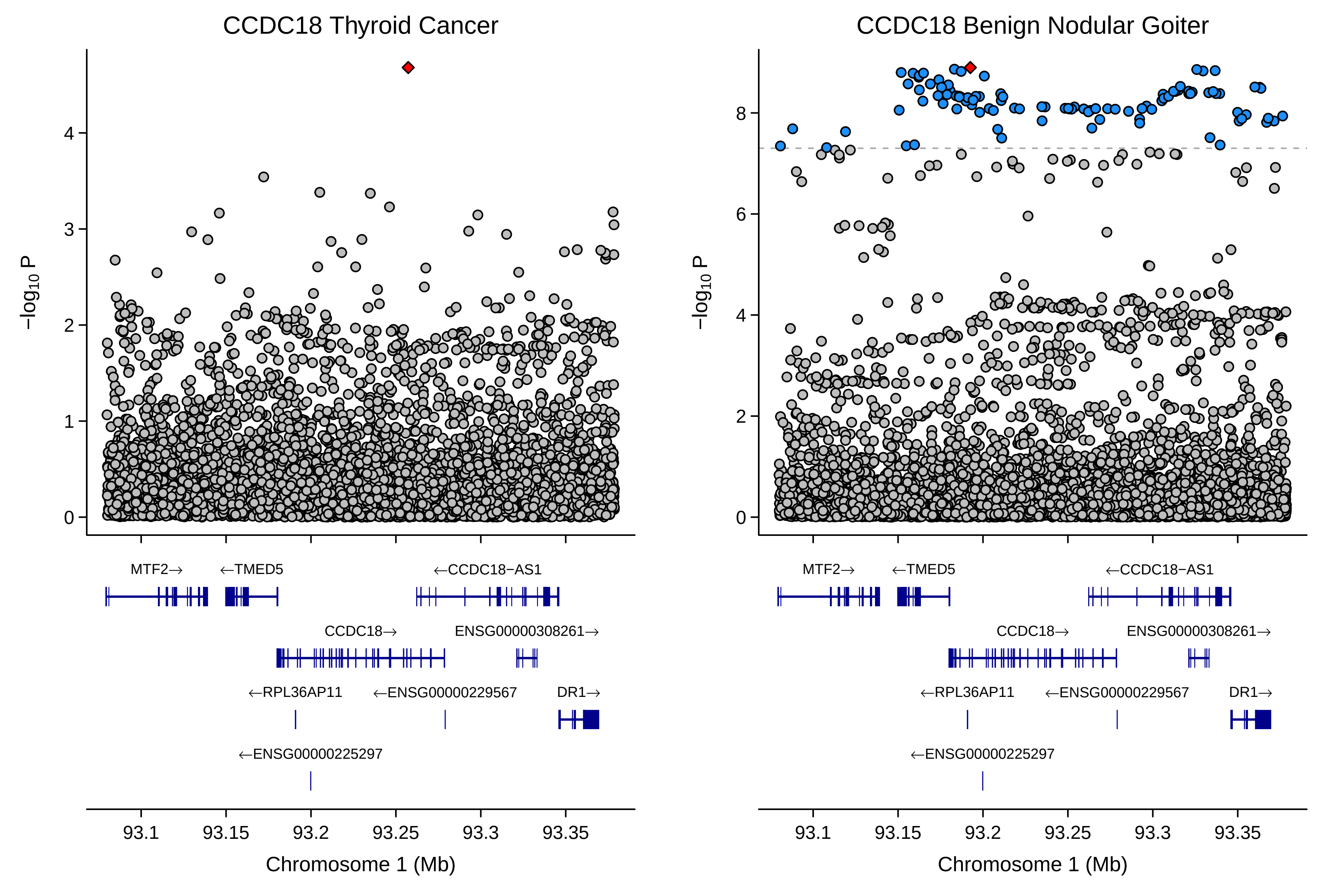
CCDC18 Thyroid Cancer
CCDC18 Benign Nodular Goiter
−log10 P
MTF2→
←TMED5
←CCDC18−AS1
CCDC18→
ENSG00000308261→
←RPL36AP11
←ENSG00000229567
DR1→
←ENSG00000225297
93.1
93.15
93.2
93.25
93.3
93.35
Chromosome 1 (Mb)

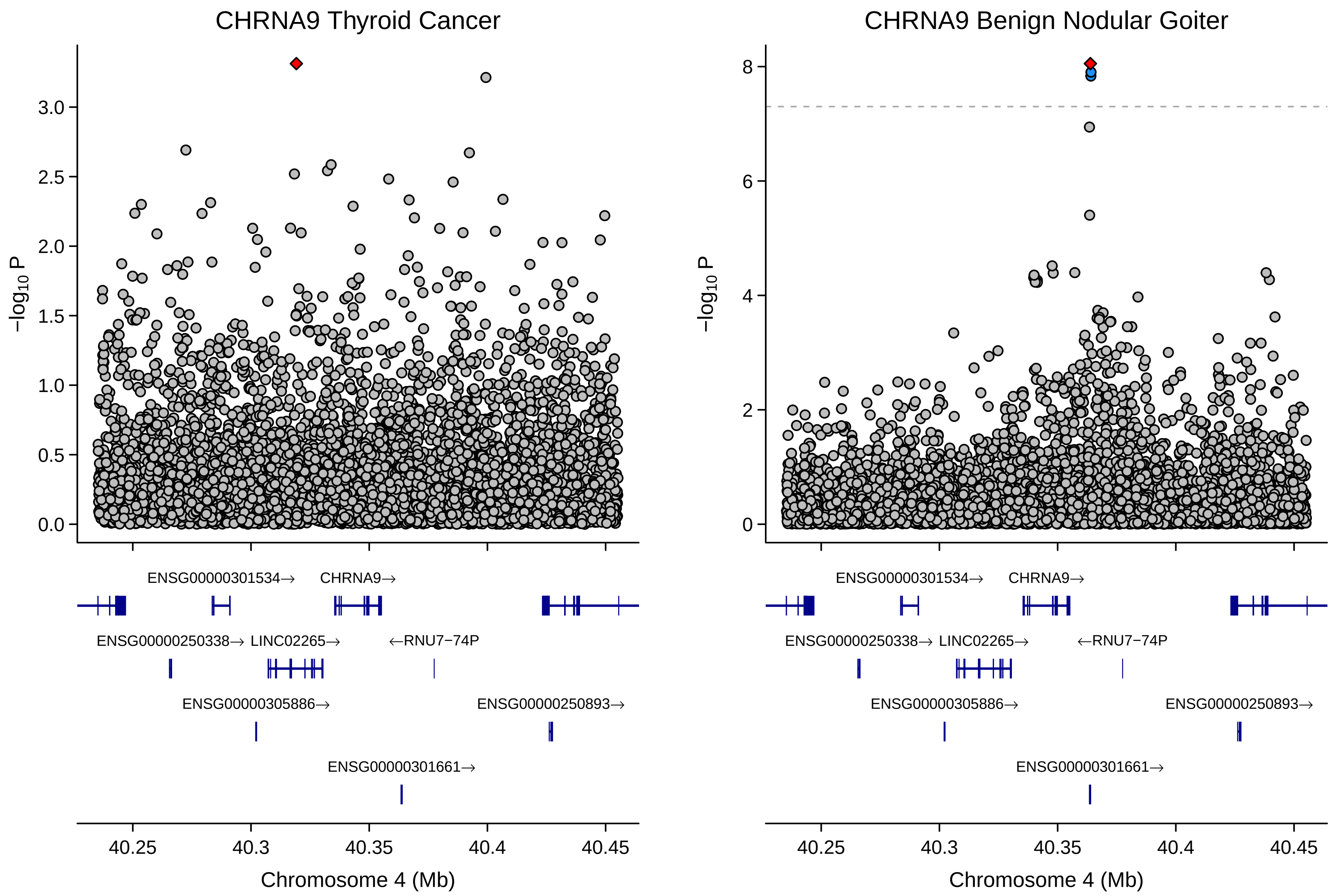

CHRNA9 Thyroid Cancer
CHRNA9 Benign Nodular Goiter
$-\log_{10}$ P
0.0
0.5
1.0
1.5
2.0
2.5
3.0
0
2
4
6
8
ENSG00000301534→
CHRNA9→
ENSG00000250338→
LINC02265→
←RNU7−74P
ENSG00000305886→
ENSG00000250893→
ENSG00000301661→
40.25
40.3
40.35
40.4
40.45
Chromosome 4 (Mb)

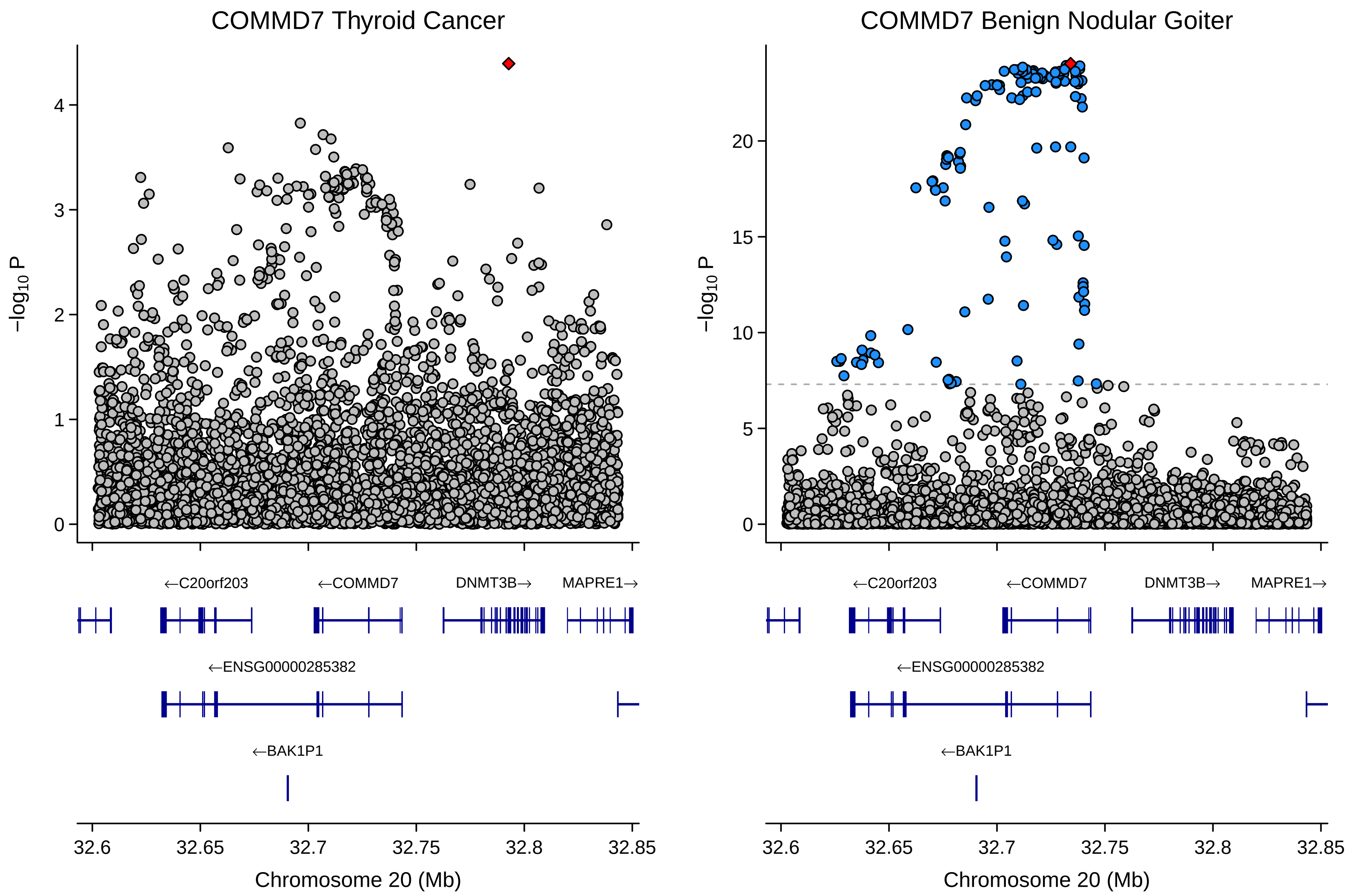

COMMD7 Thyroid Cancer
COMMD7 Benign Nodular Goiter
−log10 P
0
1
2
3
4
5
10
15
20
←C20orf203
←COMMD7
DNMT3B→
MAPRE1→
←ENSG00000285382
←BAK1P1
32.6
32.65
32.7
32.75
32.8
32.85
Chromosome 20 (Mb)

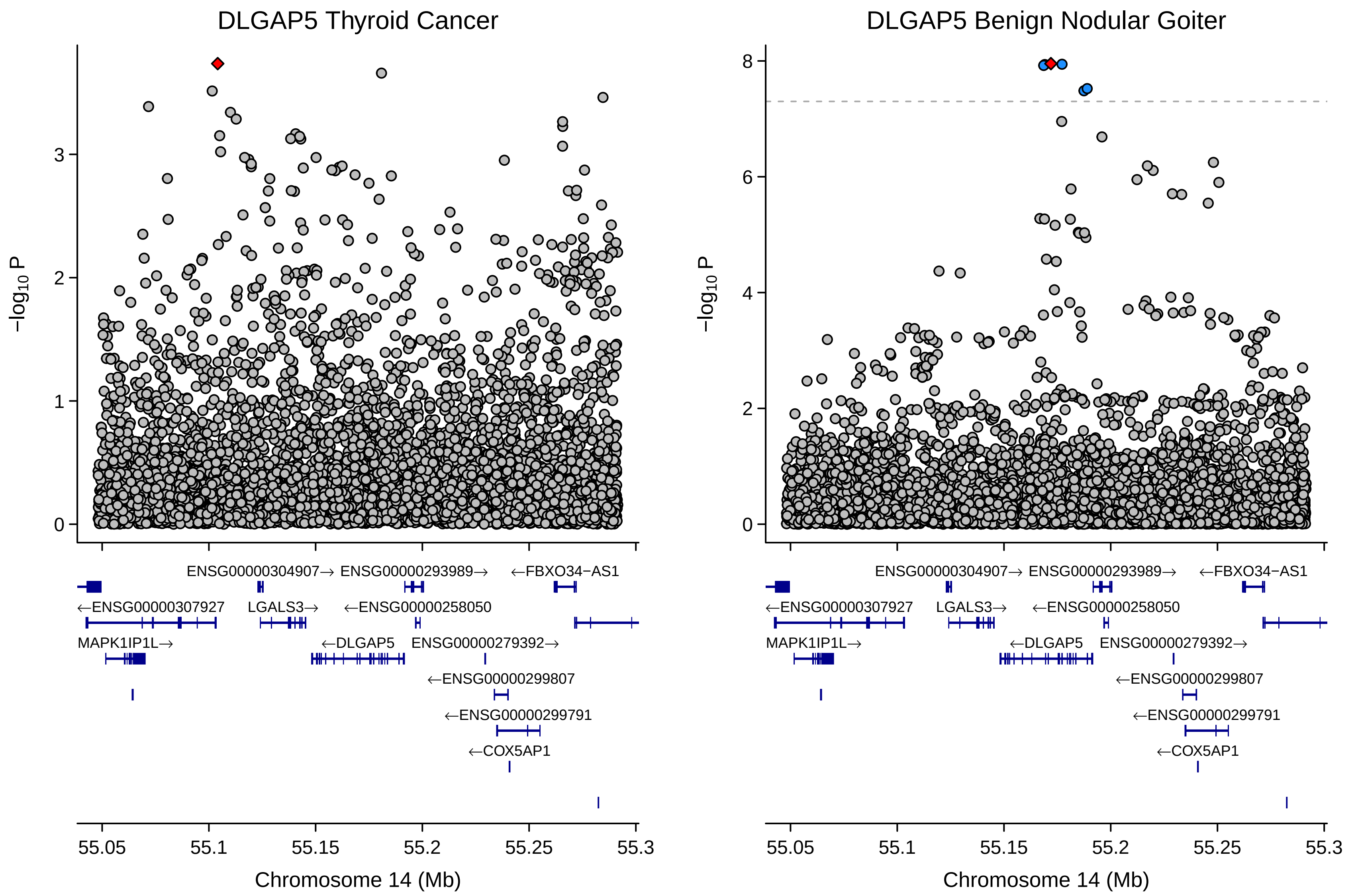

DLGAP5 Thyroid Cancer
DLGAP5 Benign Nodular Goiter
−log10 P
Chromosome 14 (Mb)
55.05
55.1
55.15
55.2
55.25
55.3
ENSG00000304907→
ENSG00000293989→
←FBXO34−AS1
←ENSG00000307927
LGALS3→
←ENSG00000258050
MAPK1IP1L→
←DLGAP5
ENSG00000279392→
←ENSG00000299807
←ENSG00000299791
←COX5AP1

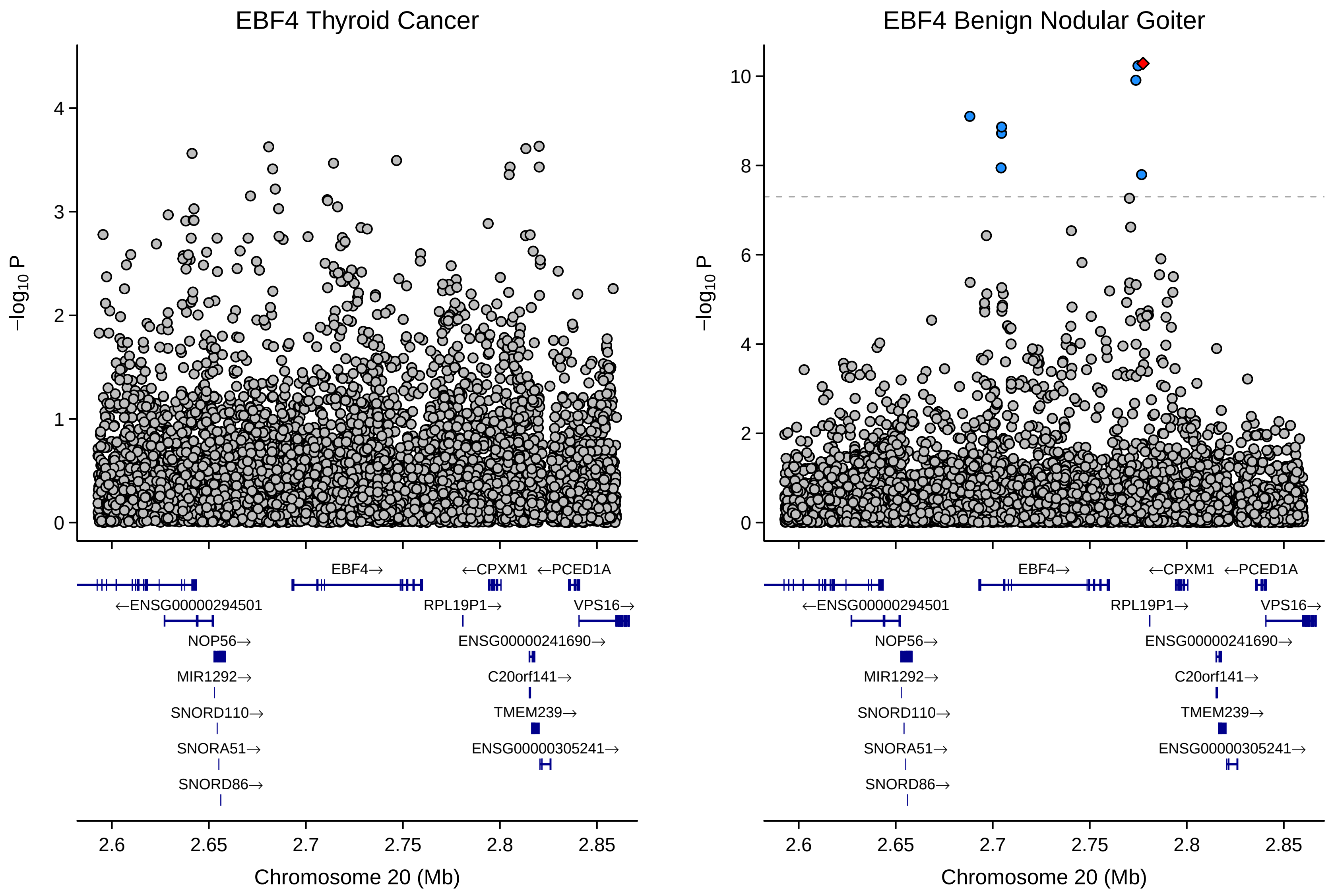

EBF4 Thyroid Cancer
EBF4 Benign Nodular Goiter
−log10 P
0
1
2
3
4
6
8
10
EBF4→
←CPXM1
←PCED1A
←ENSG00000294501
RPL19P1→
VPS16→
NOP56→
ENSG00000241690→
MIR1292→
C20orf141→
SNORD110→
TMEM239→
SNORA51→
ENSG00000305241→
SNORD86→
2.6
2.65
2.7
2.75
2.8
2.85
Chromosome 20 (Mb)

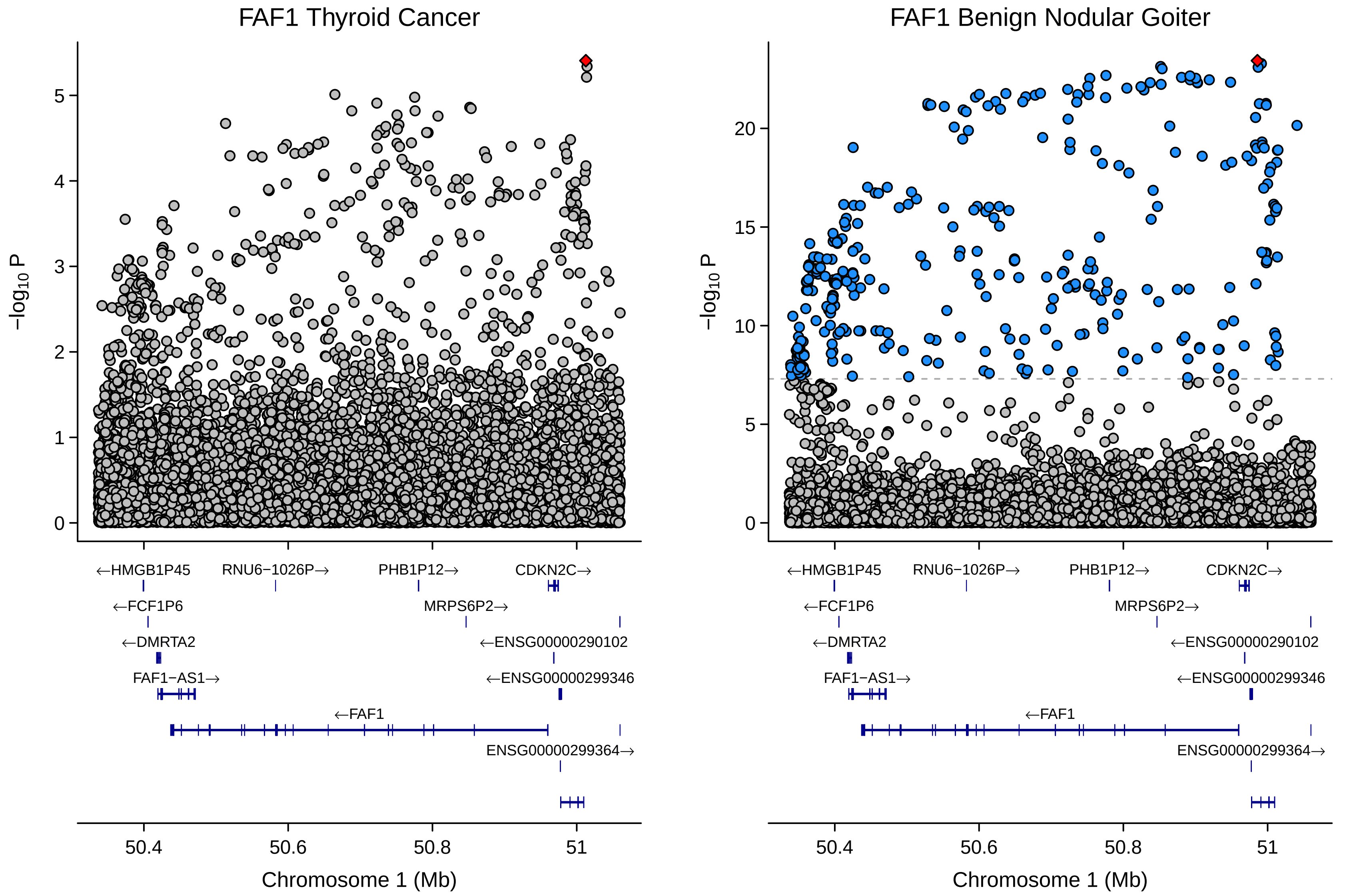
FAF1 Thyroid Cancer
FAF1 Benign Nodular Goiter
−log10 P
0
1
2
3
4
5
10
15
20
←HMGB1P45
RNU6−1026P→
PHB1P12→
CDKN2C→
←FCF1P6
MRPS6P2→
←DMRTA2
←ENSG00000290102
FAF1−AS1→
←ENSG00000299346
←FAF1
ENSG00000299364→
50.4
50.6
50.8
51
Chromosome 1 (Mb)

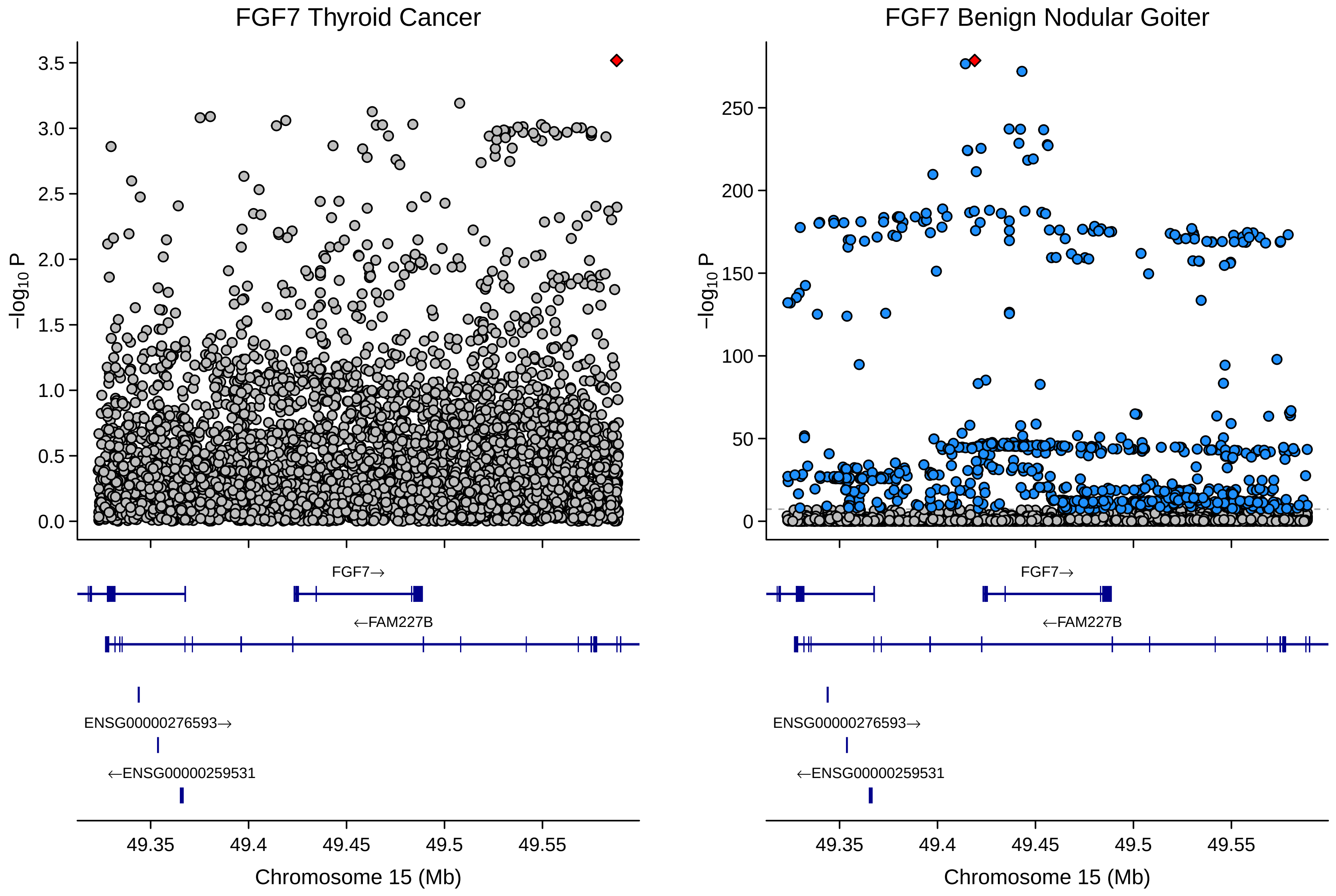
FGF7 Thyroid Cancer
FGF7 Benign Nodular Goiter
−log10 P
FGF7→
←FAM227B
ENSG00000276593→
←ENSG00000259531
49.35
49.4
49.45
49.5
49.55
Chromosome 15 (Mb)

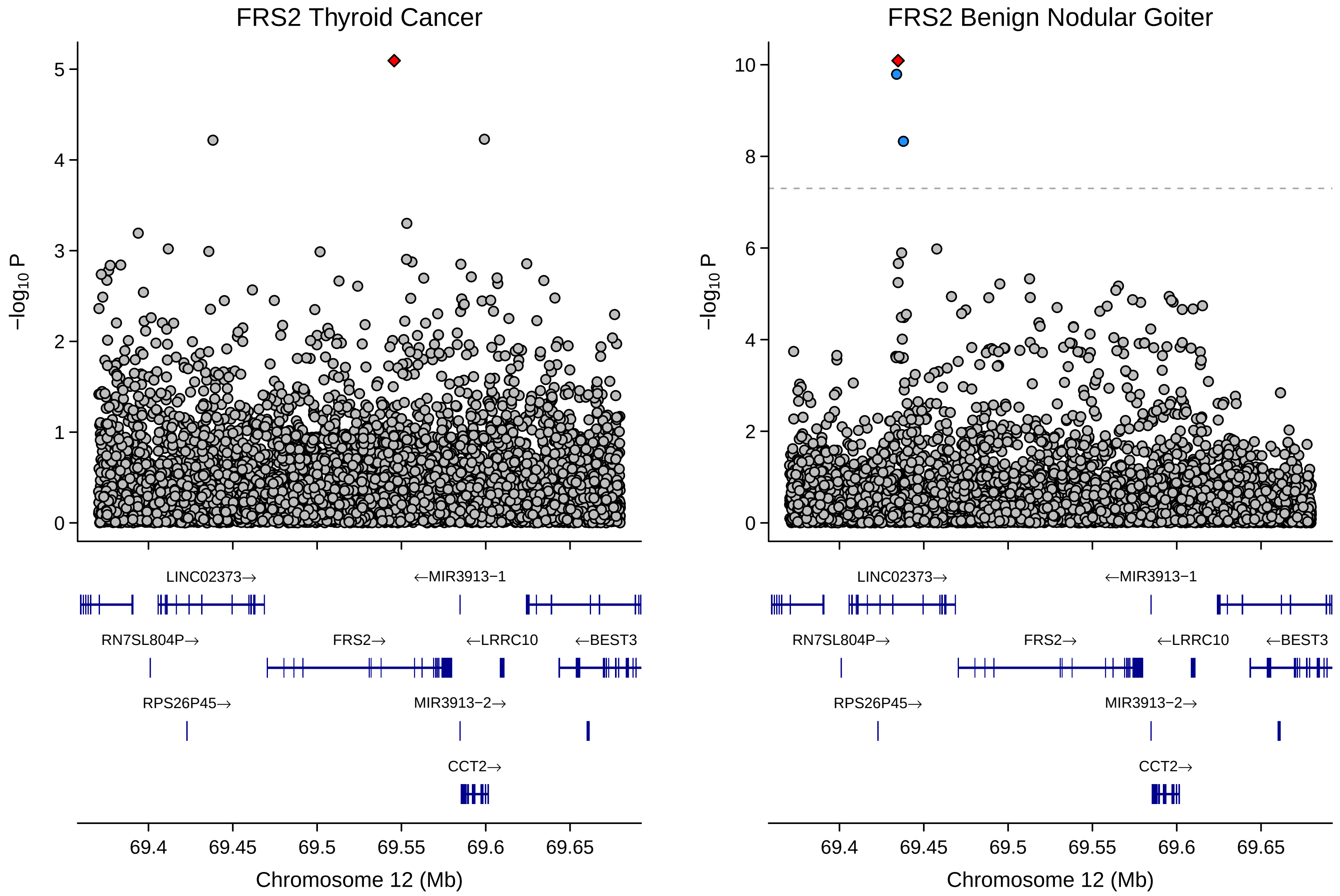

FRS2 Thyroid Cancer
FRS2 Benign Nodular Goiter
$-\log_{10}$ P
LINC02373→
←MIR3913−1
RN7SL804P→
FRS2→
←LRRC10
←BEST3
RPS26P45→
MIR3913−2→
CCT2→
69.4
69.45
69.5
69.55
69.6
69.65
Chromosome 12 (Mb)

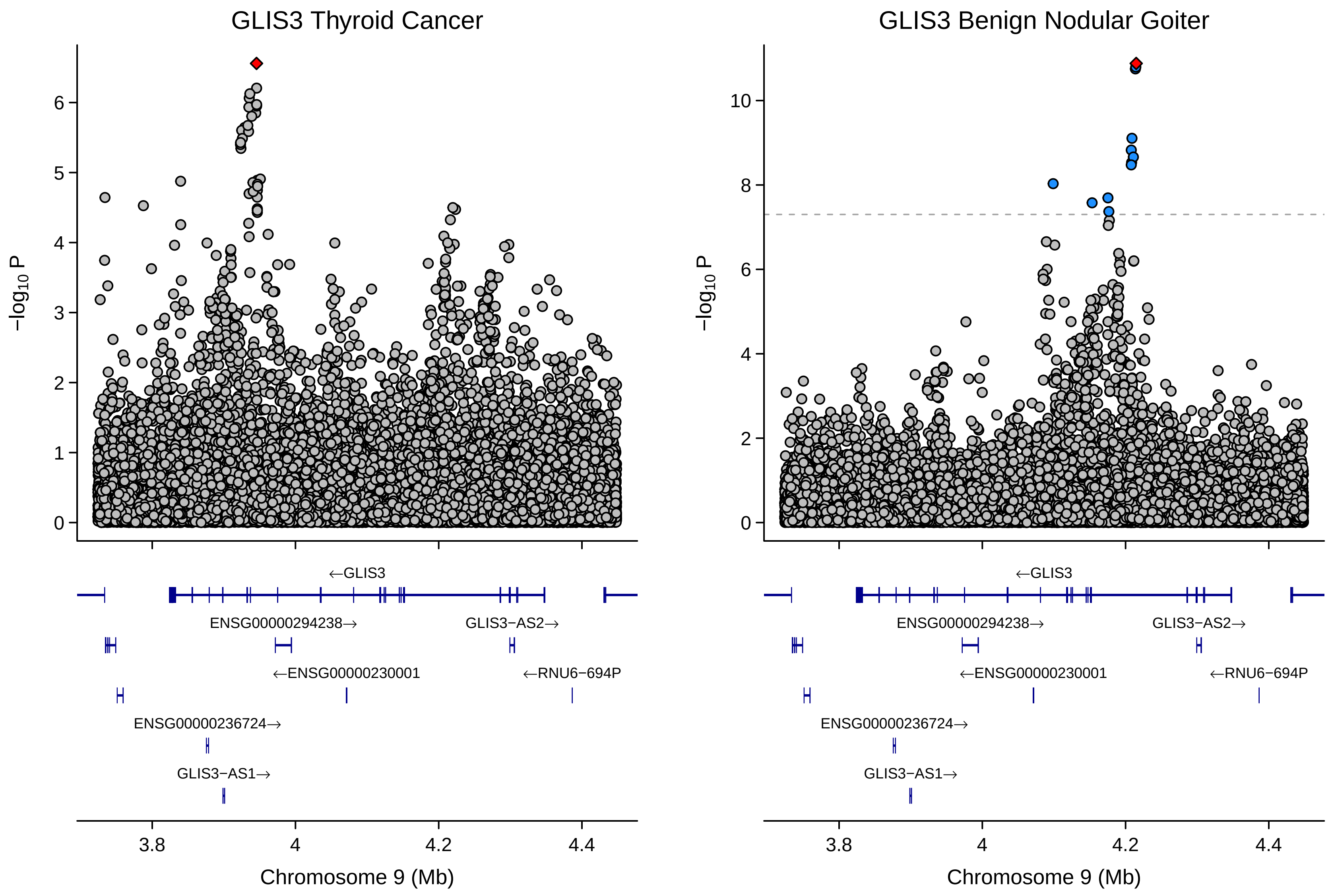

GLIS3 Thyroid Cancer
GLIS3 Benign Nodular Goiter
−log10 P
0
1
2
3
4
5
6
8
10
←GLIS3
ENSG00000294238→
GLIS3−AS2→
←ENSG00000230001
←RNU6−694P
ENSG00000236724→
GLIS3−AS1→
3.8
4
4.2
4.4
Chromosome 9 (Mb)

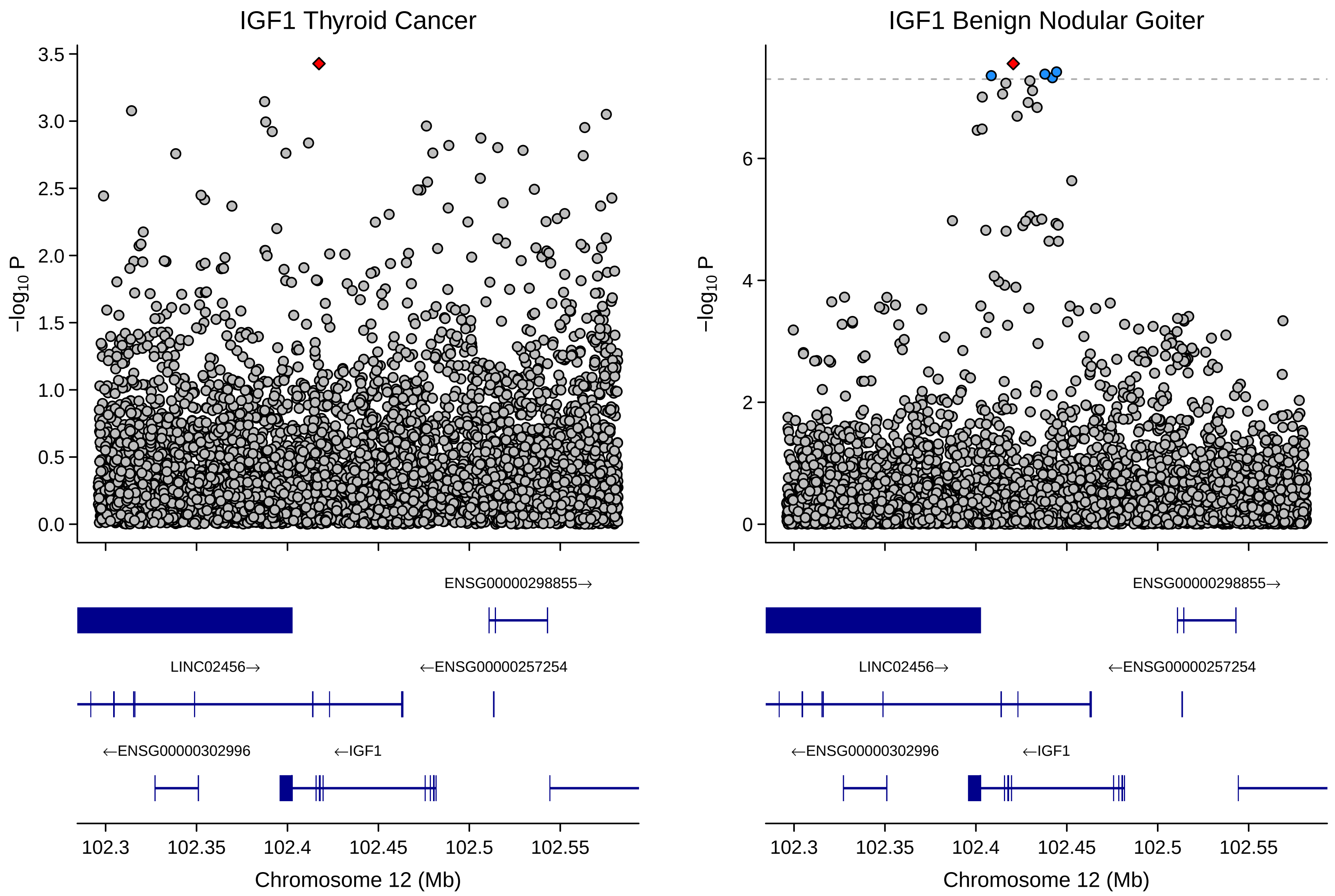

IGF1 Thyroid Cancer
IGF1 Benign Nodular Goiter
−log10 P
3.5
3.0
2.5
2.0
1.5
1.0
0.5
0.0
6
4
2
0
ENSG00000298855→
LINC02456→
←ENSG00000257254
←ENSG00000302996
←IGF1
102.3
102.35
102.4
102.45
102.5
102.55
Chromosome 12 (Mb)

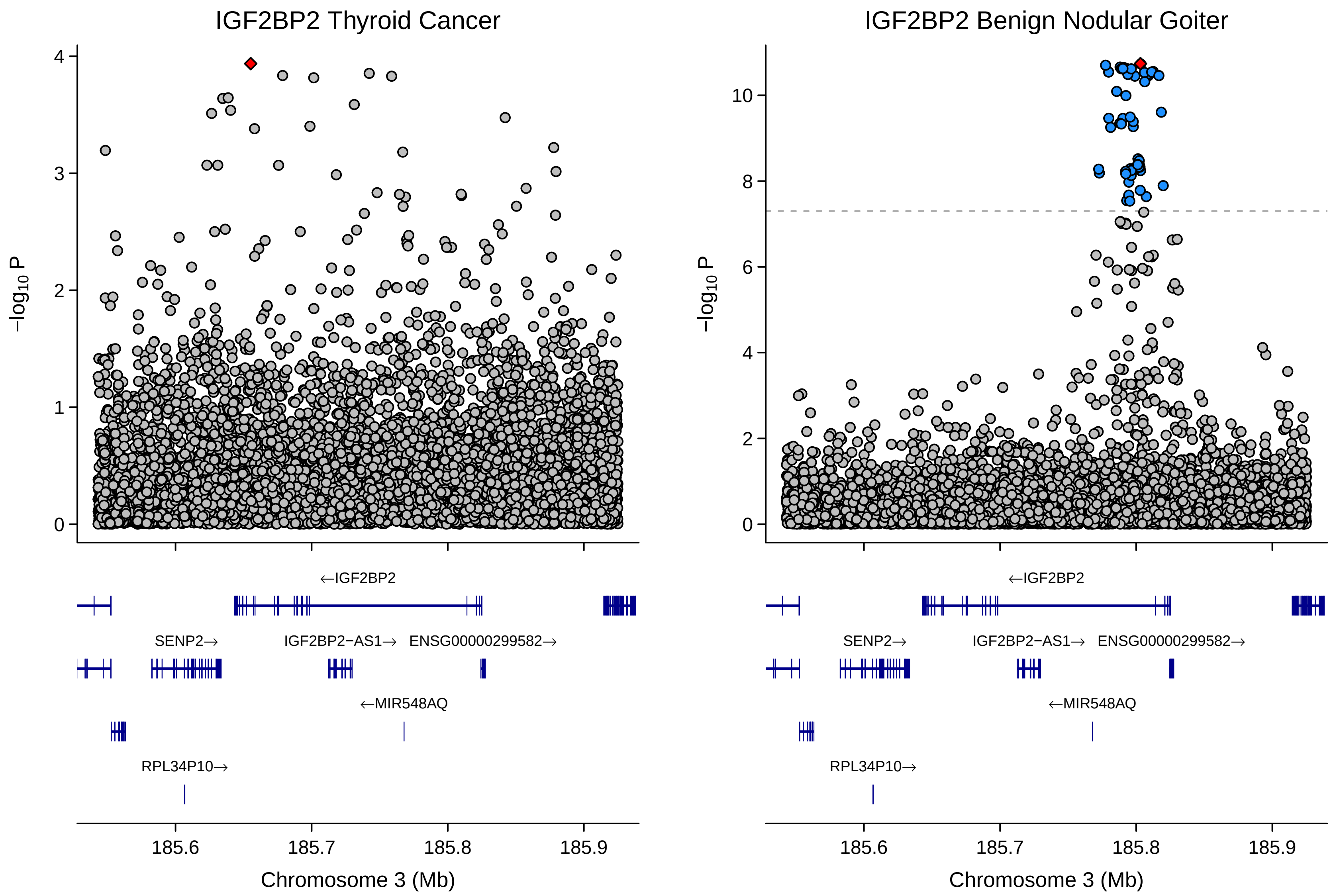
IGF2BP2 Thyroid Cancer
IGF2BP2 Benign Nodular Goiter
−log10 P
Chromosome 3 (Mb)
185.6
185.7
185.8
185.9
←IGF2BP2
SENP2→
IGF2BP2−AS1→
ENSG00000299582→
←MIR548AQ
RPL34P10→

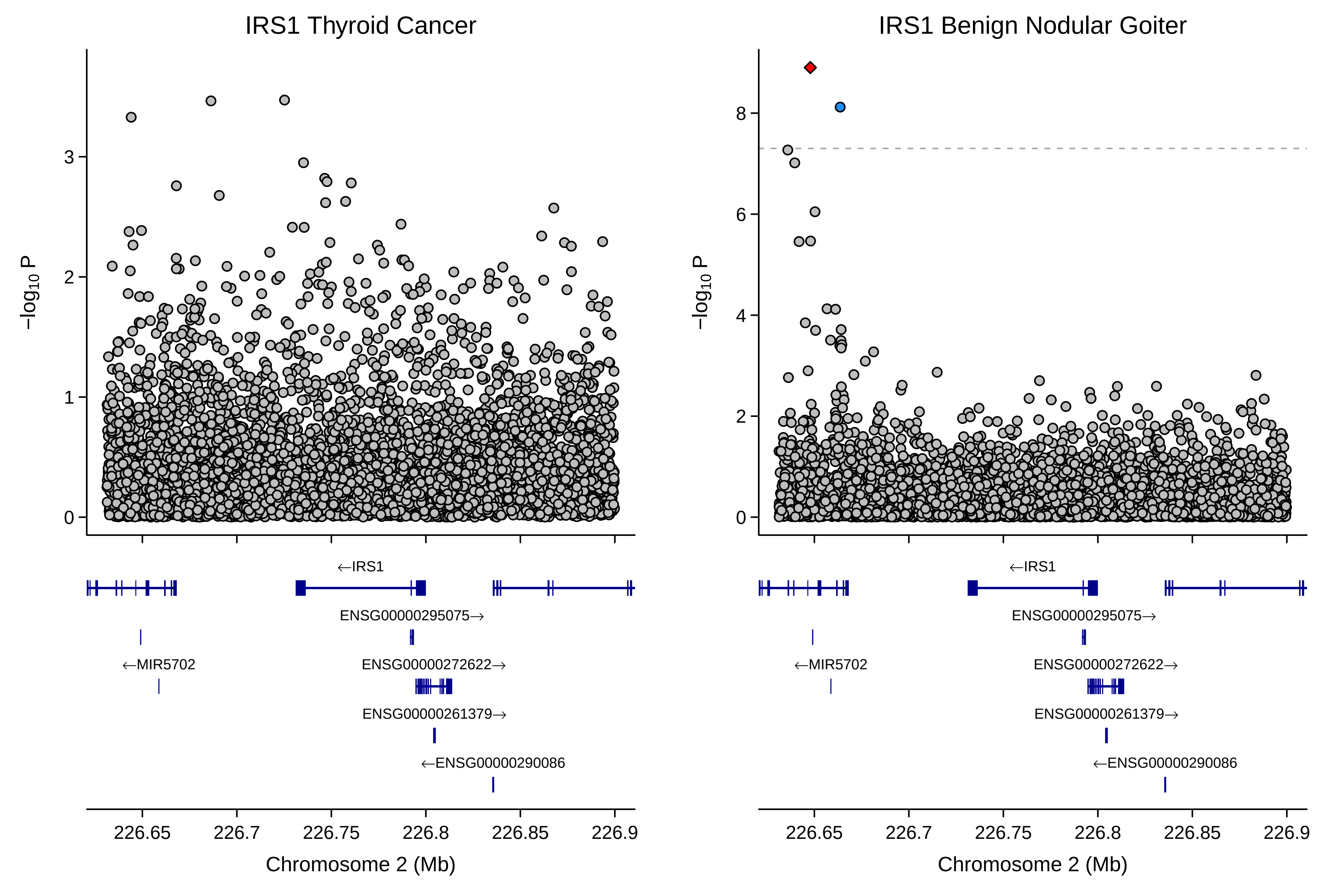

IRS1 Thyroid Cancer
IRS1 Benign Nodular Goiter
−log10 P
←IRS1
ENSG00000295075→
←MIR5702
ENSG00000272622→
ENSG00000261379→
←ENSG00000290086
226.65
226.7
226.75
226.8
226.85
226.9
Chromosome 2 (Mb)

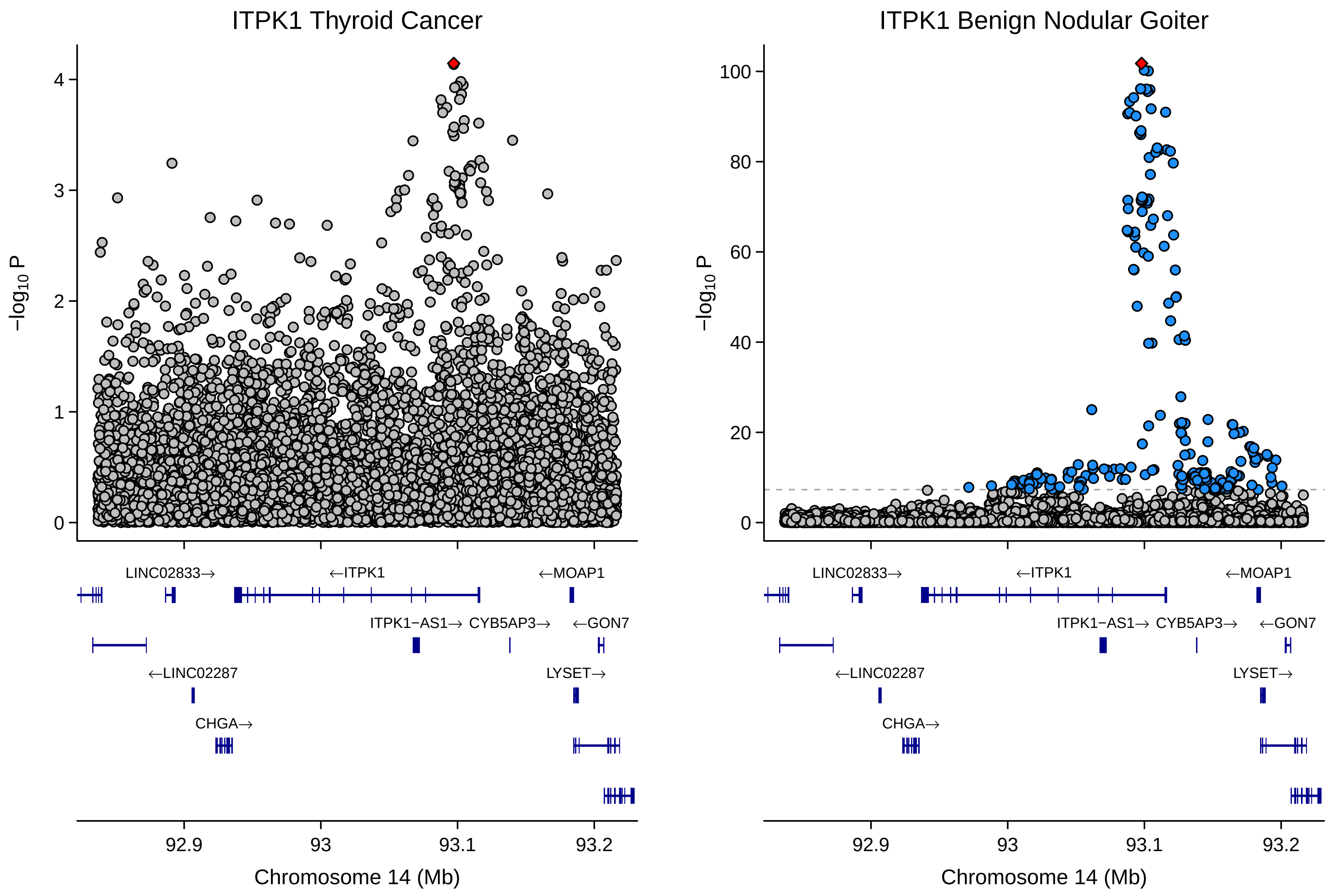
ITPK1 Thyroid Cancer
ITPK1 Benign Nodular Goiter
−log10 P
0
1
2
3
4
0
20
40
60
80
100
LINC02833→
←ITPK1
←MOAP1
ITPK1−AS1→
CYB5AP3→
←GON7
←LINC02287
LYSET→
CHGA→
92.9
93
93.1
93.2
Chromosome 14 (Mb)

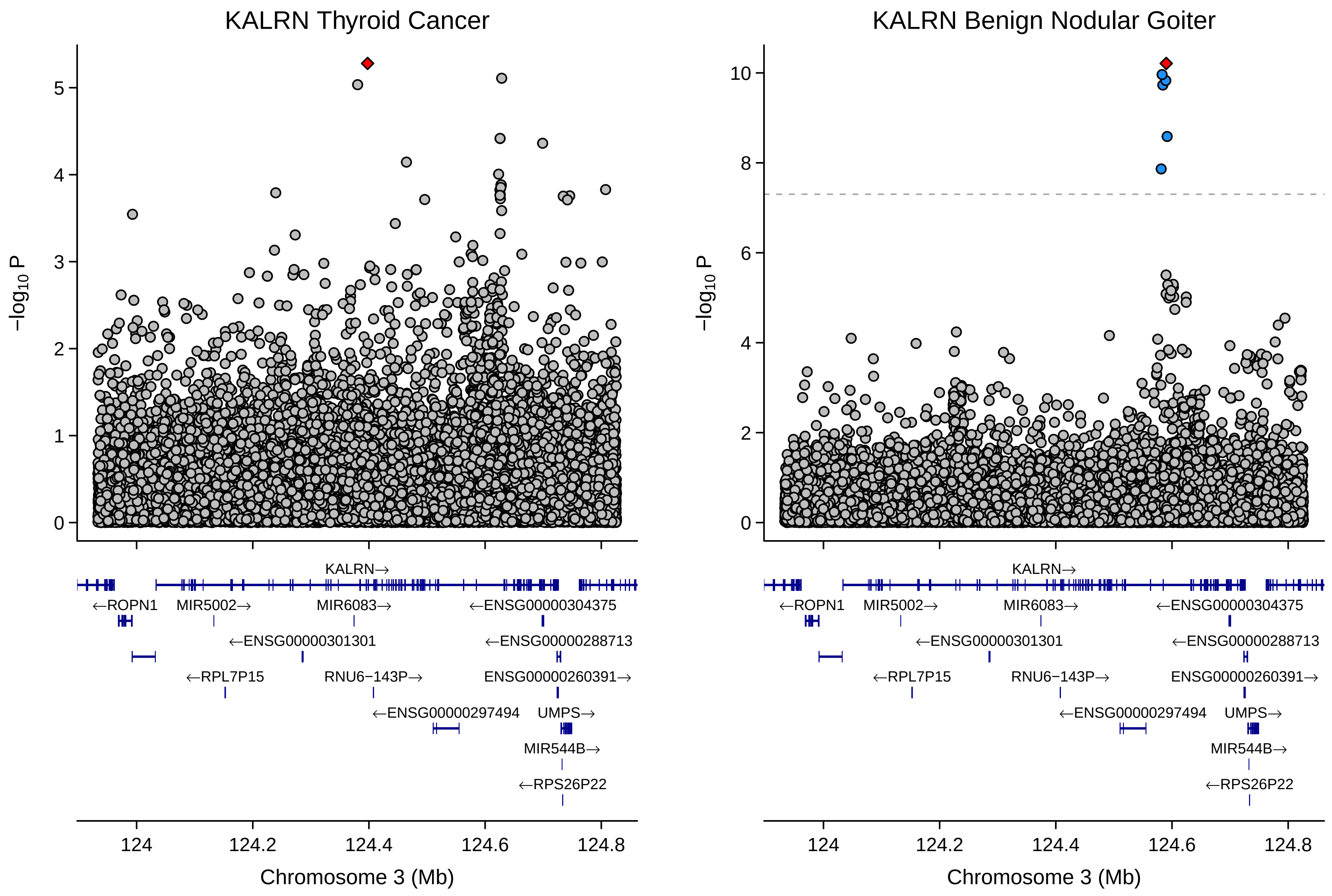

KALRN Thyroid Cancer
KALRN Benign Nodular Goiter
−log10 P
KALRN→
←ROPN1
MIR5002→
MIR6083→
←ENSG00000304375
←ENSG00000301301
←ENSG00000288713
←RPL7P15
RNU6−143P→
ENSG00000260391→
←ENSG00000297494
UMPS→
MIR544B→
←RPS26P22
Chromosome 3 (Mb)

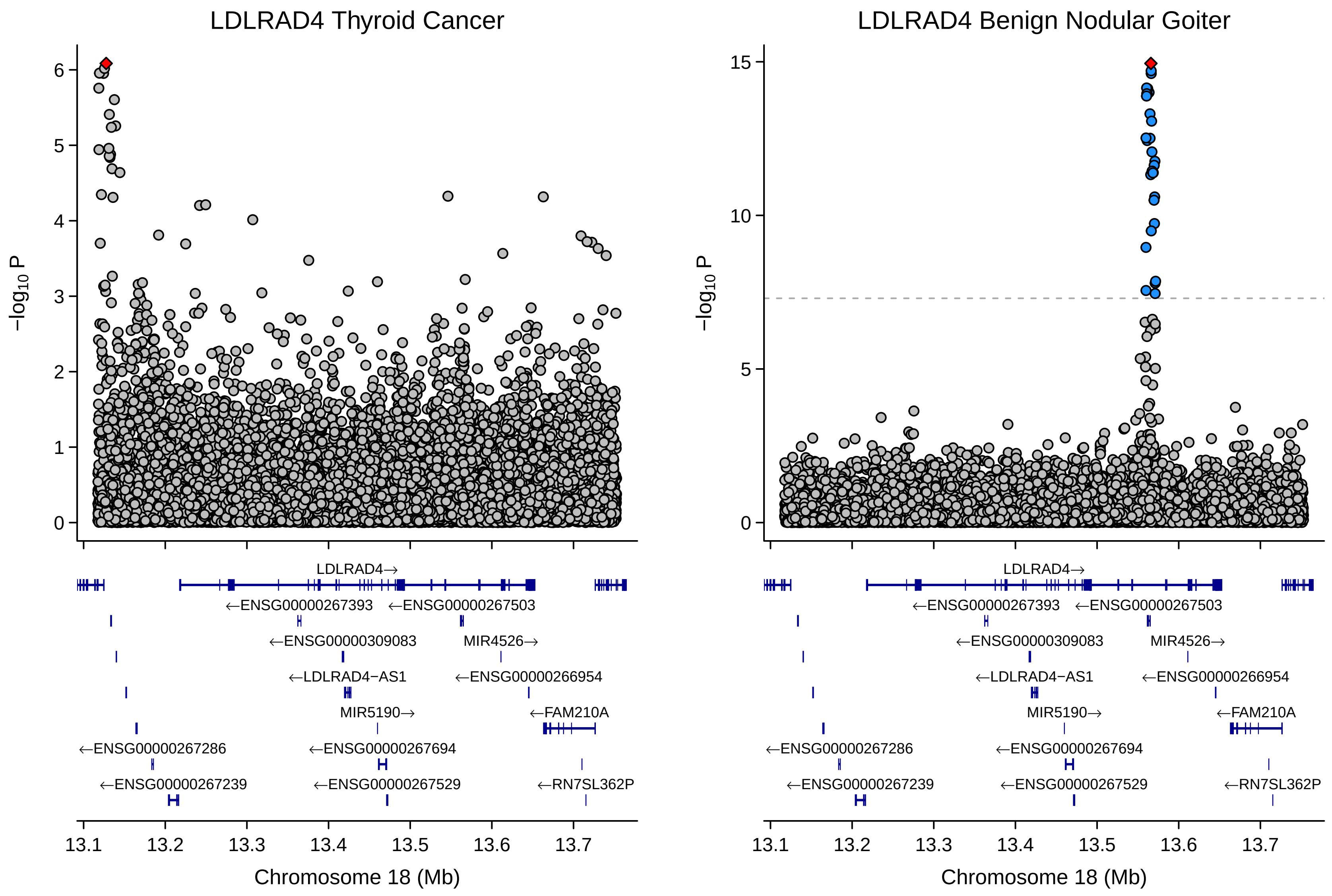

LDLRAD4 Thyroid Cancer
LDLRAD4 Benign Nodular Goiter
−log10 P
Chromosome 18 (Mb)
13.1
13.2
13.3
13.4
13.5
13.6
13.7
LDLRAD4→
←ENSG00000267393
←ENSG00000267503
←ENSG00000309083
MIR4526→
←LDLRAD4−AS1
←ENSG00000266954
MIR5190→
←FAM210A
←ENSG00000267286
←ENSG00000267694
←ENSG00000267239
←ENSG00000267529
←RN7SL362P

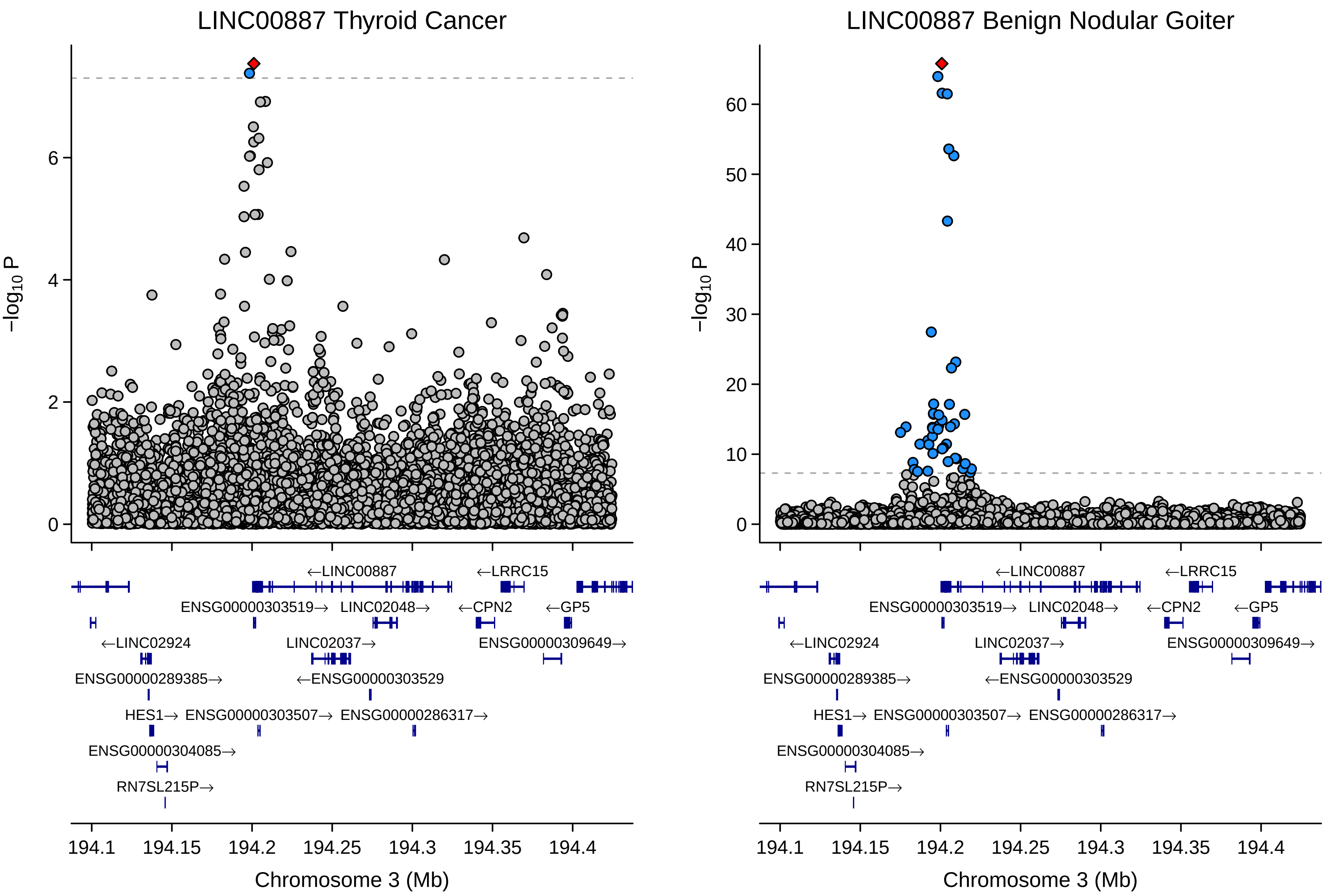
LINC00887 Thyroid Cancer
LINC00887 Benign Nodular Goiter
−log10 P
0
2
4
6
10
20
30
40
50
60
←LINC00887
←LRRC15
ENSG00000303519→
LINC02048→
←CPN2
←GP5
←LINC02924
LINC02037→
ENSG00000309649→
ENSG00000289385→
←ENSG00000303529
HES1→
ENSG00000303507→
ENSG00000286317→
ENSG00000304085→
RN7SL215P→
194.1
194.15
194.2
194.25
194.3
194.35
194.4
Chromosome 3 (Mb)

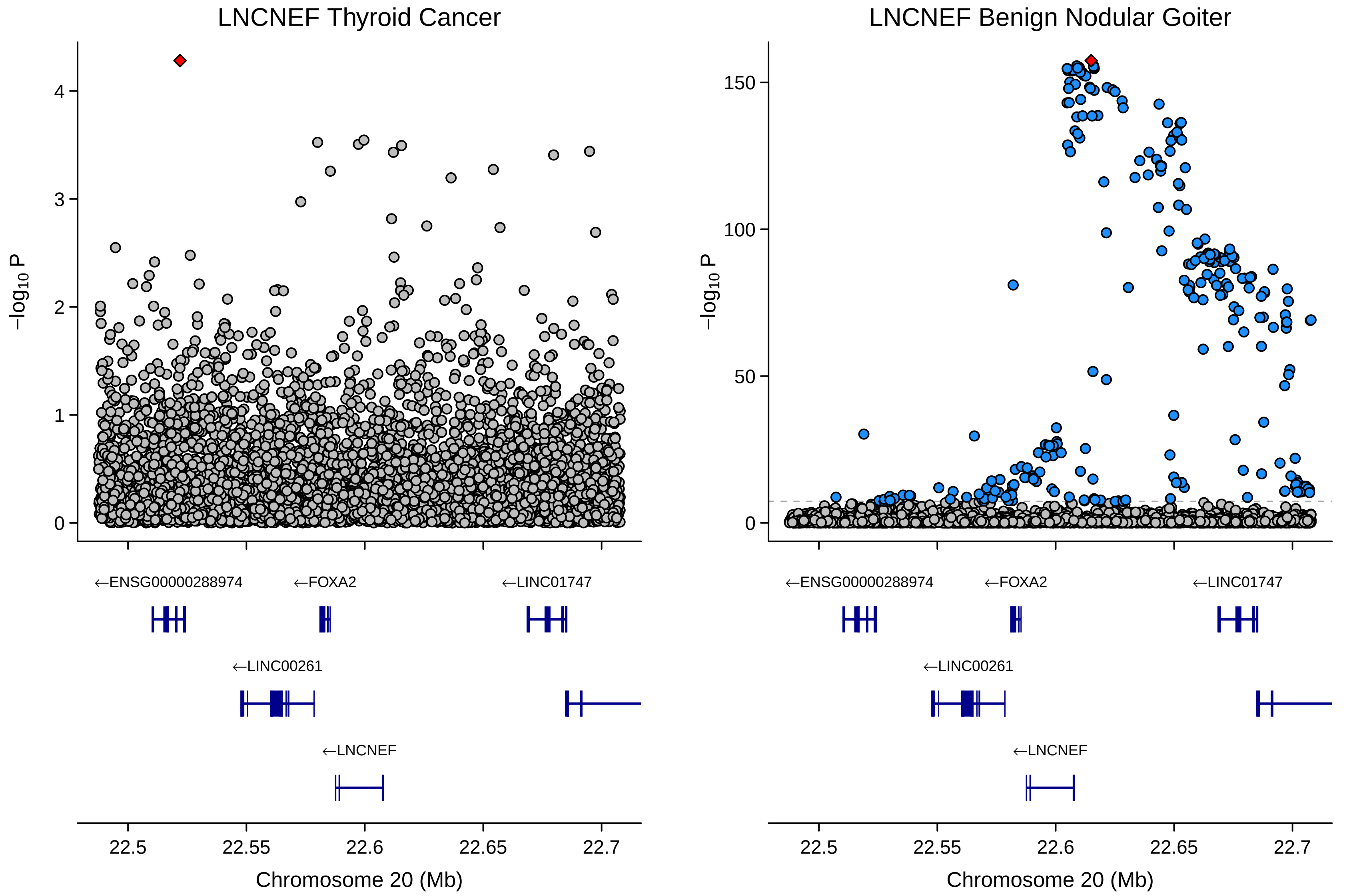

LNCNEF Thyroid Cancer
LNCNEF Benign Nodular Goiter
−log10 P
←ENSG00000288974
←FOXA2
←LINC01747
←LINC00261
←LNCNEF
22.5
22.55
22.6
22.65
22.7
Chromosome 20 (Mb)

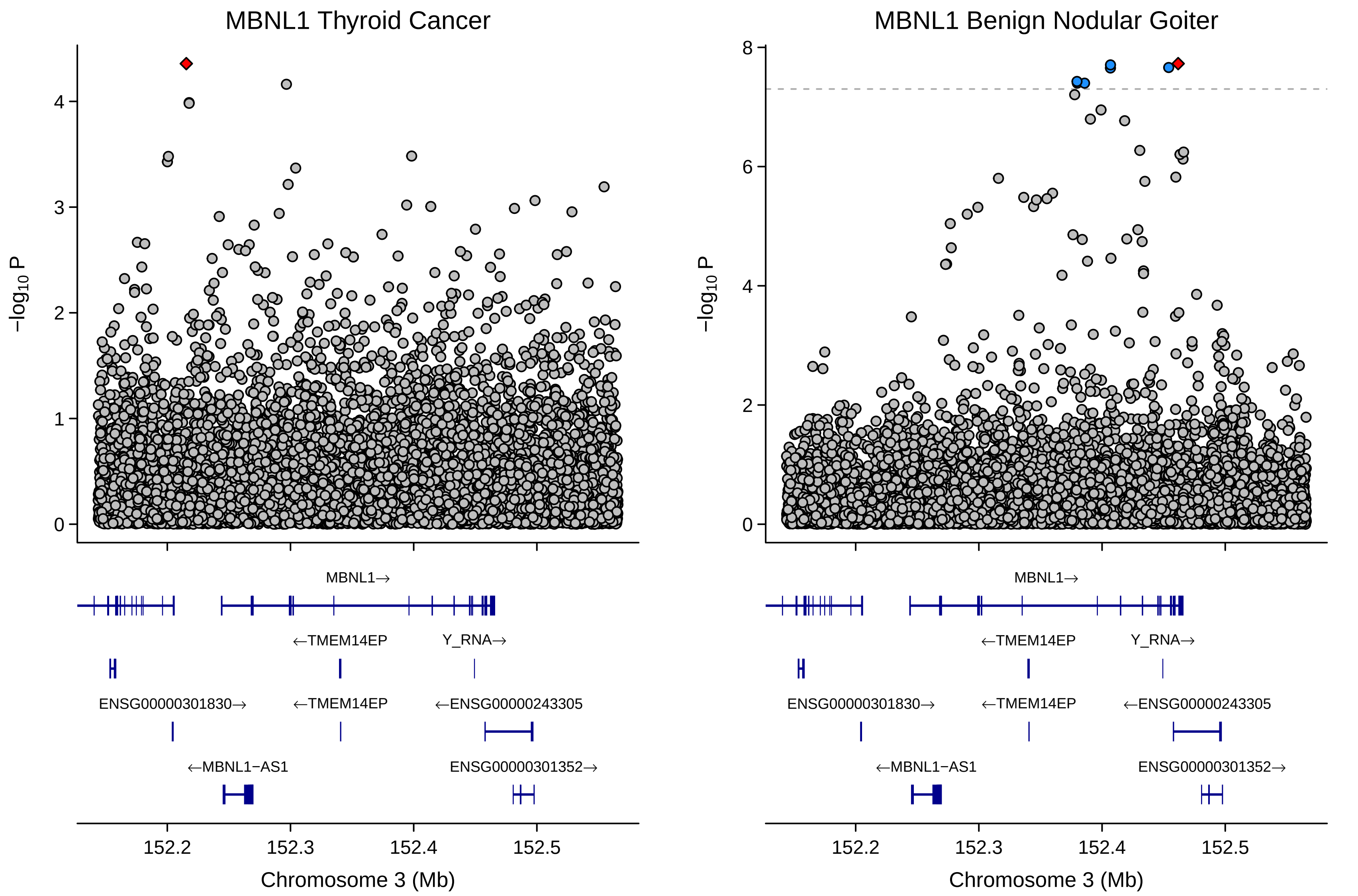
MBNL1 Thyroid Cancer
MBNL1 Benign Nodular Goiter
−log10 P
MBNL1→
←TMEM14EP
Y_RNA→
ENSG00000301830→
←TMEM14EP
←ENSG00000243305
←MBNL1−AS1
ENSG00000301352→
152.2
152.3
152.4
152.5
Chromosome 3 (Mb)

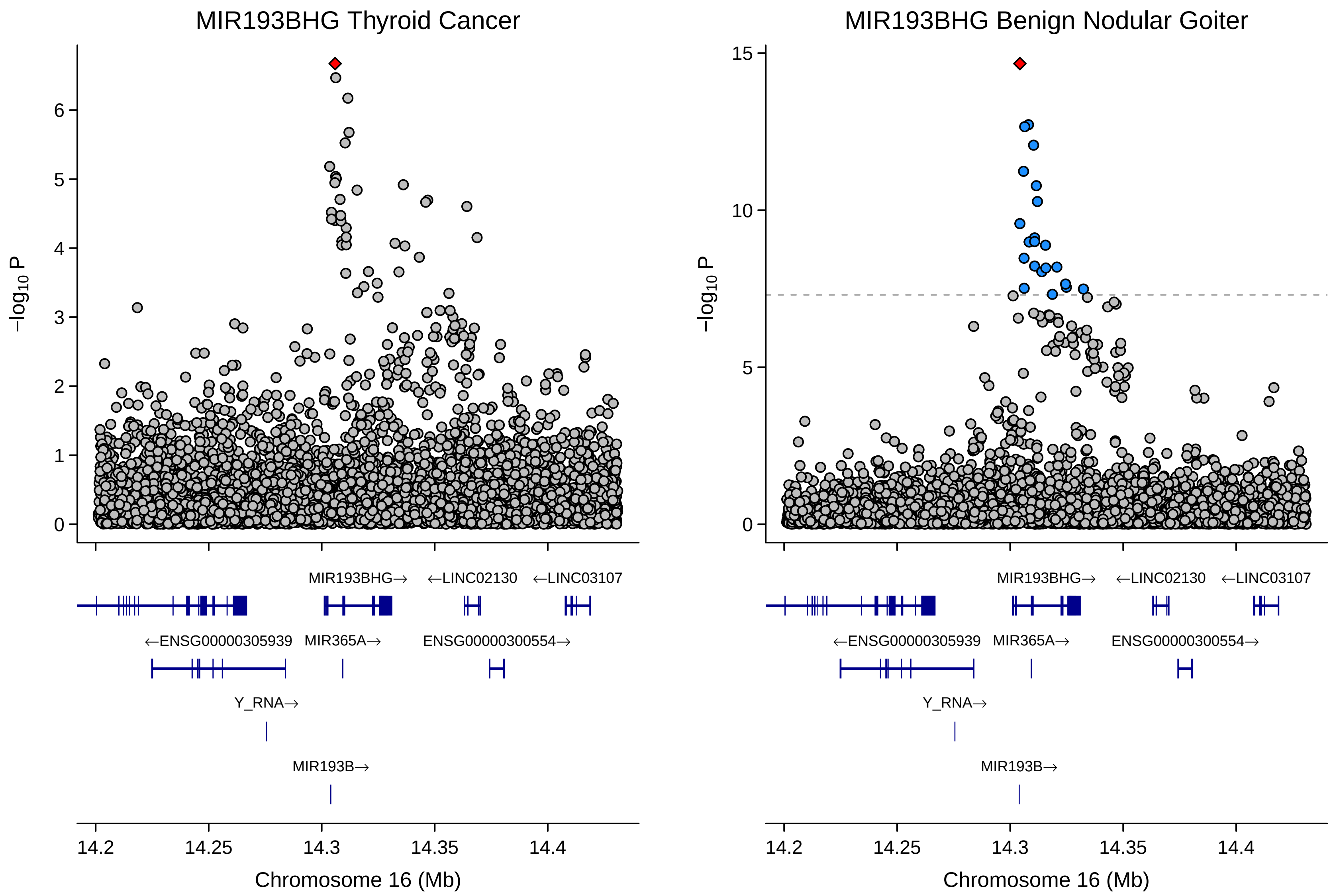

MIR193BHG Thyroid Cancer
MIR193BHG Benign Nodular Goiter
−log10 P
Chromosome 16 (Mb)
14.2
14.25
14.3
14.35
14.4
MIR193BHG→
←LINC02130
←LINC03107
←ENSG00000305939
MIR365A→
ENSG00000300554→
Y_RNA→
MIR193B→

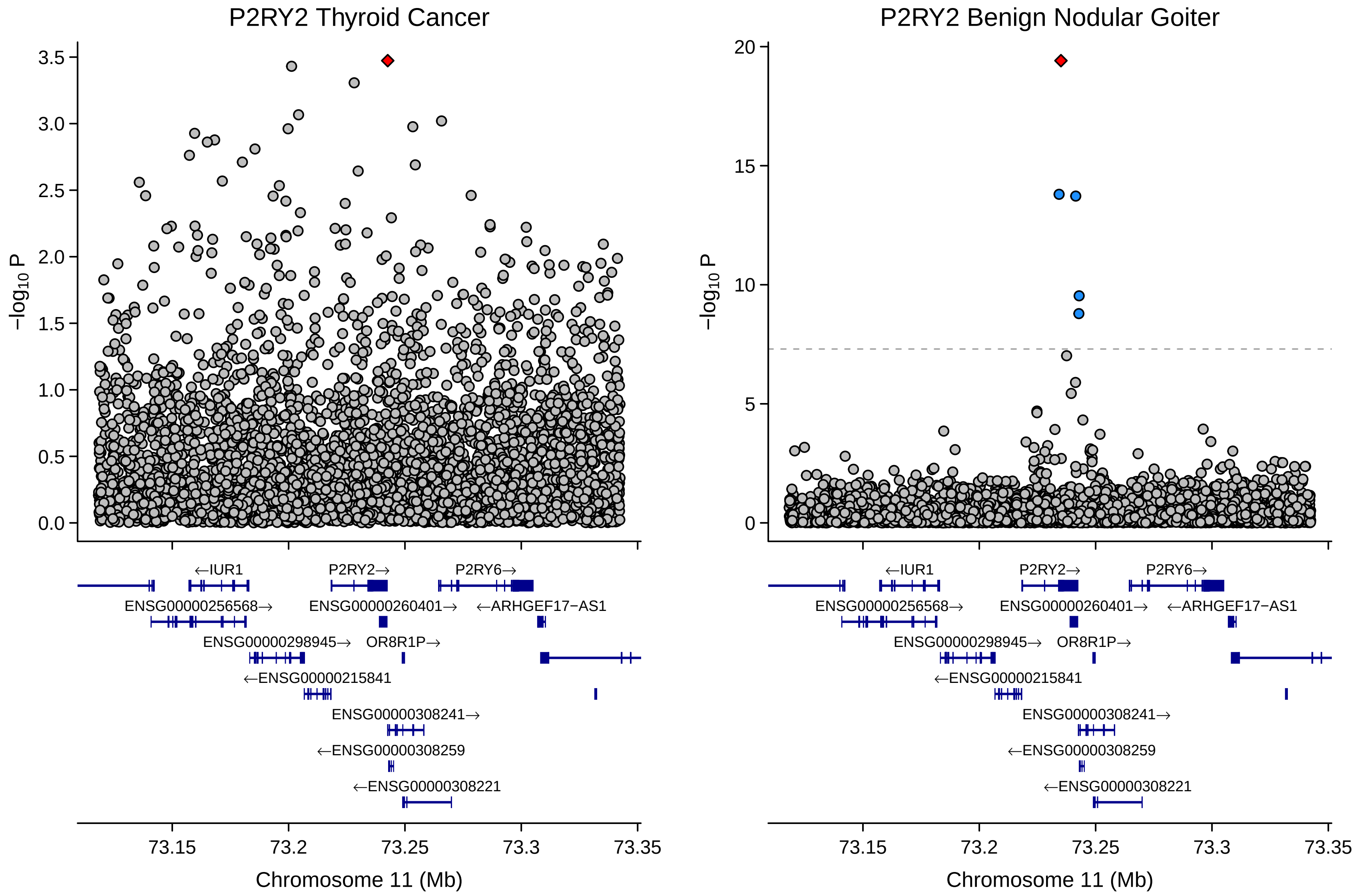
P2RY2 Thyroid Cancer
P2RY2 Benign Nodular Goiter
$-\log_{10}$ P
Chromosome 11 (Mb)
73.15
73.2
73.25
73.3
73.35
←IUR1
P2RY2→
P2RY6→
ENSG00000256568→
ENSG00000260401→
←ARHGEF17−AS1
ENSG00000298945→
OR8R1P→
←ENSG00000215841
ENSG00000308241→
←ENSG00000308259
←ENSG00000308221

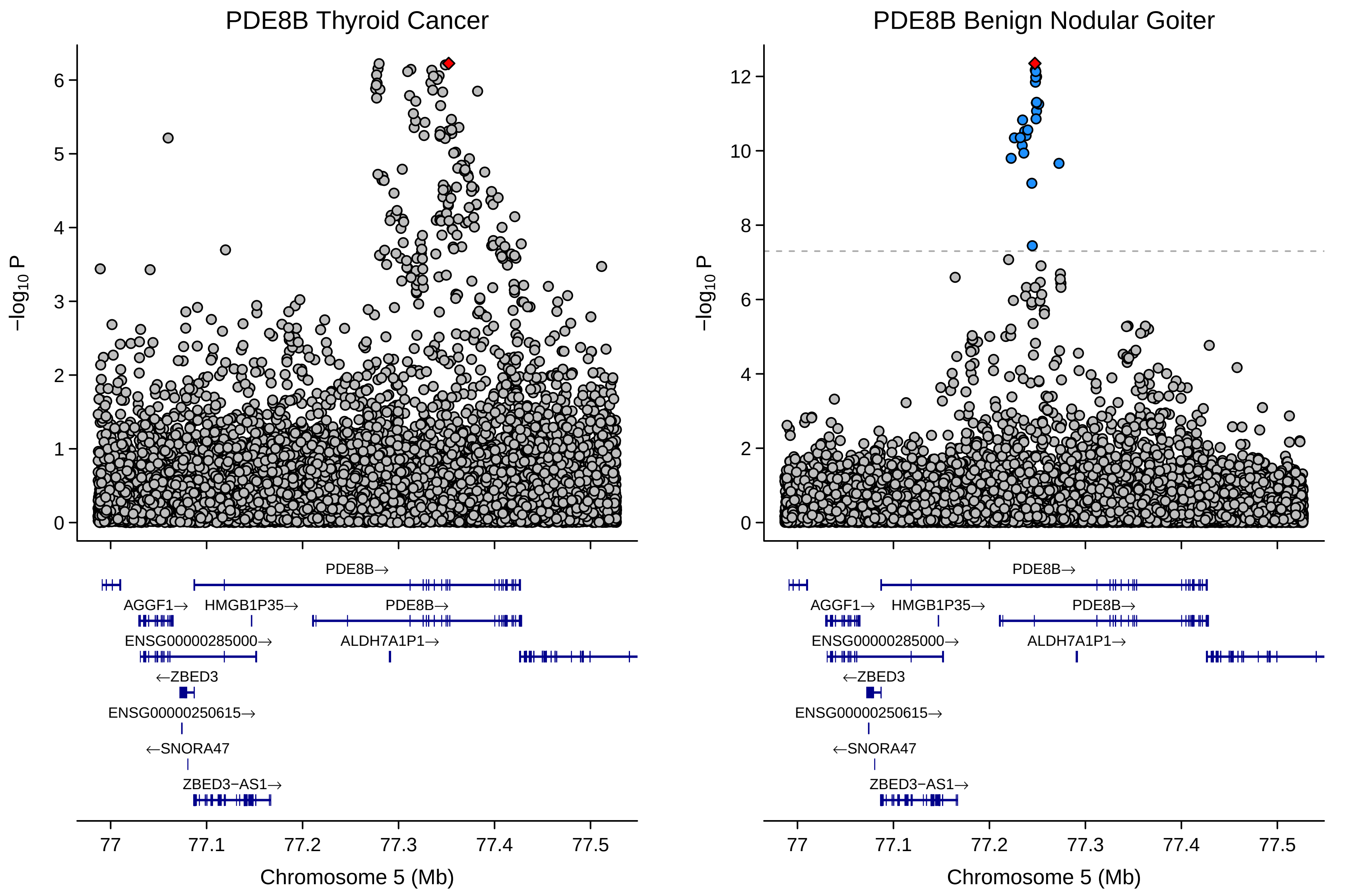

PDE8B Thyroid Cancer
PDE8B Benign Nodular Goiter
−log10 P
0
1
2
3
4
5
6
8
10
12
PDE8B→
AGGF1→
HMGB1P35→
ENSG00000285000→
ALDH7A1P1→
←ZBED3
ENSG00000250615→
←SNORA47
ZBED3−AS1→
77
77.1
77.2
77.3
77.4
77.5
Chromosome 5 (Mb)

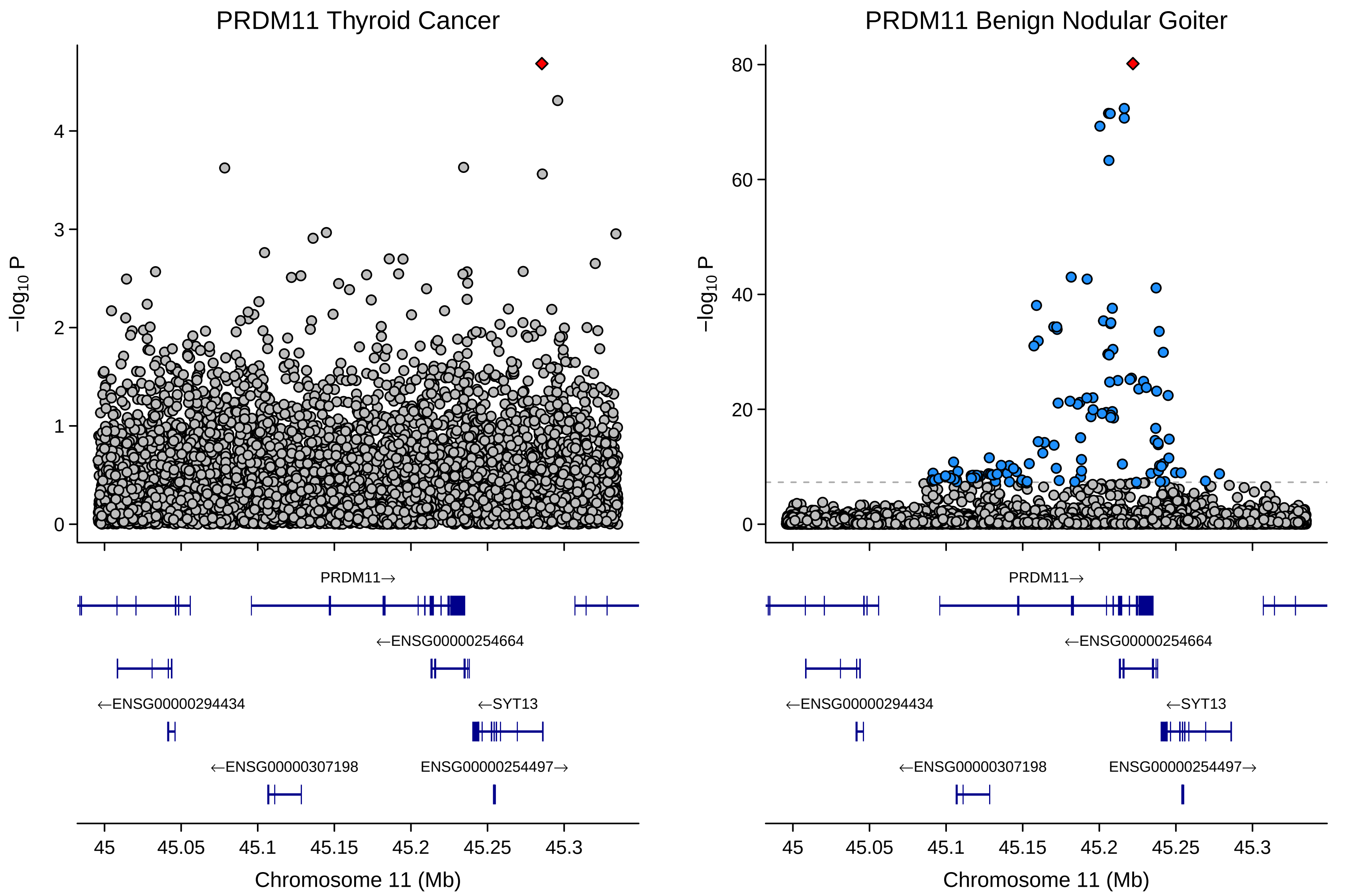

PRDM11 Thyroid Cancer
PRDM11 Benign Nodular Goiter
−log10 P
0
1
2
3
4
20
40
60
80
PRDM11→
←ENSG00000254664
←ENSG00000294434
←SYT13
←ENSG00000307198
ENSG00000254497→
45
45.05
45.1
45.15
45.2
45.25
45.3
Chromosome 11 (Mb)

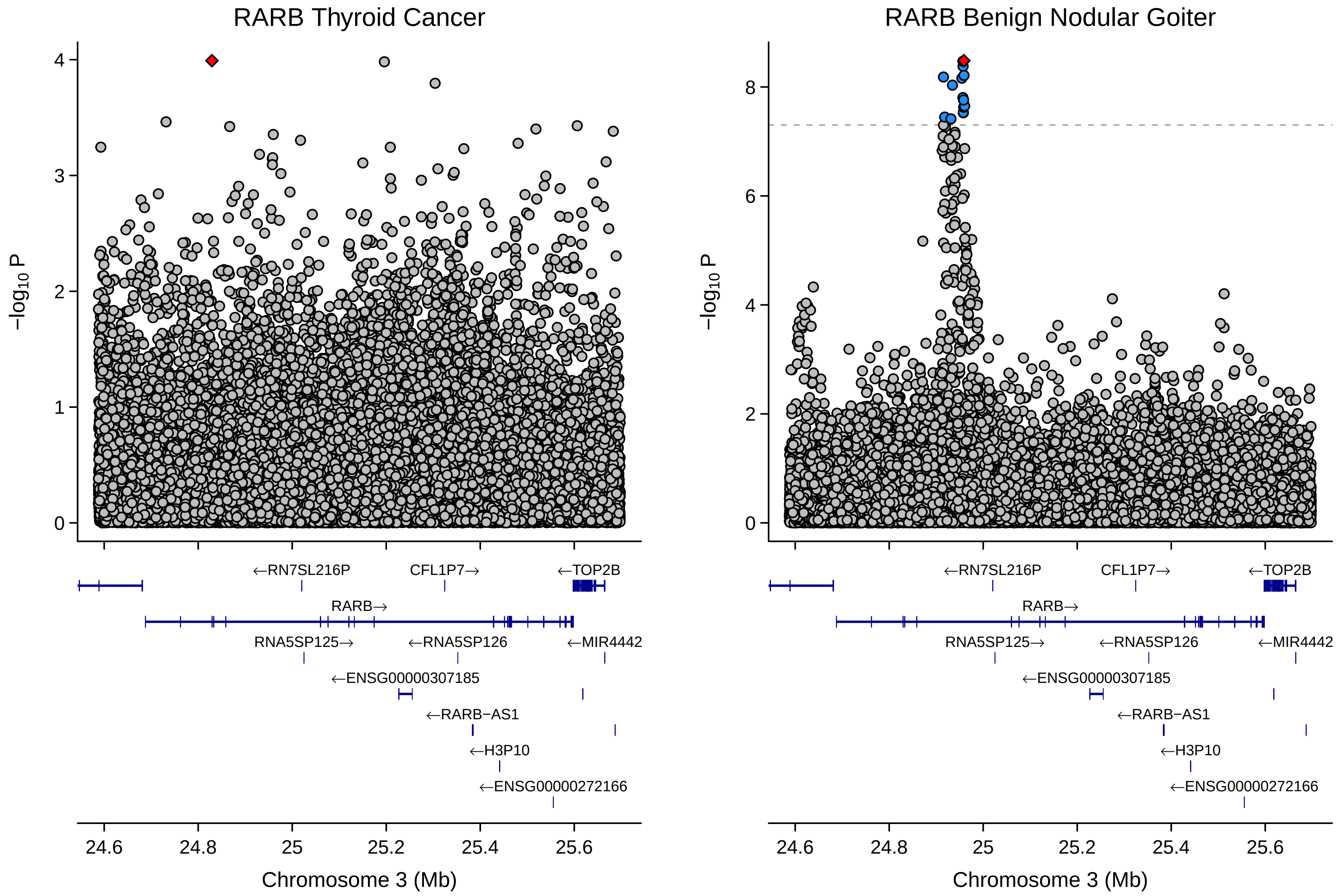

RARB Thyroid Cancer
RARB Benign Nodular Goiter
−log10 P
←RN7SL216P
CFL1P7→
←TOP2B
RARB→
RNA5SP125→
←RNA5SP126
←MIR4442
←ENSG00000307185
←RARB−AS1
←H3P10
←ENSG00000272166
24.6
24.8
25
25.2
25.4
25.6
Chromosome 3 (Mb)

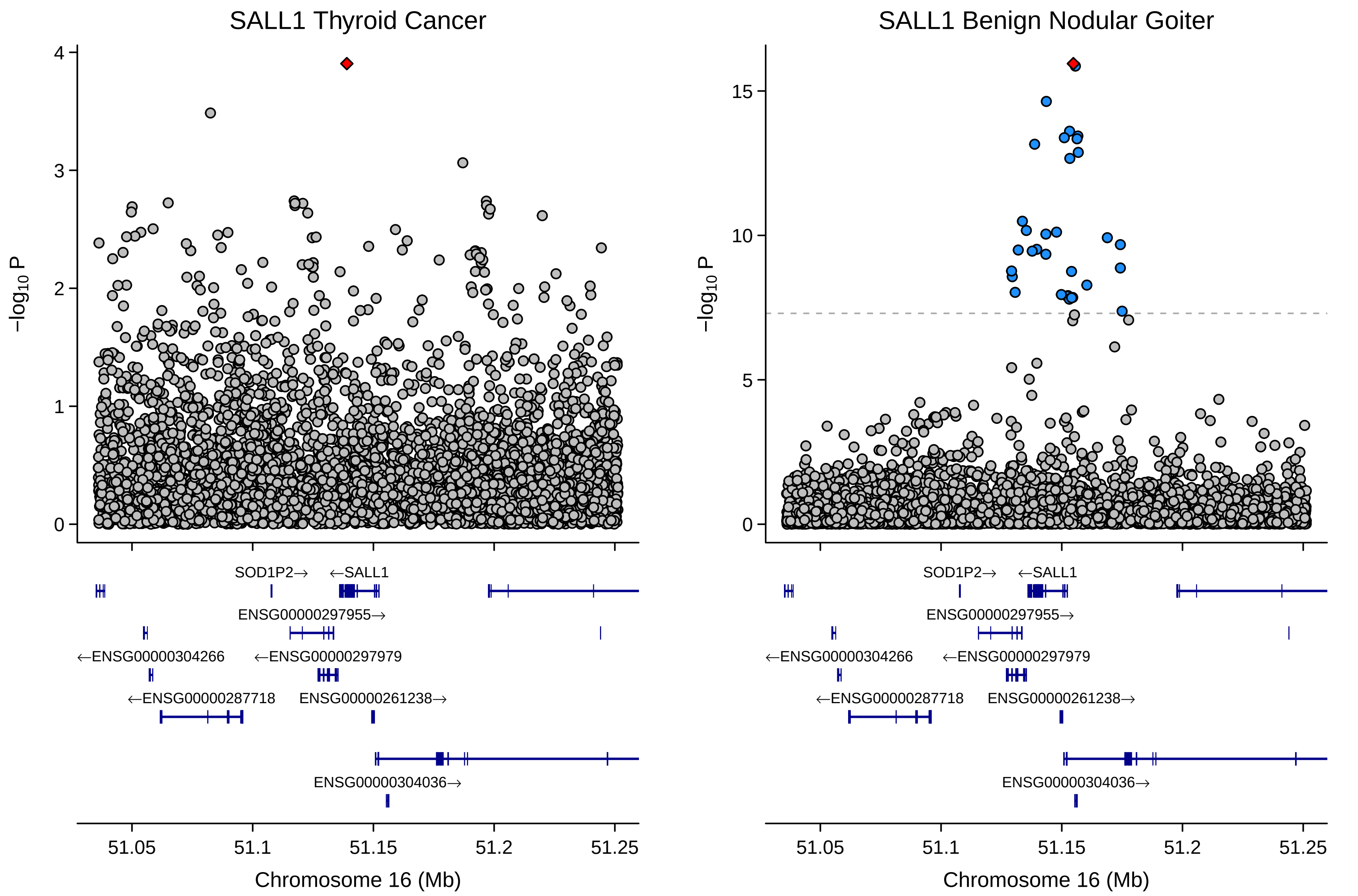

SALL1 Thyroid Cancer
SALL1 Benign Nodular Goiter
−log10 P
Chromosome 16 (Mb)
51.05
51.1
51.15
51.2
51.25
SOD1P2→
←SALL1
ENSG00000297955→
←ENSG00000304266
←ENSG00000297979
←ENSG00000287718
ENSG00000261238→
ENSG00000304036→

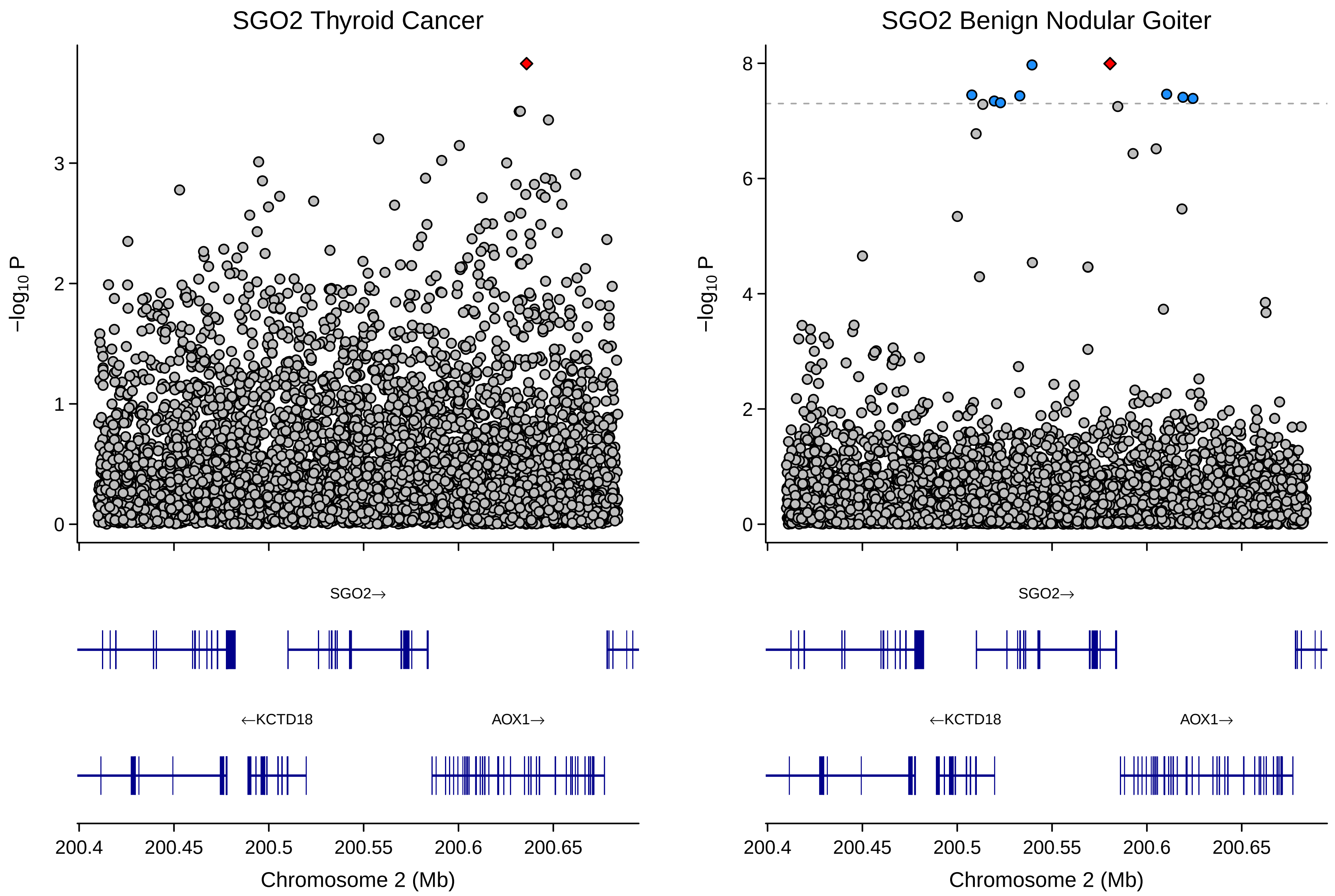

SGO2 Thyroid Cancer
SGO2 Benign Nodular Goiter
−log10 P
0
1
2
3
0
2
4
6
8
SGO2→
←KCTD18
AOX1→
200.4
200.45
200.5
200.55
200.6
200.65
Chromosome 2 (Mb)

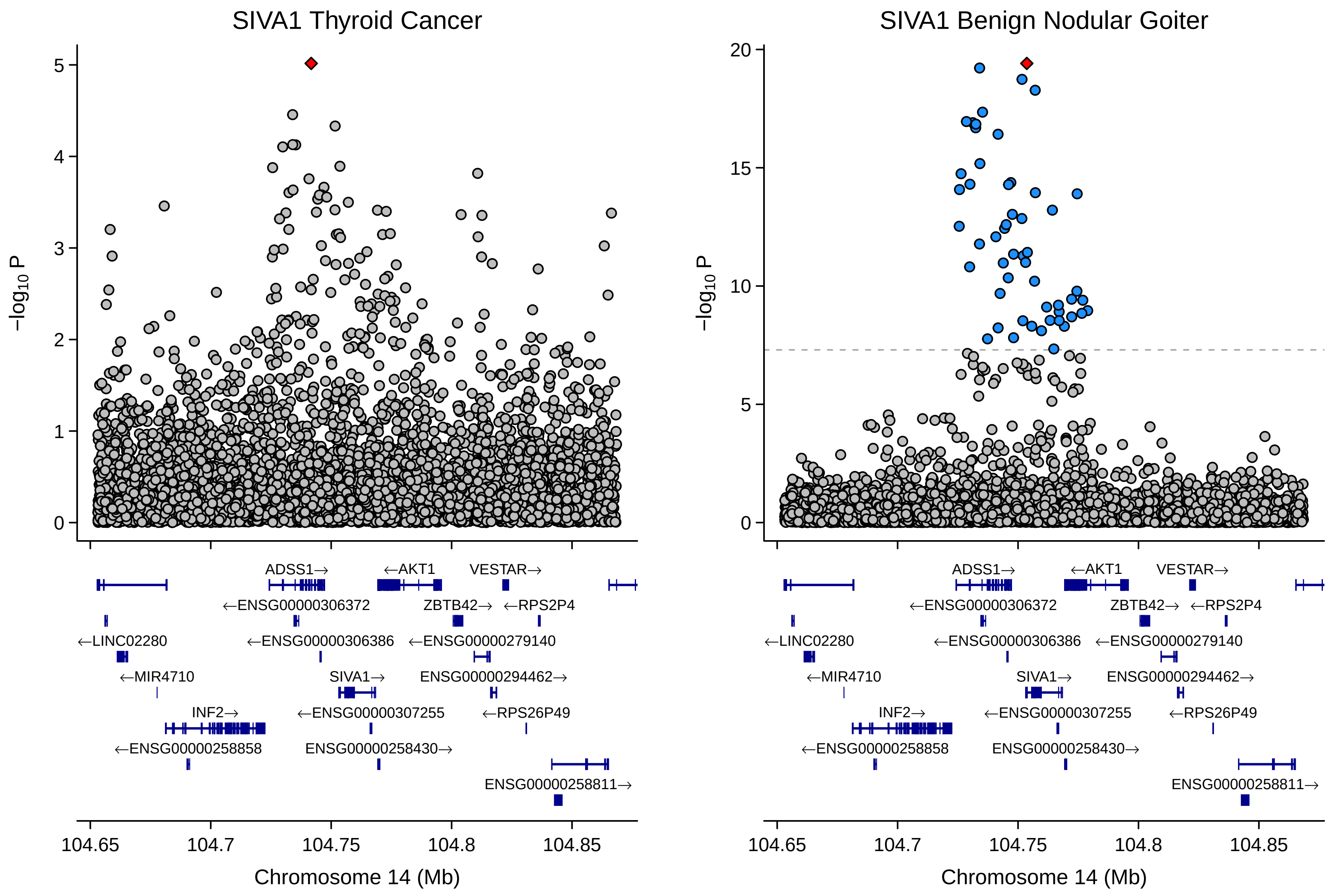
SIVA1 Thyroid Cancer
SIVA1 Benign Nodular Goiter
$-\log_{10}$ P
Chromosome 14 (Mb)
104.65
104.7
104.75
104.8
104.85
ADSS1→
←AKT1
VESTAR→
←ENSG00000306372
ZBTB42→
←RPS2P4
←LINC02280
←ENSG00000306386
←ENSG00000279140
←MIR4710
SIVA1→
ENSG00000294462→
INF2→
←ENSG00000307255
←RPS26P49
←ENSG00000258858
ENSG00000258430→
ENSG00000258811→

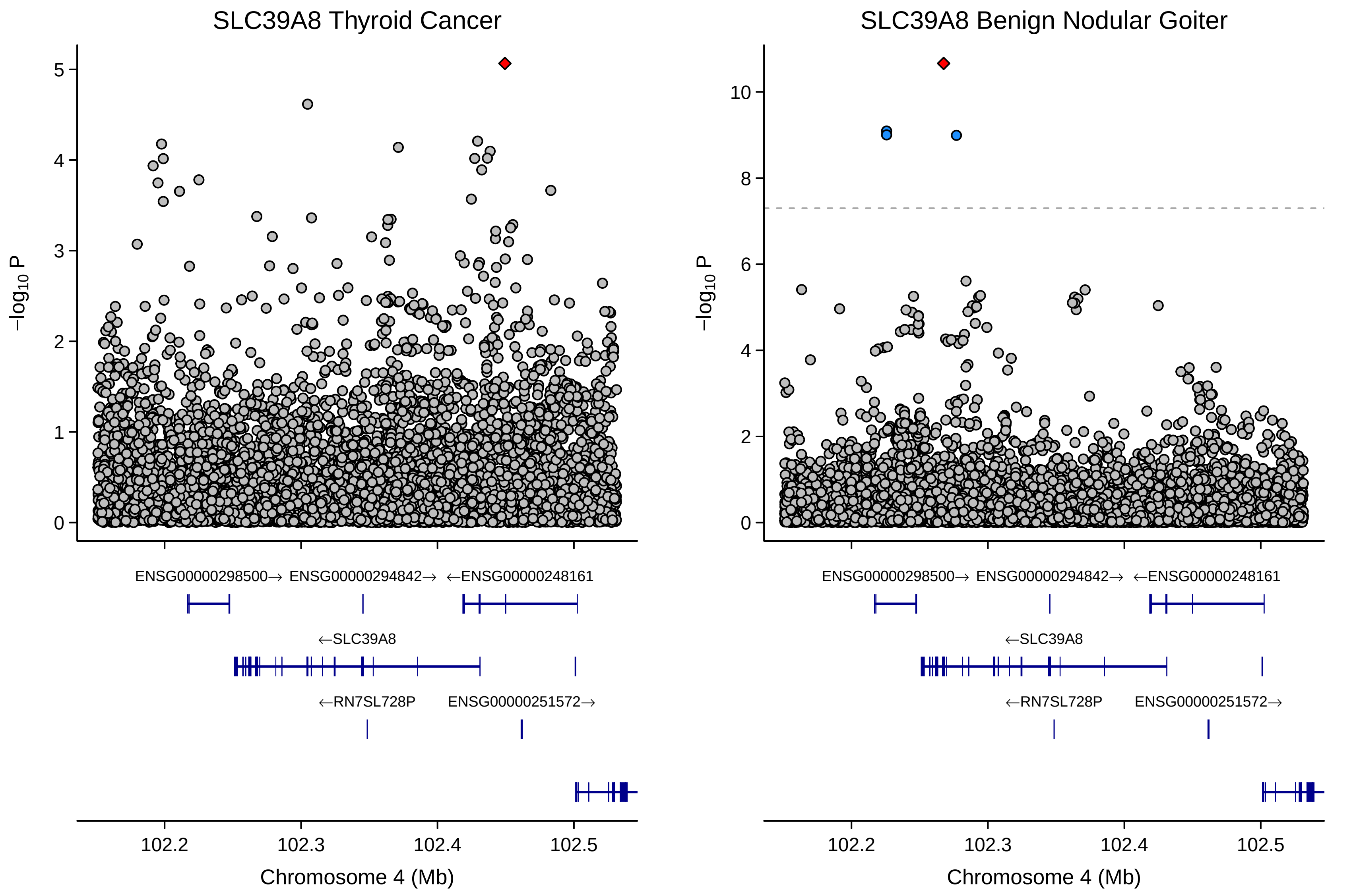

SLC39A8 Thyroid Cancer
SLC39A8 Benign Nodular Goiter
−log10 P
ENSG00000298500→
ENSG00000294842→
←ENSG00000248161
←SLC39A8
←RN7SL728P
ENSG00000251572→
102.2
102.3
102.4
102.5
Chromosome 4 (Mb)

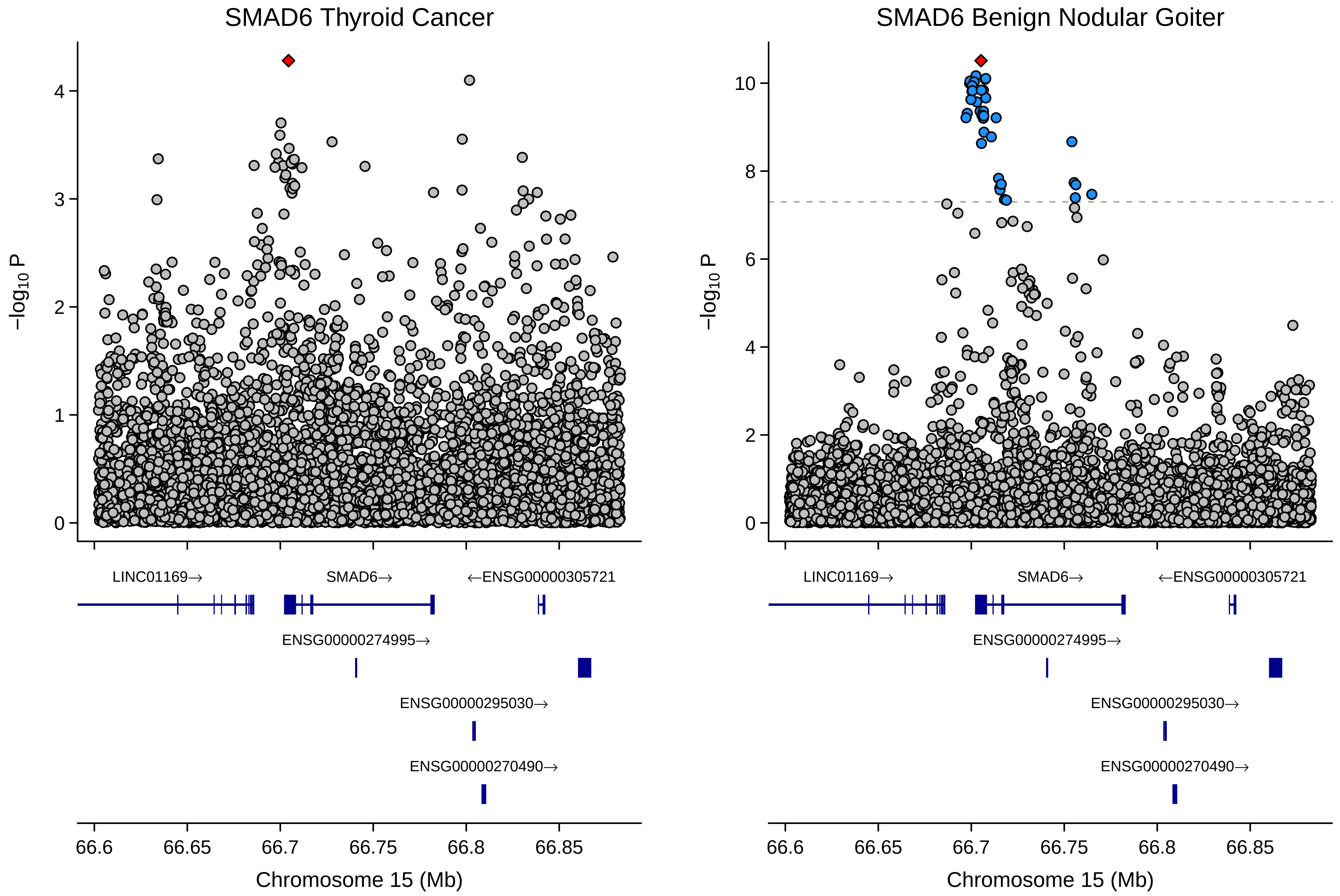
SMAD6 Thyroid Cancer
SMAD6 Benign Nodular Goiter
−log10 P
LINC01169→
SMAD6→
←ENSG00000305721
ENSG00000274995→
ENSG00000295030→
ENSG00000270490→
66.6
66.65
66.7
66.75
66.8
66.85
Chromosome 15 (Mb)

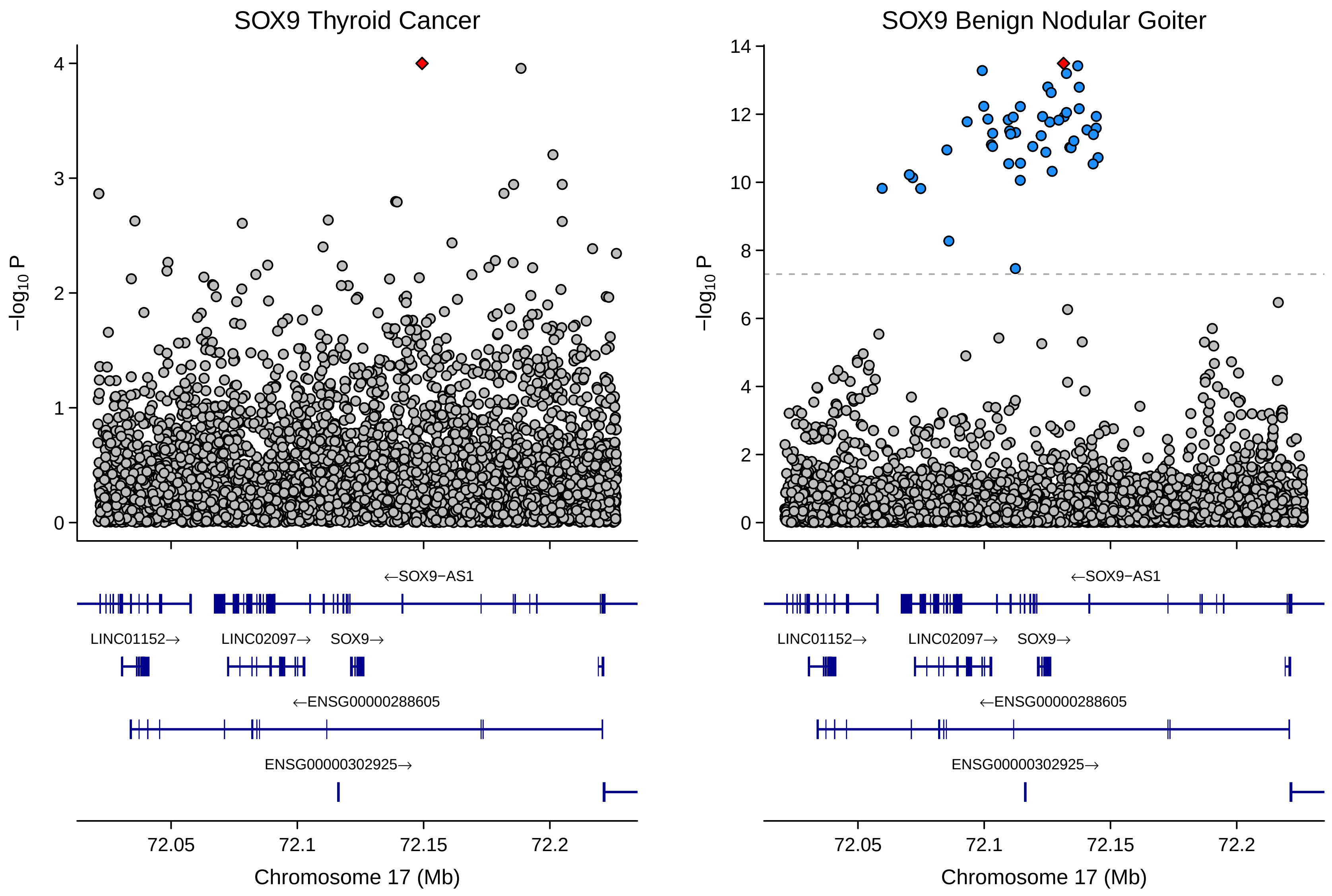
SOX9 Thyroid Cancer
SOX9 Benign Nodular Goiter
−log10 P
←SOX9−AS1
LINC01152→
LINC02097→
SOX9→
←ENSG00000288605
ENSG00000302925→
72.05
72.1
72.15
72.2
Chromosome 17 (Mb)

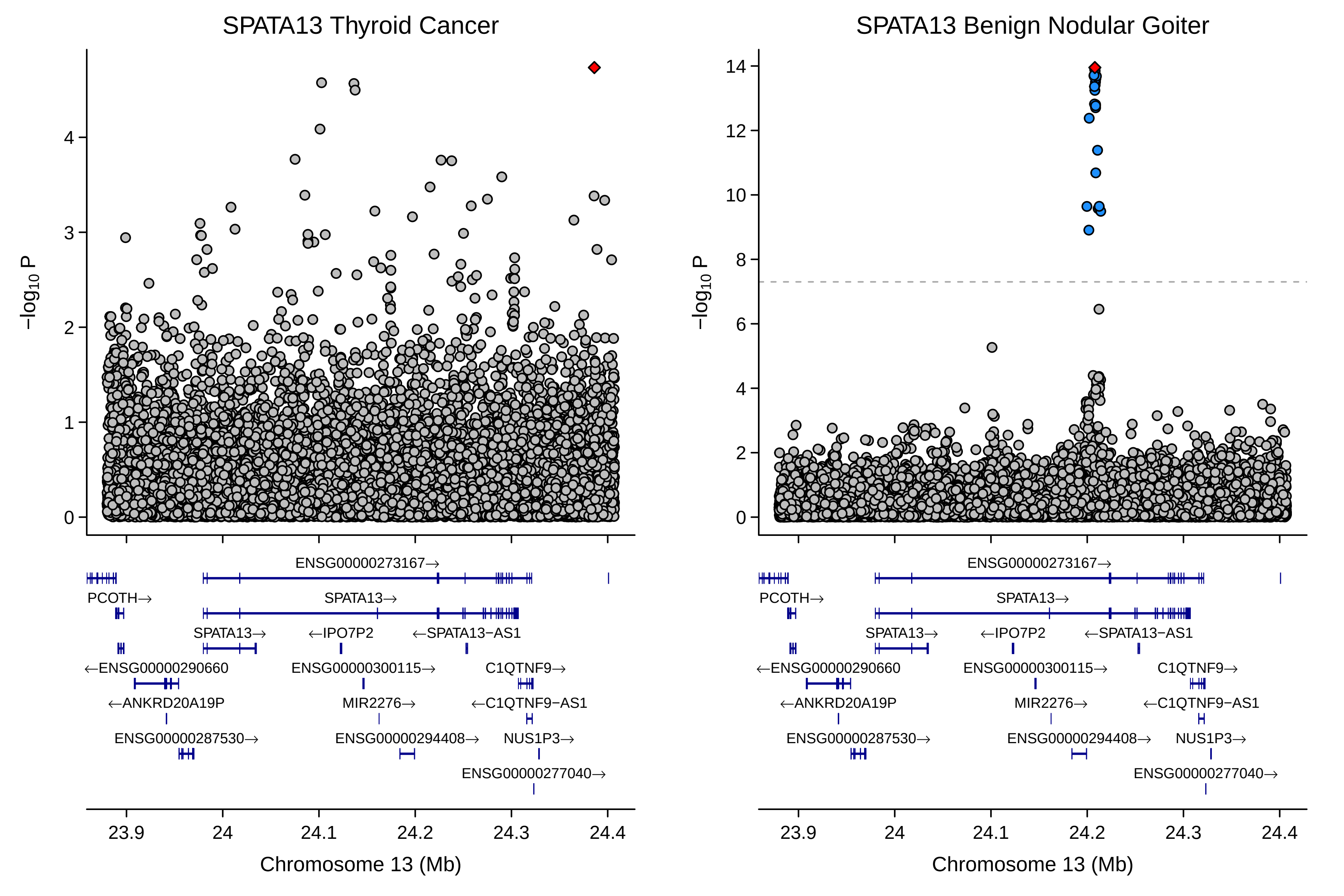
SPATA13 Thyroid Cancer
SPATA13 Benign Nodular Goiter
−log10 P
Chromosome 13 (Mb)
23.9
24
24.1
24.2
24.3
24.4
ENSG00000273167→
PCOTH→
SPATA13→
SPATA13→
←IPO7P2
←SPATA13−AS1
←ENSG00000290660
ENSG00000300115→
C1QTNF9→
←ANKRD20A19P
MIR2276→
←C1QTNF9−AS1
ENSG00000287530→
ENSG00000294408→
NUS1P3→
ENSG00000277040→

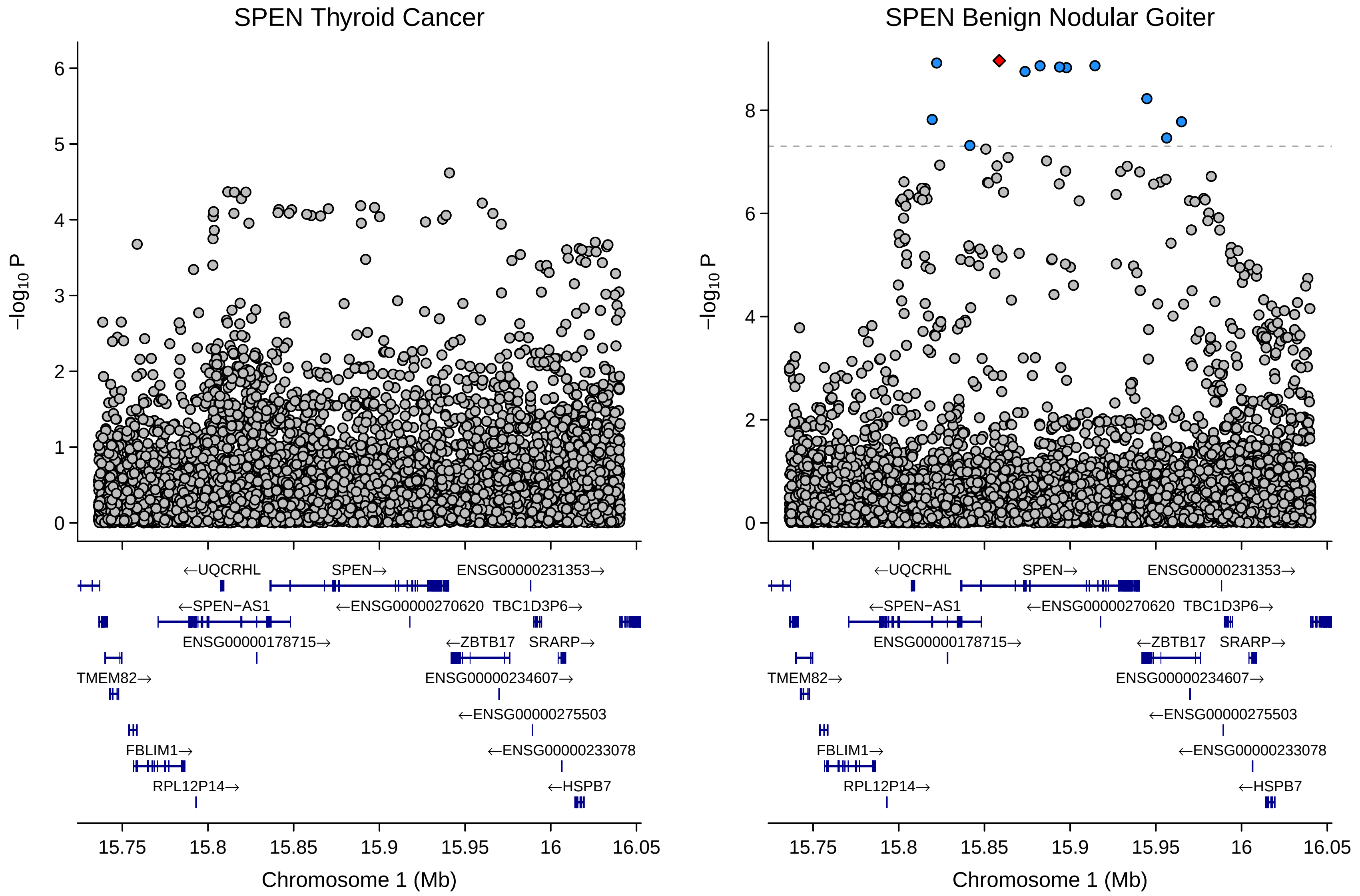

SPEN Thyroid Cancer
SPEN Benign Nodular Goiter
−log10 P
Chromosome 1 (Mb)
15.75
15.8
15.85
15.9
15.95
16
16.05
←UQCRHL
SPEN→
ENSG00000231353→
←SPEN−AS1
←ENSG00000270620
TBC1D3P6→
ENSG00000178715→
←ZBTB17
SRARP→
TMEM82→
ENSG00000234607→
←ENSG00000275503
FBLIM1→
←ENSG00000233078
RPL12P14→
←HSPB7

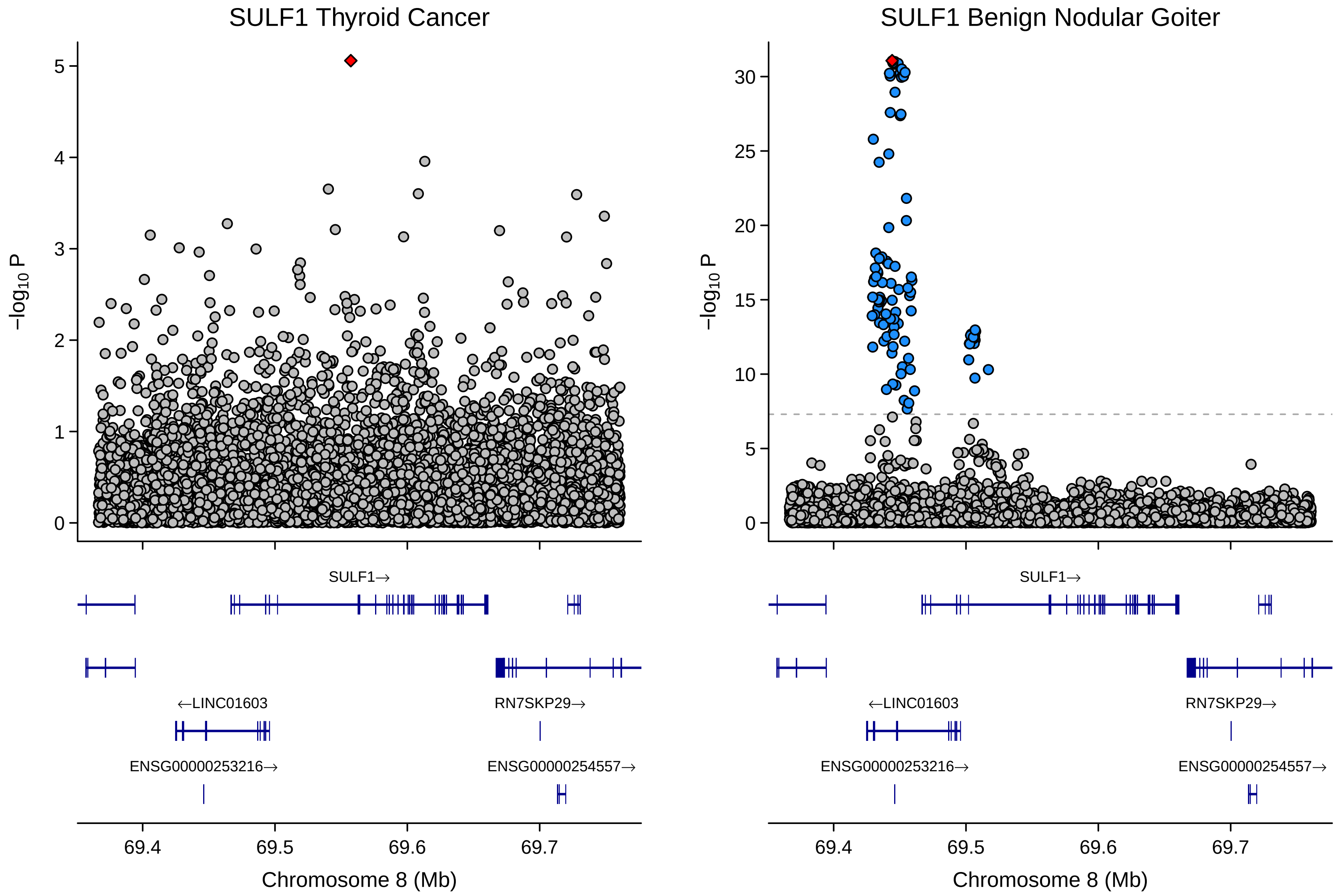

SULF1 Thyroid Cancer
SULF1 Benign Nodular Goiter
−log10 P
0
1
2
3
4
5
10
15
20
25
30
SULF1→
←LINC01603
RN7SKP29→
ENSG00000253216→
ENSG00000254557→
69.4
69.5
69.6
69.7
Chromosome 8 (Mb)

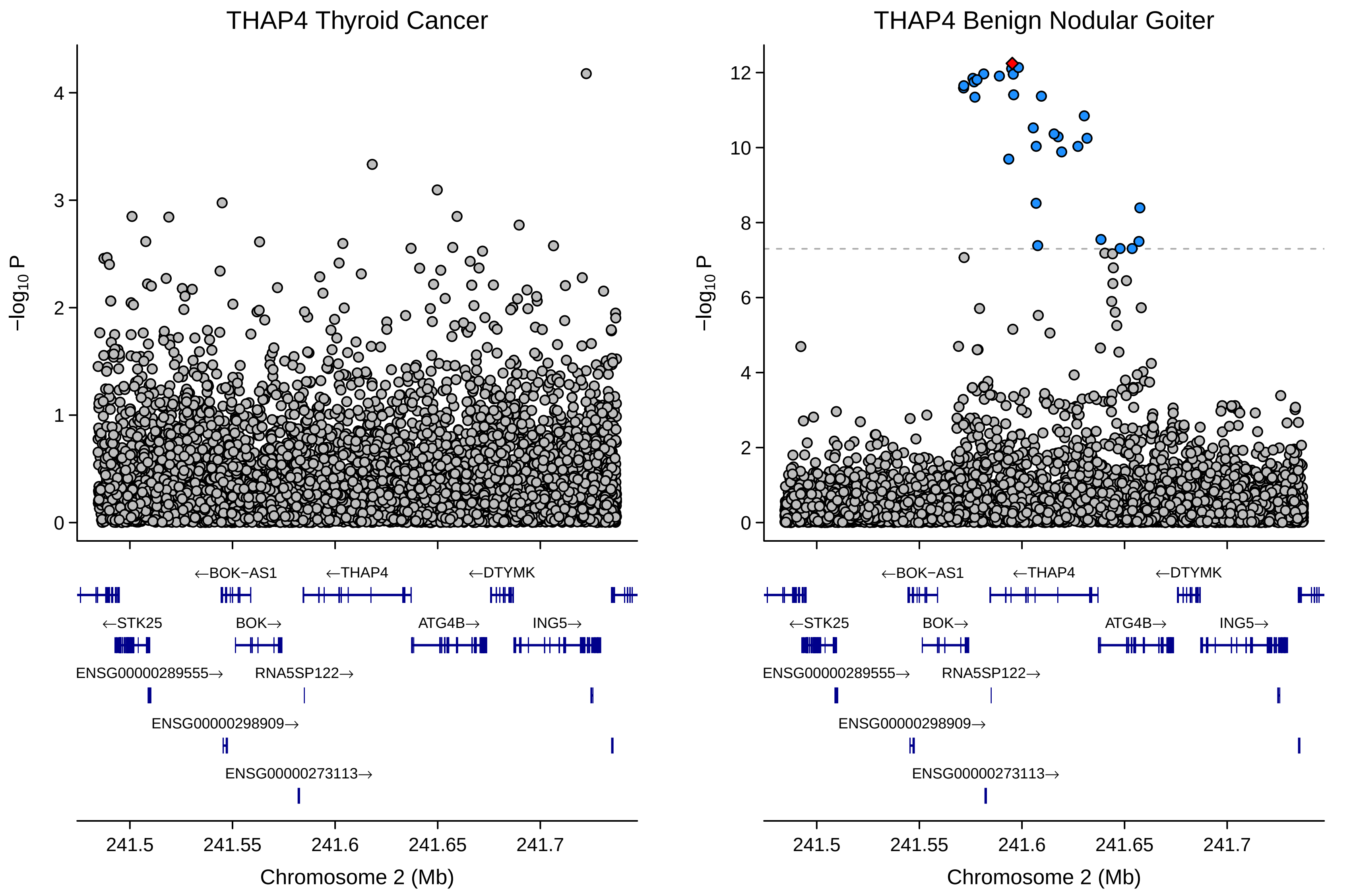
THAP4 Thyroid Cancer
THAP4 Benign Nodular Goiter
−log10 P
0
1
2
3
4
6
8
10
12
←BOK-AS1
←THAP4
←DTYMK
←STK25
BOK→
ATG4B→
ING5→
ENSG00000289555→
RNA5SP122→
ENSG00000298909→
ENSG00000273113→
241.5
241.55
241.6
241.65
241.7
Chromosome 2 (Mb)

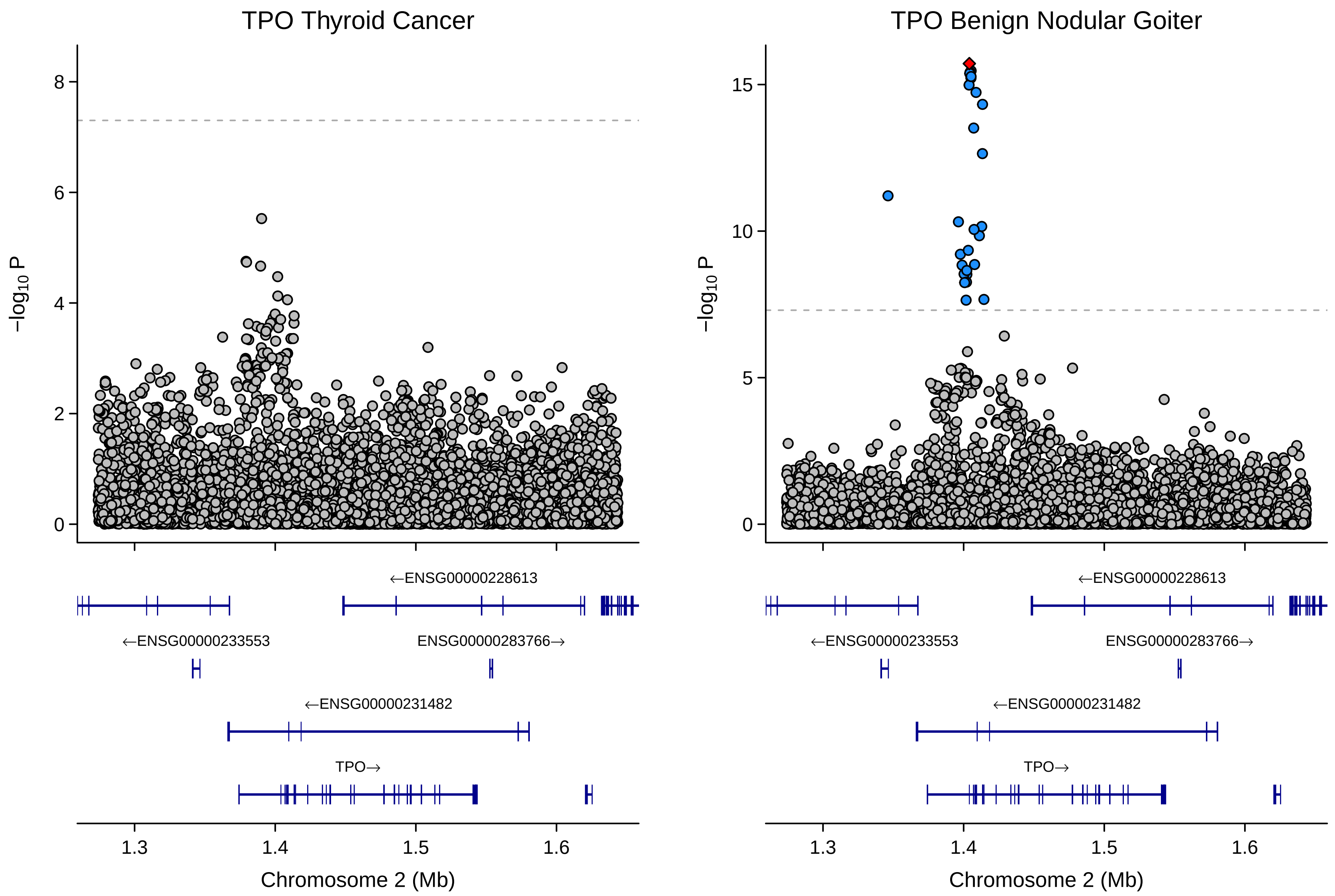

TPO Thyroid Cancer
TPO Benign Nodular Goiter
−log10 P
8
6
4
2
0
15
10
5
←ENSG00000228613
←ENSG00000233553
ENSG00000283766→
←ENSG00000231482
TPO→
1.3
1.4
1.5
1.6
Chromosome 2 (Mb)
