## Supplementary Figure 1.3 for "Global multi-ancestry genetic study elucidates genes and biological pathways associated with thyroid cancer and benign thyroid diseases"

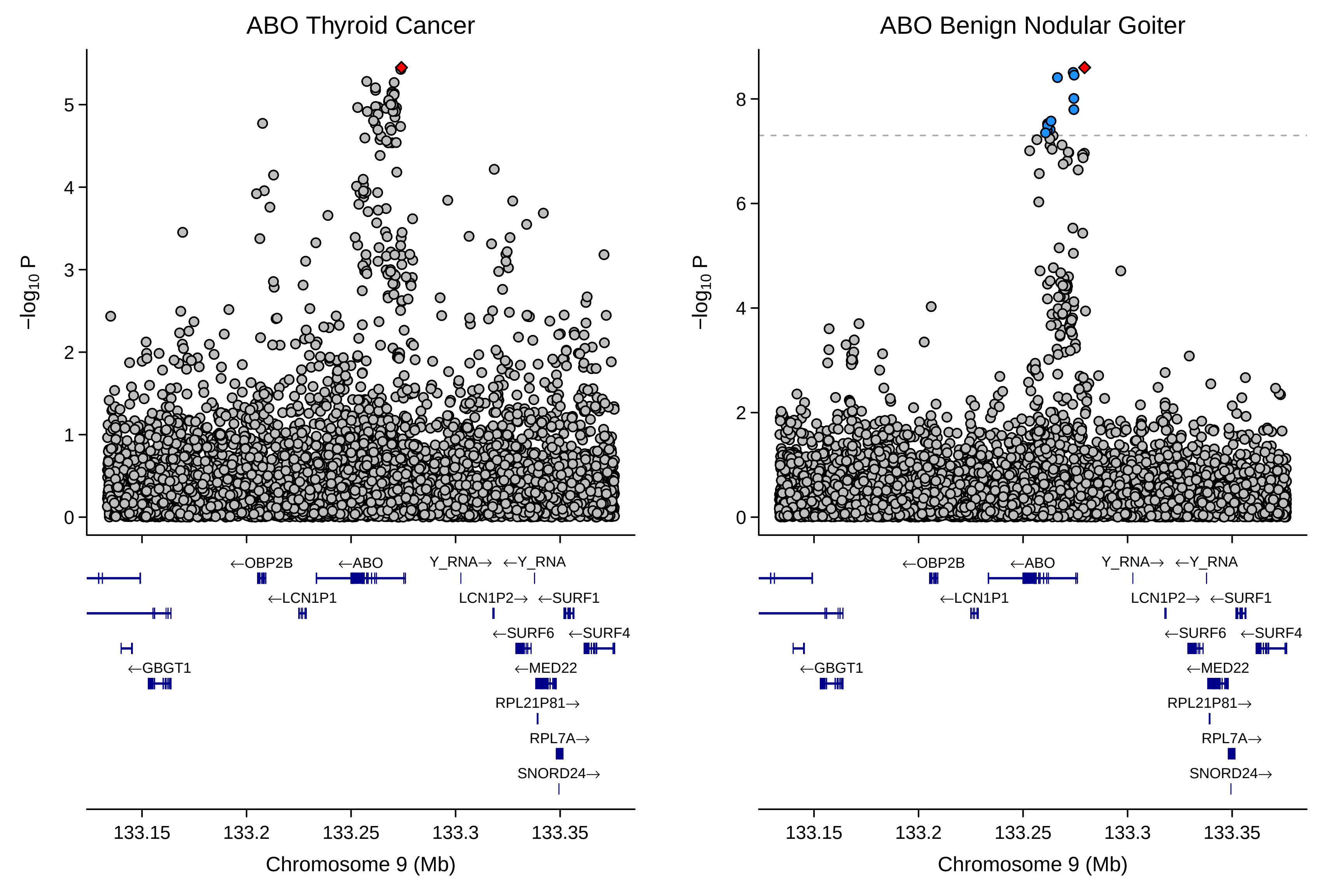

ABO Thyroid Cancer
ABO Benign Nodular Goiter
−log10 P
←OBP2B
←ABO
Y_RNA→
←Y_RNA
←LCN1P1
LCN1P2→
←SURF1
←SURF6
←SURF4
←GBGT1
←MED22
RPL21P81→
RPL7A→
SNORD24→
133.15
133.2
133.25
133.3
133.35
Chromosome 9 (Mb)

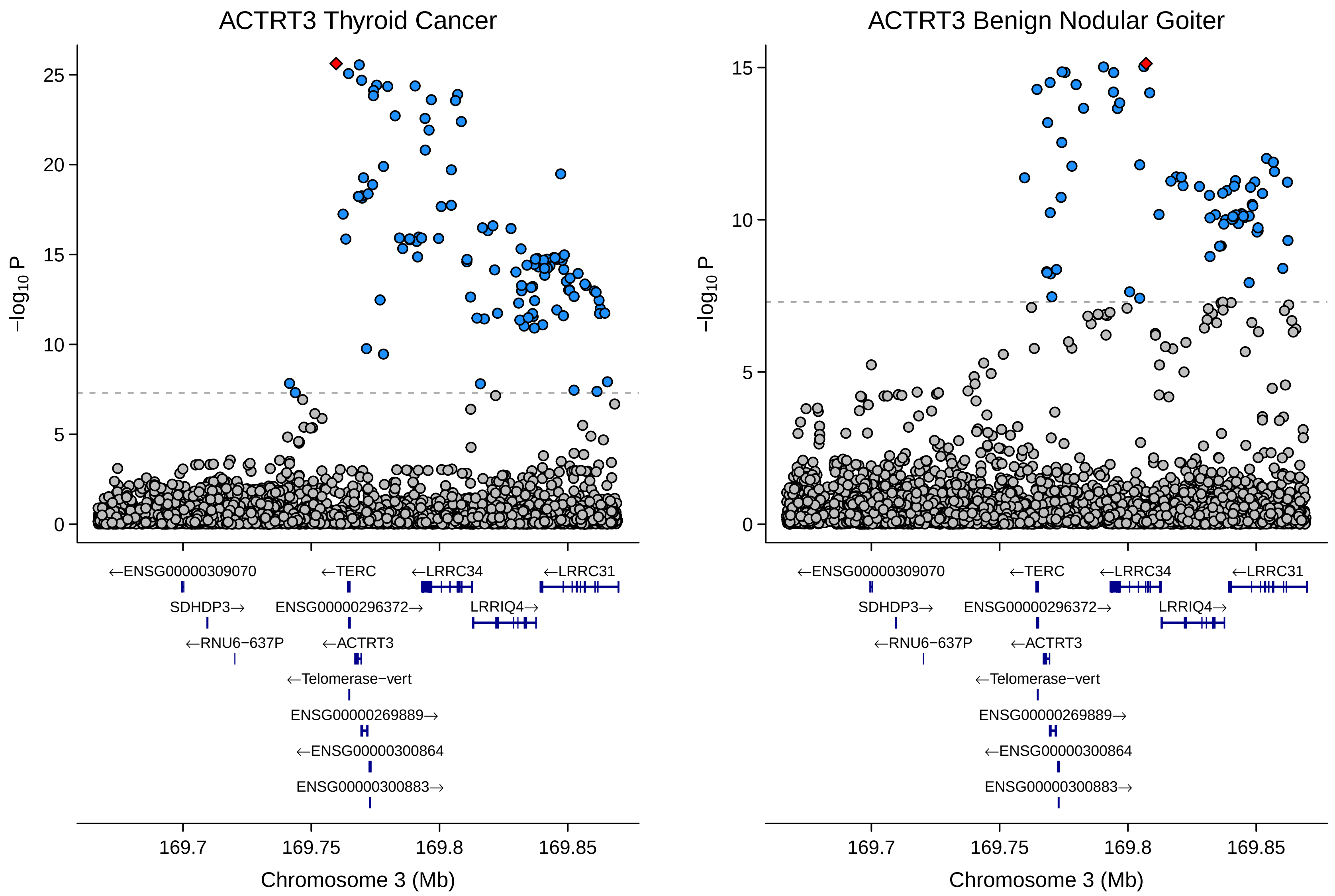

ACTRT3 Thyroid Cancer
ACTRT3 Benign Nodular Goiter
−log10 P
0
5
10
15
20
25
←ENSG00000309070
SDHDP3→
←RNU6−637P
←TERC
ENSG00000296372→
←ACTRT3
←Telomerase−vert
ENSG00000269889→
←ENSG00000300864
ENSG00000300883→
←LRRC34
LRRIQ4→
←LRRC31
169.7
169.75
169.8
169.85
Chromosome 3 (Mb)

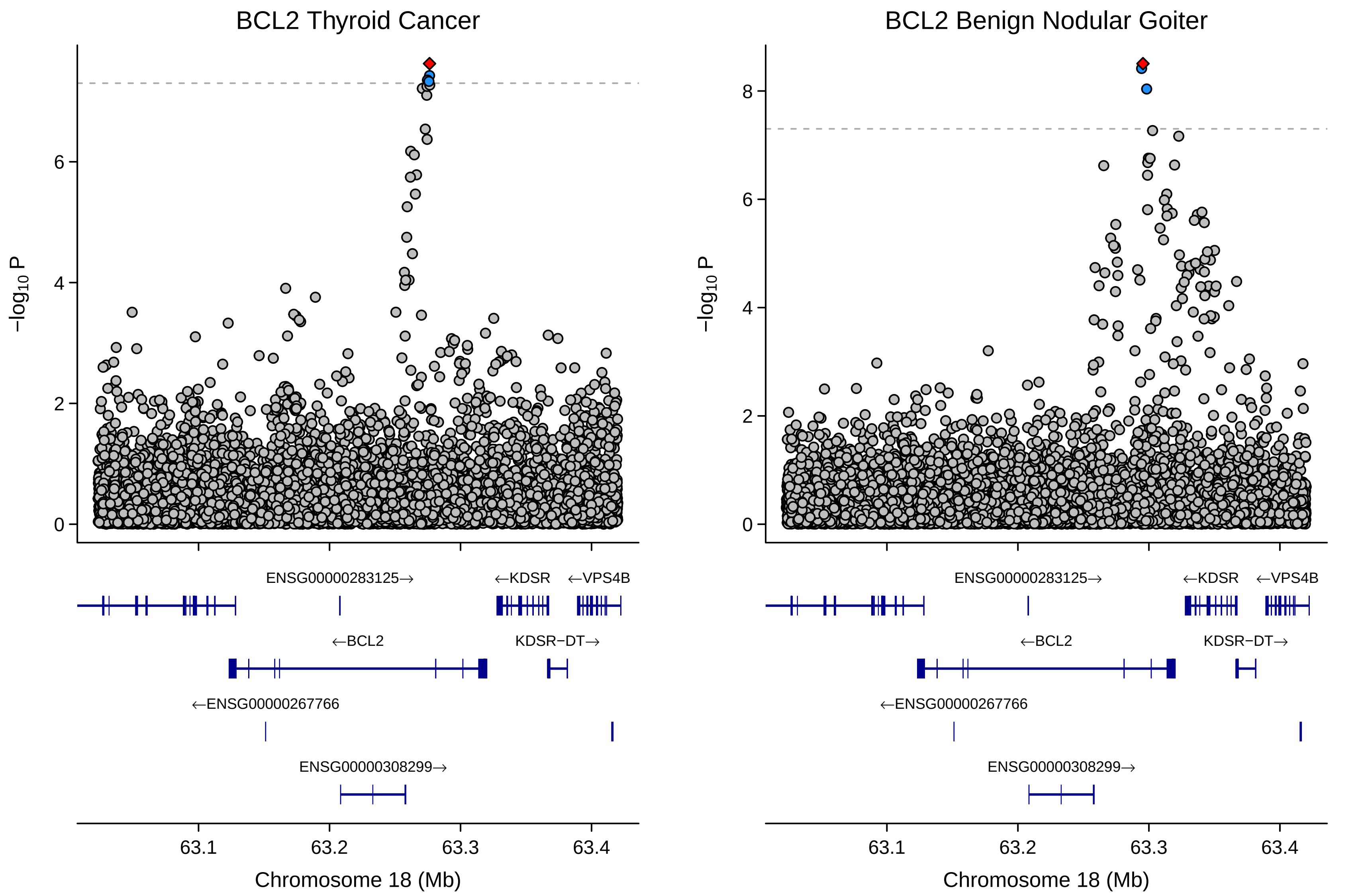
BCL2 Thyroid Cancer
BCL2 Benign Nodular Goiter
−log10 P
0
2
4
6
8
ENSG00000283125→
←KDSR
←VPS4B
←BCL2
KDSR−DT→
←ENSG00000267766
ENSG00000308299→
63.1
63.2
63.3
63.4
Chromosome 18 (Mb)

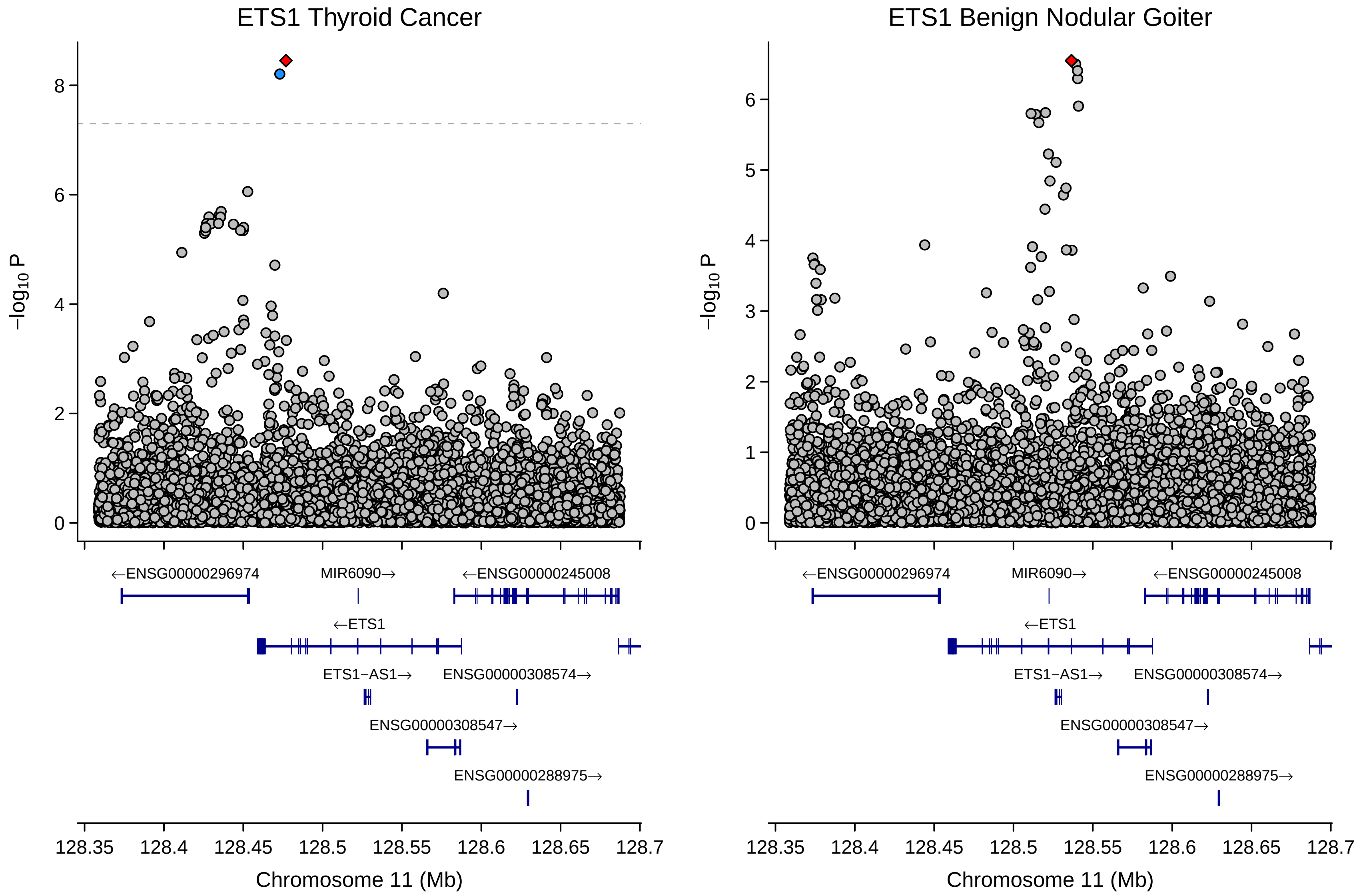
ETS1 Thyroid Cancer
ETS1 Benign Nodular Goiter
−log10 P
Chromosome 11 (Mb)
128.35
128.4
128.45
128.5
128.55
128.6
128.65
128.7
←ENSG00000296974
MIR6090→
←ENSG00000245008
←ETS1
ETS1−AS1→
ENSG00000308574→
ENSG00000308547→
ENSG00000288975→

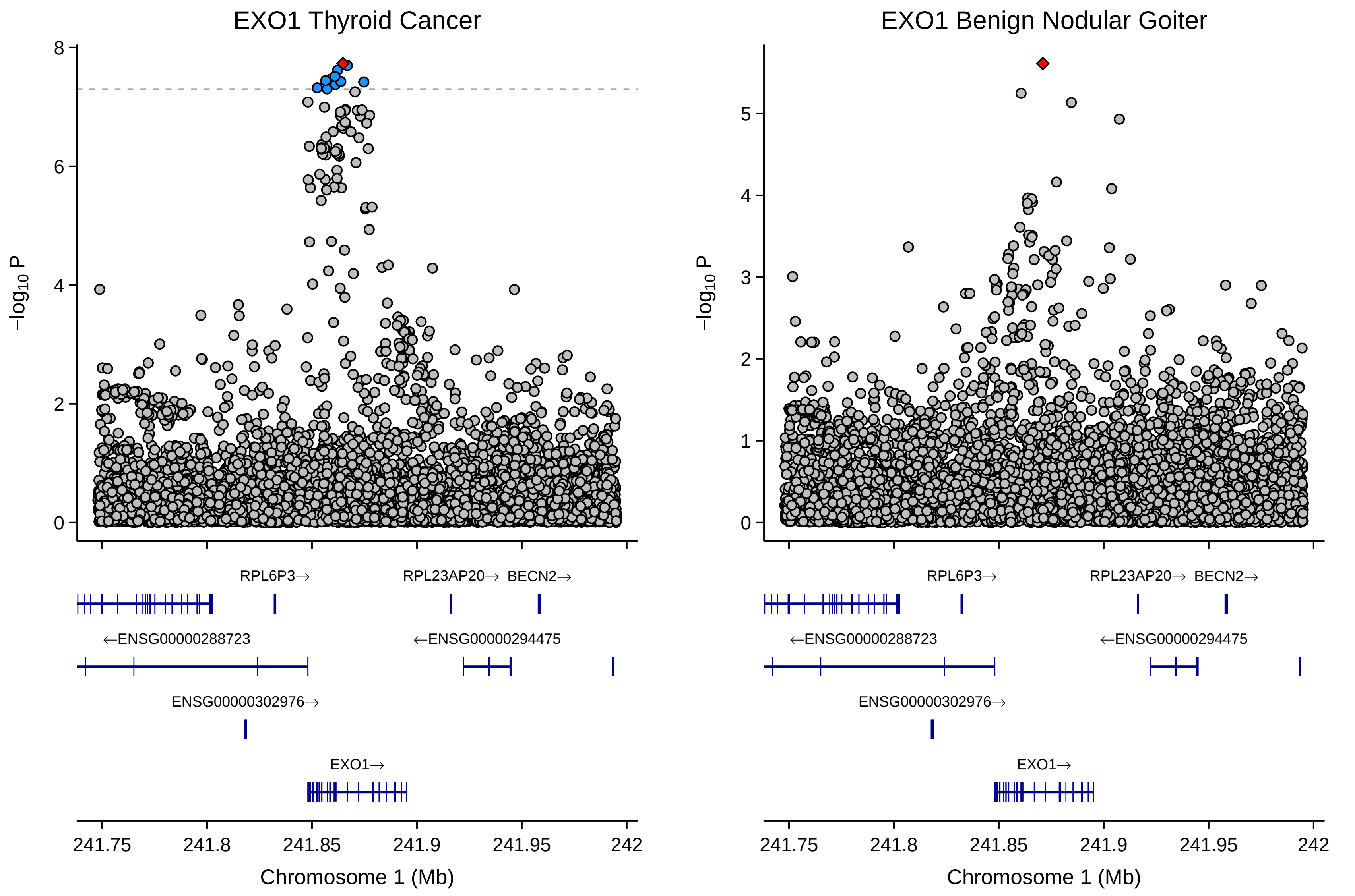
EXO1 Thyroid Cancer
EXO1 Benign Nodular Goiter
−log10 P
RPL6P3→
RPL23AP20→
BECN2→
←ENSG00000288723
←ENSG00000294475
ENSG00000302976→
EXO1→
241.75
241.8
241.85
241.9
241.95
242
Chromosome 1 (Mb)

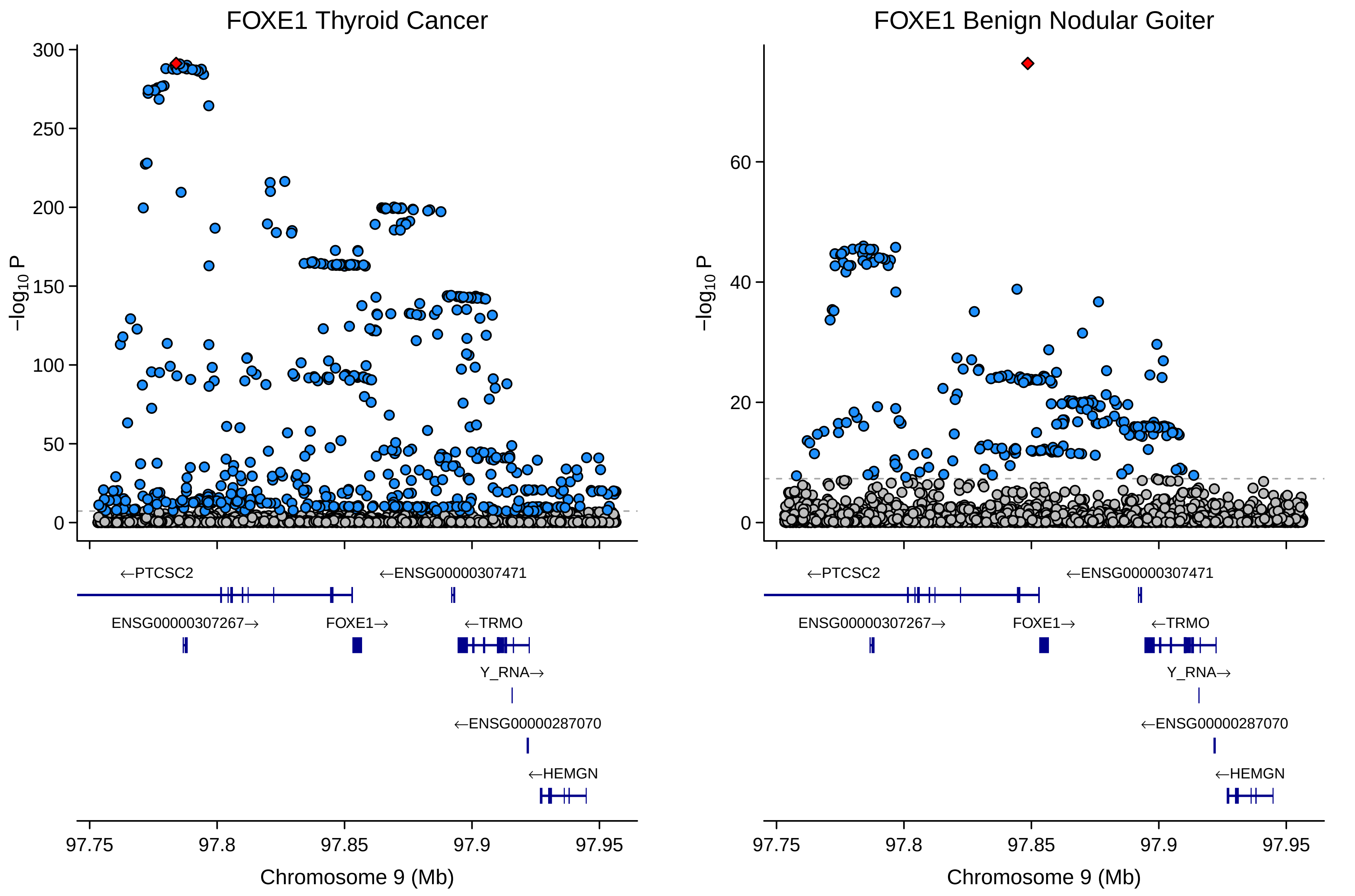
FOXE1 Thyroid Cancer
FOXE1 Benign Nodular Goiter
−log10 P
0
50
100
150
200
250
300
20
40
60
←PTCSC2
←ENSG00000307471
ENSG00000307267→
FOXE1→
←TRMO
Y_RNA→
←ENSG00000287070
←HEMGN
97.75
97.8
97.85
97.9
97.95
Chromosome 9 (Mb)

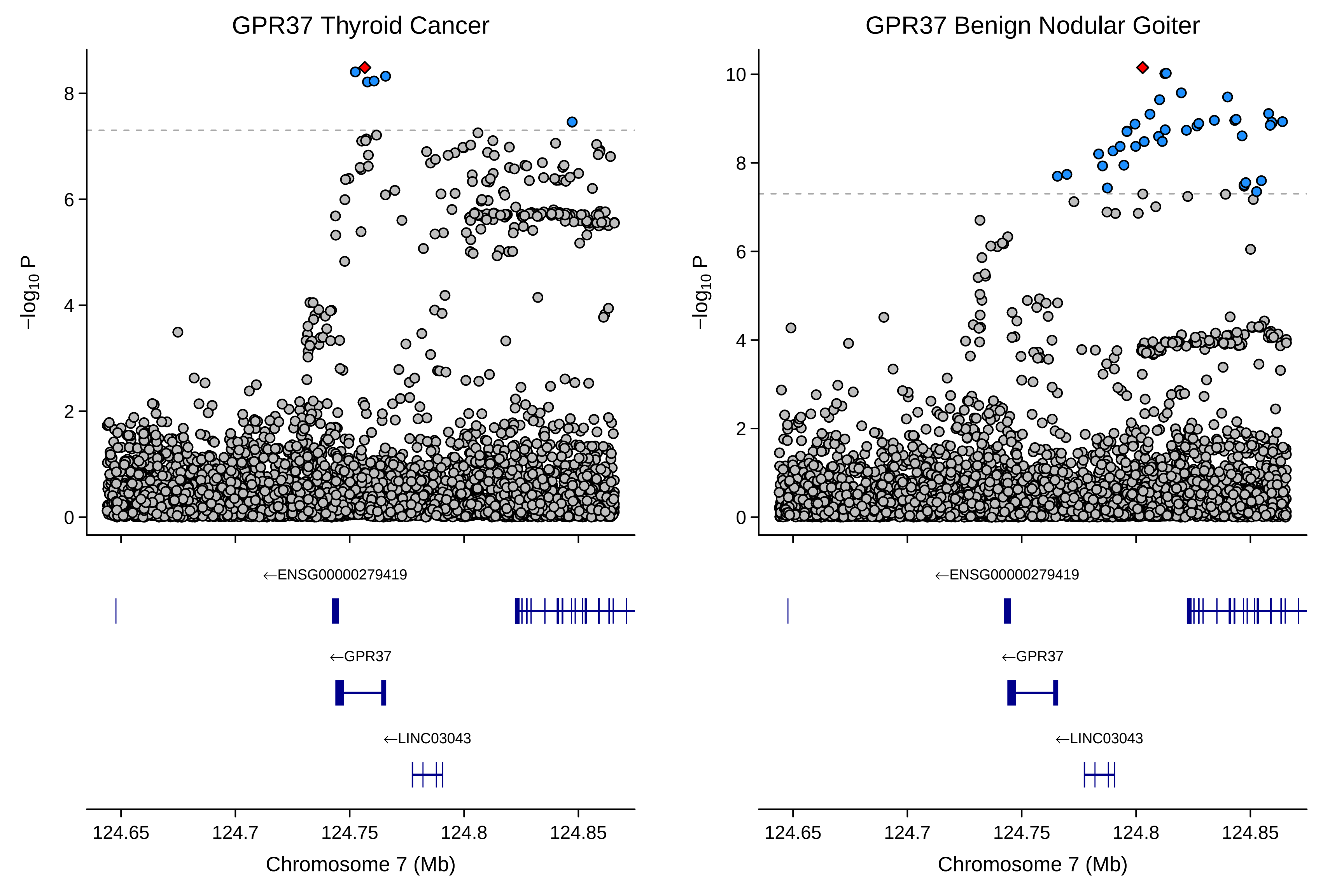

GPR37 Thyroid Cancer
GPR37 Benign Nodular Goiter
−log10 P
0
2
4
6
8
10
←ENSG00000279419
←GPR37
←LINC03043
124.65
124.7
124.75
124.8
124.85
Chromosome 7 (Mb)

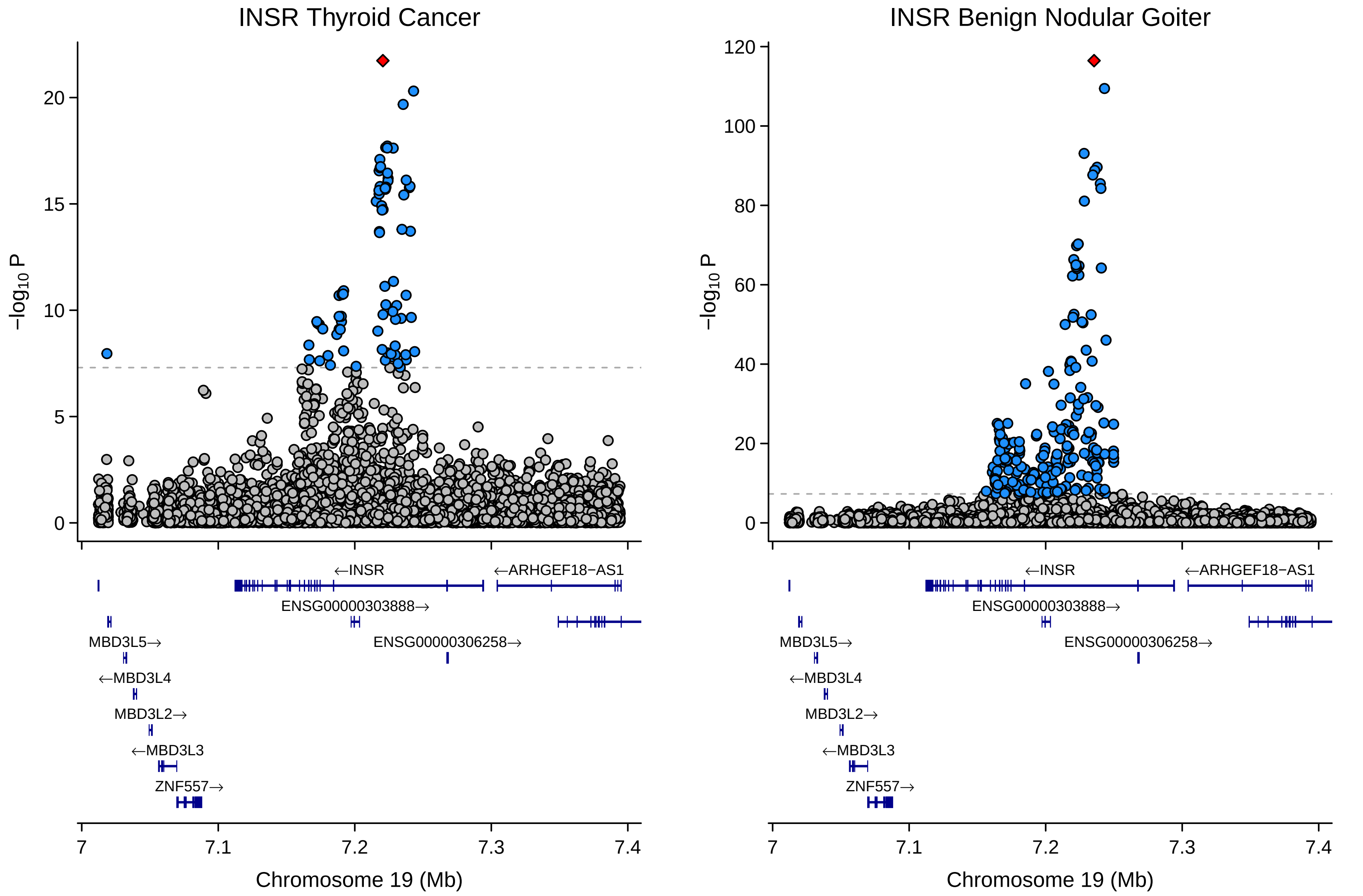

INSR Thyroid Cancer
INSR Benign Nodular Goiter
−log10 P
0
5
10
15
20
40
60
80
100
120
←INSR
←ARHGEF18−AS1
ENSG00000303888→
MBD3L5→
ENSG00000306258→
←MBD3L4
MBD3L2→
←MBD3L3
ZNF557→
7
7.1
7.2
7.3
7.4
Chromosome 19 (Mb)

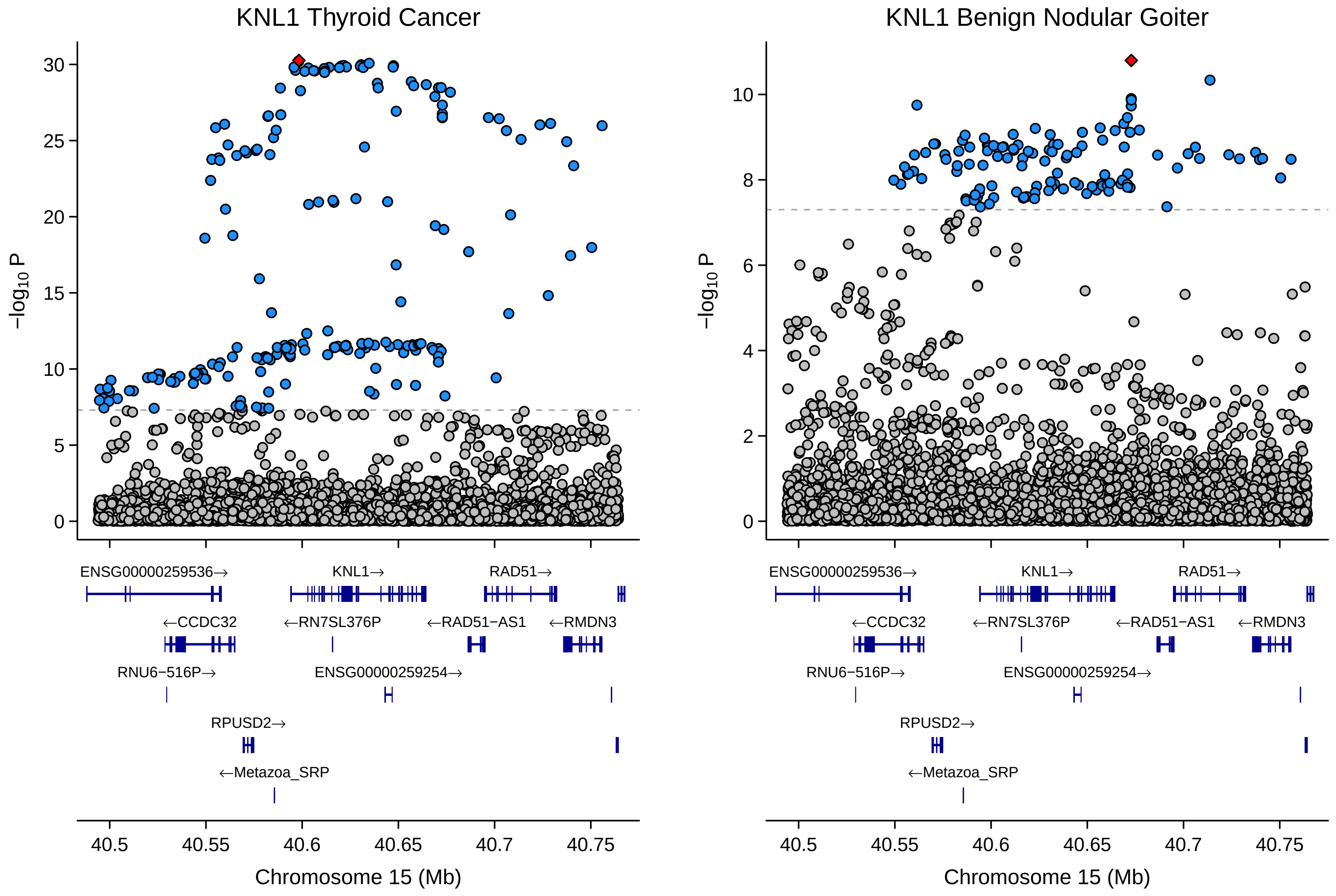
KNL1 Thyroid Cancer
KNL1 Benign Nodular Goiter
−log10 P
ENSG00000259536→
KNL1→
RAD51→
←CCDC32
←RN7SL376P
←RAD51−AS1
←RMDN3
RNU6−516P→
ENSG00000259254→
RPUSD2→
←Metazoa_SRP
Chromosome 15 (Mb)

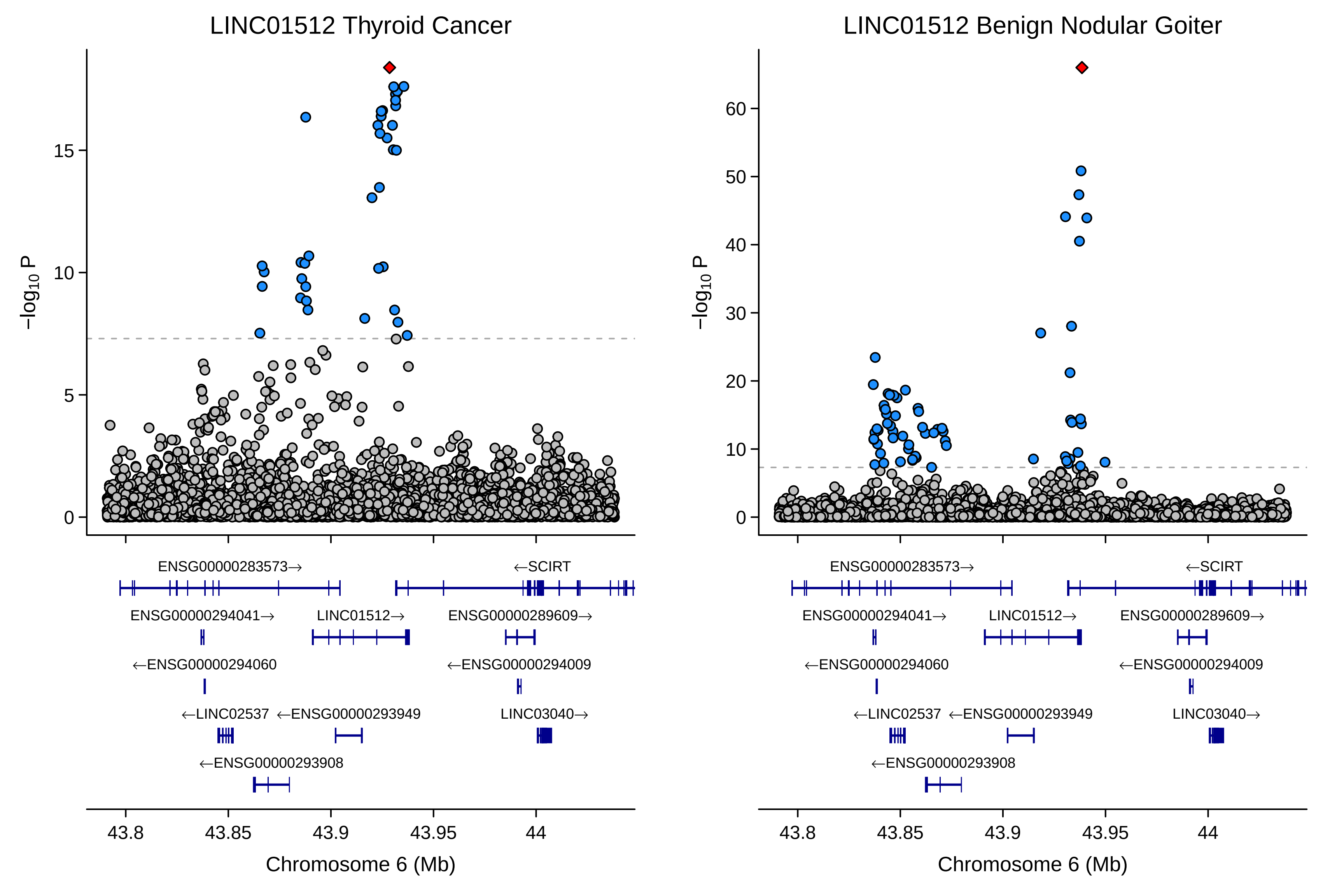

LINC01512 Thyroid Cancer
LINC01512 Benign Nodular Goiter
−log10 P
0
5
10
15
20
30
40
50
60
ENSG00000283573→
←SCIRT
ENSG00000294041→
LINC01512→
ENSG00000289609→
←ENSG00000294060
←ENSG00000294009
←LINC02537
←ENSG00000293949
LINC03040→
←ENSG00000293908
43.8
43.85
43.9
43.95
44
Chromosome 6 (Mb)

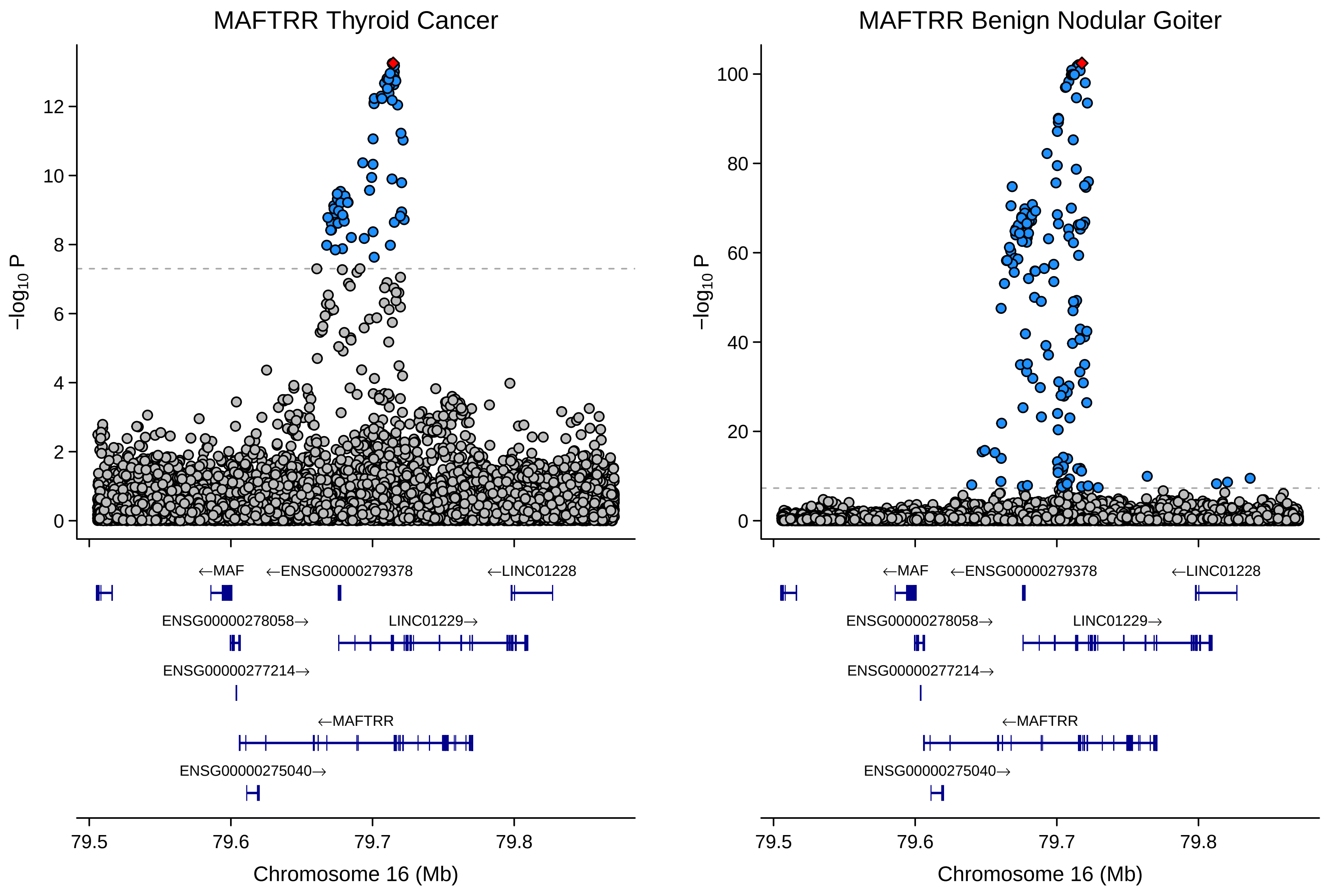
MAFTRR Thyroid Cancer
MAFTRR Benign Nodular Goiter
−log10 P
0
2
4
6
8
10
12
20
40
60
80
100
←MAF
←ENSG00000279378
←LINC01228
ENSG00000278058→
LINC01229→
ENSG00000277214→
←MAFTRR
ENSG00000275040→
79.5
79.6
79.7
79.8
Chromosome 16 (Mb)

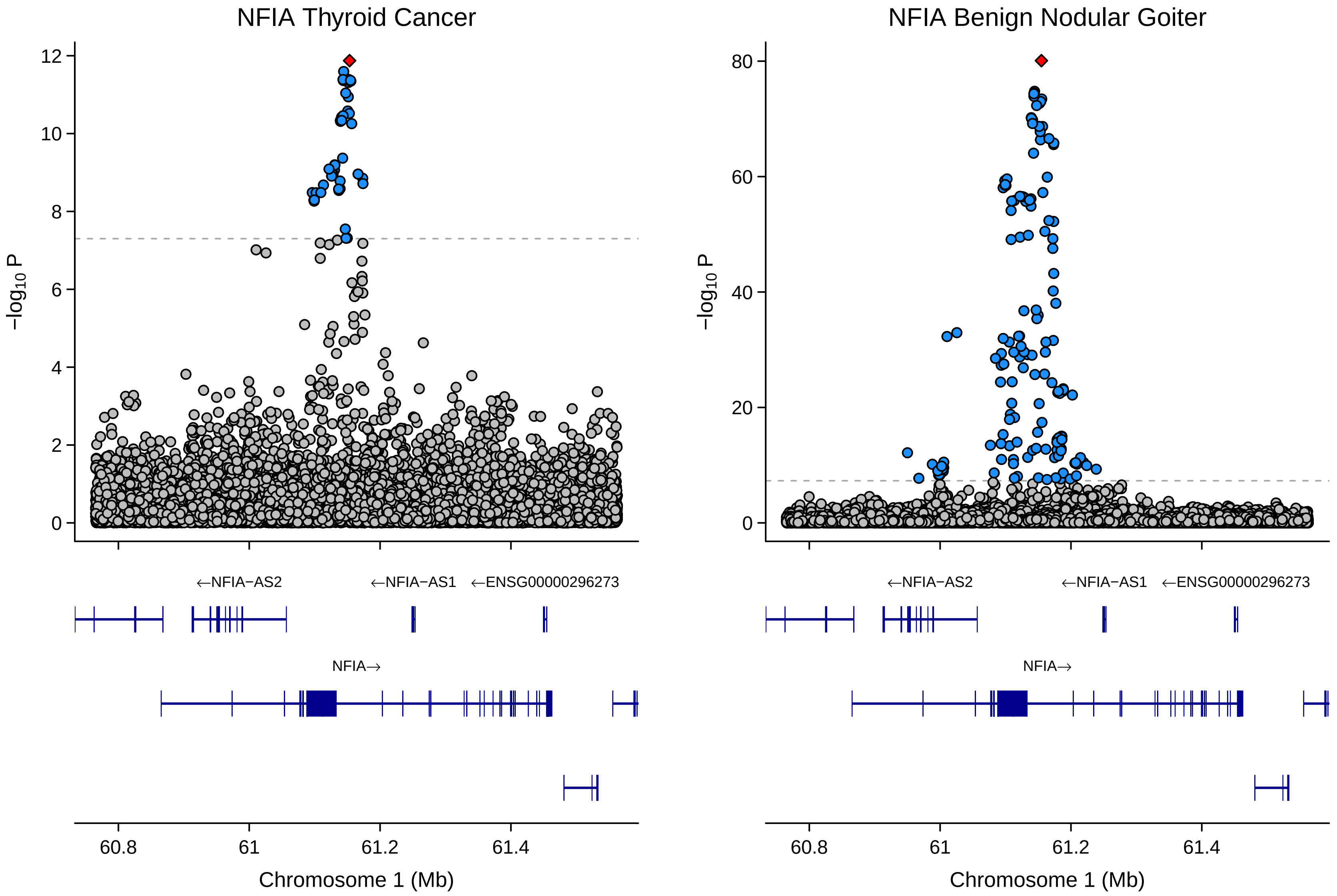

NFIA Thyroid Cancer
NFIA Benign Nodular Goiter
−log10 P
12
10
8
6
4
2
0
80
60
40
20
←NFIA−AS2
←NFIA−AS1
←ENSG00000296273
NFIA→
60.8
61
61.2
61.4
Chromosome 1 (Mb)

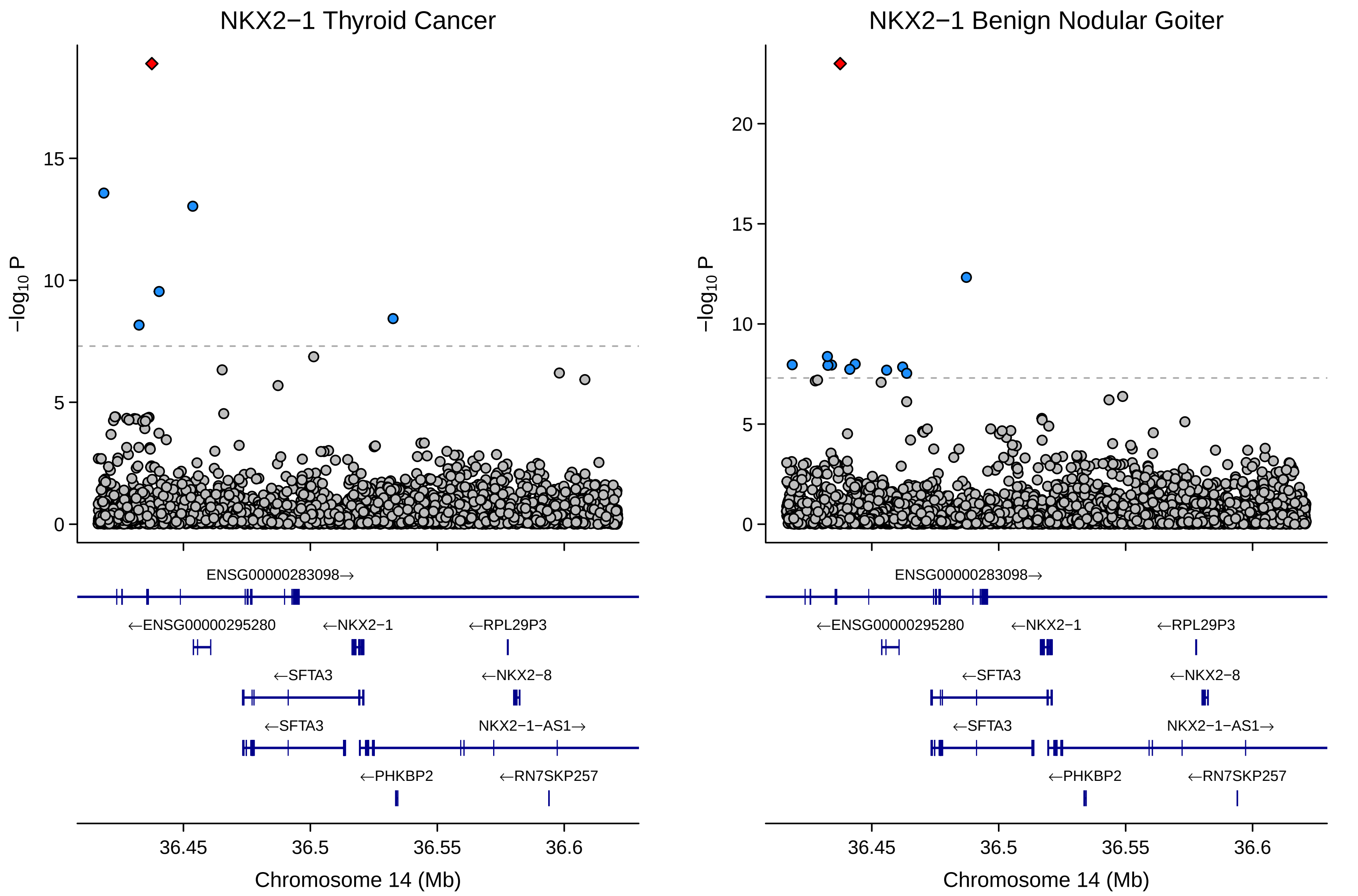

NKX2−1 Thyroid Cancer
NKX2−1 Benign Nodular Goiter
−log10 P
0
5
10
15
20
ENSG00000283098→
←ENSG00000295280
←NKX2−1
←RPL29P3
←SFTA3
←NKX2−8
←SFTA3
NKX2−1−AS1→
←PHKBP2
←RN7SKP257
36.45
36.5
36.55
36.6
Chromosome 14 (Mb)

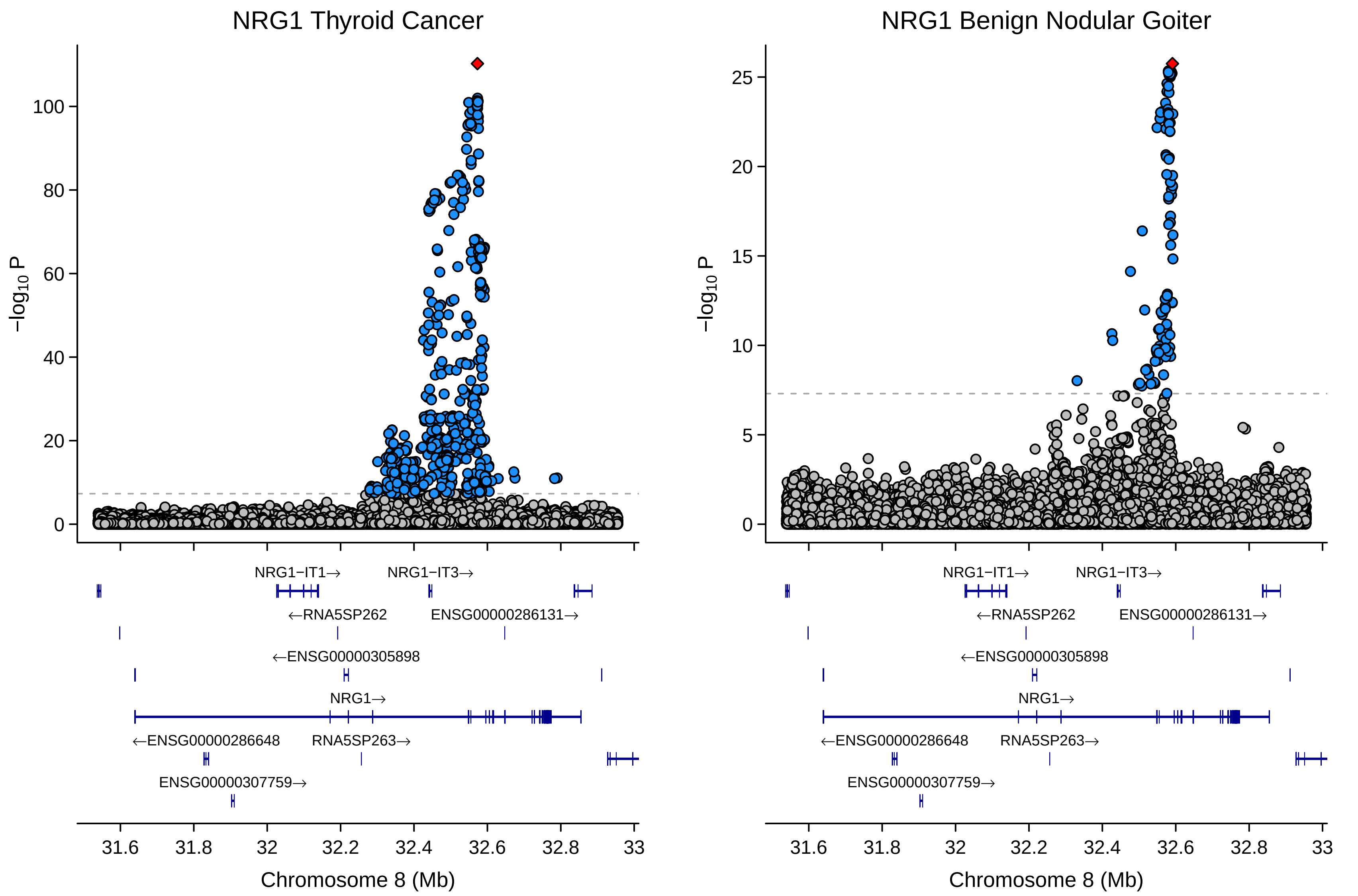

NRG1 Thyroid Cancer
NRG1 Benign Nodular Goiter
−log10 P
0
20
40
60
80
100
0
5
10
15
20
25
NRG1−IT1→
NRG1−IT3→
←RNA5SP262
ENSG00000286131→
←ENSG00000305898
NRG1→
←ENSG00000286648
RNA5SP263→
ENSG00000307759→
31.6
31.8
32
32.2
32.4
32.6
32.8
33
Chromosome 8 (Mb)

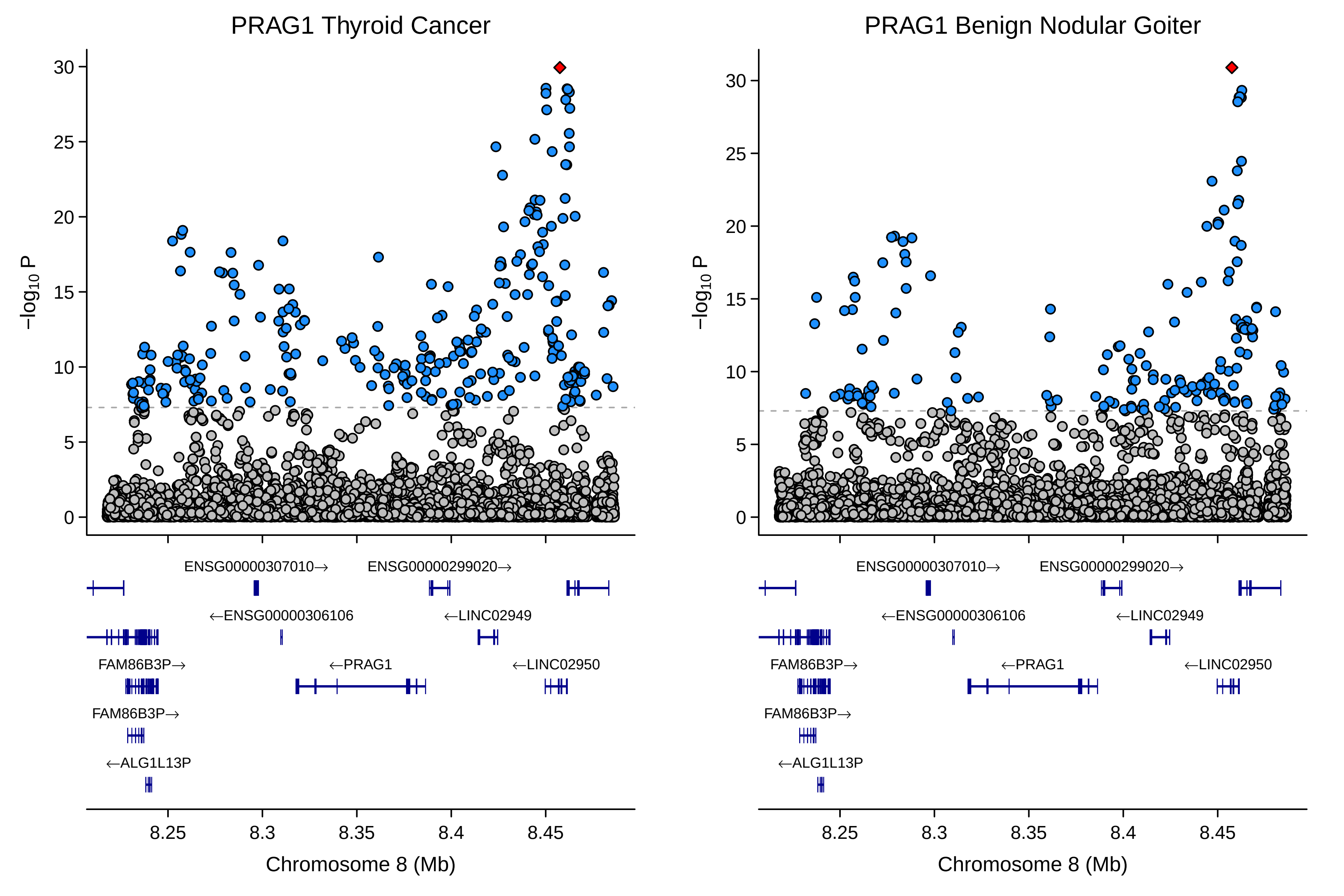
PRAG1 Thyroid Cancer
PRAG1 Benign Nodular Goiter
−log10 P
30
25
20
15
10
5
0
ENSG00000307010→
ENSG00000299020→
←ENSG00000306106
←LINC02949
FAM86B3P→
←PRAG1
←LINC02950
FAM86B3P→
←ALG1L13P
8.25
8.3
8.35
8.4
8.45
Chromosome 8 (Mb)

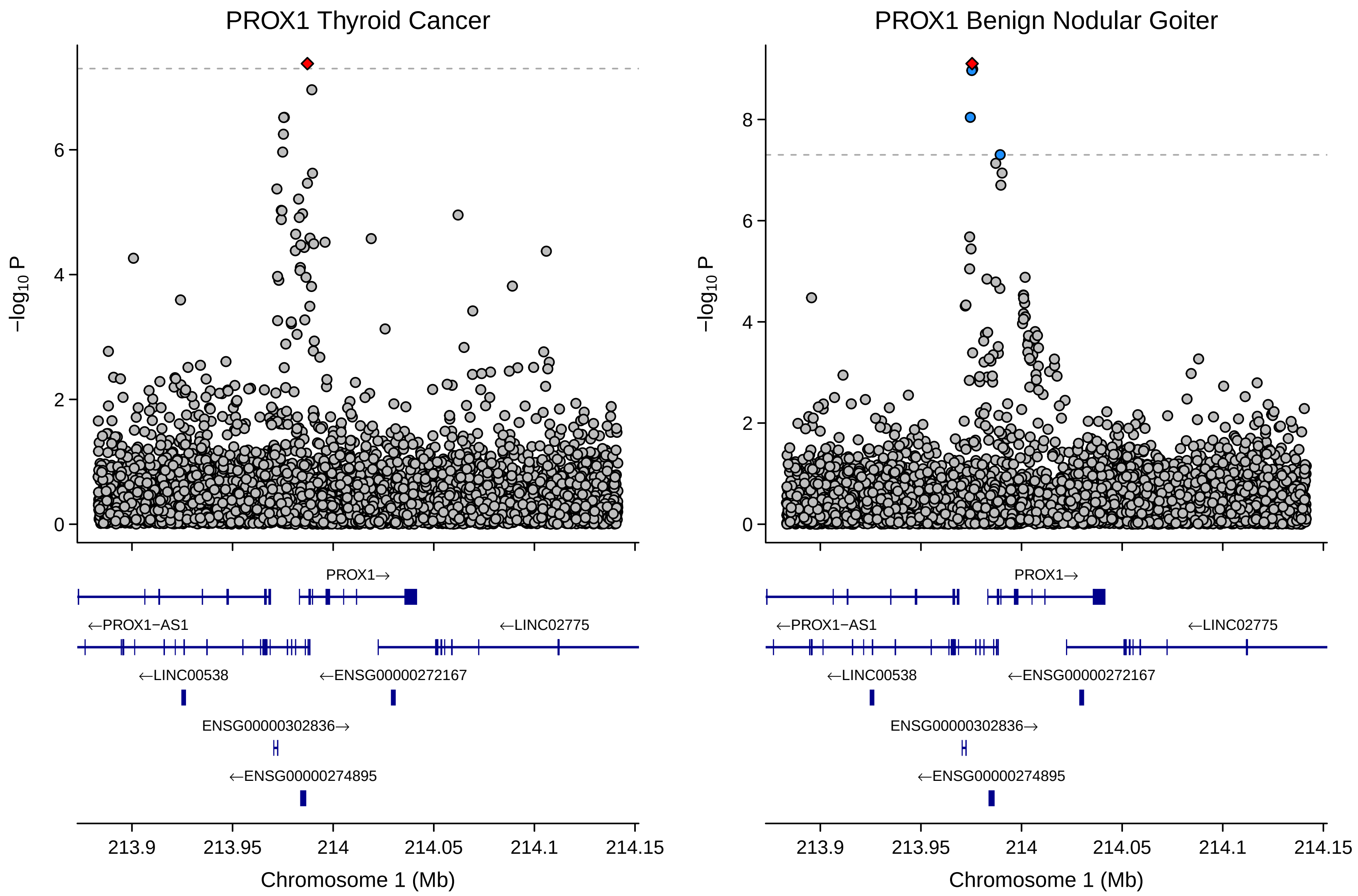
PROX1 Thyroid Cancer
PROX1 Benign Nodular Goiter
−log10 P
0
2
4
6
8
PROX1→
←PROX1−AS1
←LINC02775
←LINC00538
←ENSG00000272167
ENSG00000302836→
←ENSG00000274895
213.9
213.95
214
214.05
214.1
214.15
Chromosome 1 (Mb)

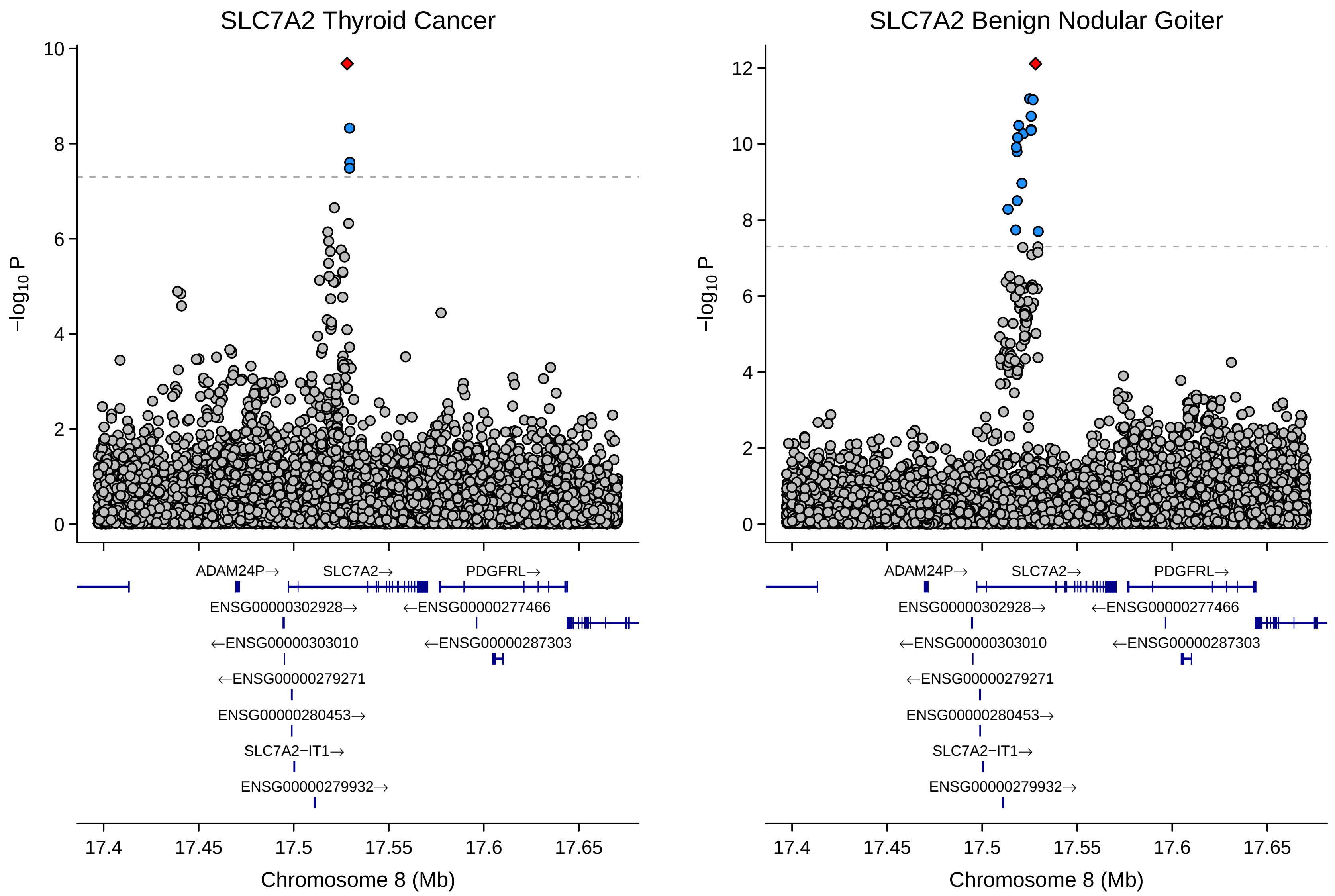

SLC7A2 Thyroid Cancer
SLC7A2 Benign Nodular Goiter
−log10 P
0
2
4
6
8
10
12
ADAM24P→
SLC7A2→
PDGFRL→
ENSG00000302928→
←ENSG00000277466
←ENSG00000303010
←ENSG00000287303
←ENSG00000279271
ENSG00000280453→
SLC7A2−IT1→
ENSG00000279932→
17.4
17.45
17.5
17.55
17.6
17.65
Chromosome 8 (Mb)

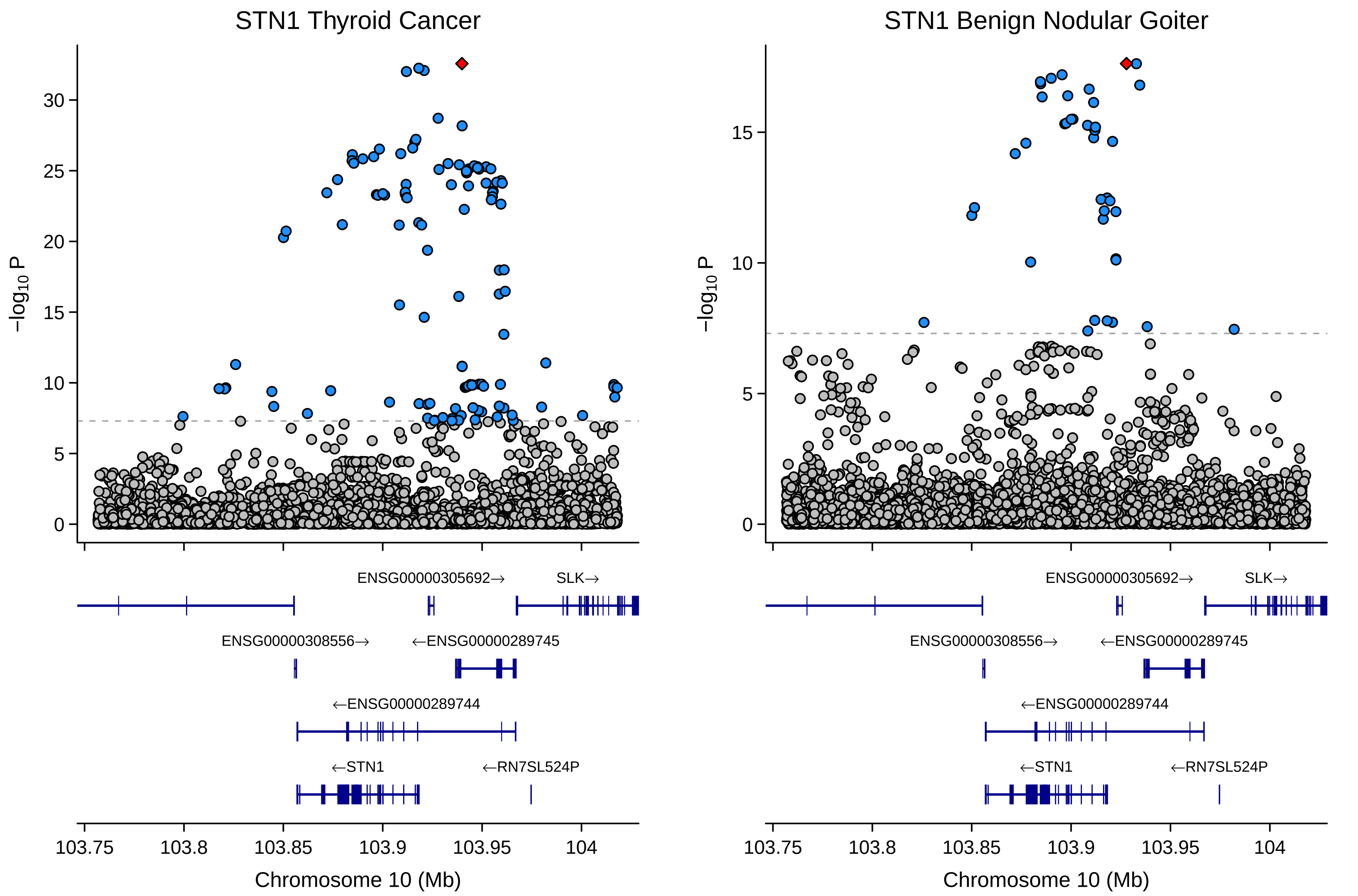

STN1 Thyroid Cancer
STN1 Benign Nodular Goiter
−log10 P
30
25
20
15
10
5
0
ENSG00000305692→
SLK→
ENSG00000308556→
←ENSG00000289745
←ENSG00000289744
←STN1
←RN7SL524P
103.75
103.8
103.85
103.9
103.95
104
Chromosome 10 (Mb)

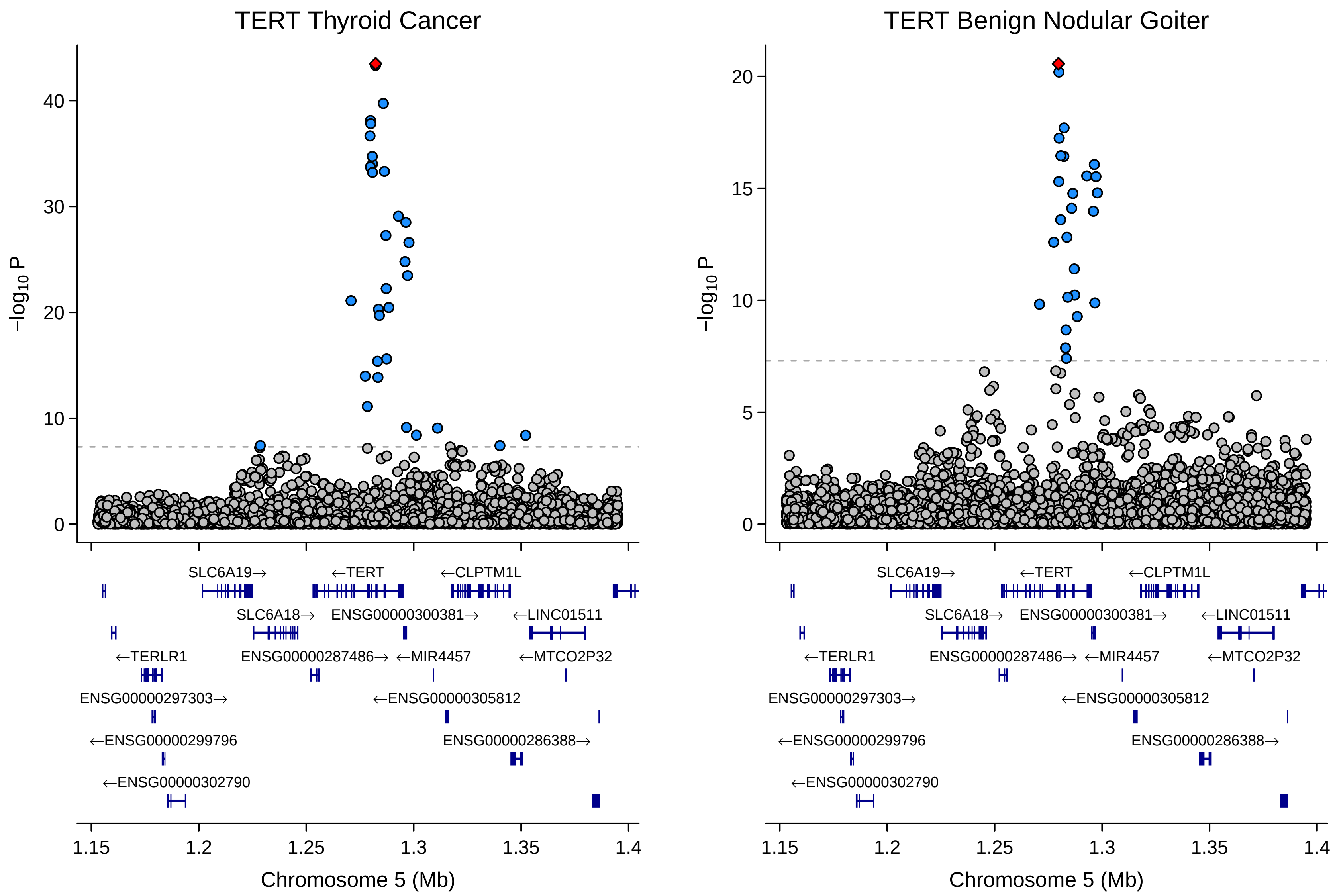

TERT Thyroid Cancer
TERT Benign Nodular Goiter
−log10 P
40
30
20
10
0
20
15
10
5
0
SLC6A19→
←TERT
←CLPTM1L
SLC6A18→
ENSG00000300381→
←LINC01511
←TERLR1
ENSG00000287486→
←MIR4457
←MTCO2P32
ENSG00000297303→
←ENSG00000305812
←ENSG00000299796
ENSG00000286388→
←ENSG00000302790
1.15
1.2
1.25
1.3
1.35
1.4
Chromosome 5 (Mb)

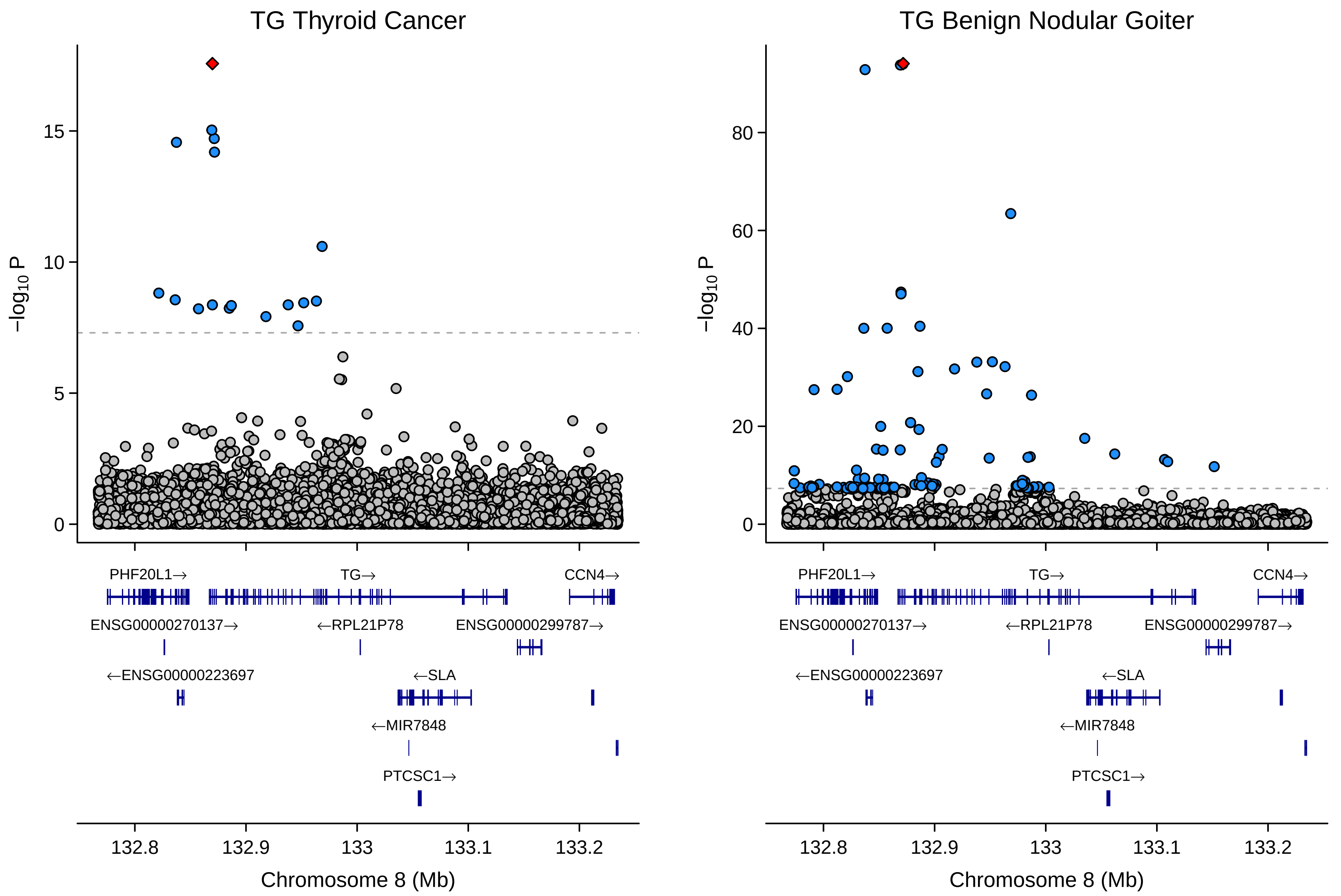

TG Thyroid Cancer
TG Benign Nodular Goiter
−log10 P
0
5
10
15
20
40
60
80
PHF20L1→
TG→
CCN4→
ENSG00000270137→
←RPL21P78
ENSG00000299787→
←ENSG00000223697
←SLA
←MIR7848
PTCSC1→
132.8
132.9
133
133.1
133.2
Chromosome 8 (Mb)

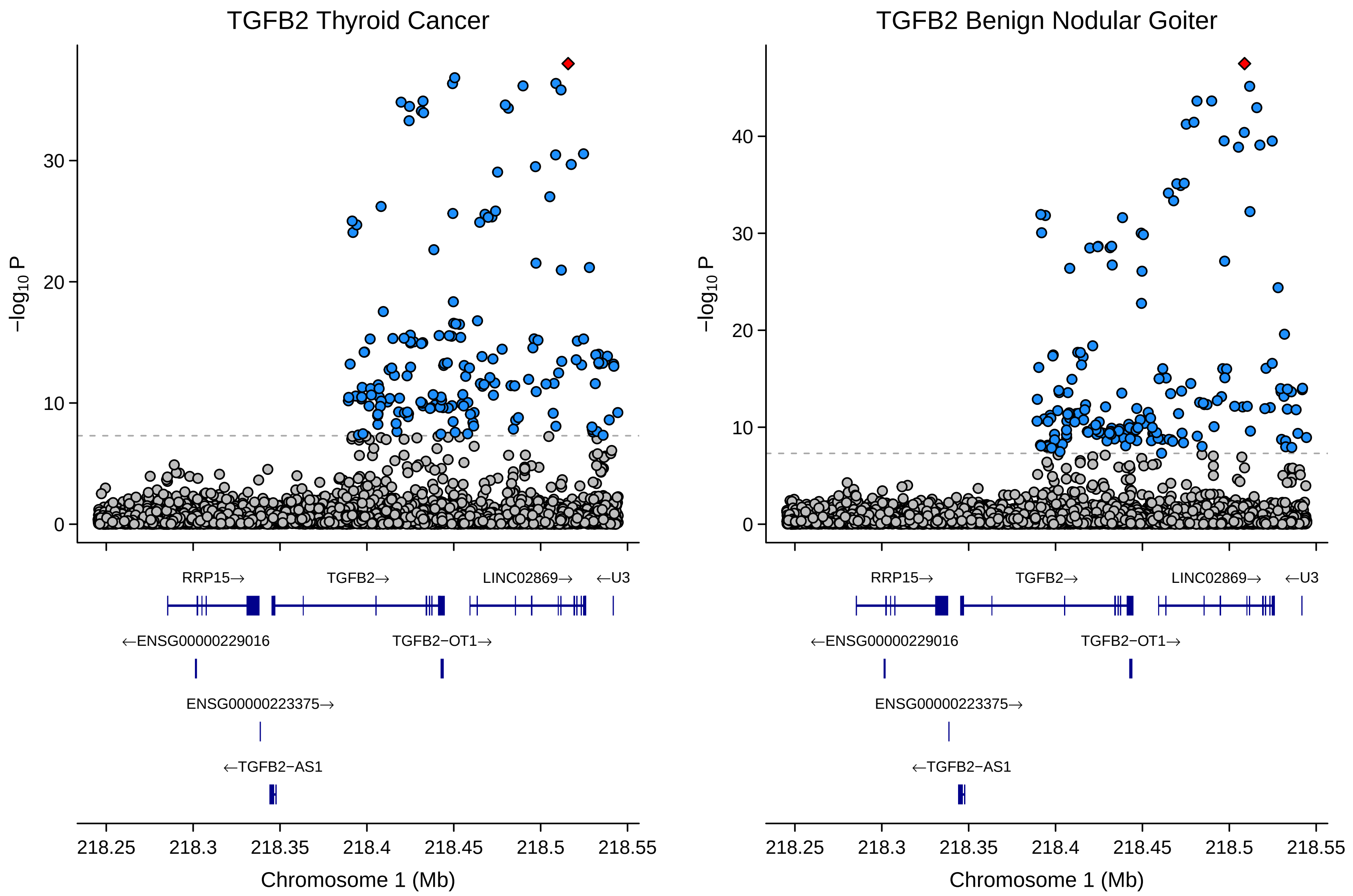
TGFB2 Thyroid Cancer
TGFB2 Benign Nodular Goiter
−log10 P
0
10
20
30
40
RRP15→
TGFB2→
LINC02869→
←U3
←ENSG00000229016
TGFB2−OT1→
ENSG00000223375→
←TGFB2−AS1
218.25
218.3
218.35
218.4
218.45
218.5
218.55
Chromosome 1 (Mb)

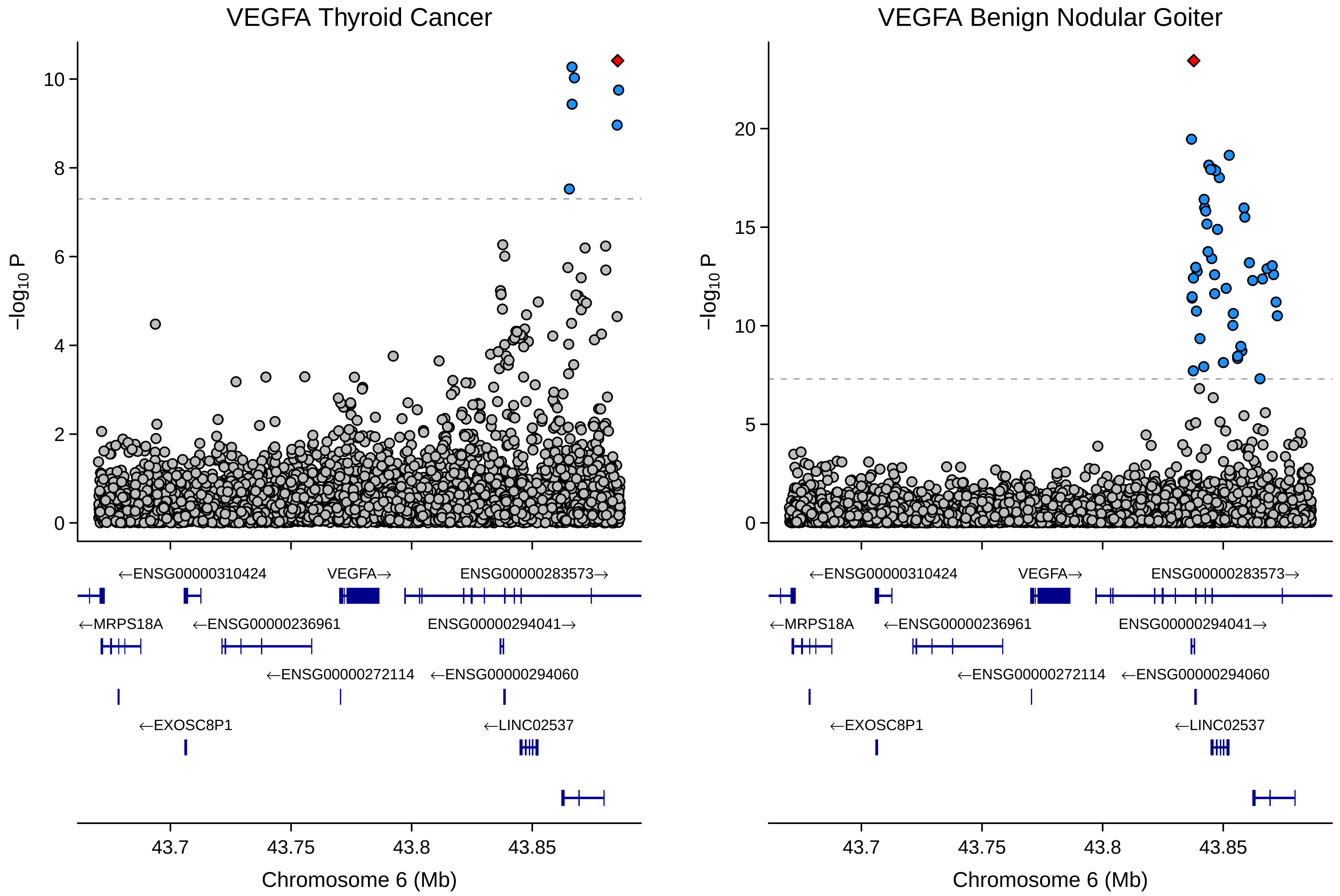

VEGFA Thyroid Cancer
VEGFA Benign Nodular Goiter
−log10 P
Chromosome 6 (Mb)
43.7
43.75
43.8
43.85
←ENSG00000310424
VEGFA→
ENSG00000283573→
←MRPS18A
←ENSG00000236961
ENSG00000294041→
←ENSG00000272114
←ENSG00000294060
←EXOSC8P1
←LINC02537

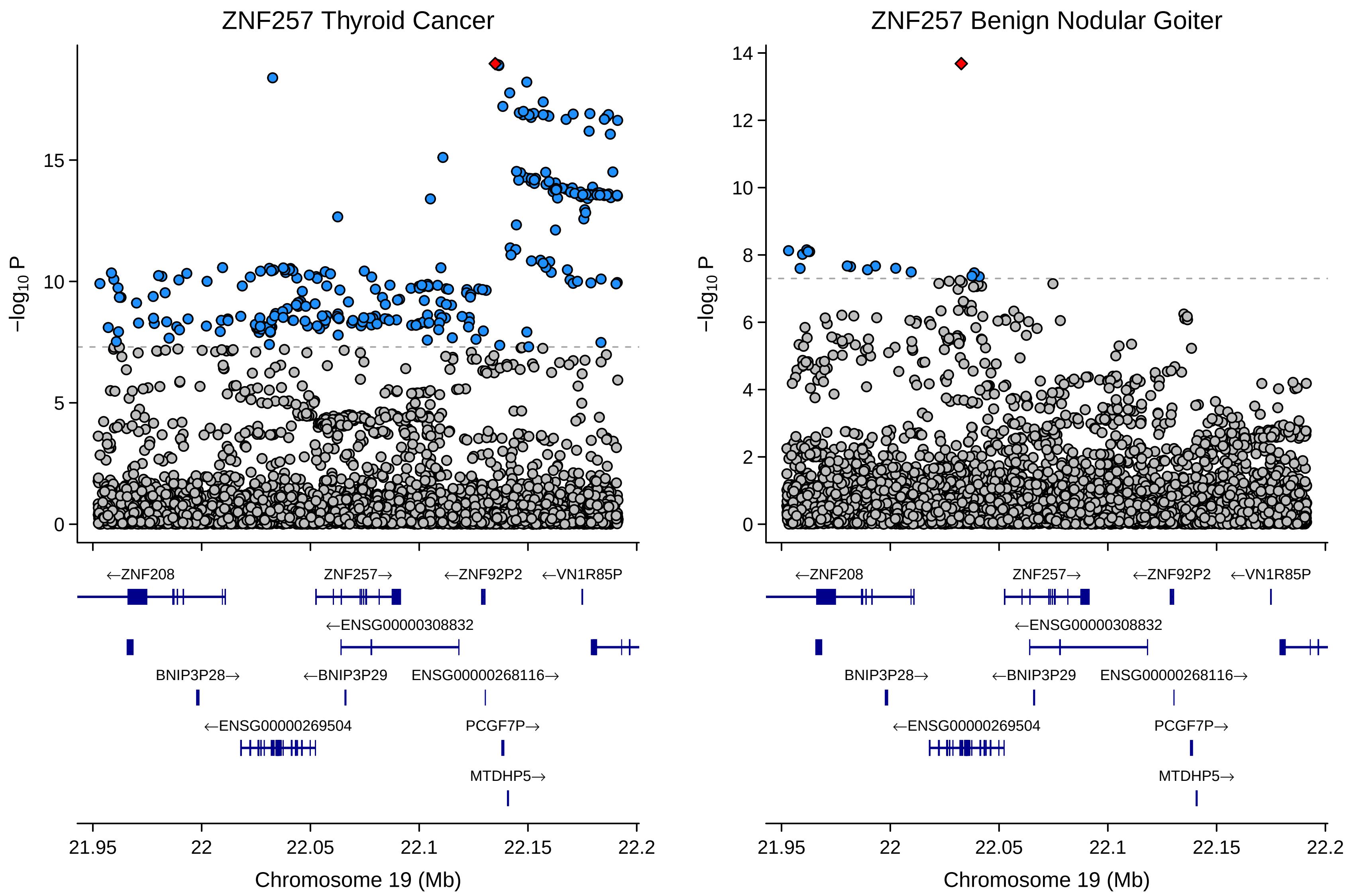
ZNF257 Thyroid Cancer
ZNF257 Benign Nodular Goiter
$-\log_{10}$ P
Chromosome 19 (Mb)
21.95
22
22.05
22.1
22.15
22.2
←ZNF208
ZNF257→
←ZNF92P2
←VN1R85P
←ENSG00000308832
BNIP3P28→
←BNIP3P29
ENSG00000268116→
←ENSG00000269504
PCGF7P→
MTDHP5→
