## Supplementary Figure 2.1 for "Global multi-ancestry genetic study elucidates genes and biological pathways associated with thyroid cancer and benign thyroid diseases"

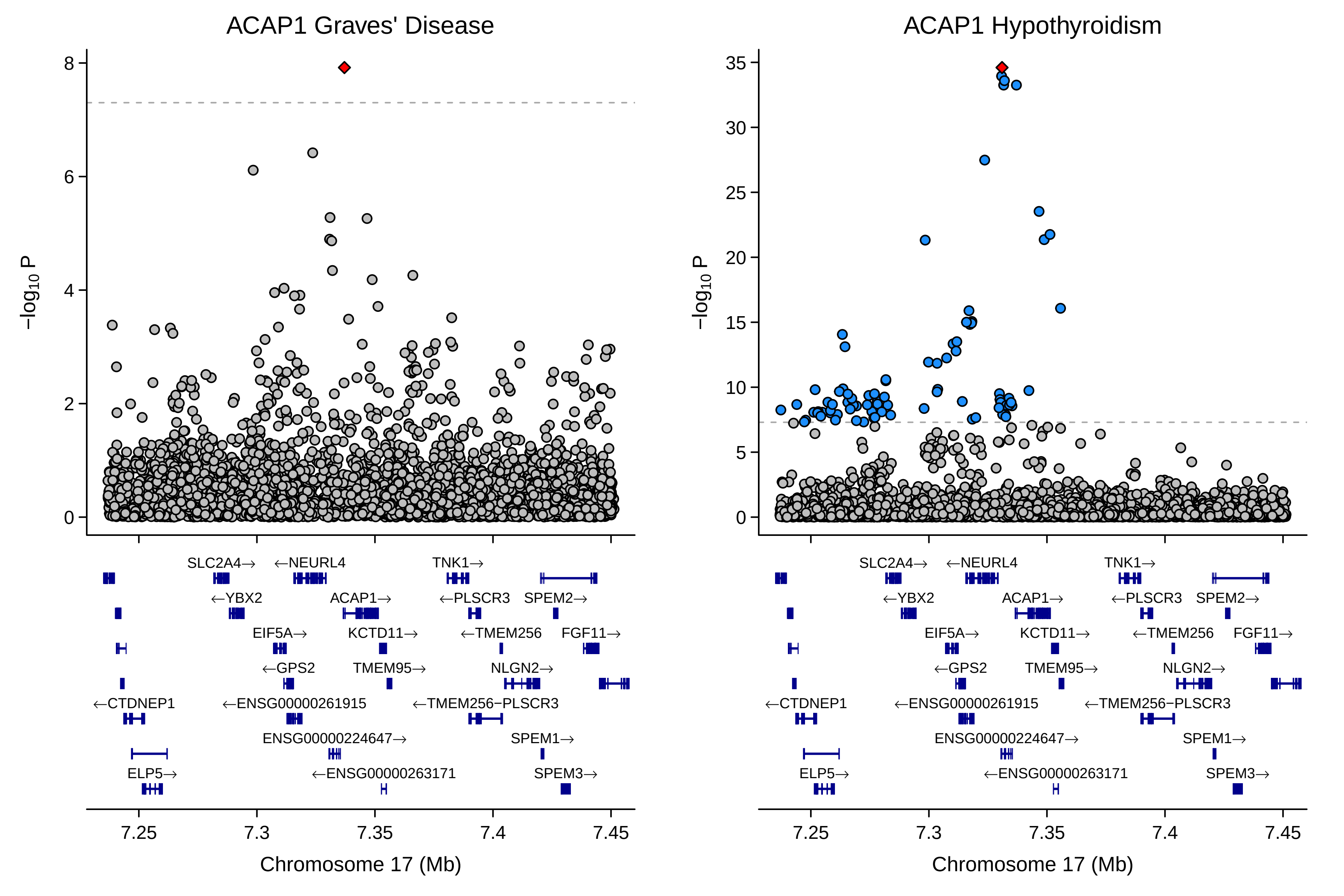
ACAP1 Graves' Disease
ACAP1 Hypothyroidism
−log10 P
Chromosome 17 (Mb)
7.25
7.3
7.35
7.4
7.45
SLC2A4→
←NEURL4
TNK1→
←YBX2
ACAP1→
←PLSCR3
SPEM2→
EIF5A→
KCTD11→
←TMEM256
FGF11→
←GPS2
TMEM95→
NLGN2→
←CTDNEP1
←ENSG00000261915
←TMEM256−PLSCR3
ENSG00000224647→
SPEM1→
ELP5→
←ENSG00000263171
SPEM3→

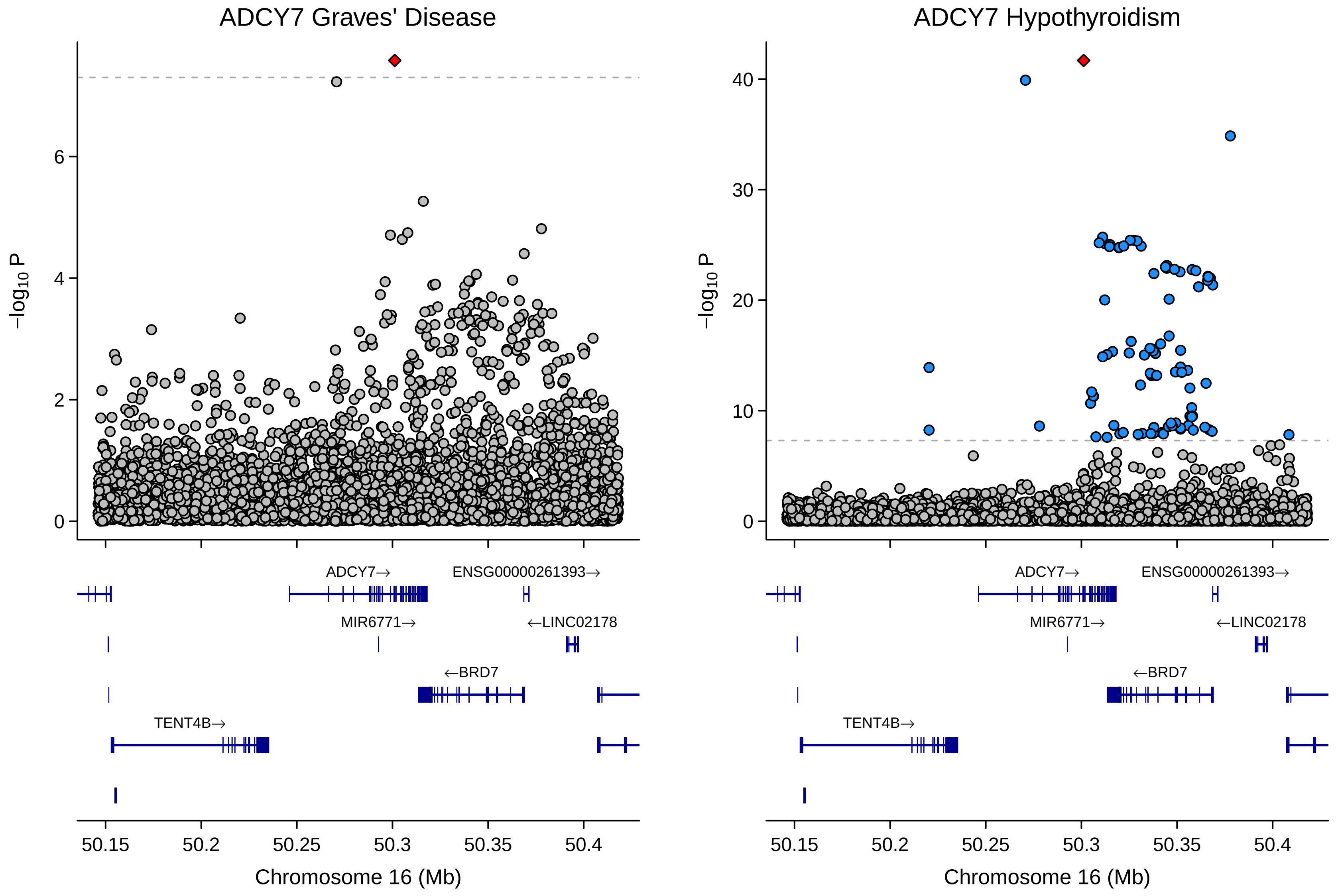
ADCY7 Graves' Disease
ADCY7 Hypothyroidism
$-\log_{10}$ P
Chromosome 16 (Mb)
50.15
50.2
50.25
50.3
50.35
50.4
ADCY7→
ENSG00000261393→
MIR6771→
←LINC02178
←BRD7
TENT4B→

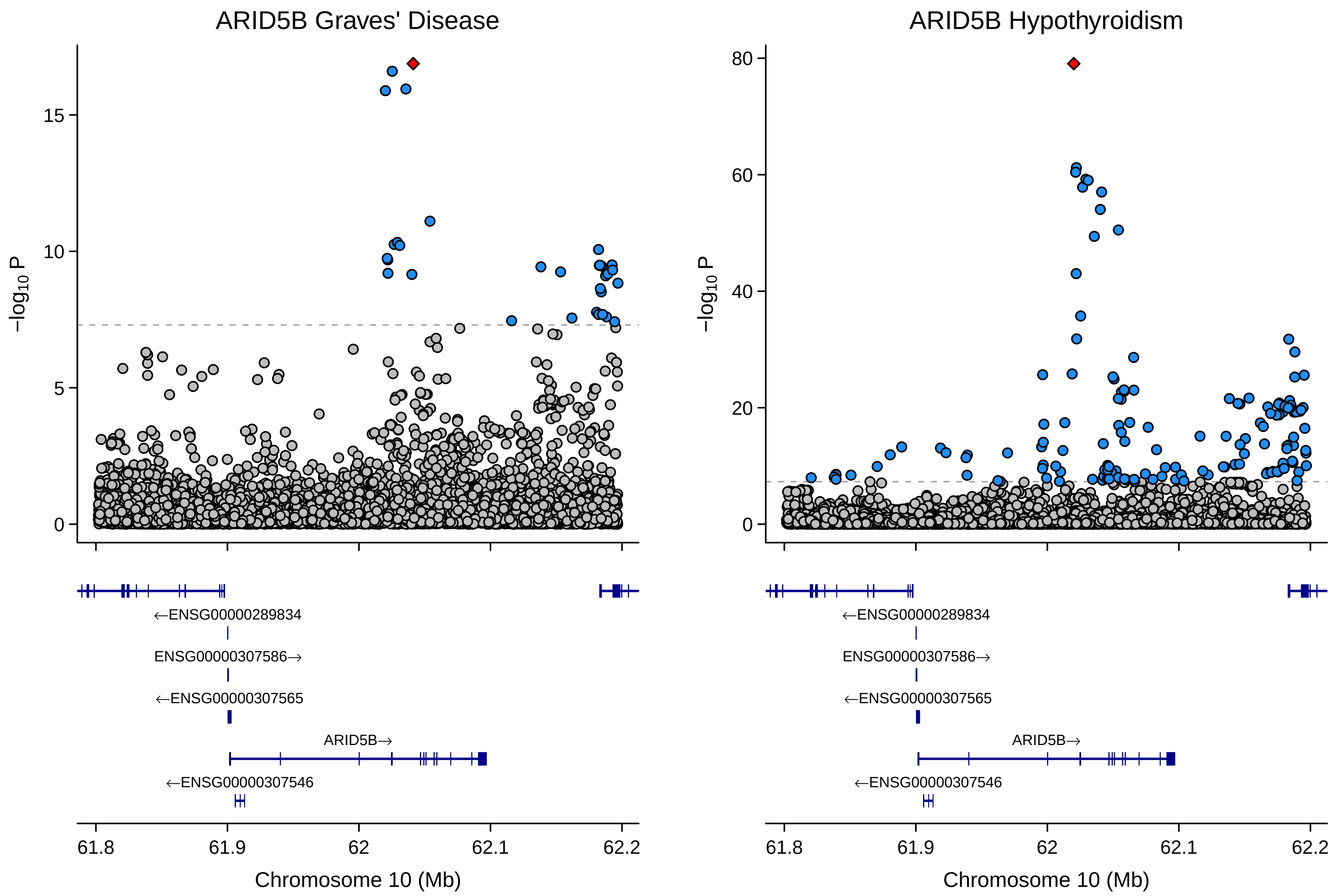

ARID5B Graves' Disease
ARID5B Hypothyroidism
−log10 P
0
5
10
15
20
40
60
80
←ENSG00000289834
ENSG00000307586→
←ENSG00000307565
ARID5B→
←ENSG00000307546
61.8
61.9
62
62.1
62.2
Chromosome 10 (Mb)

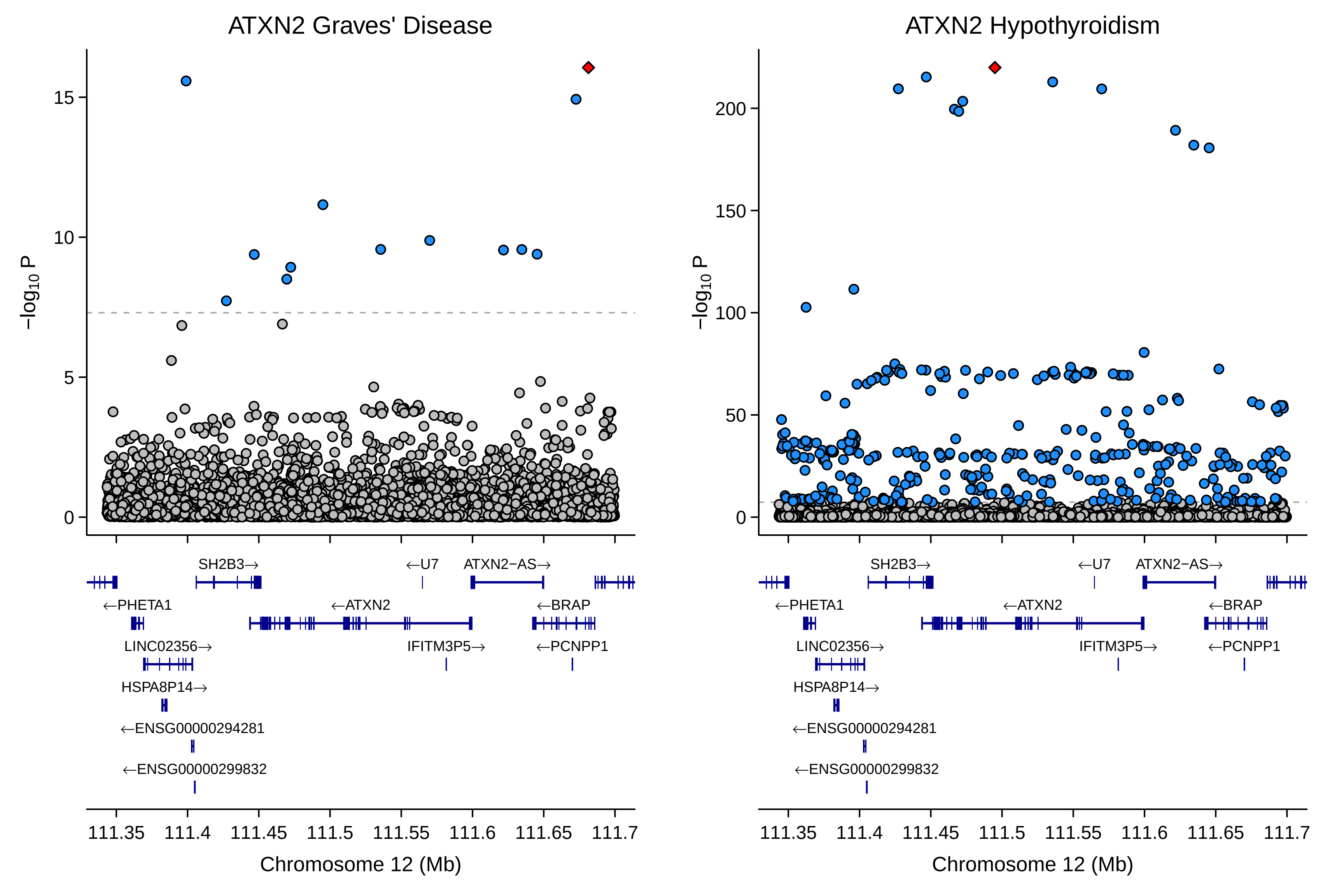
ATXN2 Graves' Disease
ATXN2 Hypothyroidism
−log10 P
0
5
10
15
0
50
100
150
200
SH2B3→
←U7
ATXN2−AS→
←PHETA1
←ATXN2
←BRAP
LINC02356→
IFITM3P5→
←PCNPP1
HSPA8P14→
←ENSG00000294281
←ENSG00000299832
111.35
111.4
111.45
111.5
111.55
111.6
111.65
111.7
Chromosome 12 (Mb)

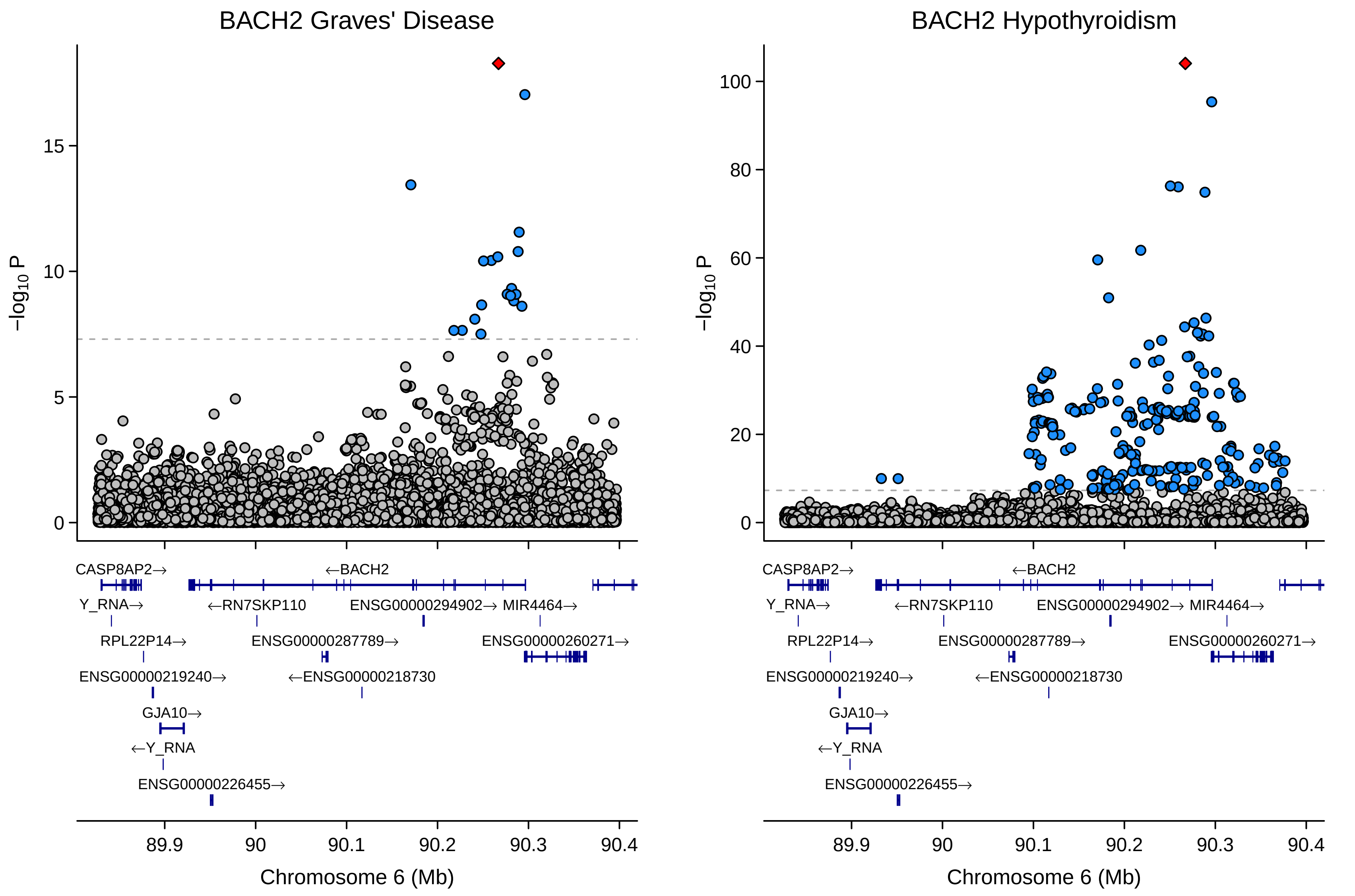

BACH2 Graves' Disease
BACH2 Hypothyroidism
−log10 P
0
5
10
15
20
40
60
80
100
CASP8AP2→
←BACH2
Y_RNA→
←RN7SKP110
ENSG00000294902→
MIR4464→
RPL22P14→
ENSG00000287789→
ENSG00000260271→
ENSG00000219240→
←ENSG00000218730
GJA10→
←Y_RNA
ENSG00000226455→
89.9
90
90.1
90.2
90.3
90.4
Chromosome 6 (Mb)

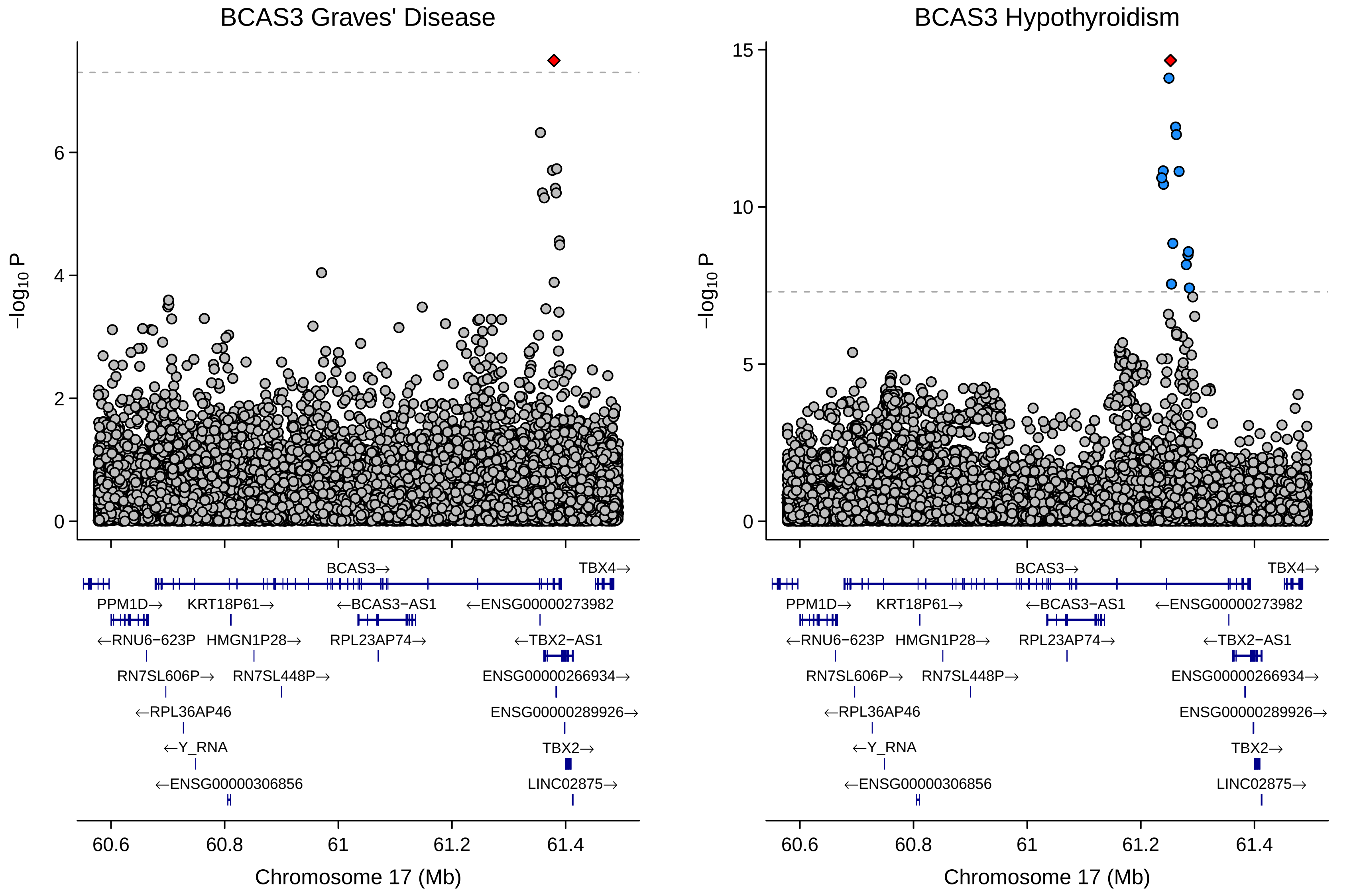

BCAS3 Graves' Disease
BCAS3 Hypothyroidism
−log10 P
0
2
4
6
5
10
15
BCAS3→
TBX4→
PPM1D→
KRT18P61→
←BCAS3−AS1
←ENSG00000273982
←RNU6−623P
HMGN1P28→
RPL23AP74→
←TBX2−AS1
RN7SL606P→
RN7SL448P→
ENSG00000266934→
←RPL36AP46
ENSG00000289926→
←Y_RNA
TBX2→
←ENSG00000306856
LINC02875→
60.6
60.8
61
61.2
61.4
Chromosome 17 (Mb)

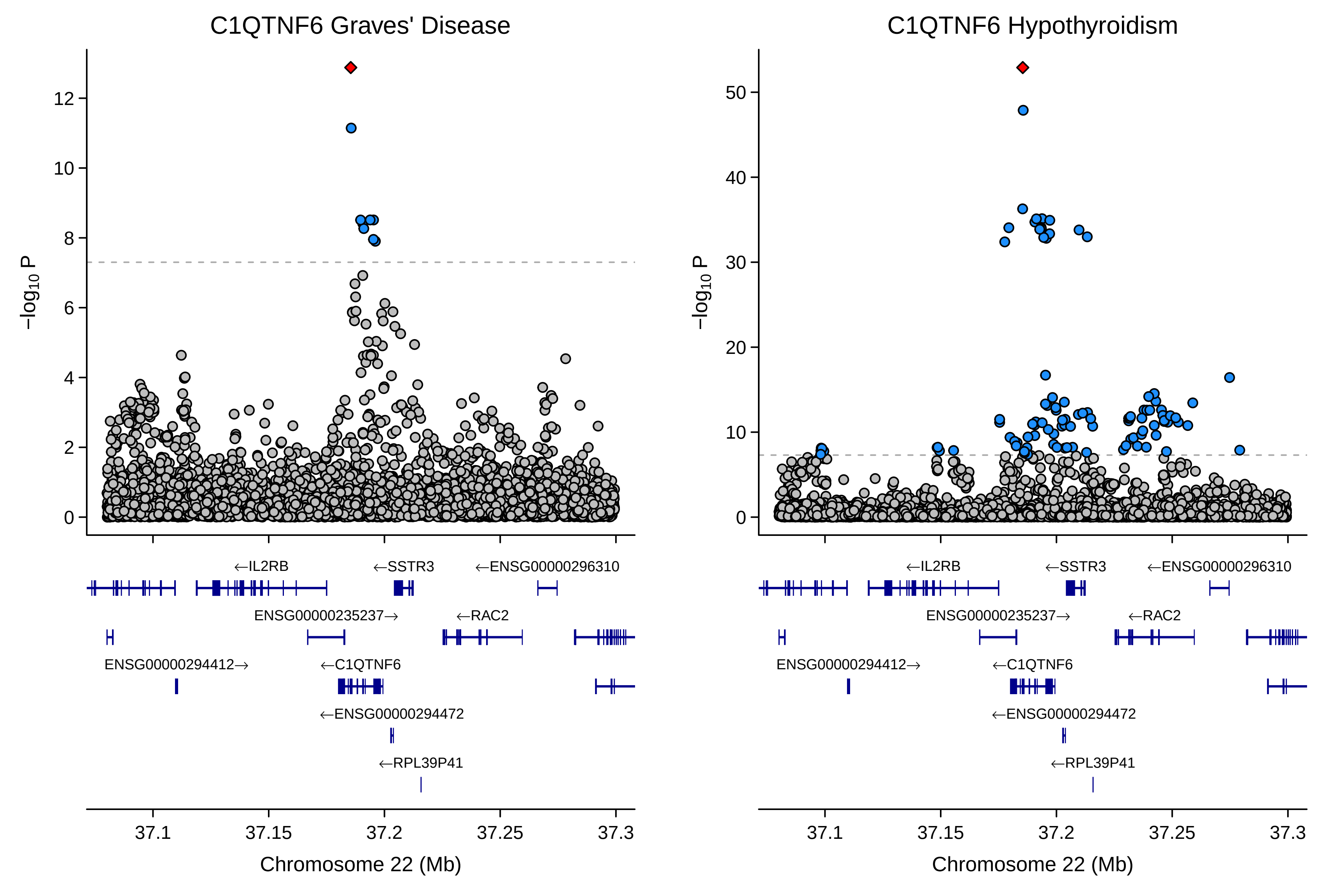

C1QTNF6 Graves' Disease
C1QTNF6 Hypothyroidism
−log10 P
0
2
4
6
8
10
12
20
30
40
50
←IL2RB
←SSTR3
←ENSG00000296310
ENSG00000235237→
←RAC2
ENSG00000294412→
←C1QTNF6
←ENSG00000294472
←RPL39P41
37.1
37.15
37.2
37.25
37.3
Chromosome 22 (Mb)

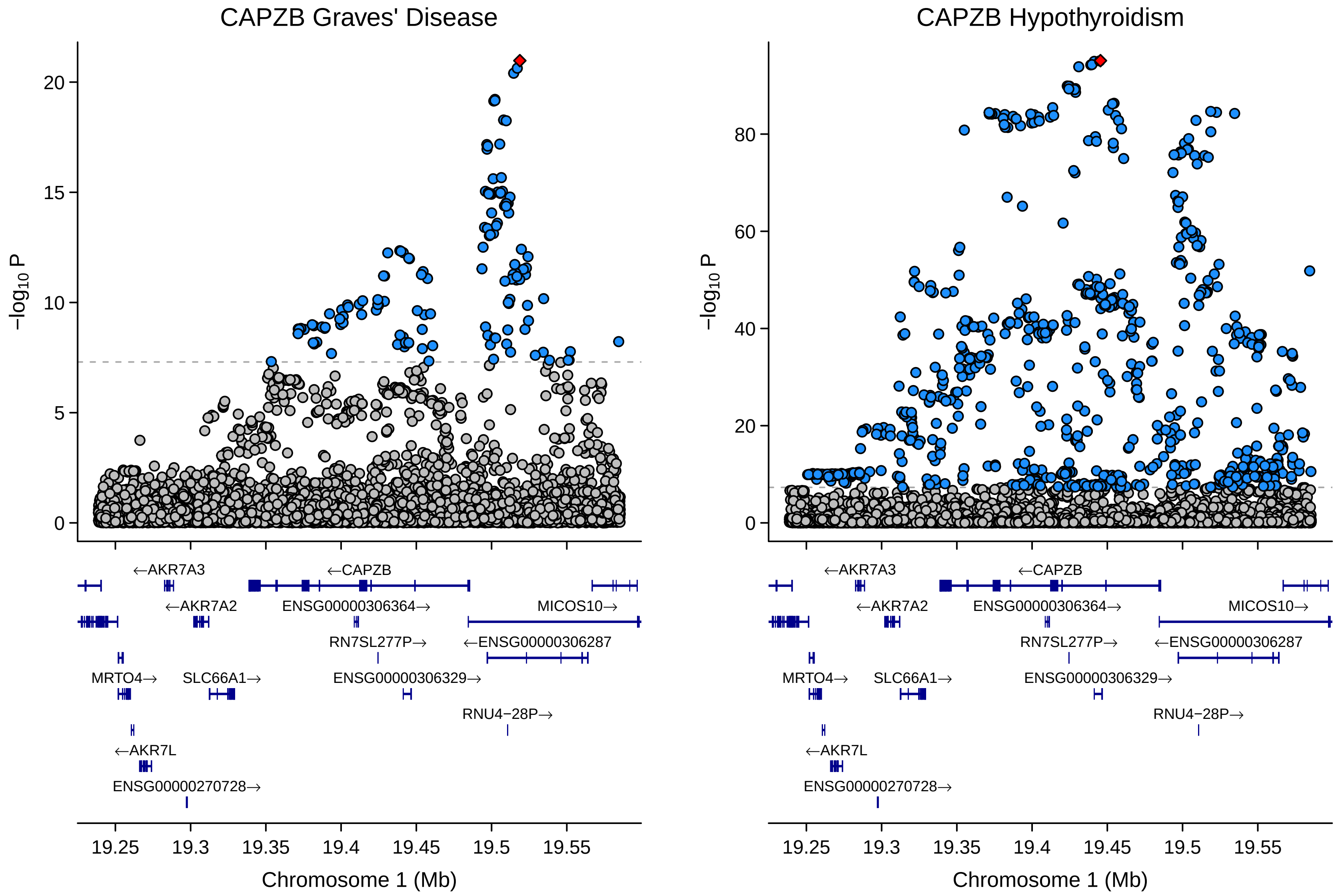

CAPZB Graves' Disease
CAPZB Hypothyroidism
−log10 P
0
5
10
15
20
40
60
80
←AKR7A3
←CAPZB
←AKR7A2
ENSG00000306364→
MICOS10→
RN7SL277P→
←ENSG00000306287
MRTO4→
SLC66A1→
ENSG00000306329→
RNU4−28P→
←AKR7L
ENSG00000270728→
19.25
19.3
19.35
19.4
19.45
19.5
19.55
Chromosome 1 (Mb)

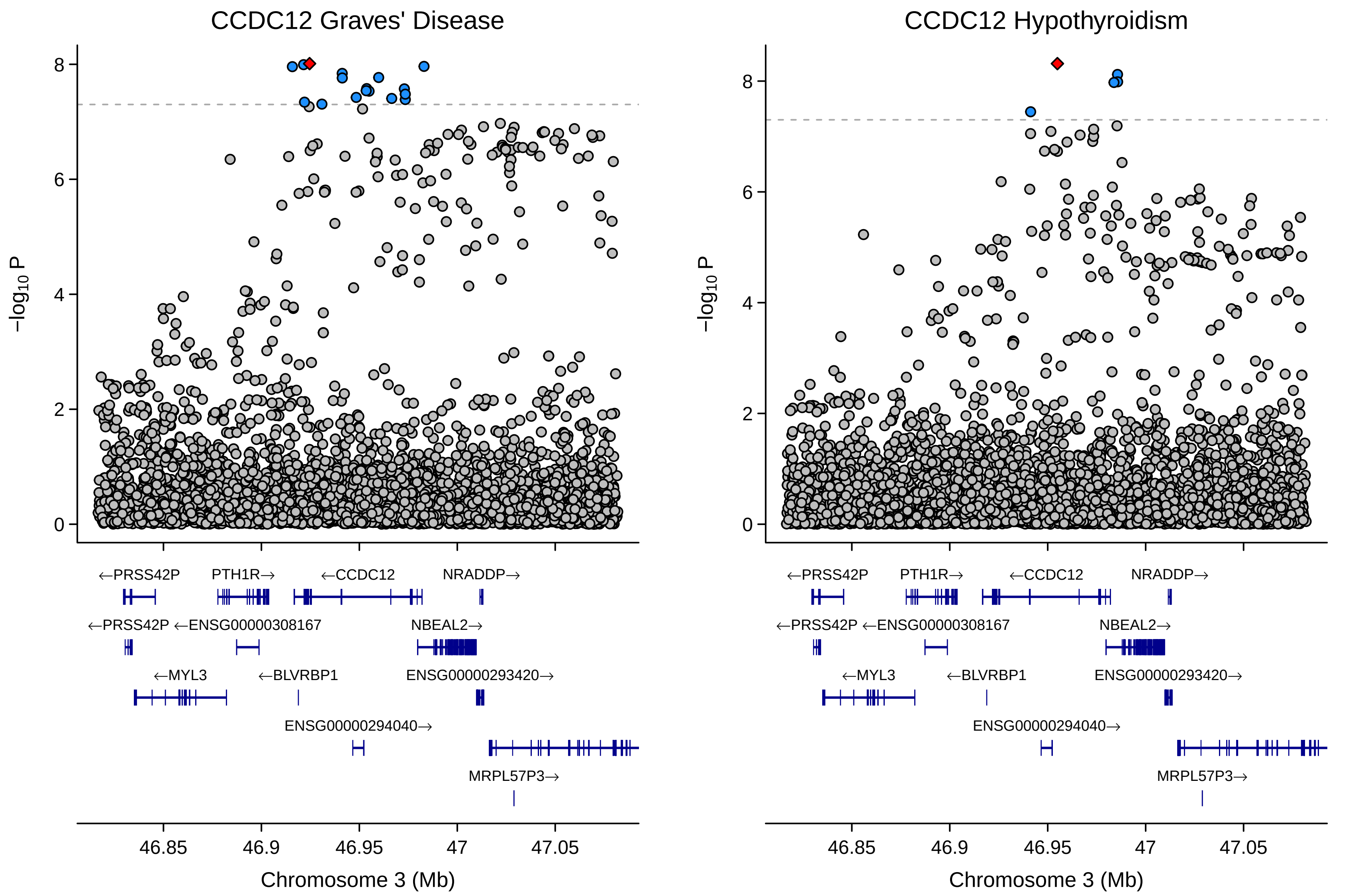

CCDC12 Graves' Disease
CCDC12 Hypothyroidism
−log10 P
8
6
4
2
0
←PRSS42P
PTH1R→
←CCDC12
NRADDP→
←PRSS42P ←ENSG00000308167
NBEAL2→
←MYL3
←BLVRBP1
ENSG00000293420→
ENSG00000294040→
MRPL57P3→
46.85
46.9
46.95
47
47.05
Chromosome 3 (Mb)

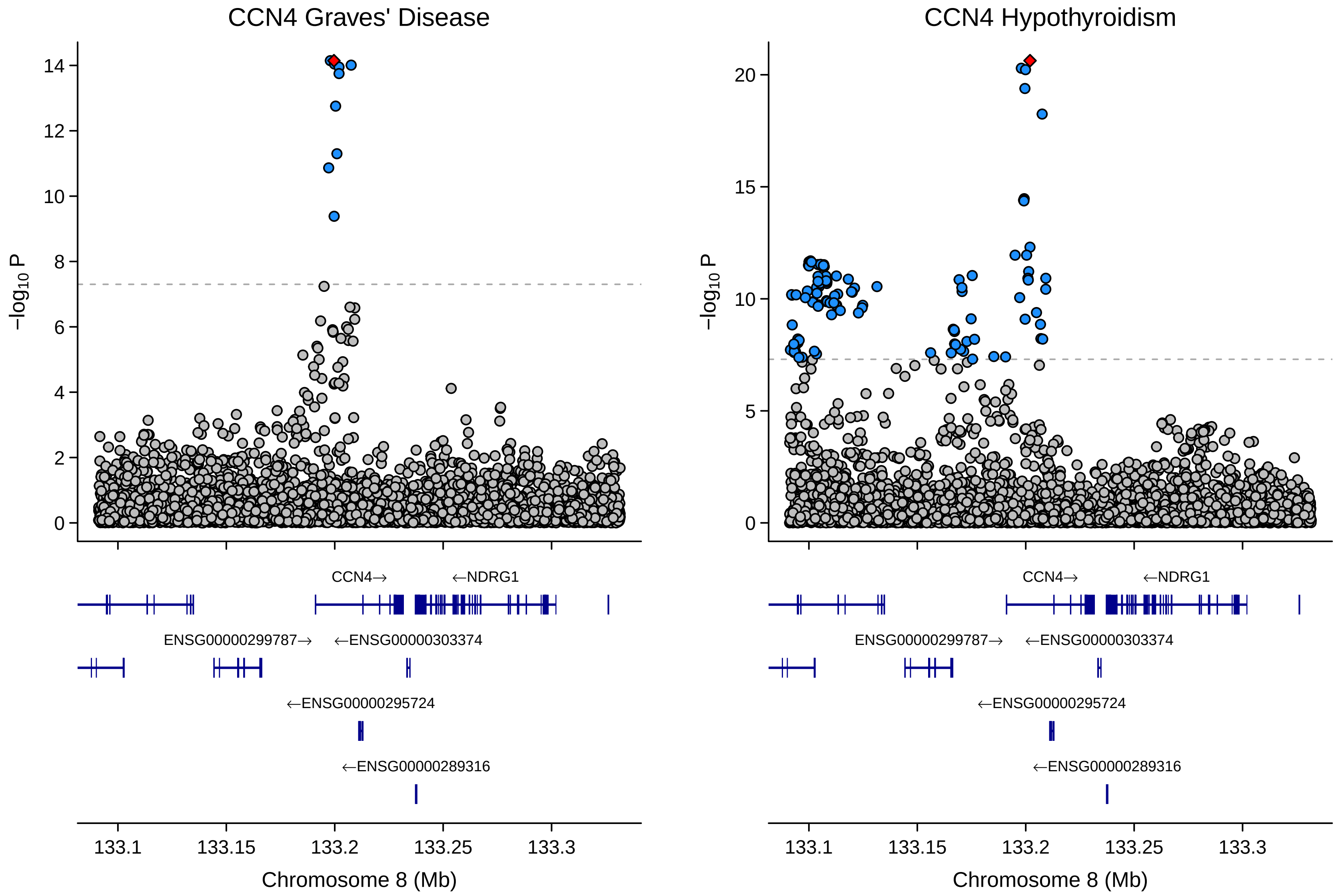

CCN4 Graves' Disease
CCN4 Hypothyroidism
−log10 P
0
2
4
6
8
10
12
14
5
15
20
CCN4→
←NDRG1
ENSG00000299787→
←ENSG00000303374
←ENSG00000295724
←ENSG00000289316
133.1
133.15
133.2
133.25
133.3
Chromosome 8 (Mb)

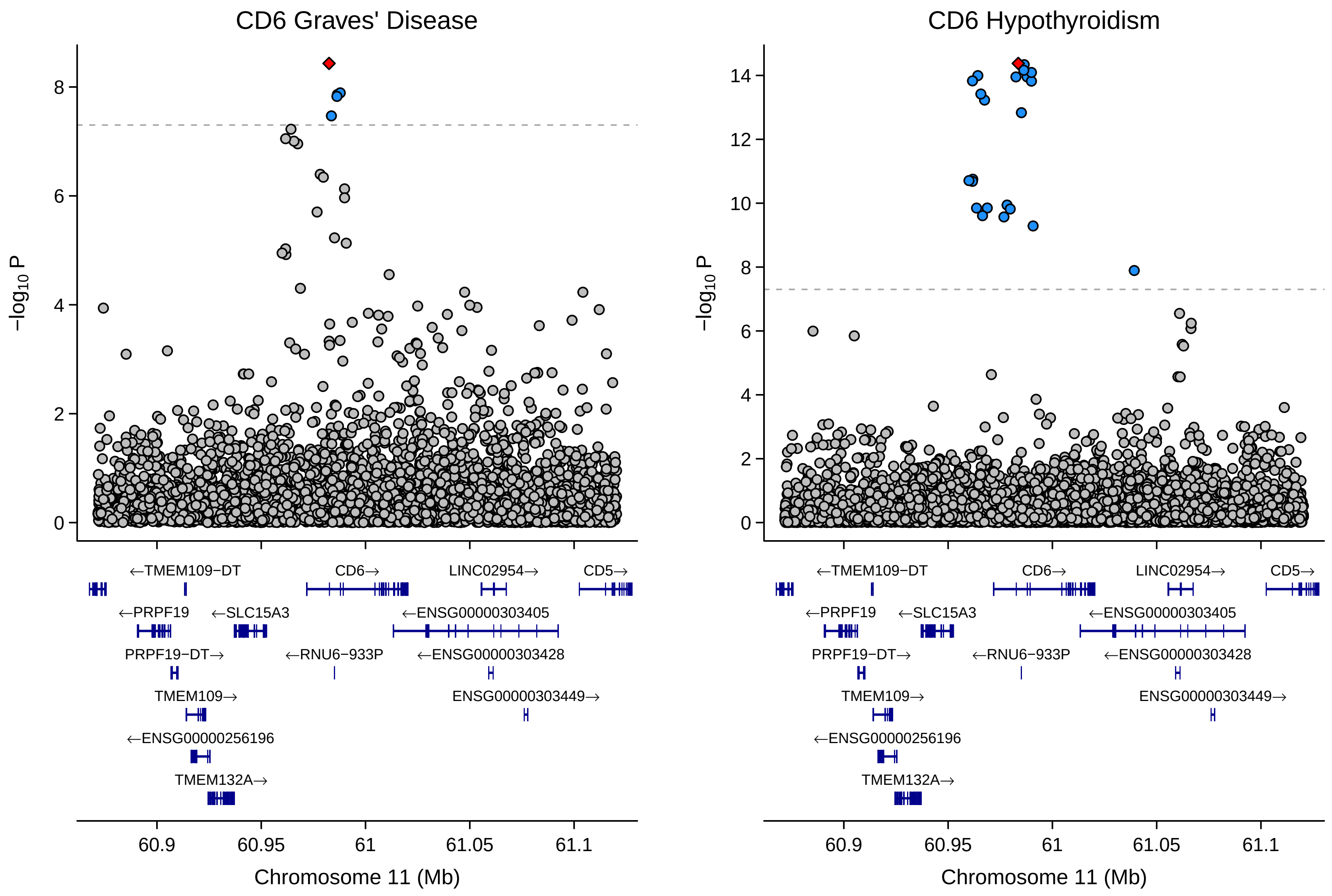
CD6 Graves' Disease
CD6 Hypothyroidism
−log10 P
0
2
4
6
8
10
12
14
←TMEM109−DT
CD6→
LINC02954→
CD5→
←PRPF19
←SLC15A3
←ENSG00000303405
PRPF19−DT→
←RNU6−933P
←ENSG00000303428
TMEM109→
ENSG00000303449→
←ENSG00000256196
TMEM132A→
60.9
60.95
61
61.05
61.1
Chromosome 11 (Mb)

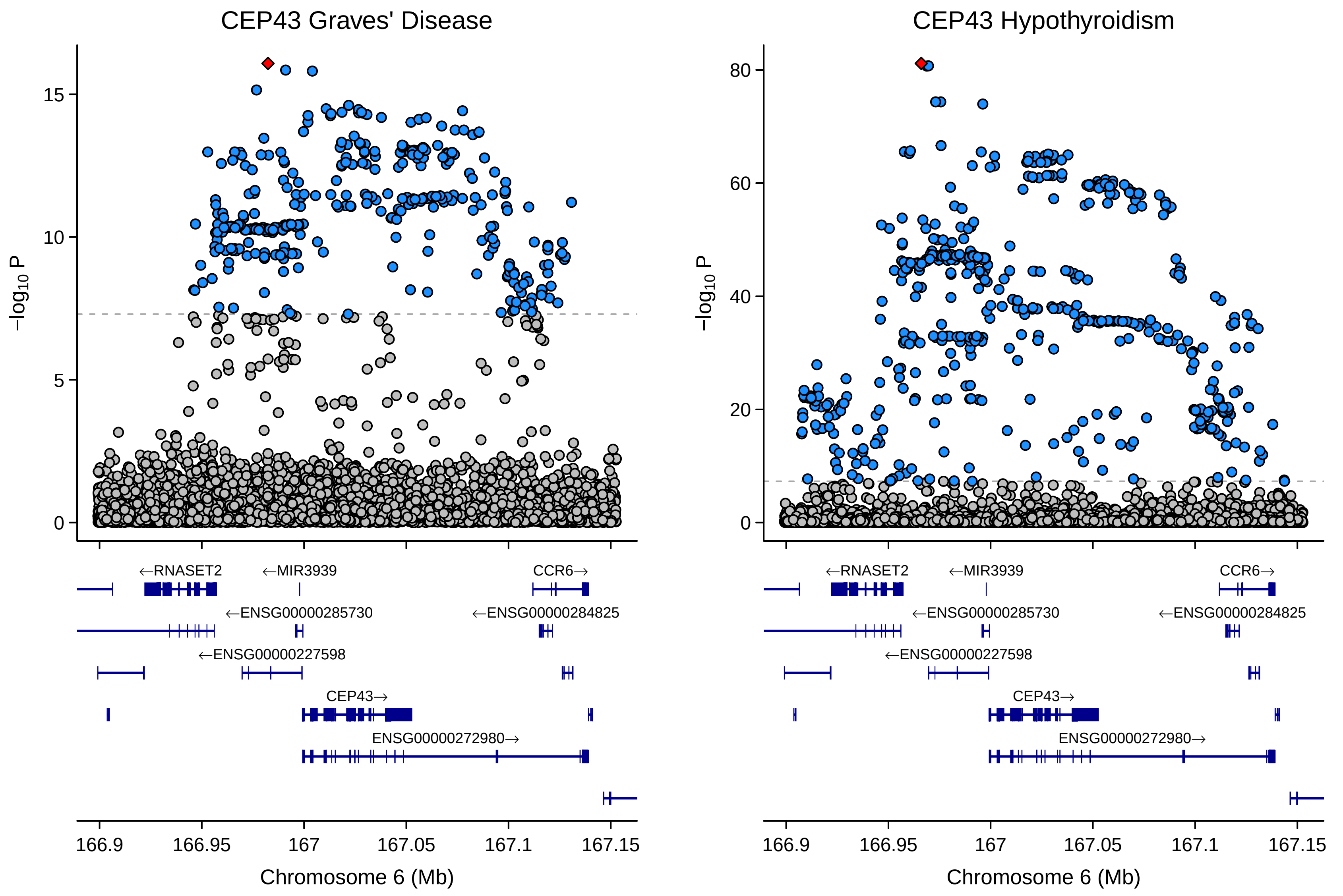
CEP43 Graves' Disease
CEP43 Hypothyroidism
−log10 P
←RNASET2
←MIR3939
CCR6→
←ENSG00000285730
←ENSG00000284825
←ENSG00000227598
CEP43→
ENSG00000272980→
166.9
166.95
167
167.05
167.1
167.15
Chromosome 6 (Mb)

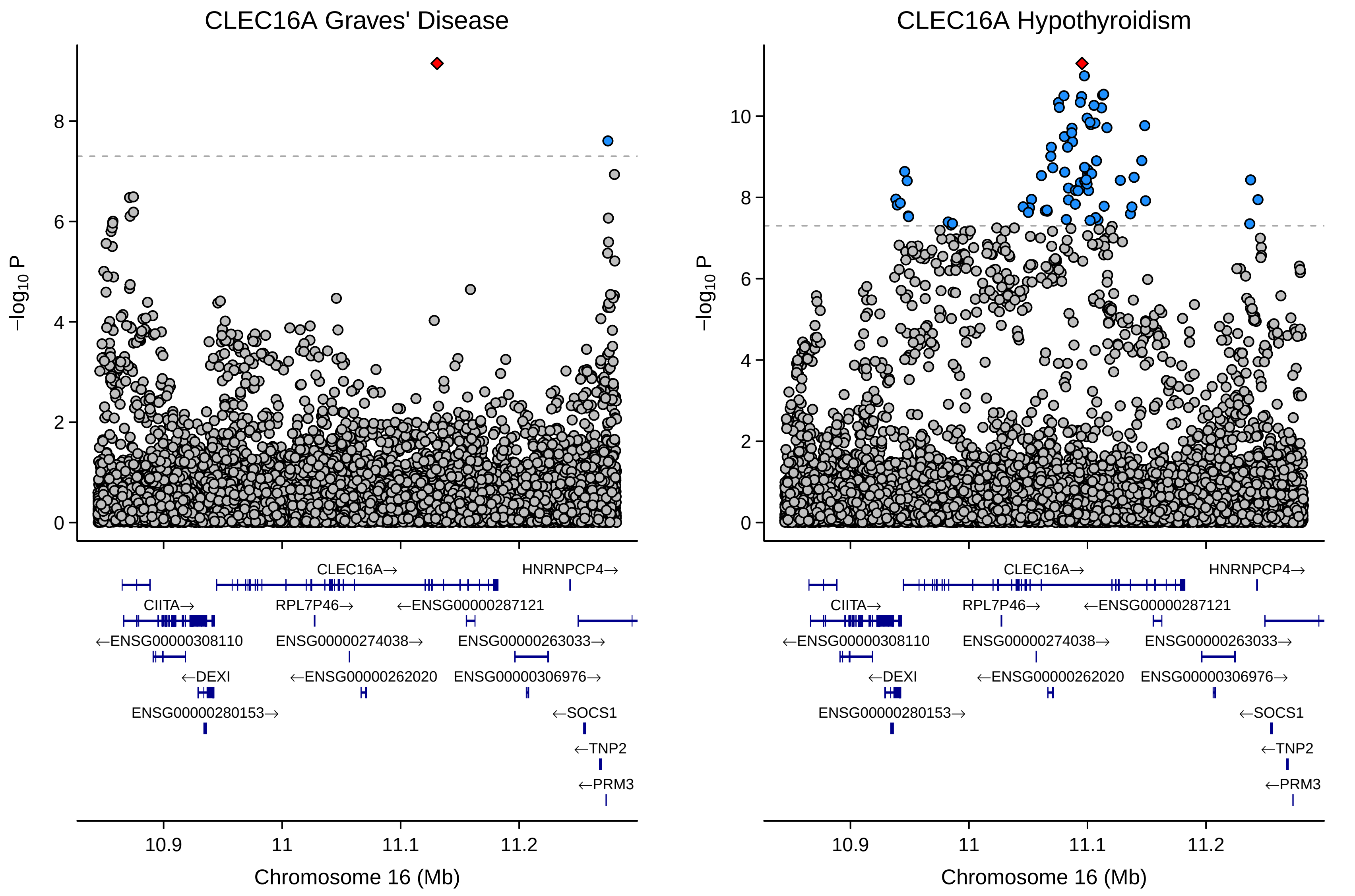

CLEC16A Graves' Disease
CLEC16A Hypothyroidism
−log10 P
0
2
4
6
8
10
CLEC16A→
HNRNPCP4→
CIITA→
RPL7P46→
←ENSG00000287121
←ENSG00000308110
ENSG00000274038→
ENSG00000263033→
←DEXI
←ENSG00000262020
ENSG00000306976→
ENSG00000280153→
←SOCS1
←TNP2
←PRM3
10.9
11
11.1
11.2
Chromosome 16 (Mb)

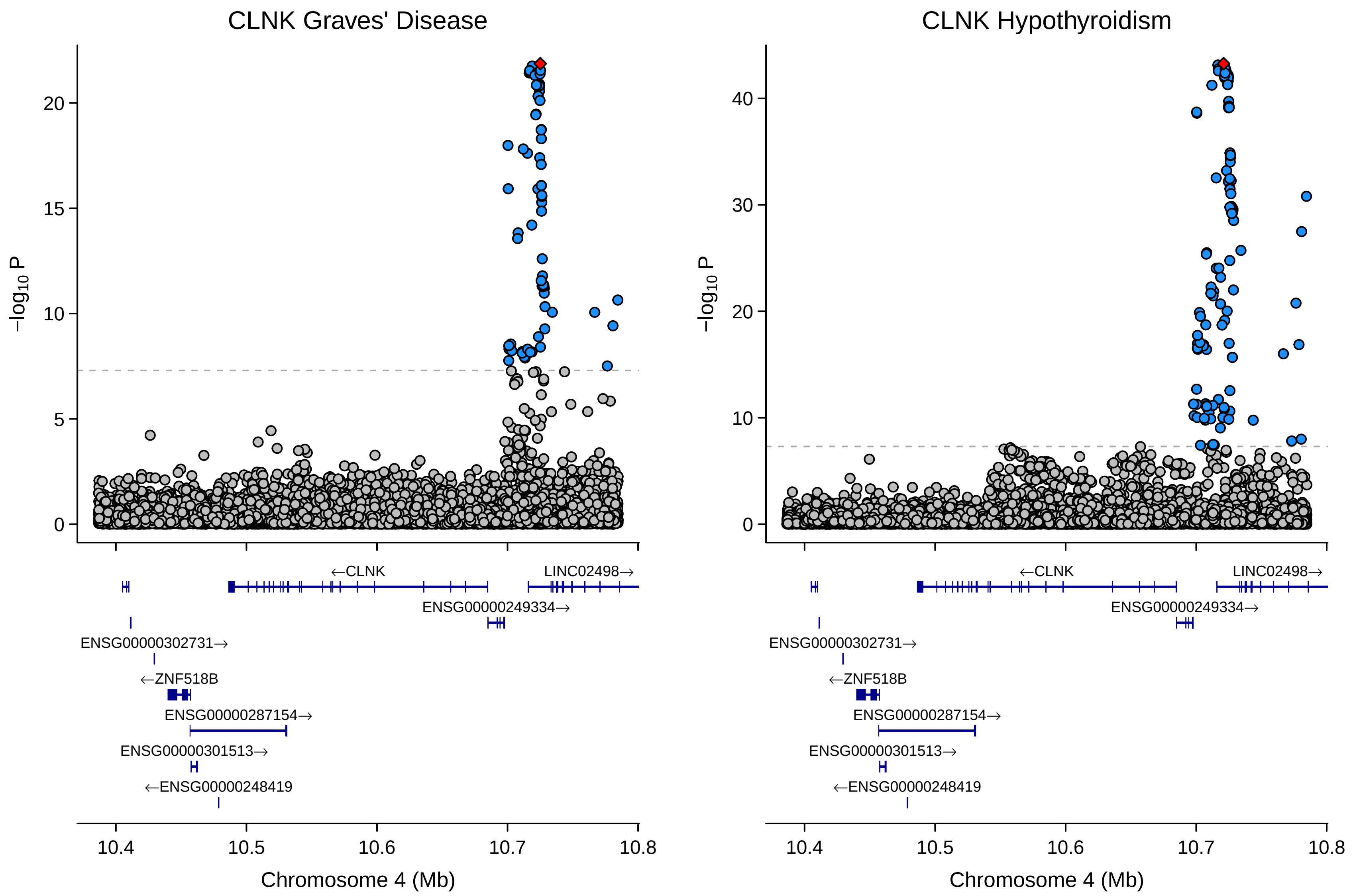

CLNK Graves' Disease
CLNK Hypothyroidism
−log10 P
0
5
10
15
20
30
40
←CLNK
LINC02498→
ENSG00000249334→
ENSG00000302731→
←ZNF518B
ENSG00000287154→
ENSG00000301513→
←ENSG00000248419
10.4
10.5
10.6
10.7
10.8
Chromosome 4 (Mb)

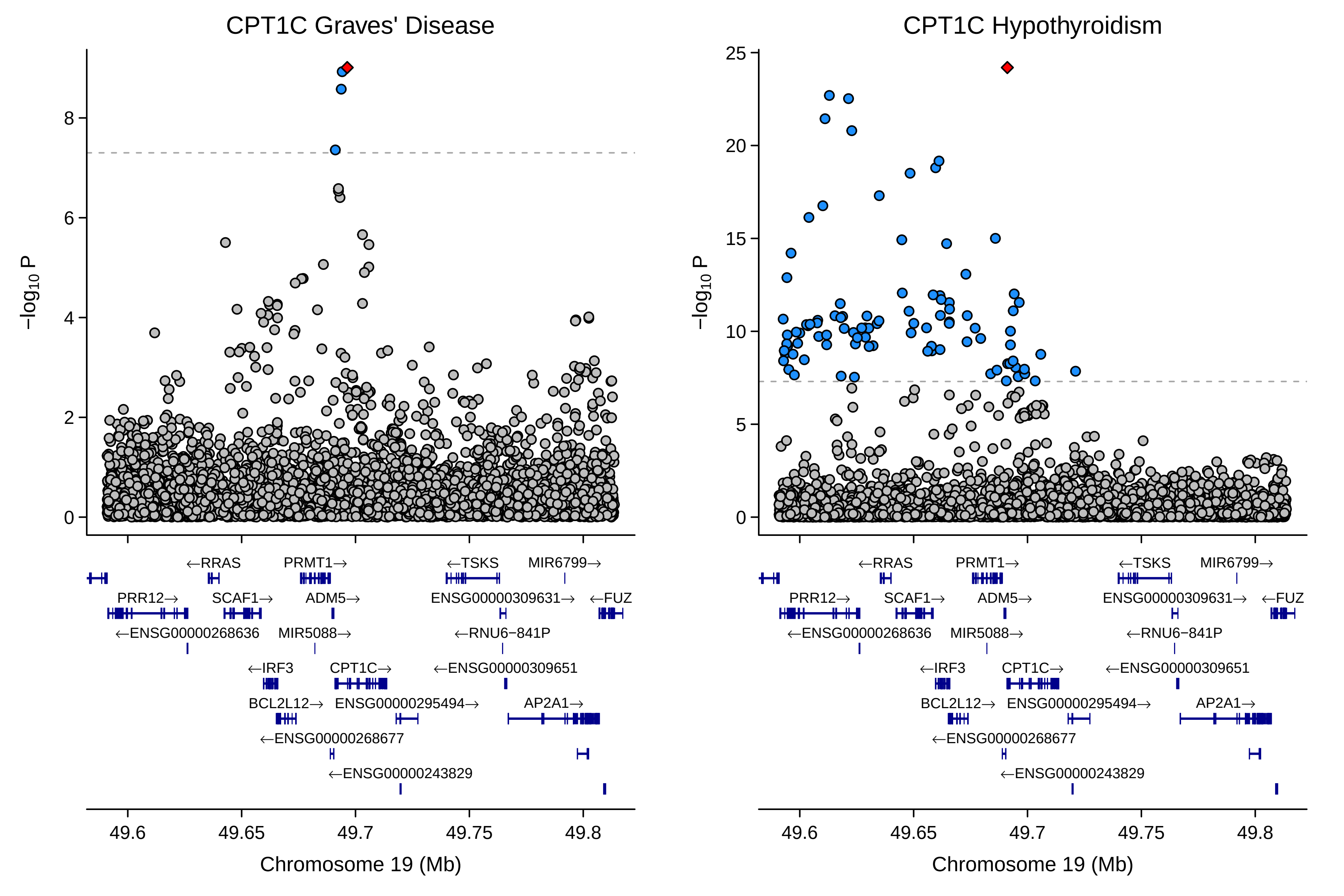
CPT1C Graves' Disease
CPT1C Hypothyroidism
−log10 P
Chromosome 19 (Mb)
49.6
49.65
49.7
49.75
49.8
←RRAS
PRMT1→
←TSKS
MIR6799→
PRR12→
SCAF1→
ADM5→
ENSG00000309631→
←FUZ
←ENSG00000268636
MIR5088→
←RNU6−841P
←IRF3
CPT1C→
←ENSG00000309651
BCL2L12→
ENSG00000295494→
AP2A1→
←ENSG00000268677
←ENSG00000243829

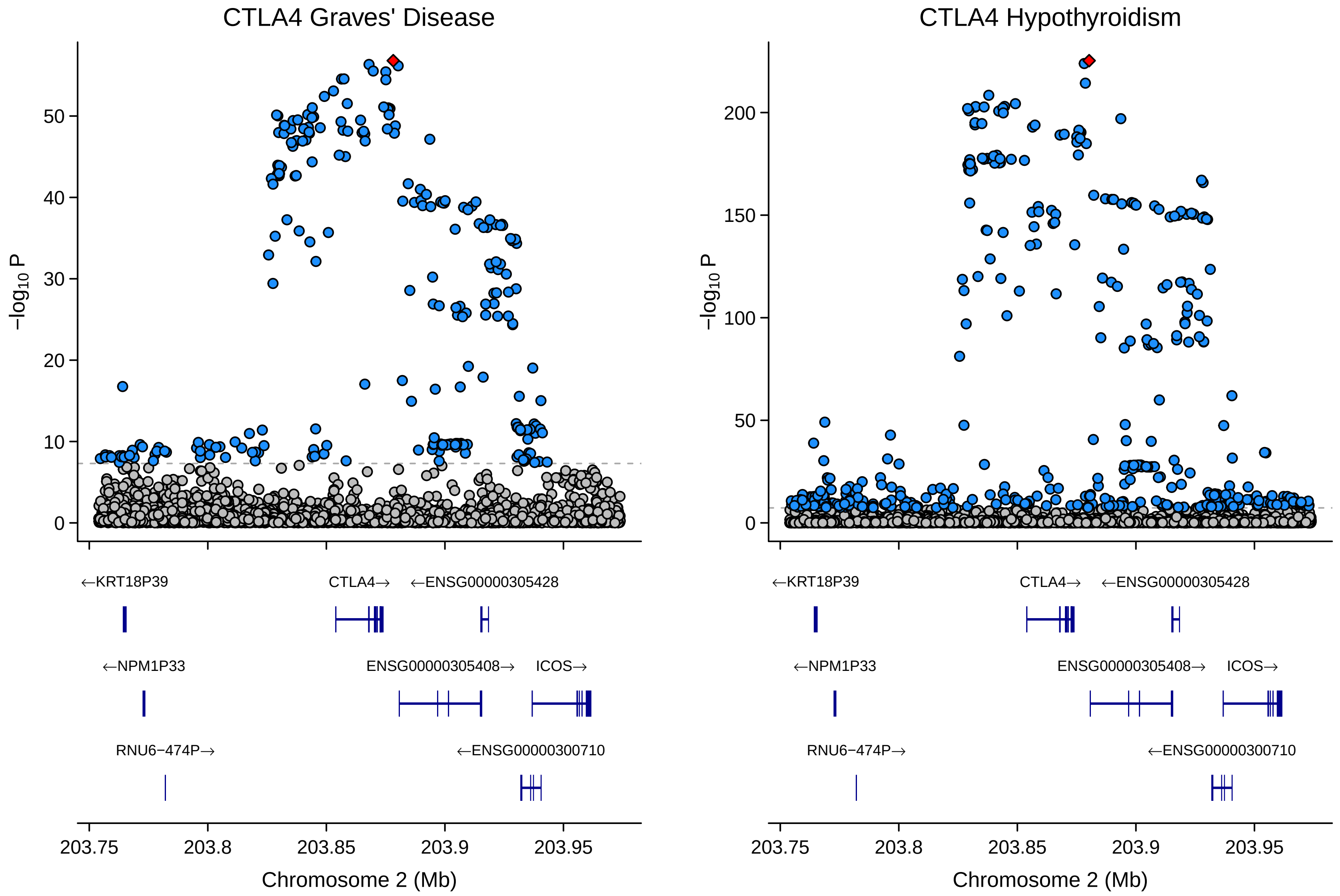

CTLA4 Graves' Disease
CTLA4 Hypothyroidism
−log10 P
0
10
20
30
40
50
0
50
100
150
200
←KRT18P39
CTLA4→
←ENSG00000305428
←NPM1P33
ENSG00000305408→
ICOS→
RNU6−474P→
←ENSG00000300710
203.75
203.8
203.85
203.9
203.95
Chromosome 2 (Mb)

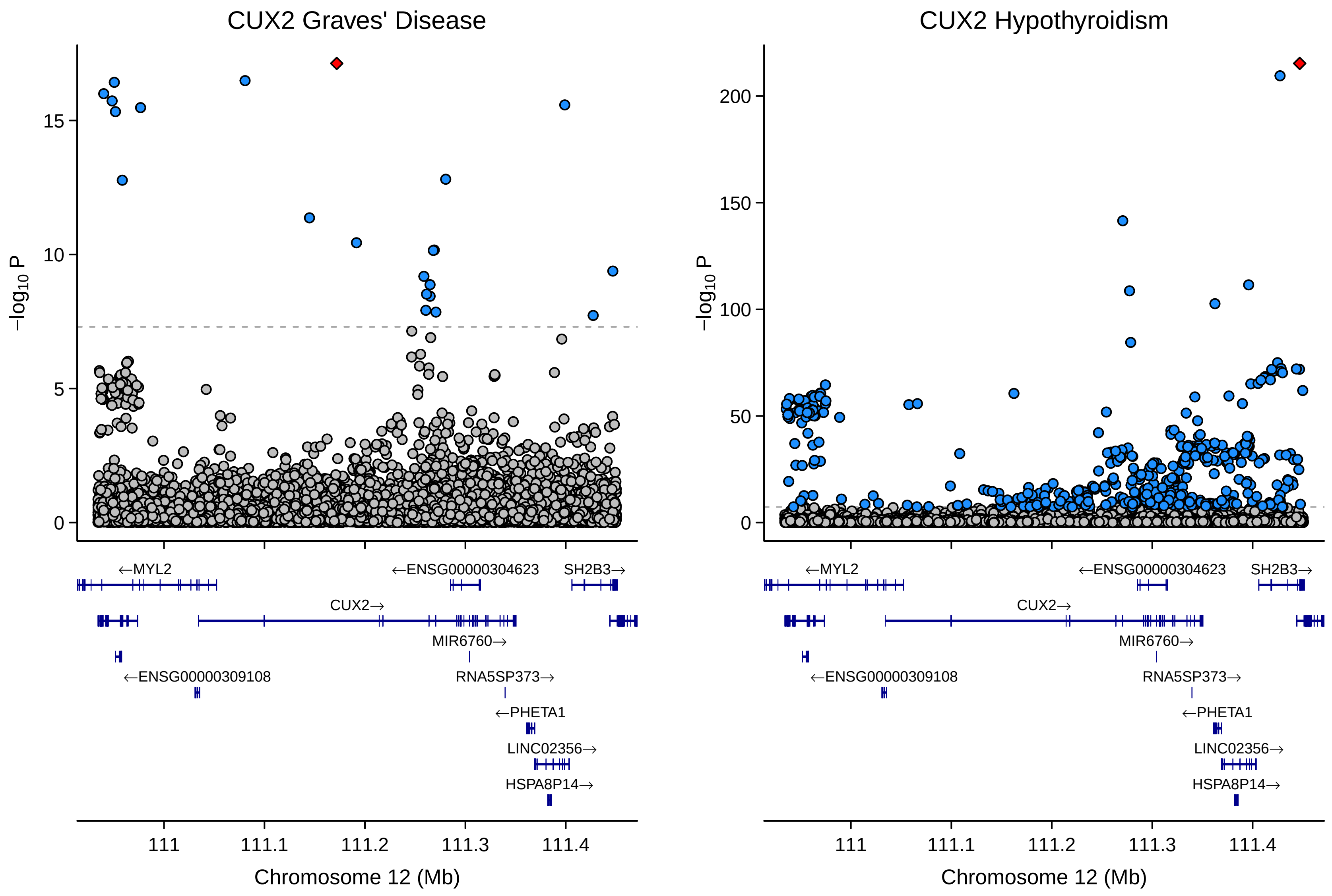

CUX2 Graves' Disease
CUX2 Hypothyroidism
−log10 P
0
5
10
15
50
100
150
200
←MYL2
←ENSG00000304623
SH2B3→
CUX2→
MIR6760→
←ENSG00000309108
RNA5SP373→
←PHETA1
LINC02356→
HSPA8P14→
111
111.1
111.2
111.3
111.4
Chromosome 12 (Mb)

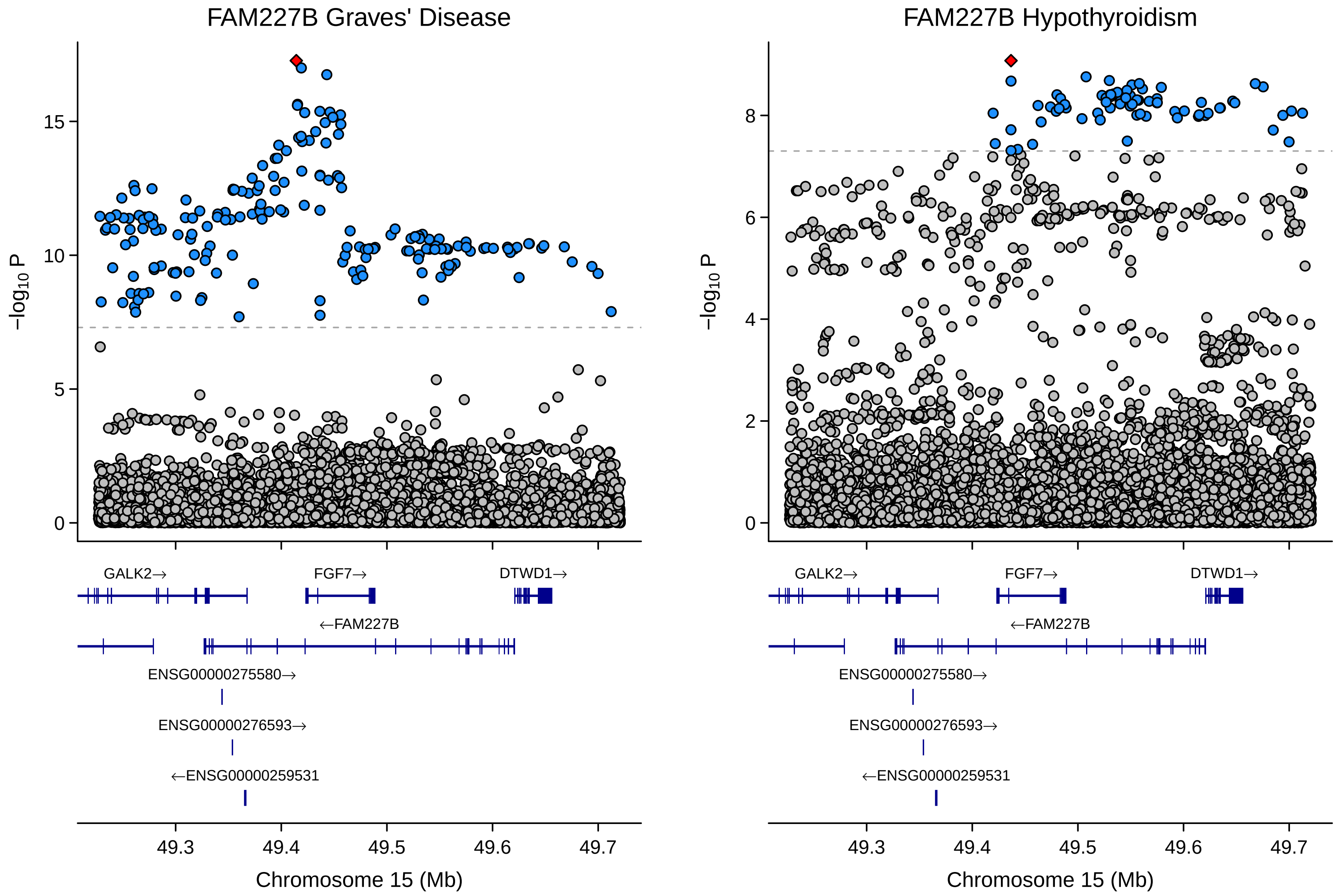
FAM227B Graves' Disease
FAM227B Hypothyroidism
−log10 P
0
5
10
15
2
4
6
8
GALK2→
FGF7→
DTWD1→
←FAM227B
ENSG00000275580→
ENSG00000276593→
←ENSG00000259531
49.3
49.4
49.5
49.6
49.7
Chromosome 15 (Mb)

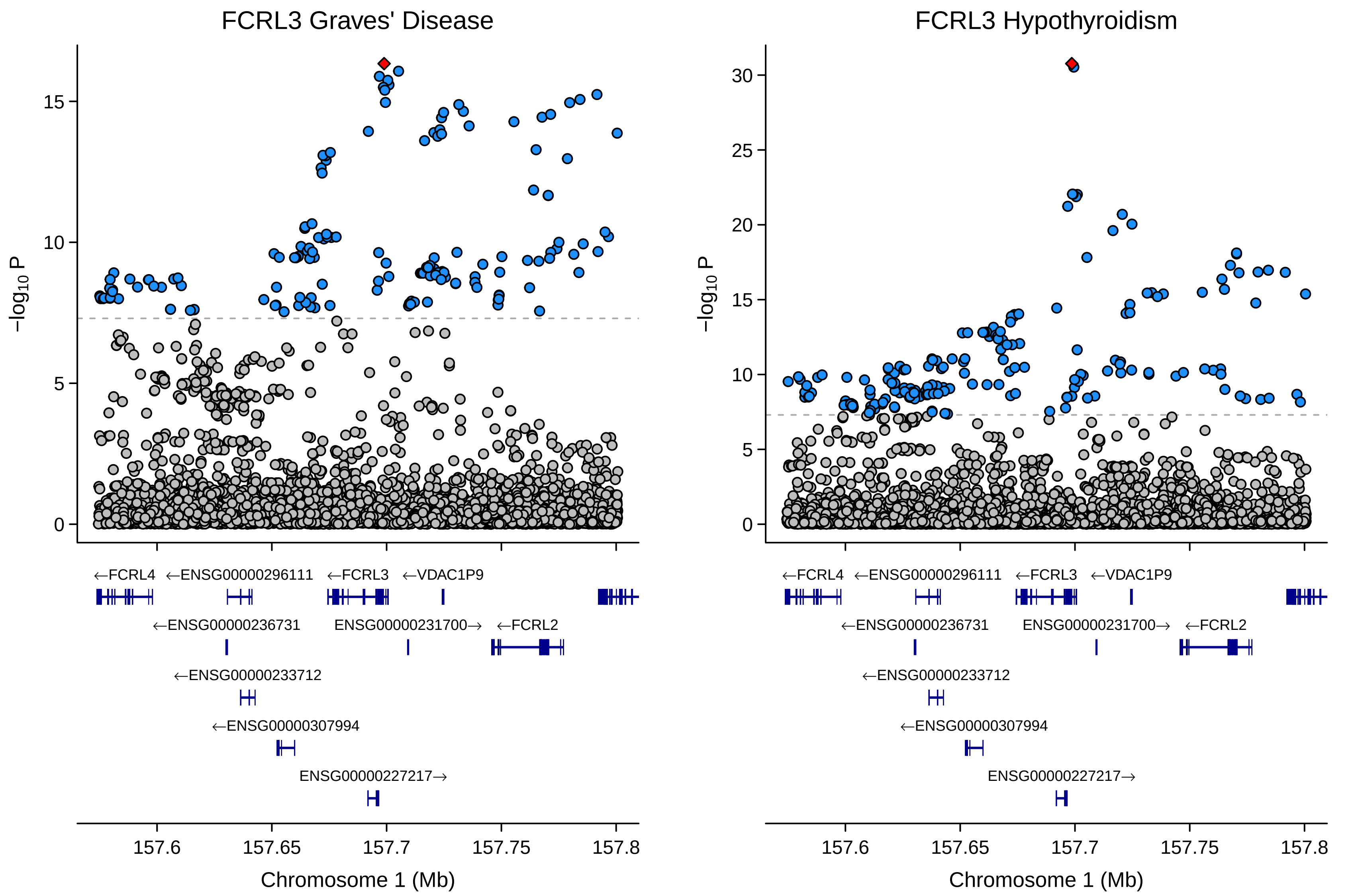

FCRL3 Graves' Disease
FCRL3 Hypothyroidism
−log10 P
0
5
10
15
20
25
30
←FCRL4
←ENSG00000296111
←FCRL3
←VDAC1P9
←ENSG00000236731
ENSG00000231700→
←FCRL2
←ENSG00000233712
←ENSG00000307994
ENSG00000227217→
157.6
157.65
157.7
157.75
157.8
Chromosome 1 (Mb)

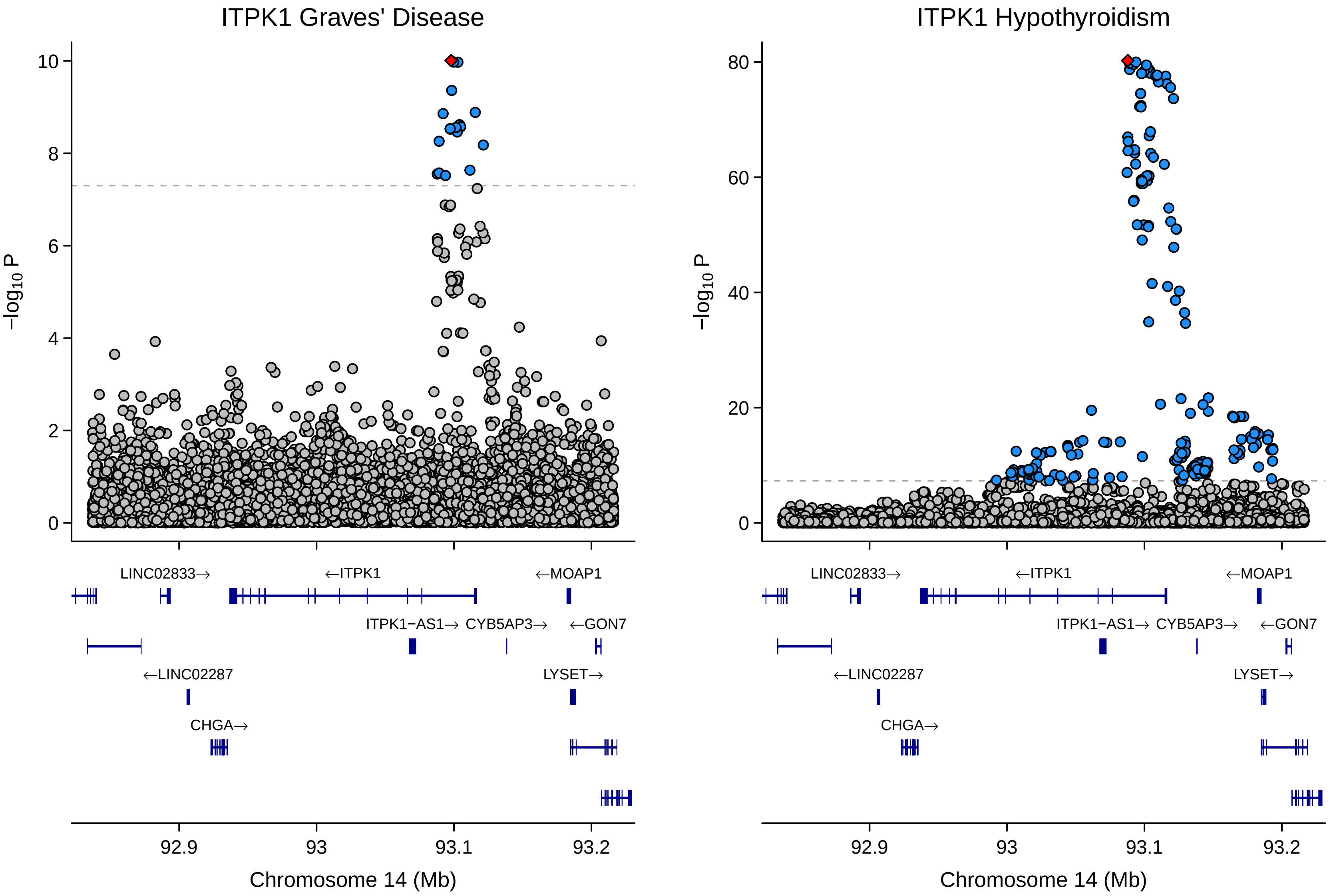
ITPK1 Graves' Disease
ITPK1 Hypothyroidism
−log10 P
10
8
6
4
2
0
80
60
40
20
LINC02833→
←ITPK1
←MOAP1
ITPK1−AS1→
CYB5AP3→
←GON7
←LINC02287
LYSET→
CHGA→
92.9
93
93.1
93.2
Chromosome 14 (Mb)

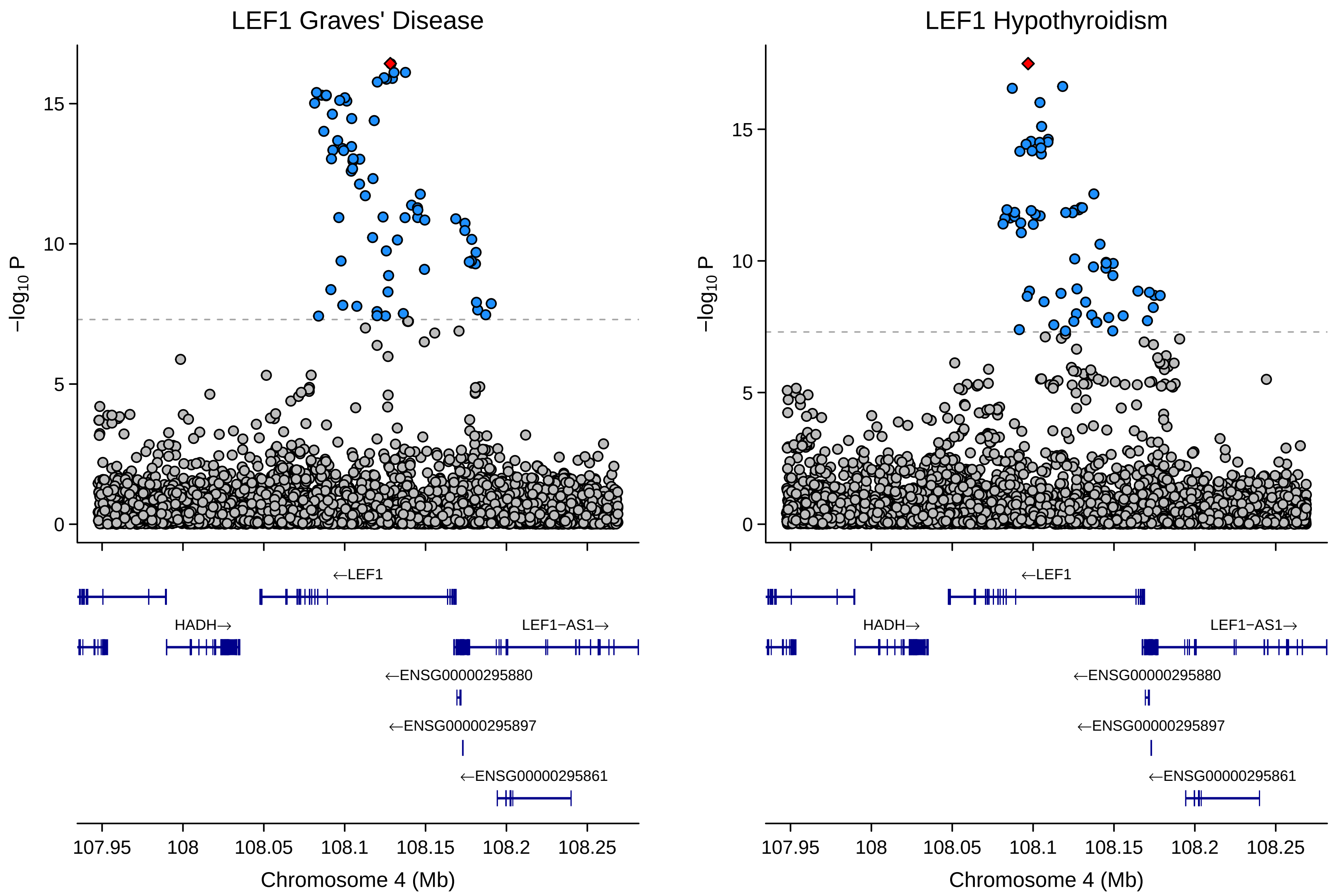

LEF1 Graves' Disease
LEF1 Hypothyroidism
−log10 P
15
10
5
0
←LEF1
HADH→
LEF1−AS1→
←ENSG00000295880
←ENSG00000295897
←ENSG00000295861
107.95
108
108.05
108.1
108.15
108.2
108.25
Chromosome 4 (Mb)

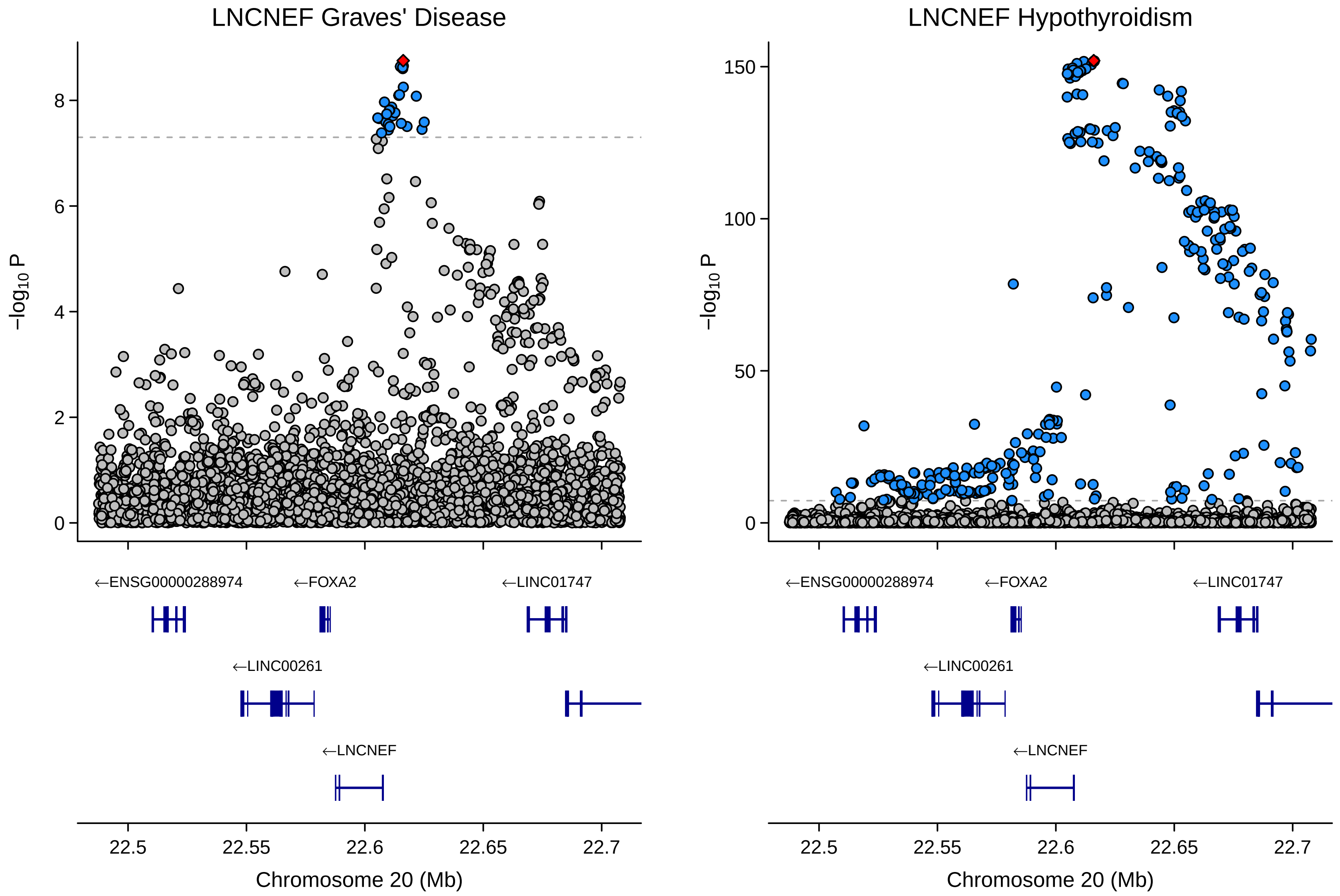

LNCNEF Graves' Disease
LNCNEF Hypothyroidism
−log10 P
0
2
4
6
8
0
50
100
150
←ENSG00000288974
←FOXA2
←LINC01747
←LINC00261
←LNCNEF
22.5
22.55
22.6
22.65
22.7
Chromosome 20 (Mb)

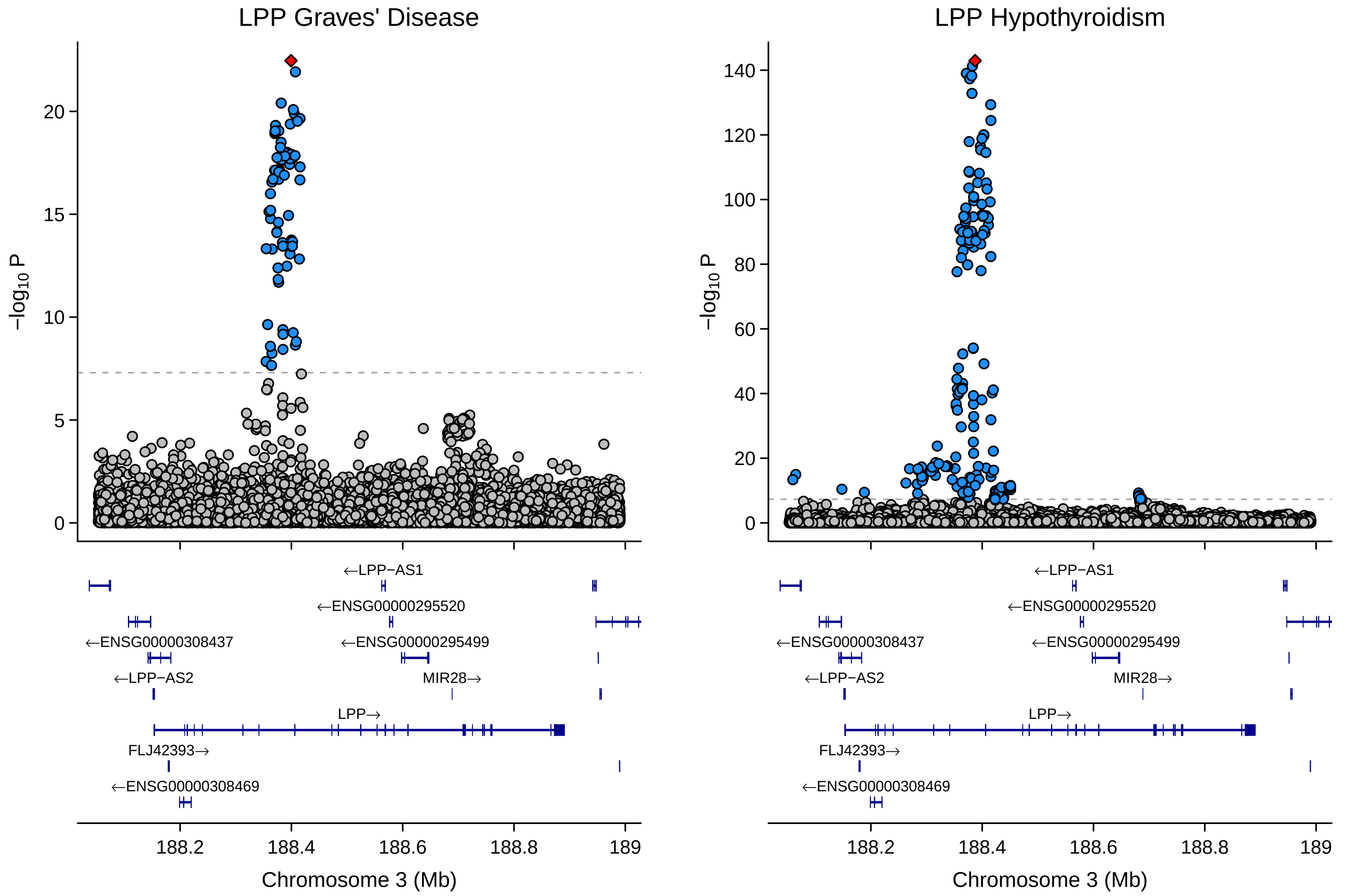
LPP Graves' Disease
LPP Hypothyroidism
−log10 P
Chromosome 3 (Mb)
188.2
188.4
188.6
188.8
189
←LPP−AS1
←ENSG00000295520
←ENSG00000308437
←ENSG00000295499
←LPP−AS2
MIR28→
LPP→
FLJ42393→
←ENSG00000308469

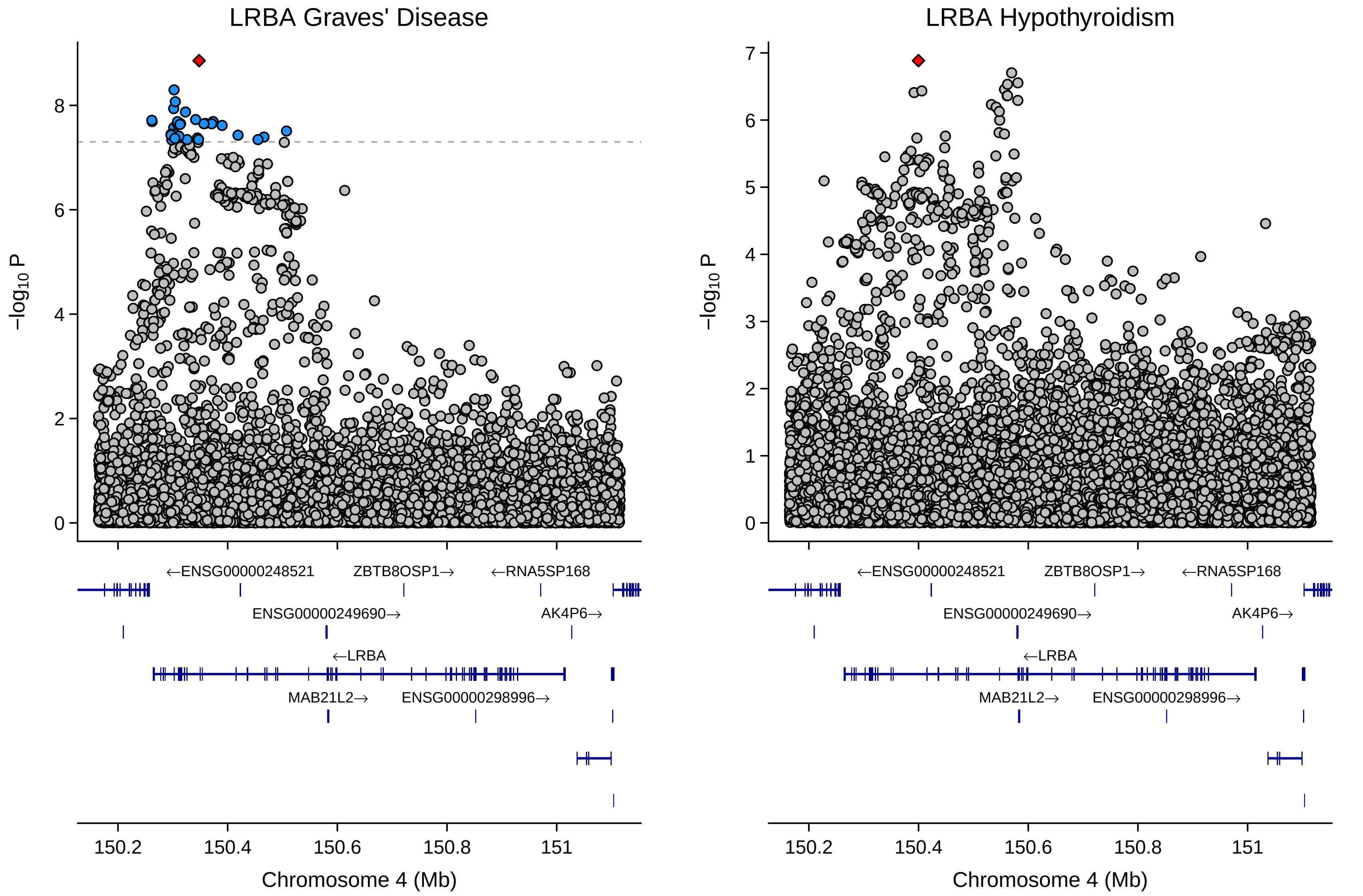
LRBA Graves' Disease
LRBA Hypothyroidism
−log10 P
Chromosome 4 (Mb)
150.2
150.4
150.6
150.8
151
←ENSG00000248521
ZBTB8OSP1→
←RNA5SP168
ENSG00000249690→
AK4P6→
←LRBA
MAB21L2→
ENSG00000298996→

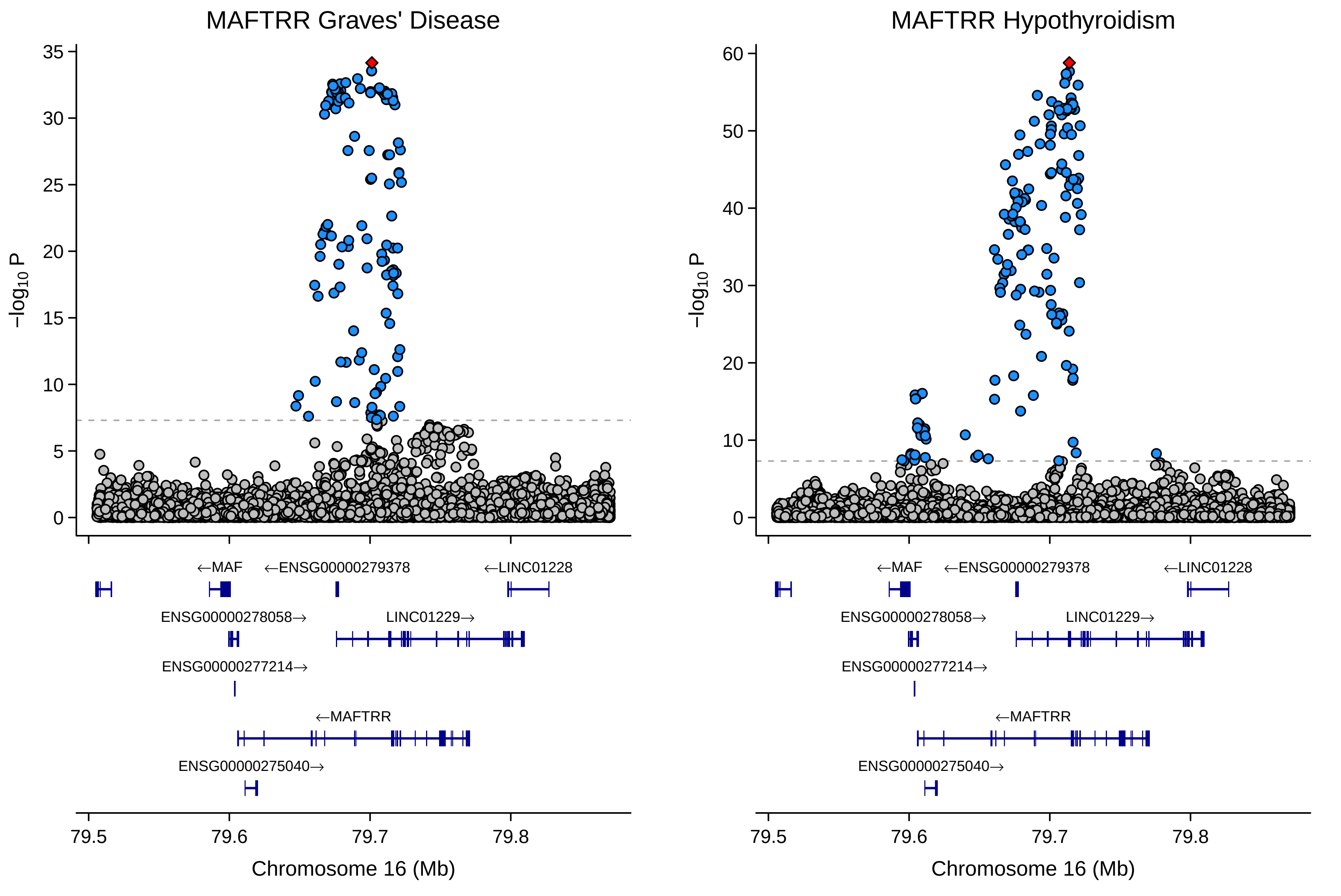

MAFTRR Graves' Disease
MAFTRR Hypothyroidism
−log10 P
35
30
25
20
15
10
5
0
60
50
40
30
20
10
←MAF
←ENSG00000279378
←LINC01228
ENSG00000278058→
LINC01229→
ENSG00000277214→
←MAFTRR
ENSG00000275040→
79.5
79.6
79.7
79.8
Chromosome 16 (Mb)

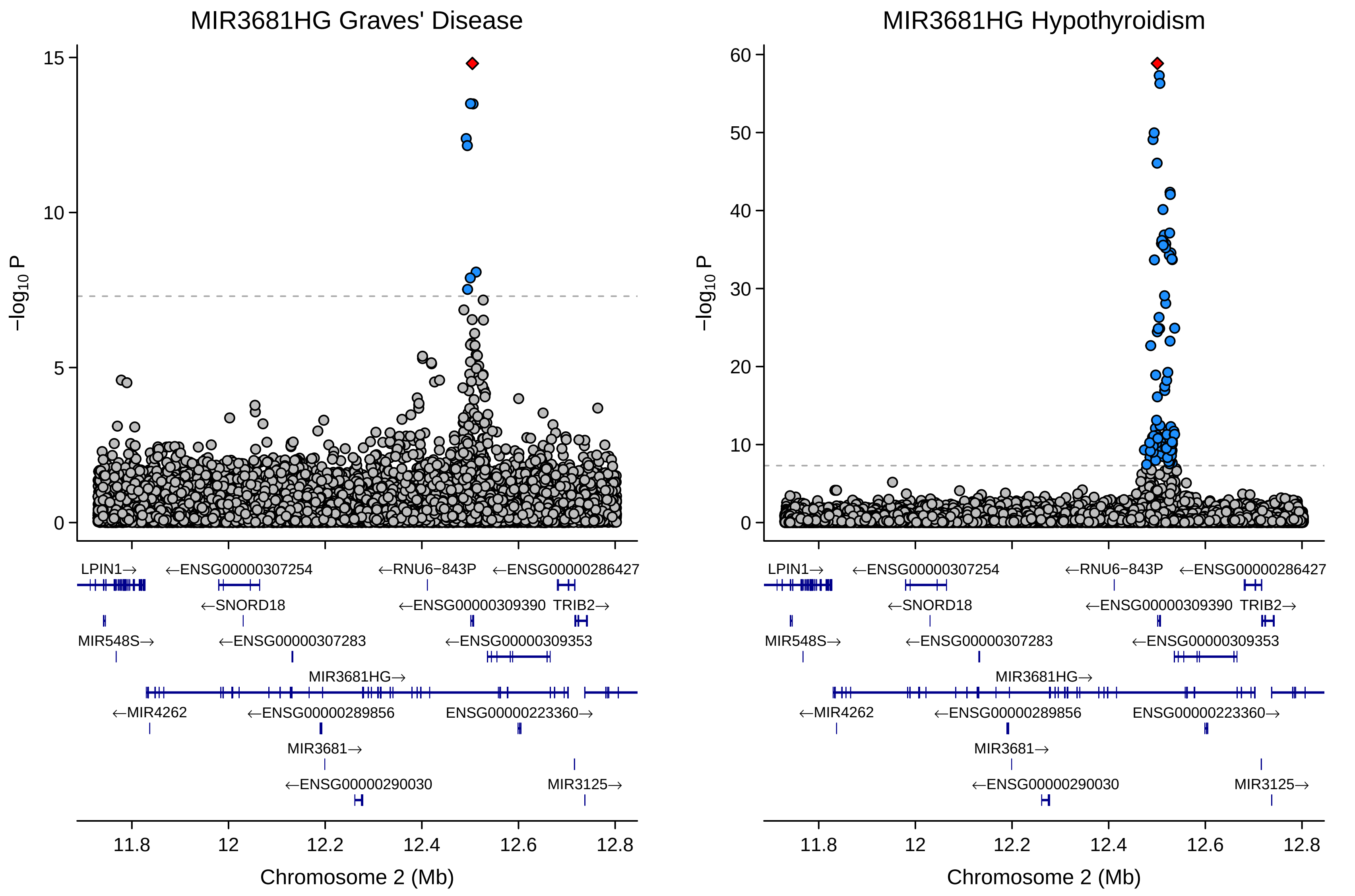

MIR3681HG Graves' Disease
MIR3681HG Hypothyroidism
−log10 P
Chromosome 2 (Mb)
LPIN1→
←ENSG00000307254
←RNU6−843P
←ENSG00000286427
←SNORD18
←ENSG00000309390
TRIB2→
MIR548S→
←ENSG00000307283
←ENSG00000309353
MIR3681HG→
←MIR4262
←ENSG00000289856
ENSG00000223360→
MIR3681→
←ENSG00000290030
MIR3125→

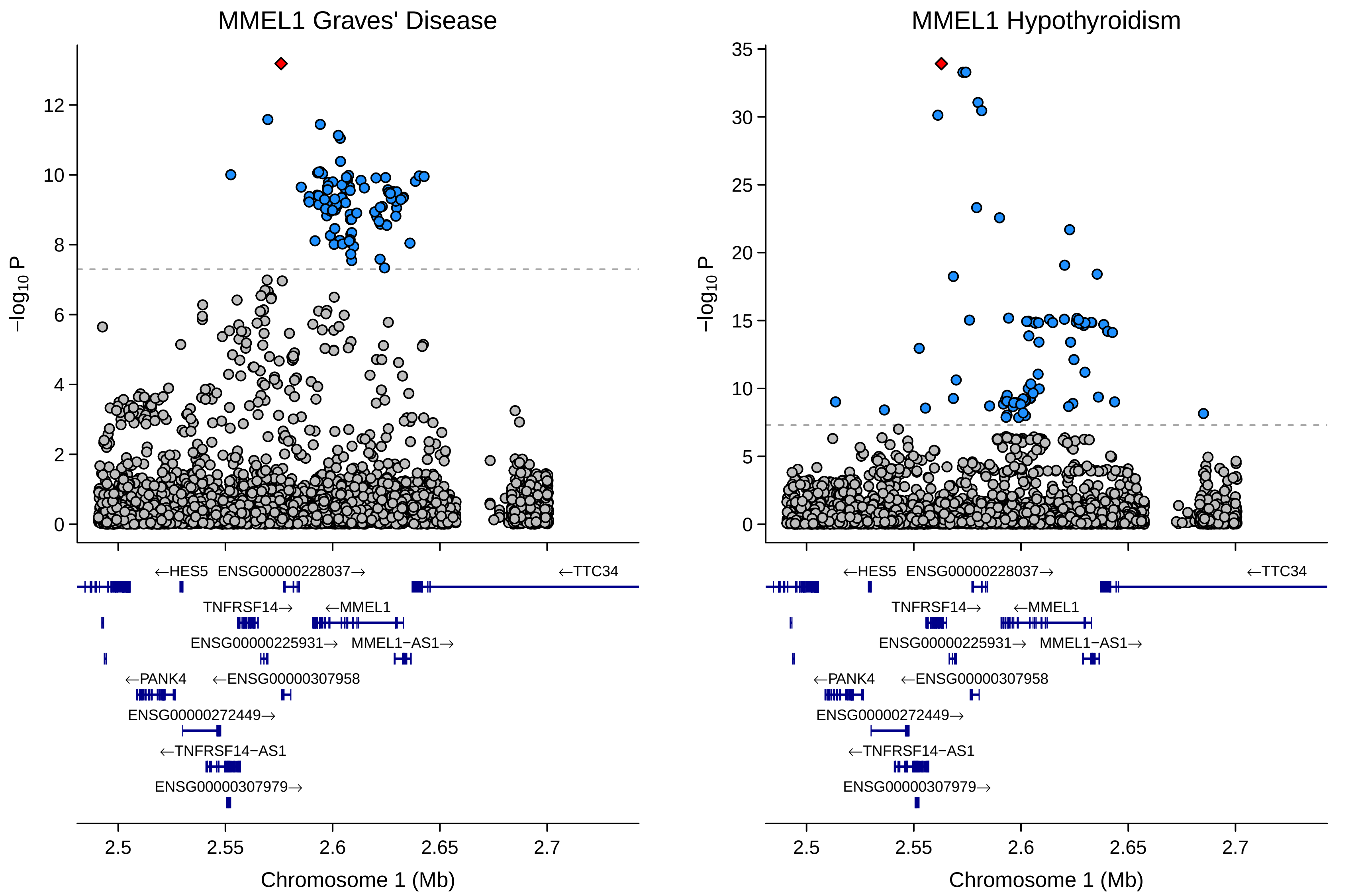

MMEL1 Graves' Disease
MMEL1 Hypothyroidism
−log10 P
Chromosome 1 (Mb)
2.5
2.55
2.6
2.65
2.7
←HES5
ENSG00000228037→
←TTC34
TNFRSF14→
←MMEL1
ENSG00000225931→
MMEL1−AS1→
←PANK4
←ENSG00000307958
ENSG00000272449→
←TNFRSF14−AS1
ENSG00000307979→

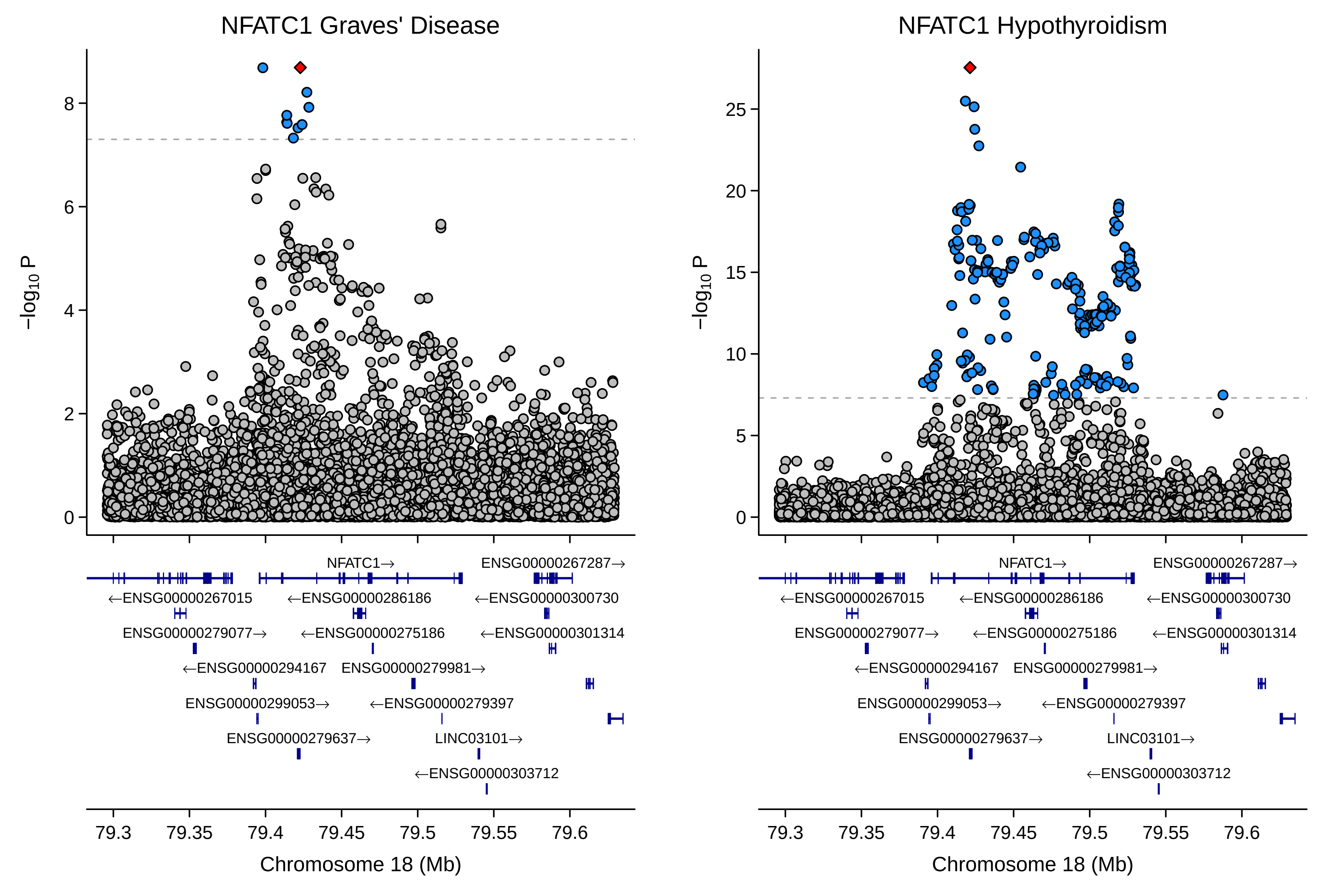
NFATC1 Graves' Disease
NFATC1 Hypothyroidism
−log10 P
Chromosome 18 (Mb)
NFATC1→
ENSG00000267287→
←ENSG00000267015
←ENSG00000286186
←ENSG00000300730
ENSG00000279077→
←ENSG00000275186
←ENSG00000301314
←ENSG00000294167
ENSG00000279981→
ENSG00000299053→
←ENSG00000279397
ENSG00000279637→
LINC03101→
←ENSG00000303712

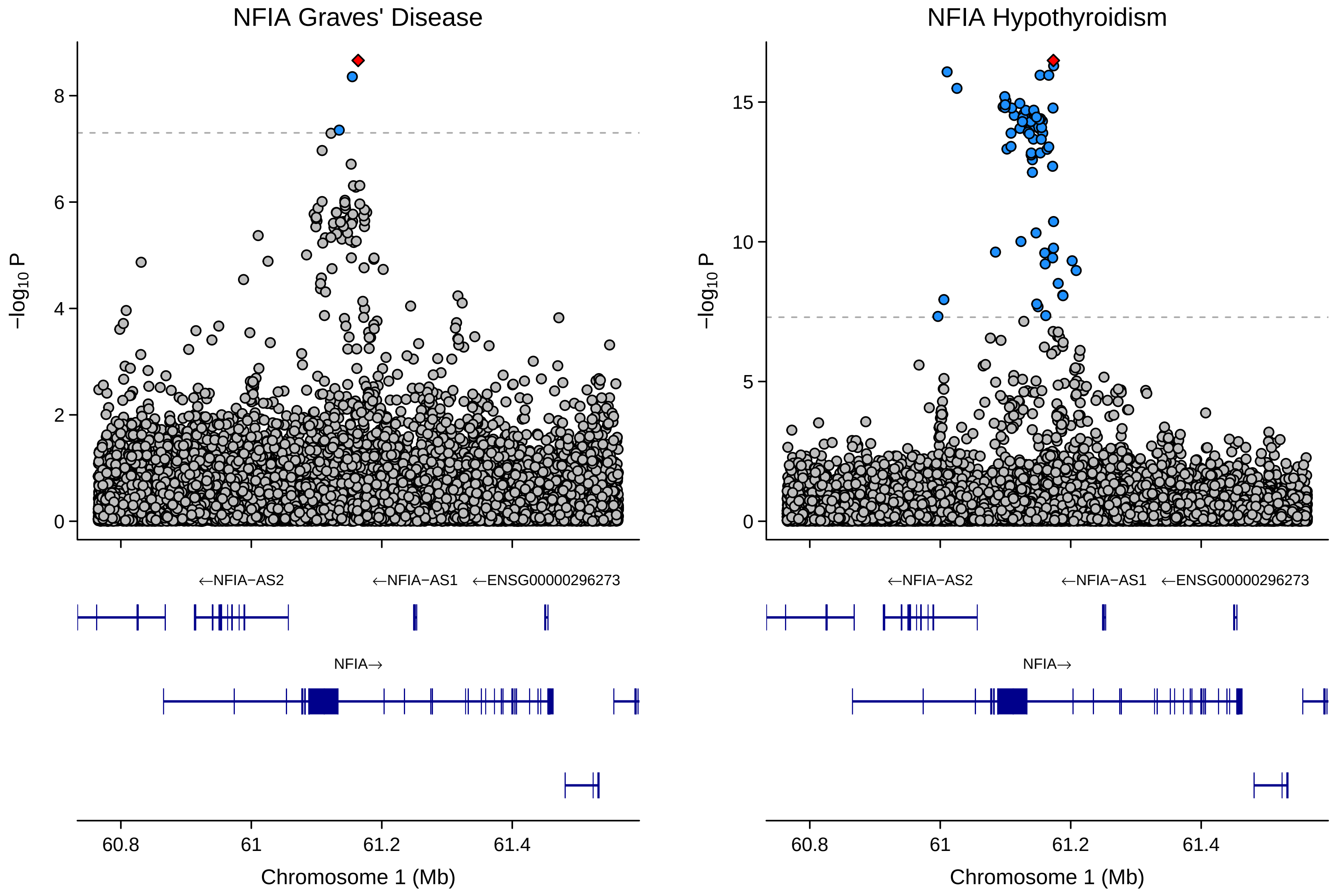
NFIA Graves' Disease
NFIA Hypothyroidism
−log10 P
8
6
4
2
0
15
10
5
0
←NFIA−AS2
←NFIA−AS1
←ENSG00000296273
NFIA→
60.8
61
61.2
61.4
Chromosome 1 (Mb)

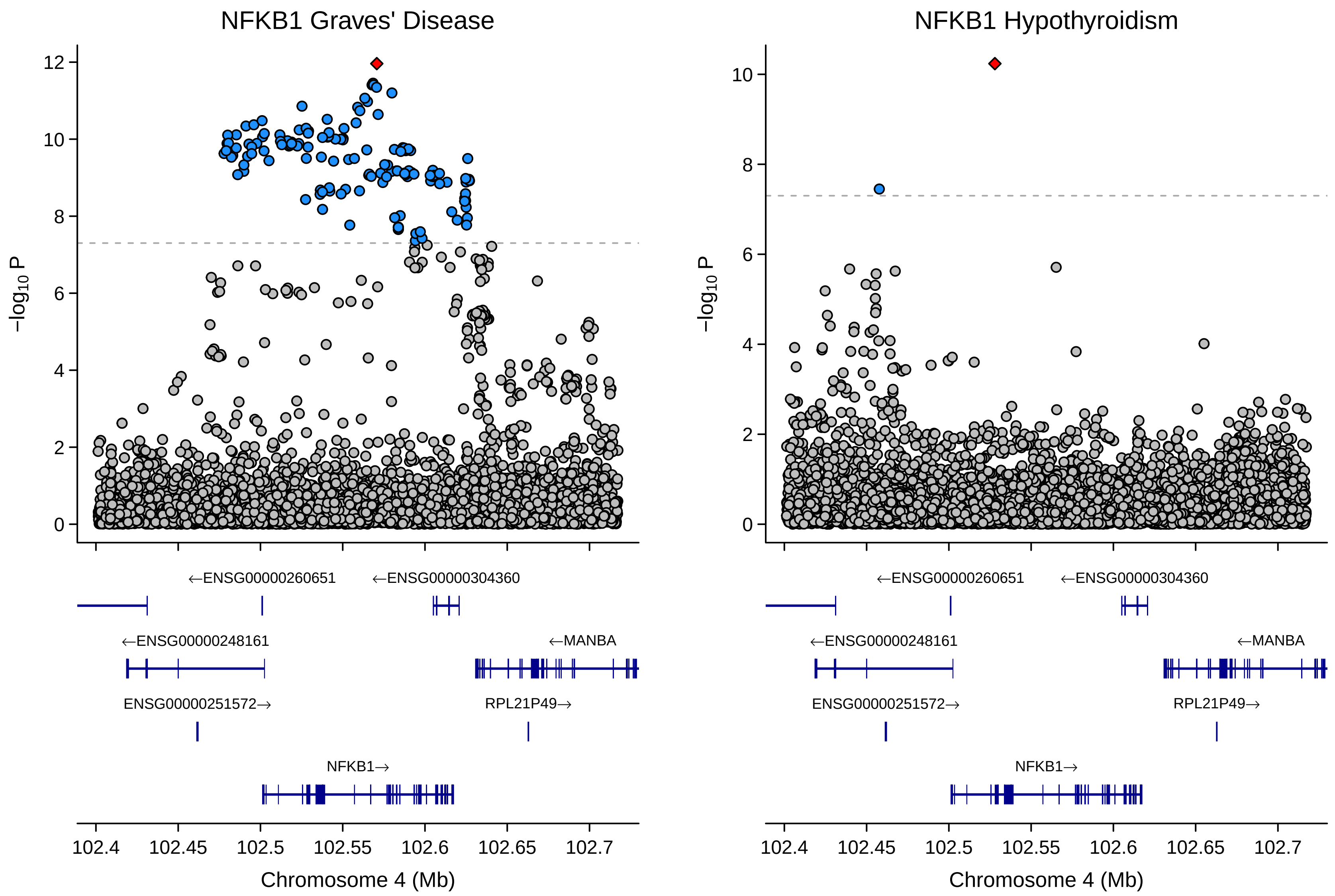
NFKB1 Graves' Disease
NFKB1 Hypothyroidism
−log10 P
Chromosome 4 (Mb)
102.4
102.45
102.5
102.55
102.6
102.65
102.7
←ENSG00000260651
←ENSG00000304360
←ENSG00000248161
←MANBA
ENSG00000251572→
RPL21P49→
NFKB1→

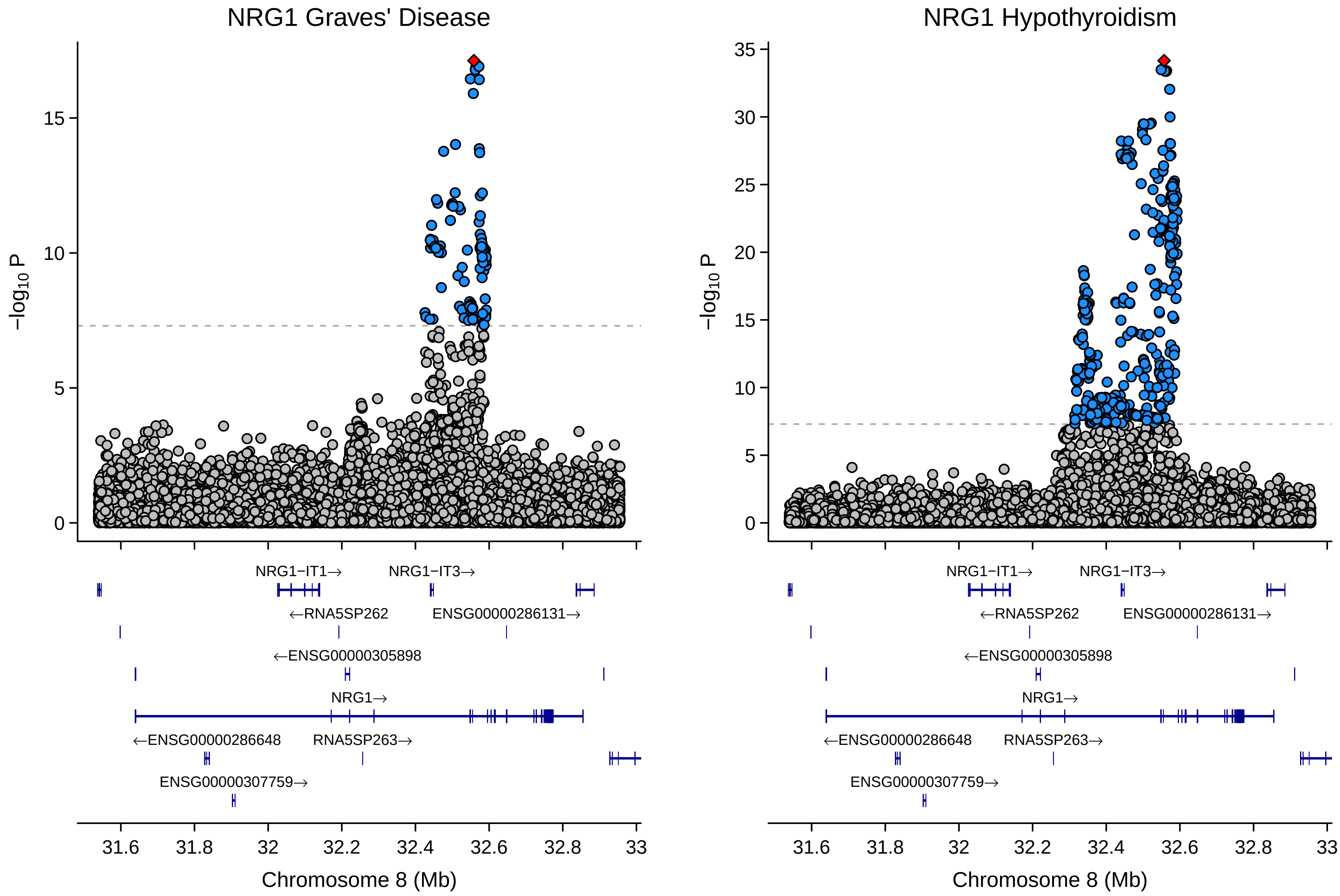
NRG1 Graves' Disease
NRG1 Hypothyroidism
−log10 P
Chromosome 8 (Mb)
NRG1−IT1→
NRG1−IT3→
←RNA5SP262
ENSG00000286131→
←ENSG00000305898
NRG1→
←ENSG00000286648
RNA5SP263→
ENSG00000307759→
31.6
31.8
32
32.2
32.4
32.6
32.8
33
0
5
10
15
20
25
30
35

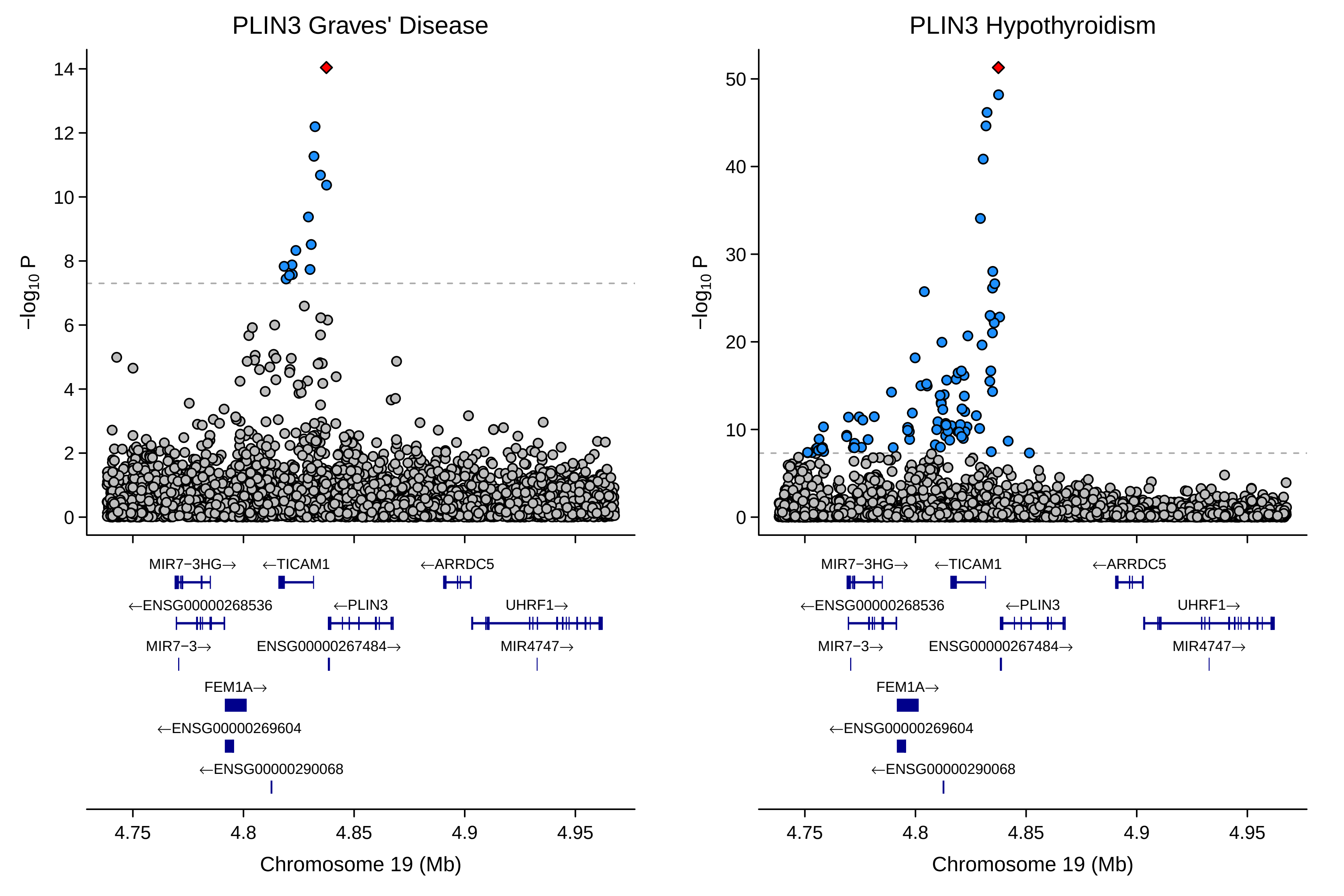
PLIN3 Graves' Disease
PLIN3 Hypothyroidism
−log10 P
MIR7−3HG→
←TICAM1
←ARRDC5
←ENSG00000268536
←PLIN3
UHRF1→
MIR7−3→
ENSG00000267484→
MIR4747→
FEM1A→
←ENSG00000269604
←ENSG00000290068
4.75
4.8
4.85
4.9
4.95
Chromosome 19 (Mb)

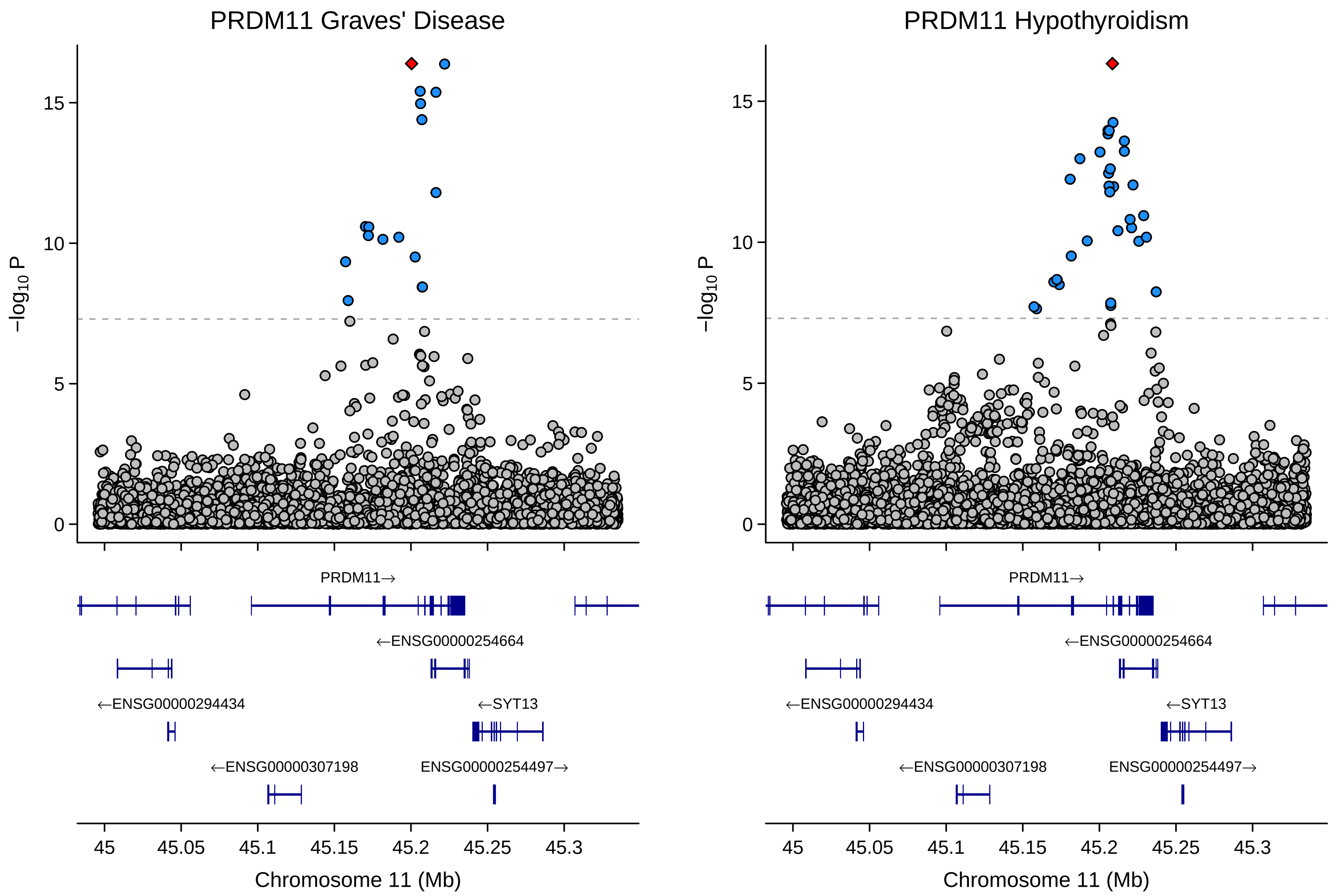
PRDM11 Graves' Disease
PRDM11 Hypothyroidism
−log10 P
0
5
10
15
PRDM11→
←ENSG00000254664
←ENSG00000294434
←SYT13
←ENSG00000307198
ENSG00000254497→
45
45.05
45.1
45.15
45.2
45.25
45.3
Chromosome 11 (Mb)

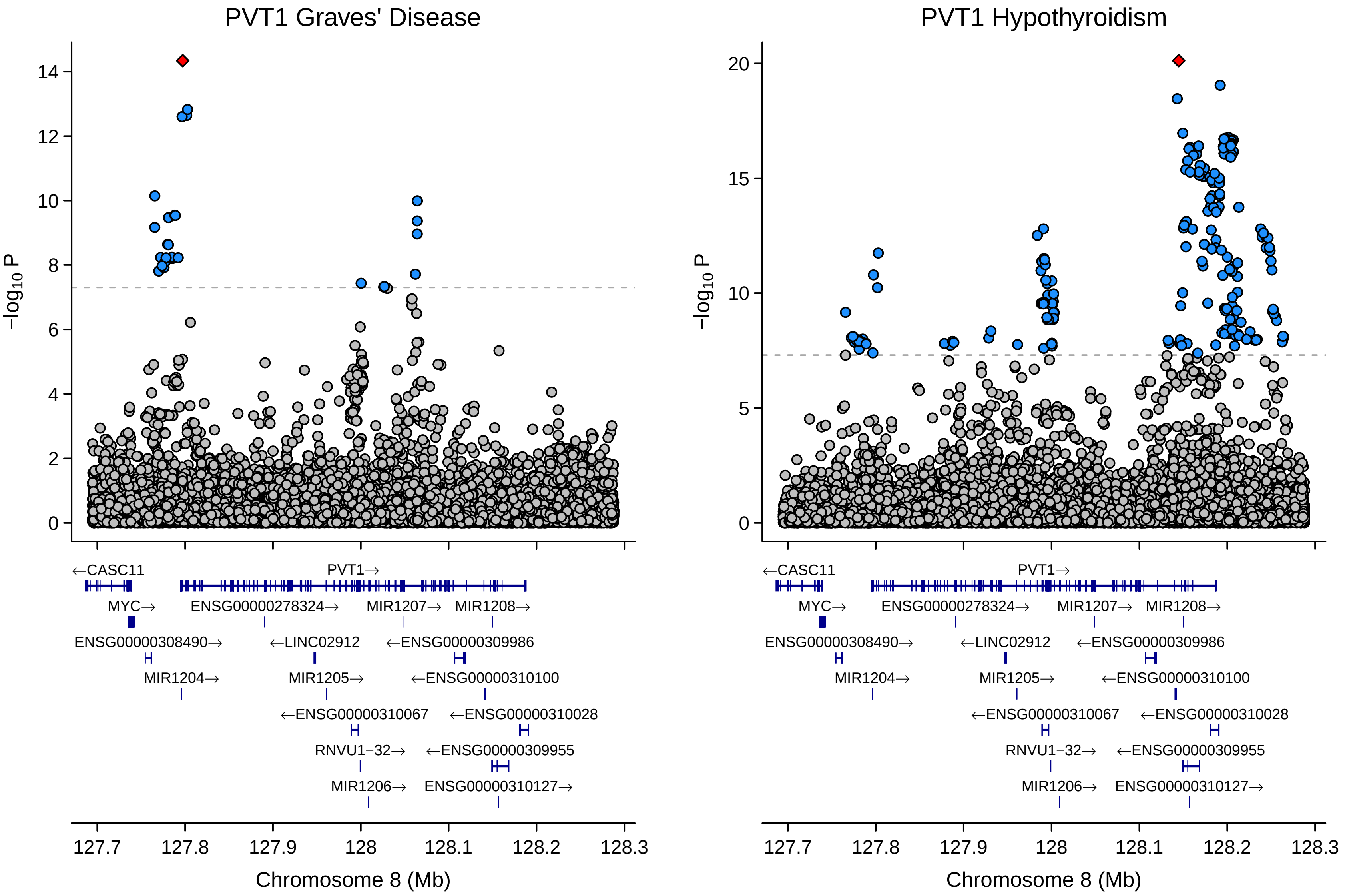
PVT1 Graves' Disease
PVT1 Hypothyroidism
$-\log_{10} P$
14
12
10
8
6
4
2
0
20
15
5
←CASC11
PVT1→
MYC→
ENSG00000278324→
MIR1207→
MIR1208→
ENSG00000308490→
←LINC02912
←ENSG00000309986
MIR1204→
MIR1205→
←ENSG00000310100
←ENSG00000310067
←ENSG00000310028
RNVU1−32→
←ENSG00000309955
MIR1206→
ENSG00000310127→
127.7
127.8
127.9
128
128.1
128.2
128.3
Chromosome 8 (Mb)

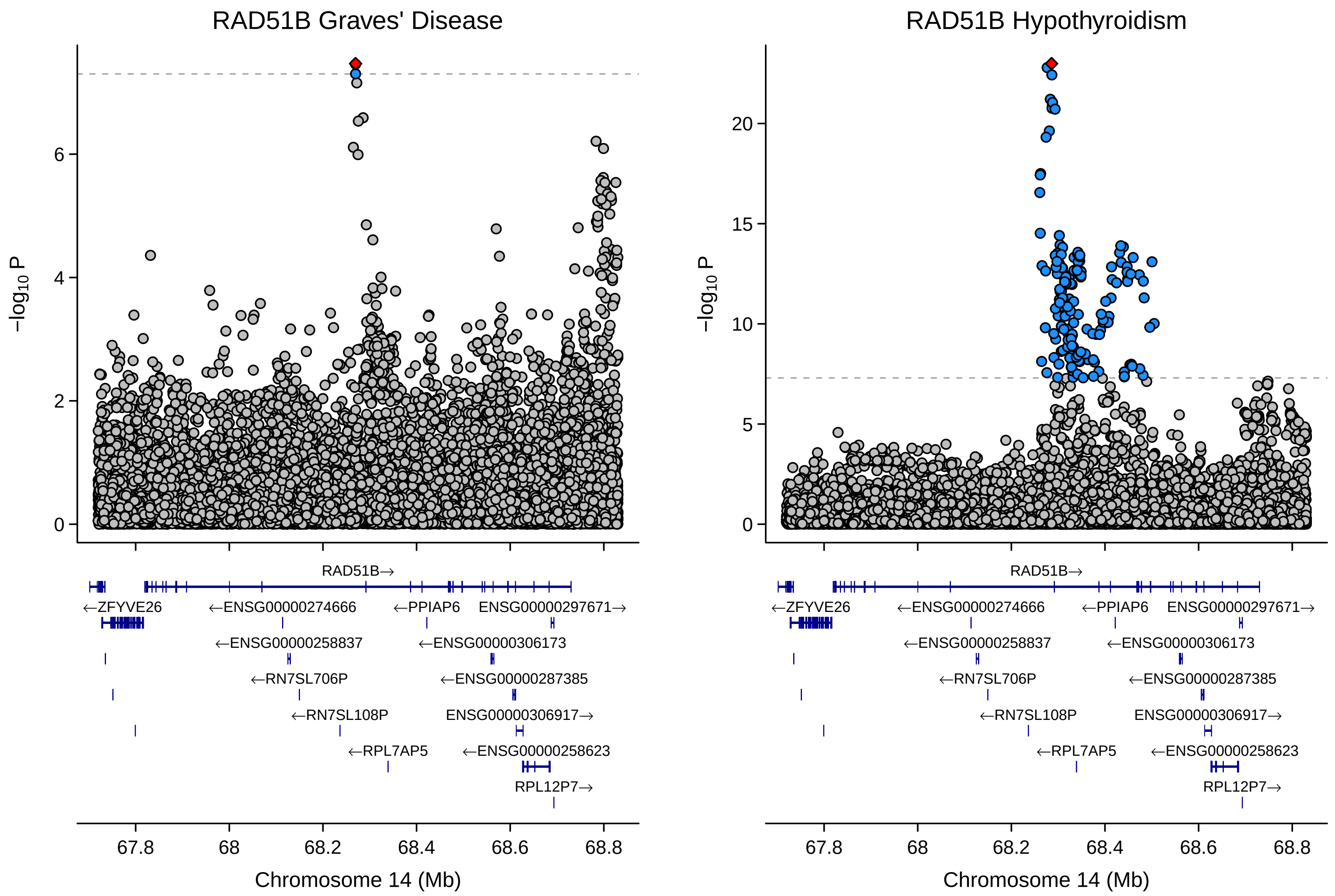
RAD51B Graves' Disease
RAD51B Hypothyroidism
−log10 P
0
2
4
6
5
10
15
20
RAD51B→
←ZFYVE26
←ENSG00000274666
←PPIAP6
ENSG00000297671→
←ENSG00000258837
←ENSG00000306173
←RN7SL706P
←ENSG00000287385
←RN7SL108P
ENSG00000306917→
←RPL7AP5
←ENSG00000258623
RPL12P7→
67.8
68
68.2
68.4
68.6
68.8
Chromosome 14 (Mb)

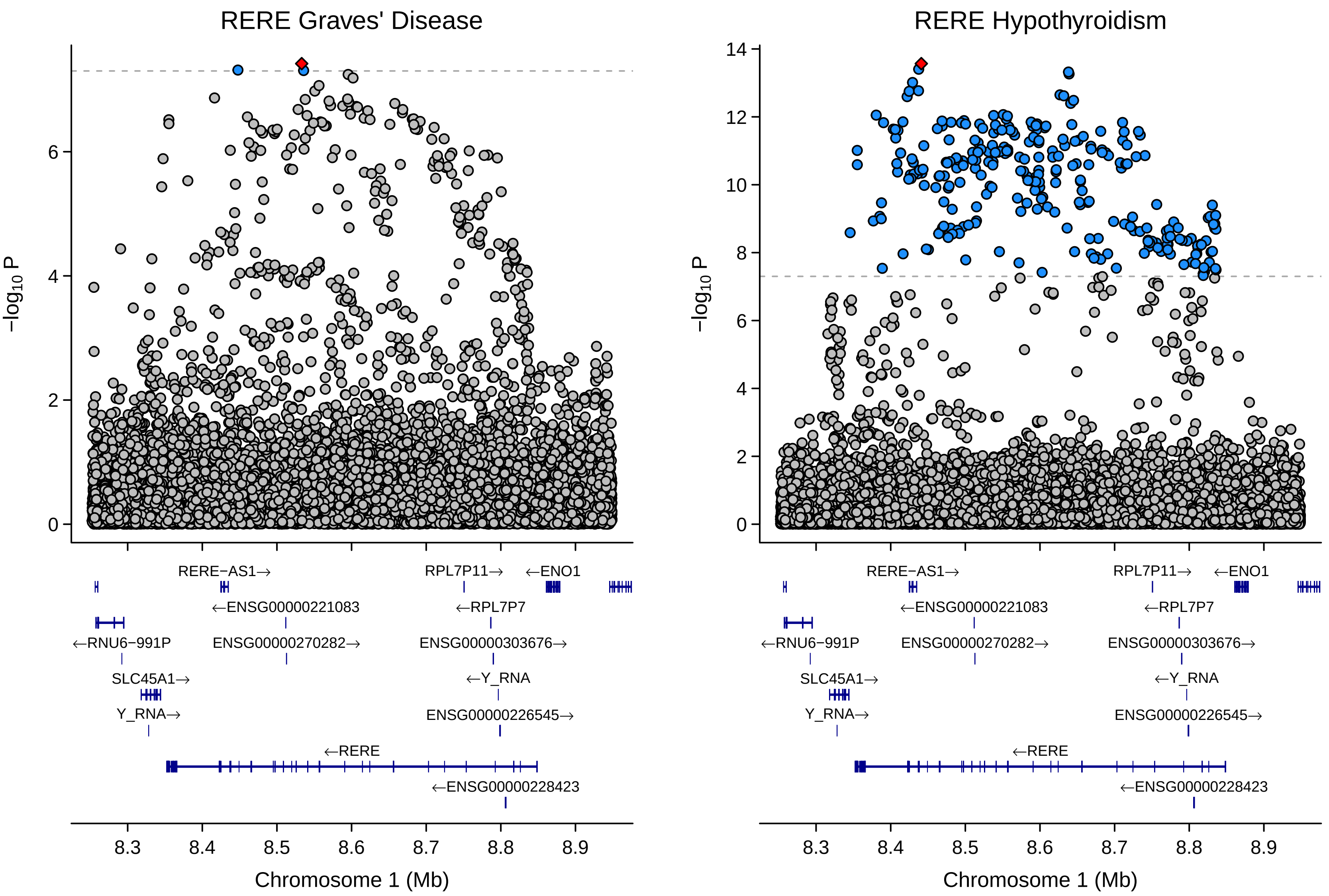
RERE Graves' Disease
RERE Hypothyroidism
−log10 P
Chromosome 1 (Mb)
8.3
8.4
8.5
8.6
8.7
8.8
8.9
RERE−AS1→
RPL7P11→
←ENO1
←ENSG00000221083
←RPL7P7
←RNU6−991P
ENSG00000270282→
ENSG0000303676→
SLC45A1→
←Y_RNA
Y_RNA→
ENSG00000226545→
←RERE
←ENSG00000228423

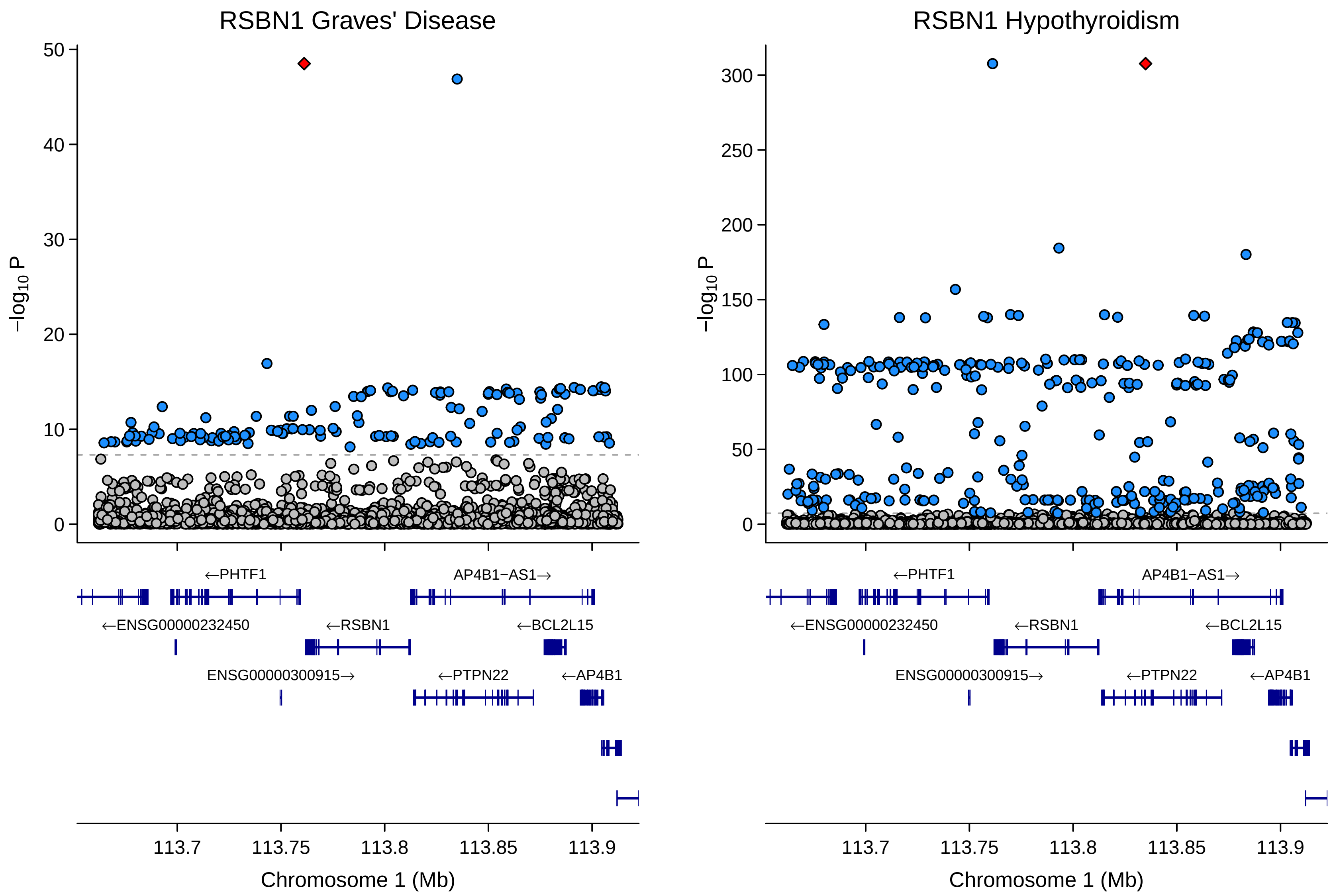
RSBN1 Graves' Disease
RSBN1 Hypothyroidism
−log10 P
0
10
20
30
40
50
0
50
100
150
200
250
300
←PHTF1
AP4B1−AS1→
←ENSG00000232450
←RSBN1
←BCL2L15
ENSG00000300915→
←PTPN22
←AP4B1
113.7
113.75
113.8
113.85
113.9
Chromosome 1 (Mb)

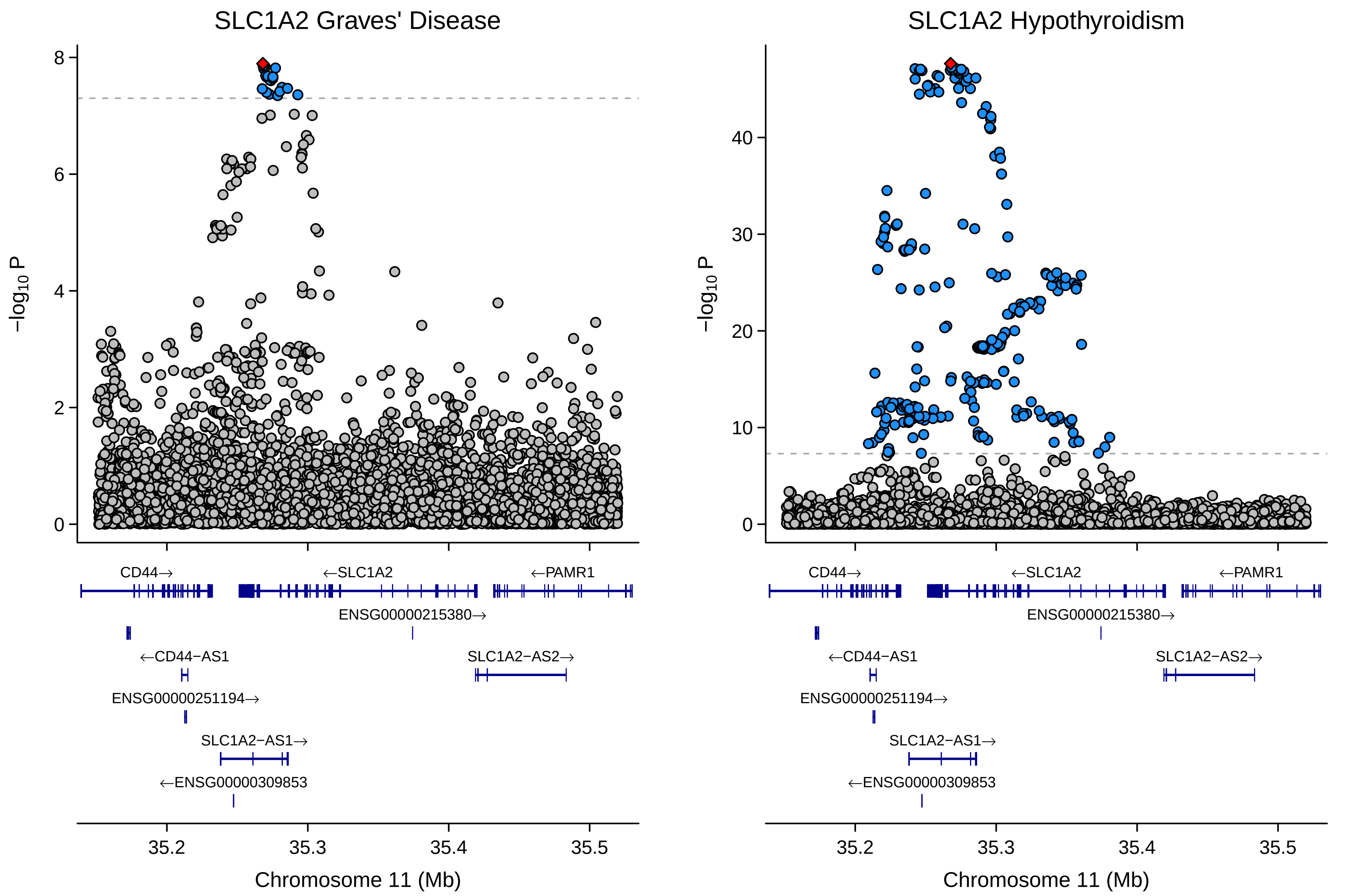

SLC1A2 Graves' Disease
SLC1A2 Hypothyroidism
−log10 P
Chromosome 11 (Mb)
35.2
35.3
35.4
35.5
CD44→
←SLC1A2
←PAMR1
ENSG00000215380→
←CD44−AS1
SLC1A2−AS2→
ENSG00000251194→
SLC1A2−AS1→
←ENSG00000309853

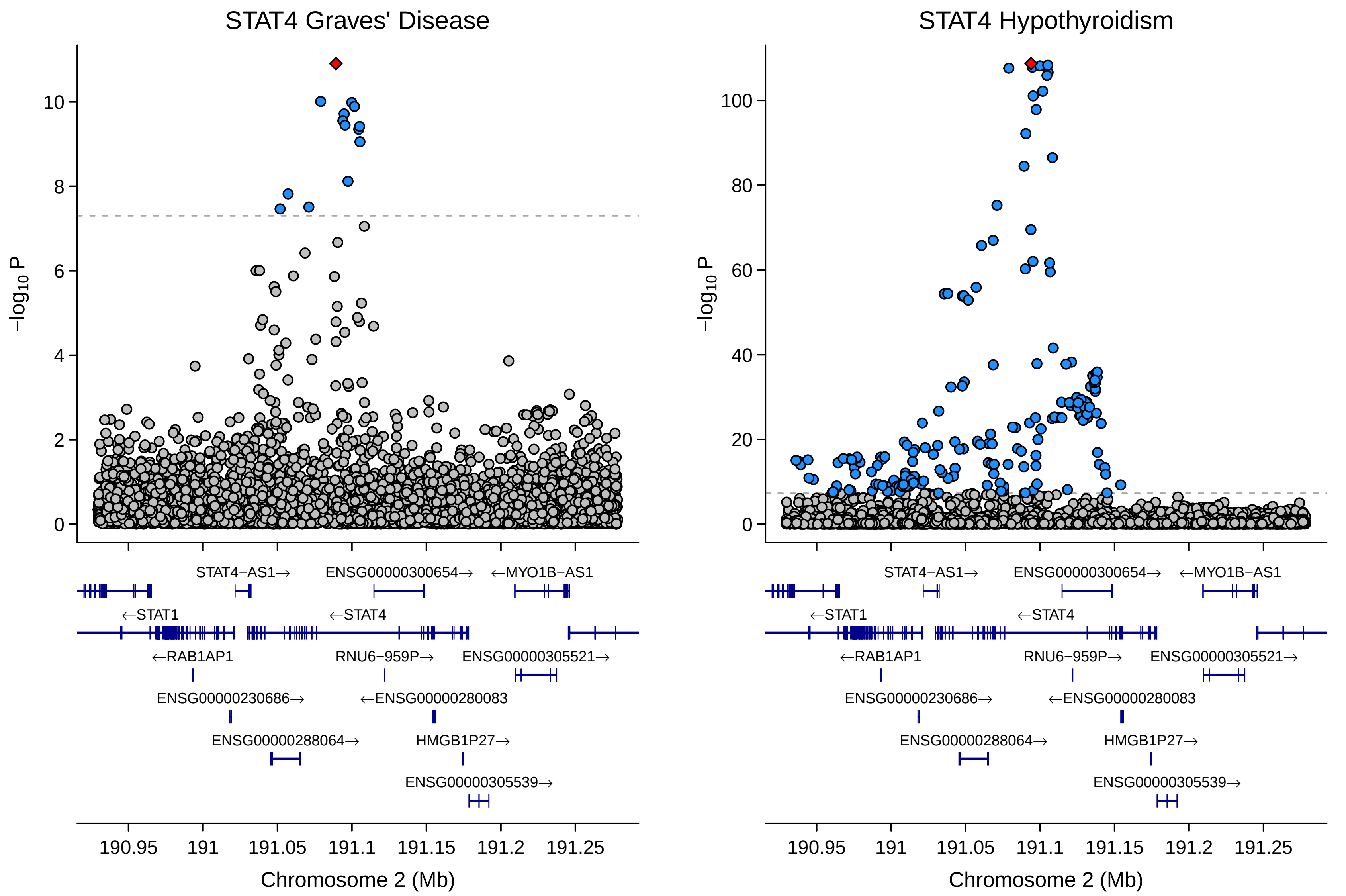
STAT4 Graves' Disease
STAT4 Hypothyroidism
−log10 P
0
2
4
6
8
10
20
40
60
80
100
STAT4−AS1→
ENSG00000300654→
←MYO1B−AS1
←STAT1
←STAT4
←RAB1AP1
RNU6−959P→
ENSG00000305521→
ENSG00000230686→
←ENSG00000280083
ENSG00000288064→
HMGB1P27→
ENSG00000305539→
190.95
191
191.05
191.1
191.15
191.2
191.25
Chromosome 2 (Mb)

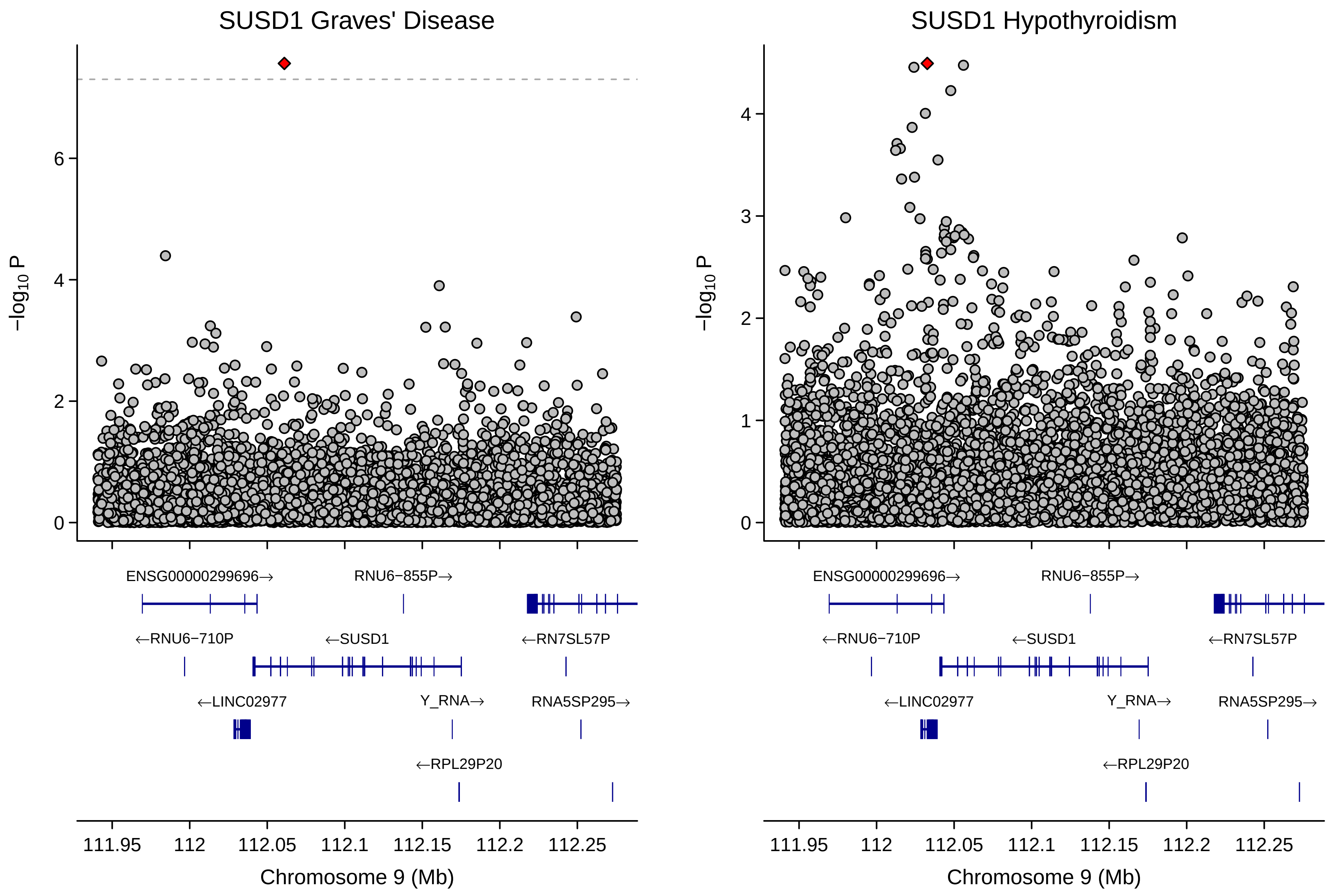
SUSD1 Graves' Disease
SUSD1 Hypothyroidism
−log10 P
ENSG00000299696→
RNU6−855P→
←RNU6−710P
←SUSD1
←RN7SL57P
←LINC02977
Y_RNA→
RNA5SP295→
←RPL29P20
111.95
112
112.05
112.1
112.15
112.2
112.25
Chromosome 9 (Mb)

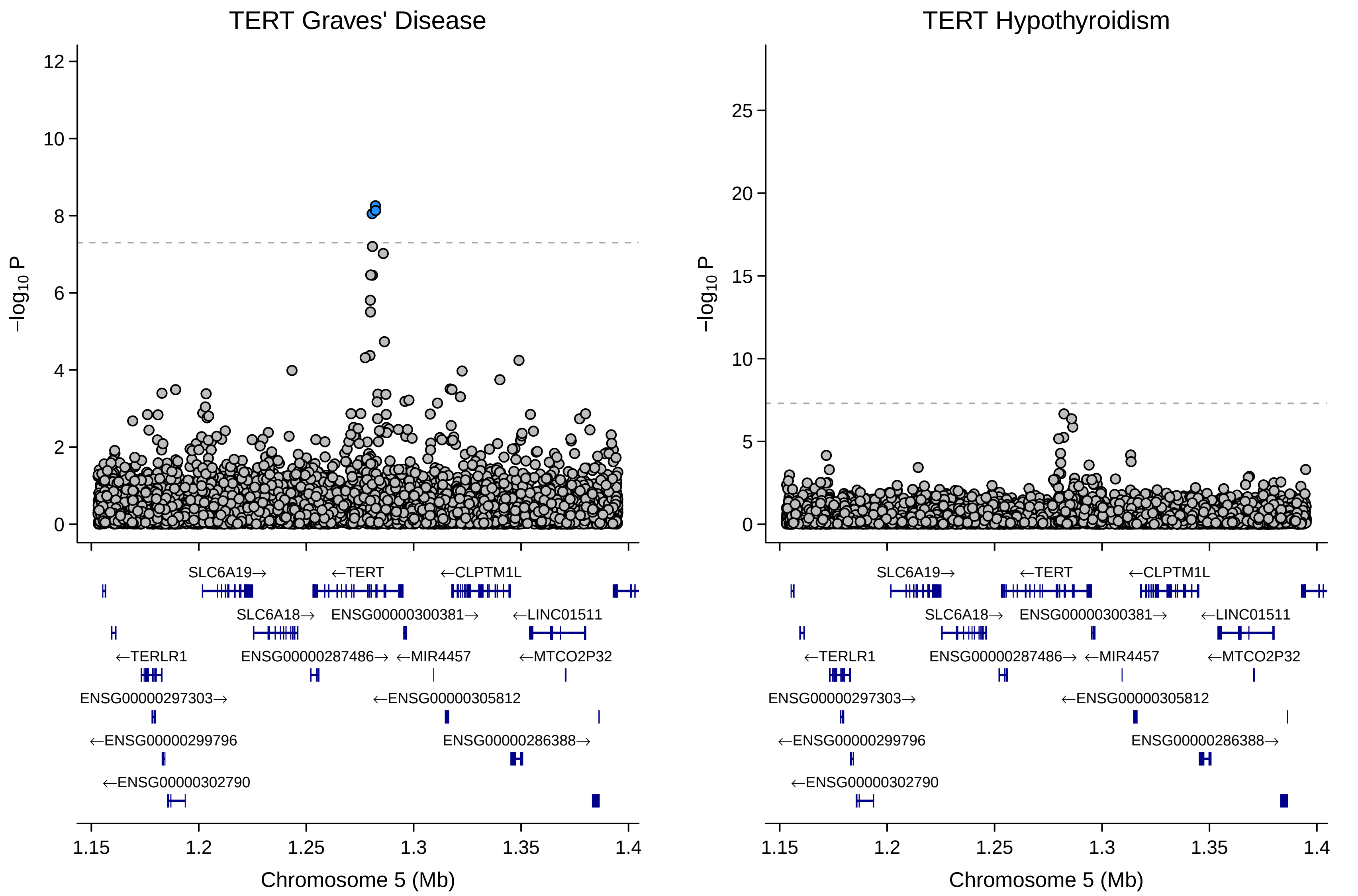

TERT Graves' Disease
TERT Hypothyroidism
−log10 P
Chromosome 5 (Mb)
SLC6A19→
←TERT
←CLPTM1L
SLC6A18→
ENSG00000300381→
←LINC01511
←TERLR1
ENSG00000287486→
←MIR4457
←MTCO2P32
ENSG00000297303→
←ENSG00000305812
←ENSG00000299796
ENSG00000286388→
←ENSG00000302790

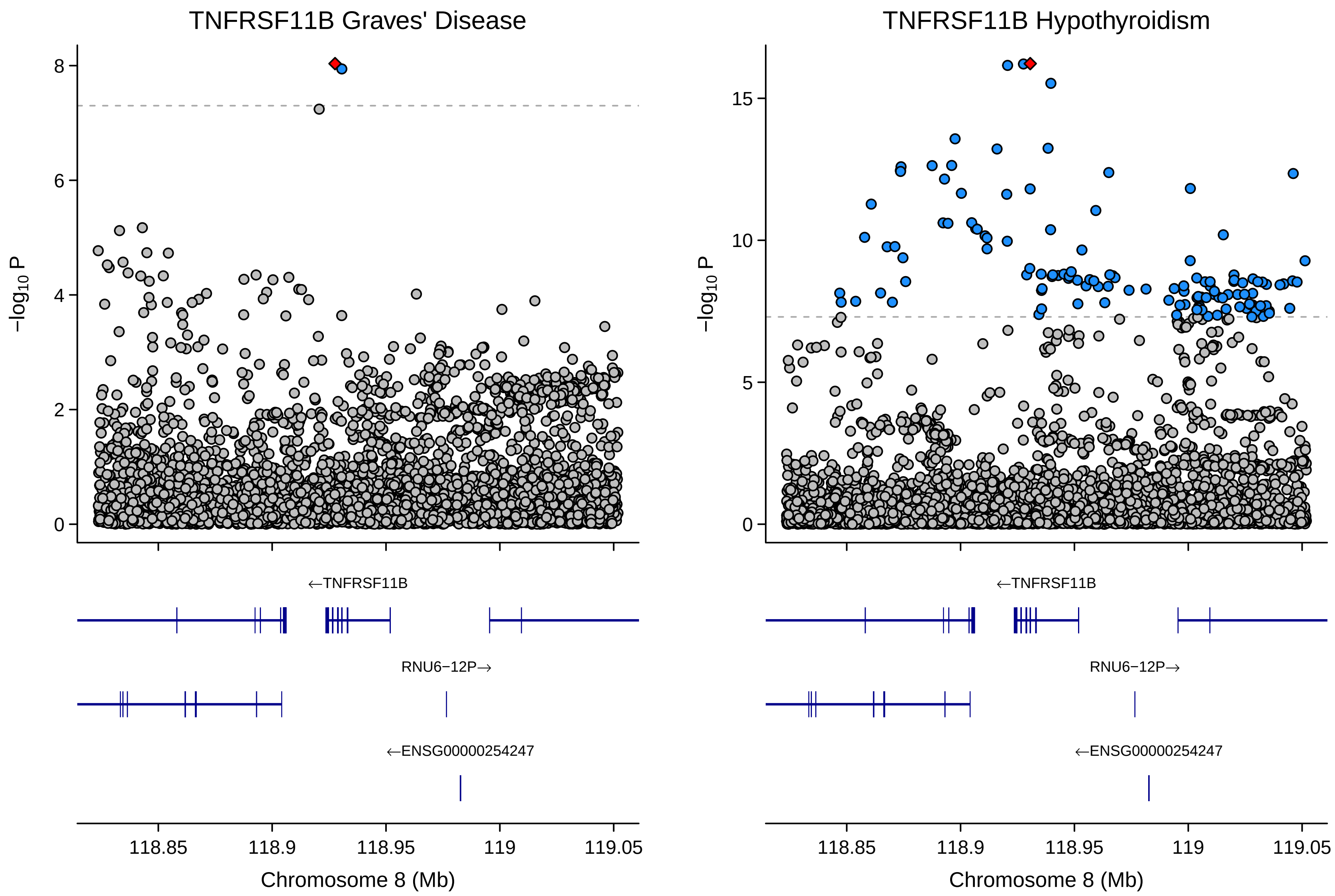
TNFRSF11B Graves' Disease
TNFRSF11B Hypothyroidism
−log10 P
8
6
4
2
0
15
10
5
0
←TNFRSF11B
RNU6−12P→
←ENSG00000254247
118.85
118.9
118.95
119
119.05
Chromosome 8 (Mb)

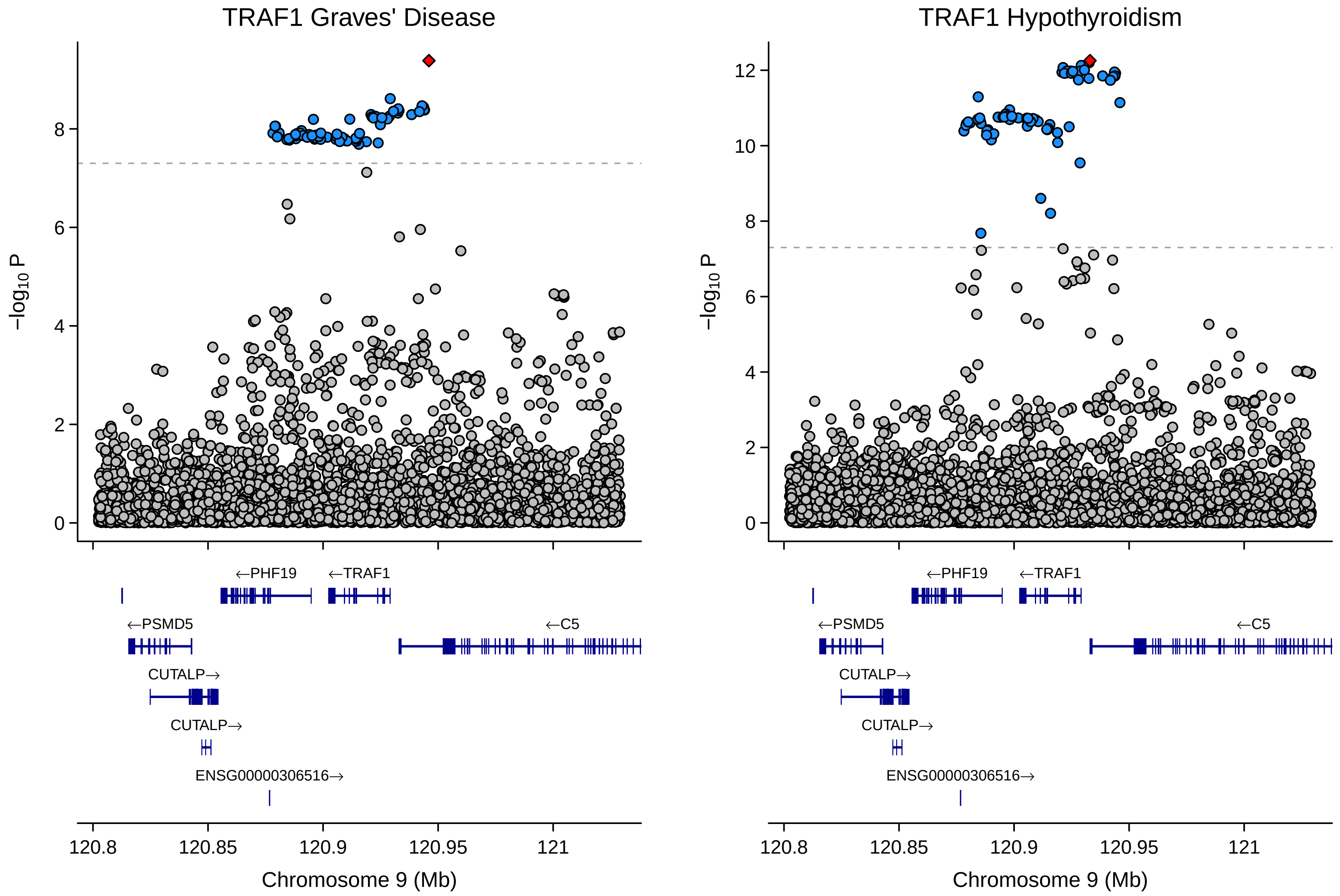

TRAF1 Graves' Disease
TRAF1 Hypothyroidism
−log10 P
0
2
4
6
8
10
12
←PHF19
←TRAF1
←PSMD5
←C5
CUTALP→
CUTALP→
ENSG00000306516→
120.8
120.85
120.9
120.95
121
Chromosome 9 (Mb)

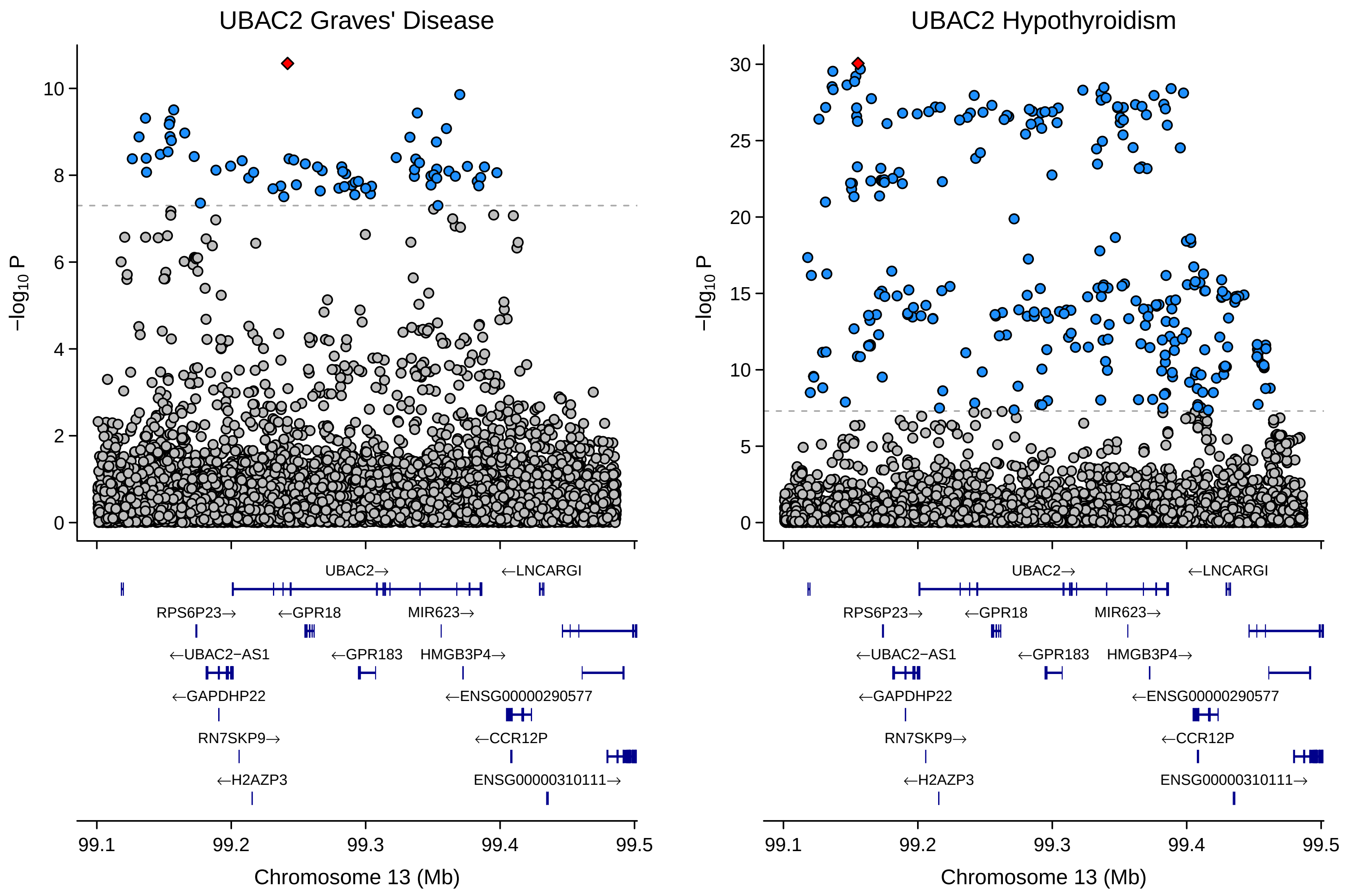

UBAC2 Graves' Disease
UBAC2 Hypothyroidism
−log10 P
0
2
4
6
8
10
0
5
10
15
20
25
30
UBAC2→
←LNCARGI
RPS6P23→
←GPR18
MIR623→
←UBAC2−AS1
←GPR183
HMGB3P4→
←GAPDHP22
←ENSG00000290577
RN7SKP9→
←CCR12P
←H2AZP3
ENSG00000310111→
99.1
99.2
99.3
99.4
99.5
Chromosome 13 (Mb)

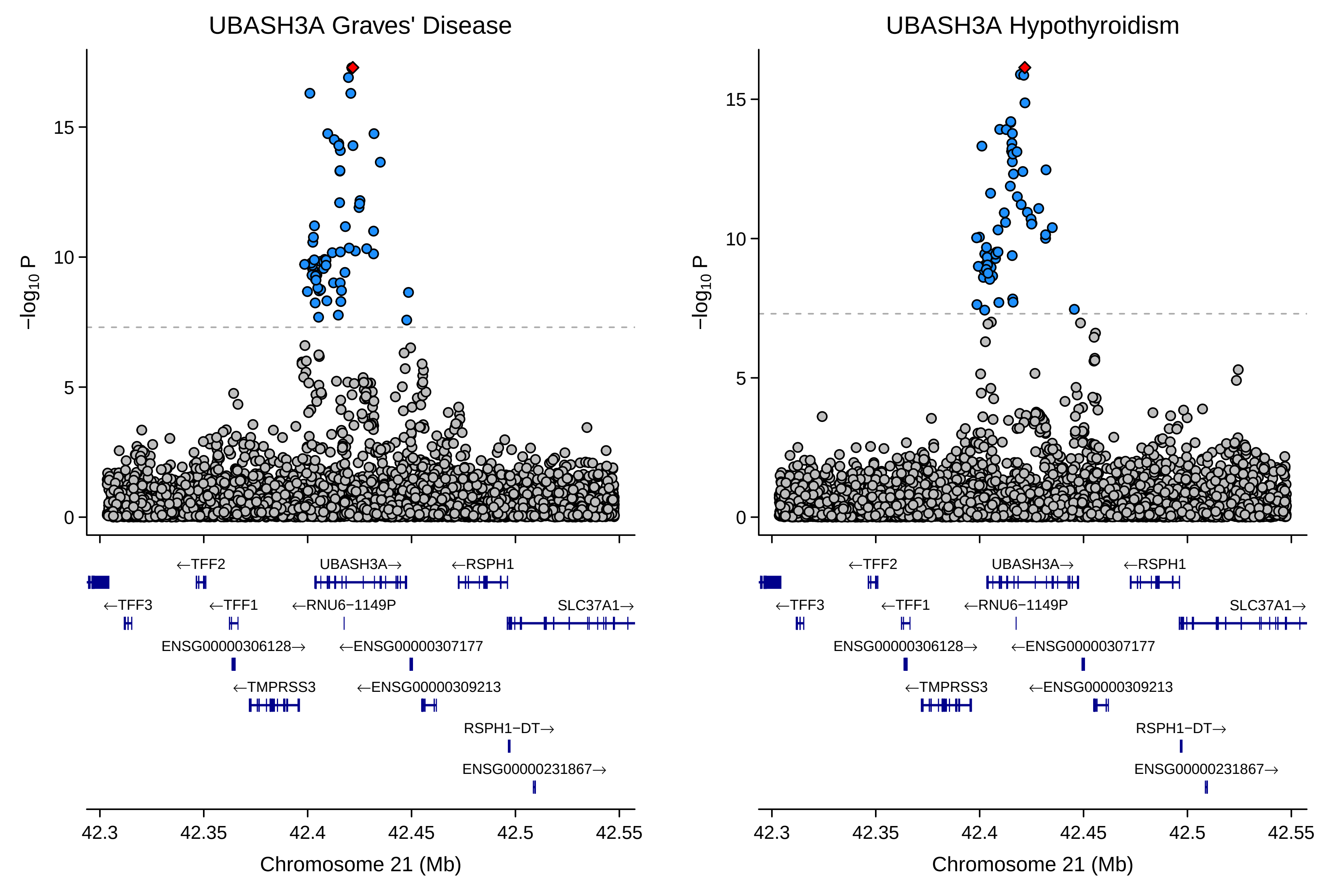
UBASH3A Graves' Disease
UBASH3A Hypothyroidism
−log10 P
0
5
10
15
Chromosome 21 (Mb)
42.3
42.35
42.4
42.45
42.5
42.55
←TFF2
UBASH3A→
←RSPH1
←TFF3
←TFF1
←RNU6−1149P
SLC37A1→
ENSG00000306128→
←ENSG00000307177
←TMPRSS3
←ENSG00000309213
RSPH1−DT→
ENSG00000231867→

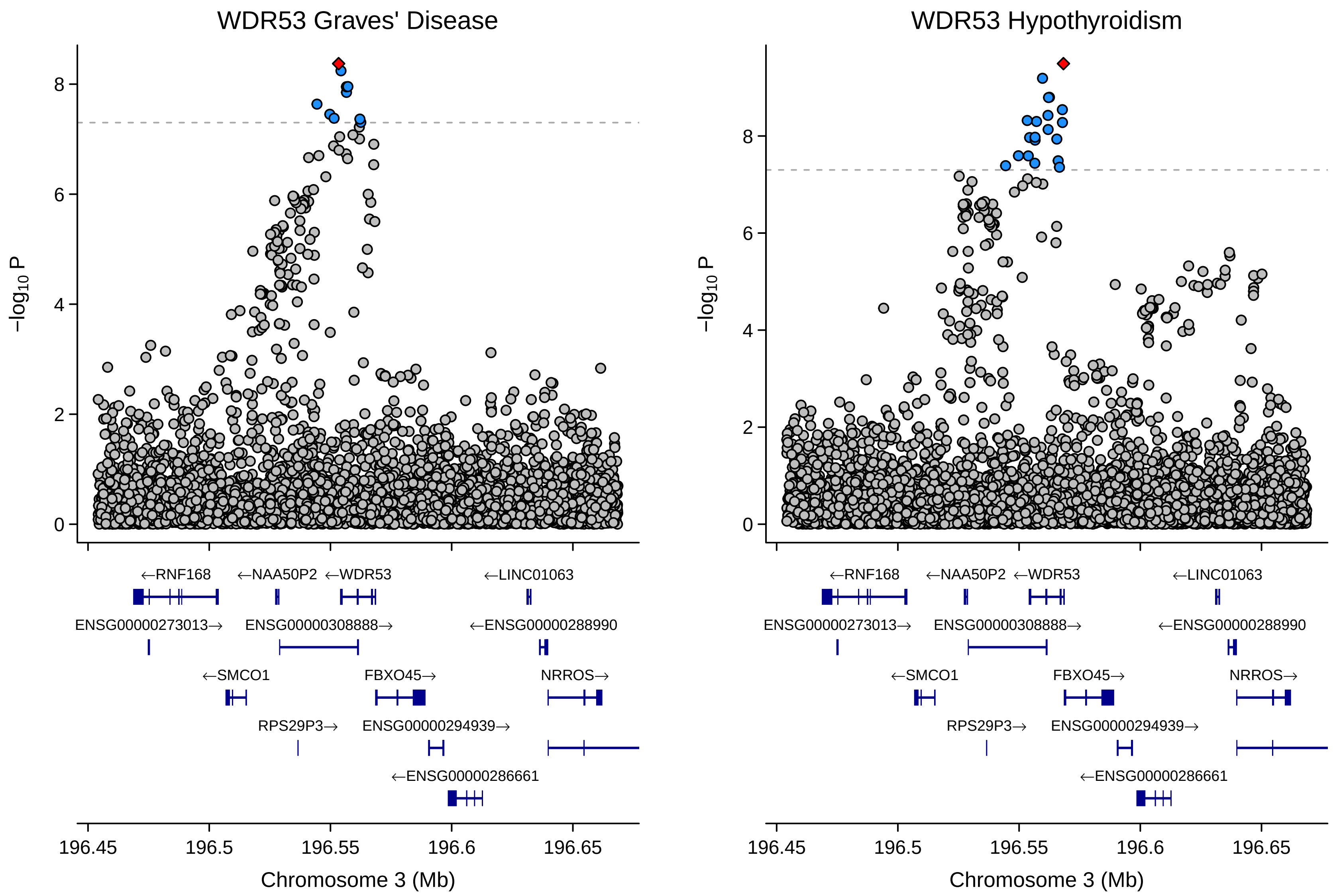

WDR53 Graves' Disease
WDR53 Hypothyroidism
−log10 P
Chromosome 3 (Mb)
196.45
196.5
196.55
196.6
196.65
←RNF168
←NAA50P2
←WDR53
←LINC01063
ENSG00000273013→
ENSG00000308888→
←ENSG00000288990
←SMCO1
FBXO45→
NRROS→
RPS29P3→
ENSG00000294939→
←ENSG00000286661

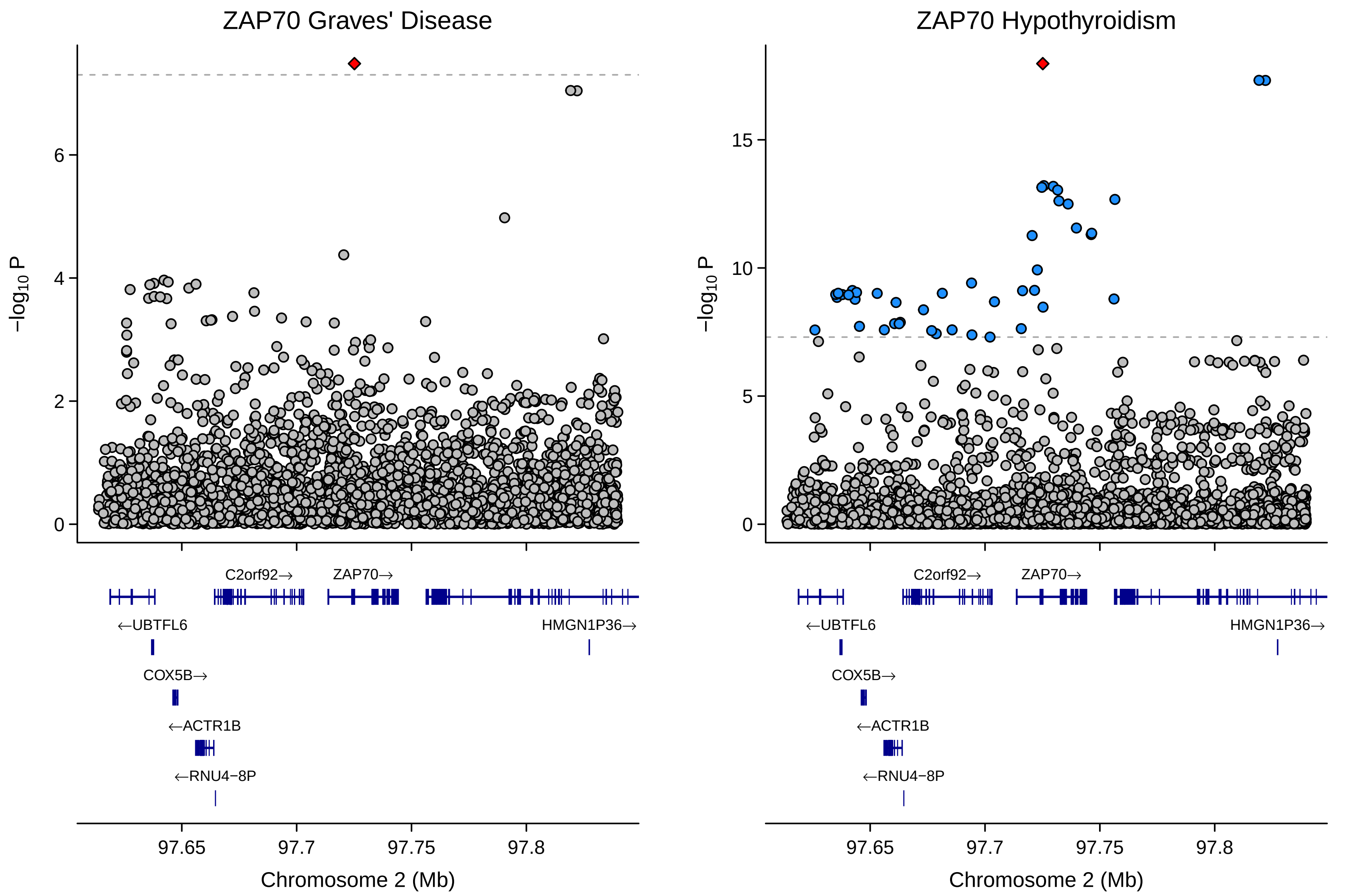
ZAP70 Graves' Disease
ZAP70 Hypothyroidism
−log10 P
0
2
4
6
5
10
15
C2orf92→
ZAP70→
←UBTFL6
HMGN1P36→
COX5B→
←ACTR1B
←RNU4−8P
97.65
97.7
97.75
97.8
Chromosome 2 (Mb)
