## Supplementary figures and images for "Global multi-ancestry genetic study elucidates genes and biological pathways associated with thyroid cancer and benign thyroid diseases"

### Supplementary Figure 2.2

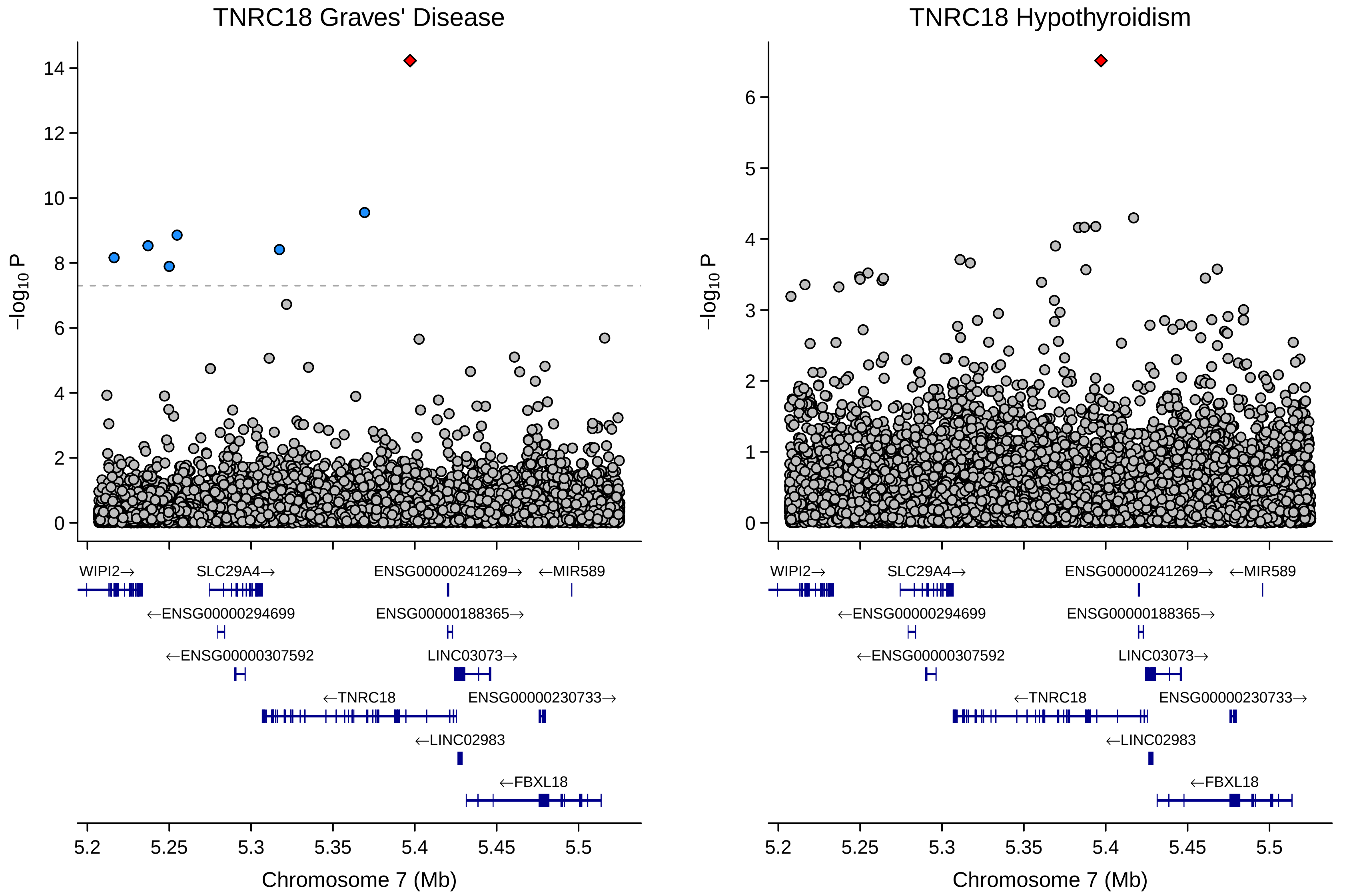

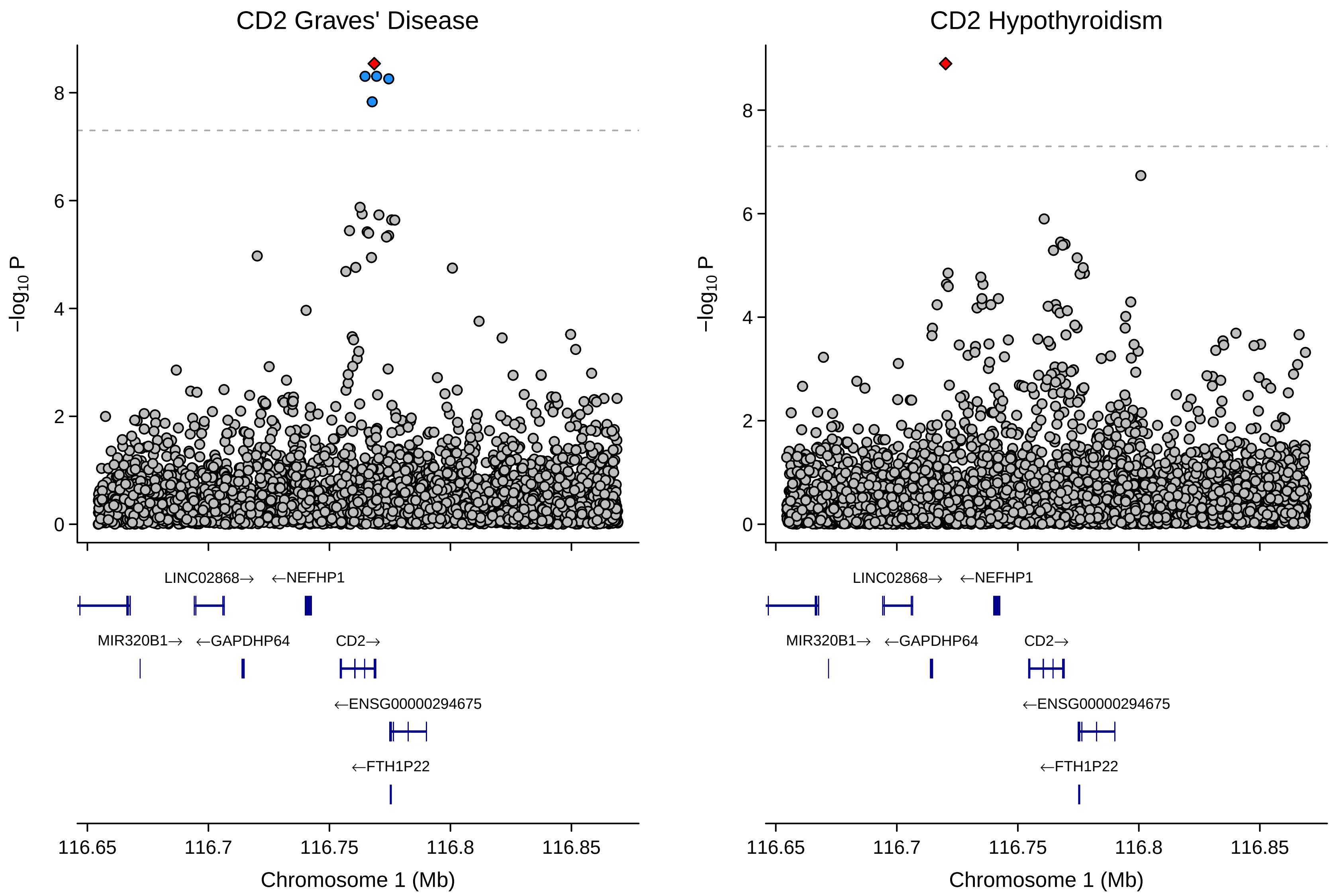

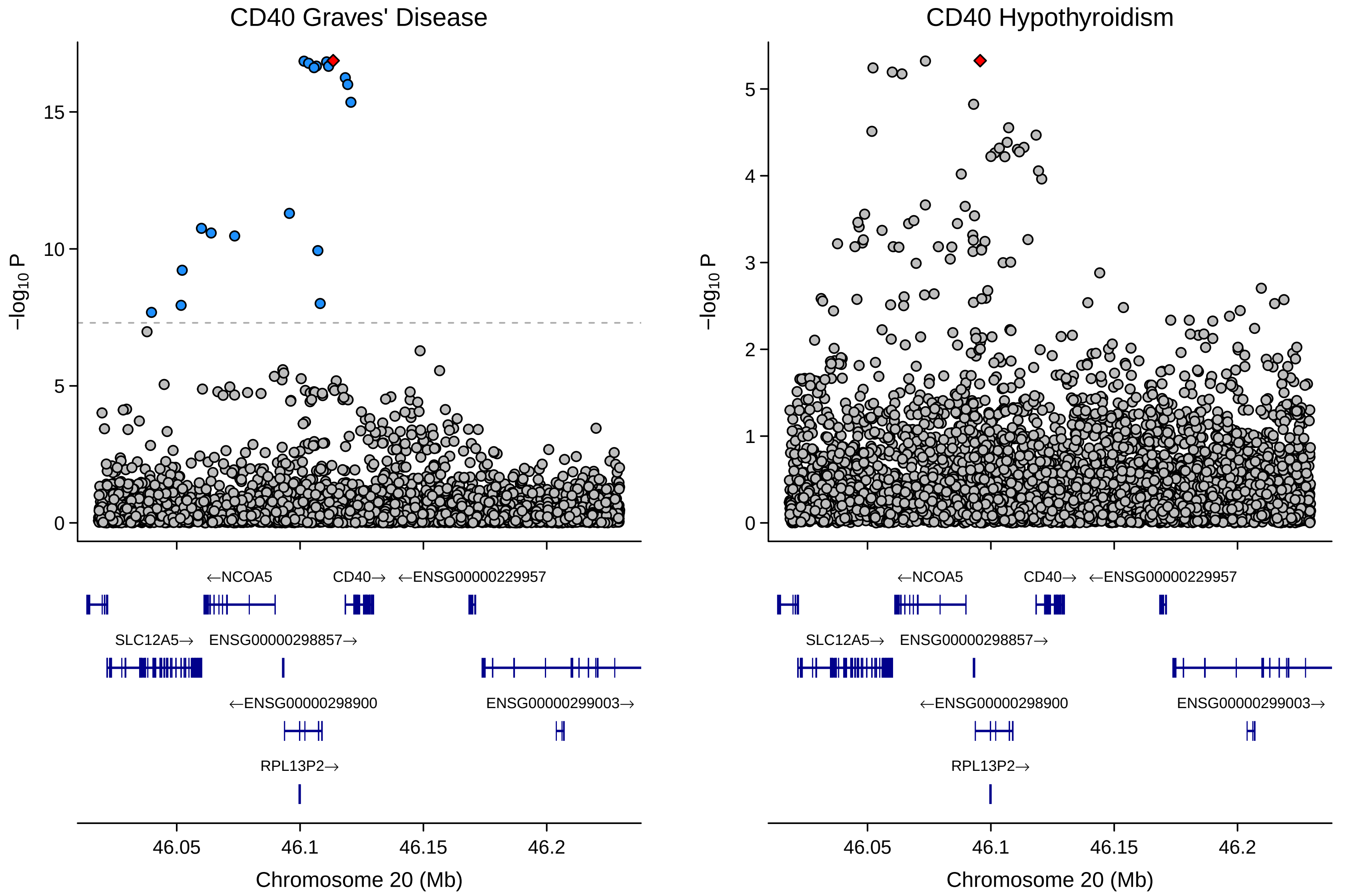

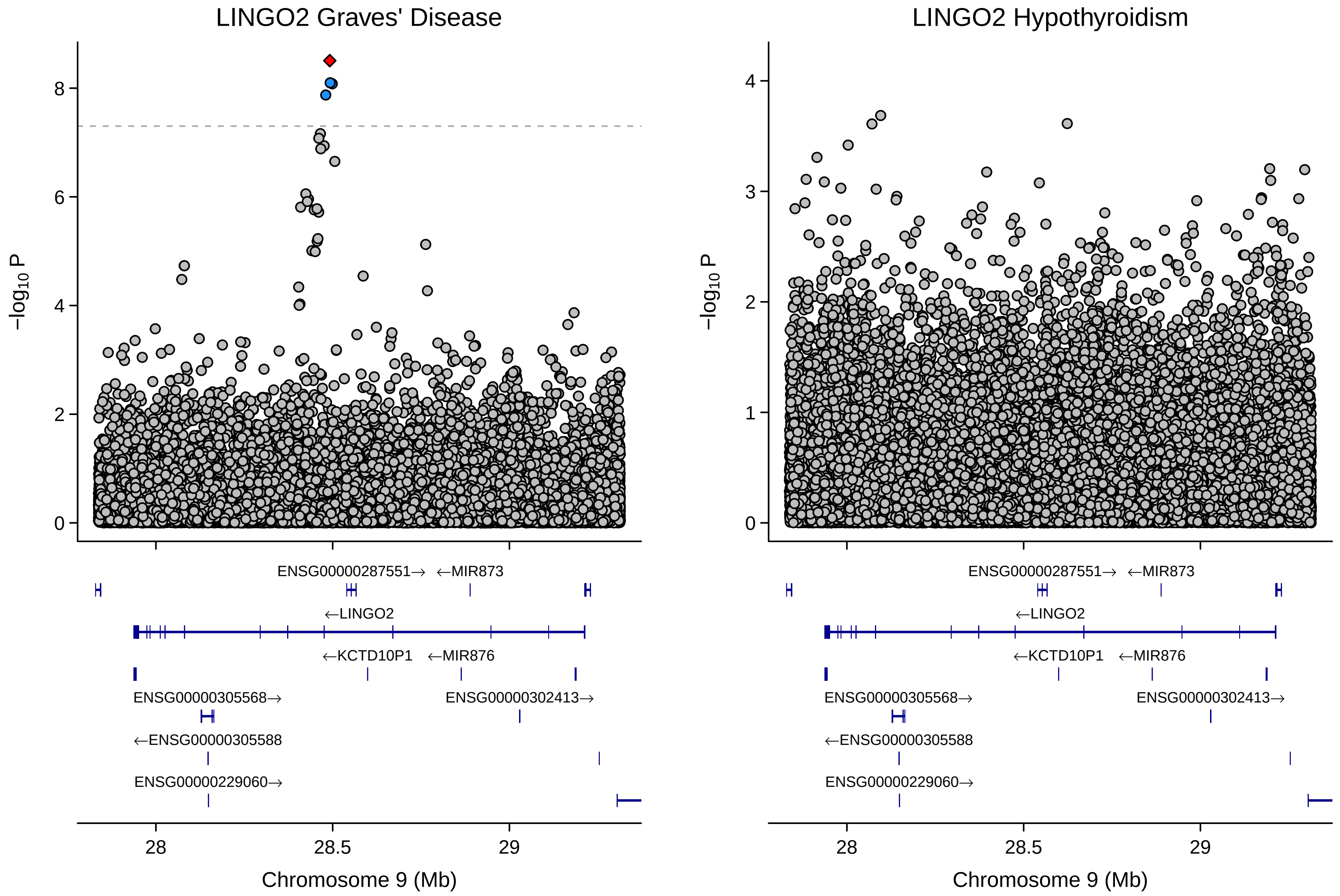
