## Supplementary material for "Global multi-ancestry genetic study elucidates genes and biological pathways associated with thyroid cancer and benign thyroid diseases": Banner Authorships

**Anderson, Heather D.**
Department of Clinical Pharmacy, University of Colorado Skaggs School of Pharmacy and Pharmaceutical Sciences, Anschutz Medical Campus

**Aquilante, Christina L.**
Department of Pharmaceutical Sciences, University of Colorado Skaggs School of Pharmacy and Pharmaceutical Sciences, Anschutz Medical Campus

**Arbogast, Kelsey**

**Arehart, Christopher H.**

**Brooks, Ian M.**
Department of Biomedical Informatics, University of Colorado School of Medicine, Anschutz Medical Campus; Health Data Compass, Office of the Vice Chancellor for Health Affairs, Anschutz Medical Campus

**Brunetti, Tonya M.**

**Brutus-Lestin, Judith**
UCHealth Core Lab

**Burke, Elizabeth E.**
CARES Innovation Center, UCHealth, Anschutz Medical Campus

**Casteel, Emily M.**
None

**Cole, Joanne B**
Department of Biomedical Informatics, University of Colorado School of Medicine, Anschutz Medical Campus

**Coughlin II, Curtis R.**
Department of Pediatrics, University of Colorado School of Medicine, Anschutz Medical Campus

**Crooks, Kristy**
Department of Pathology, University of Colorado School of Medicine, Anschutz Medical Campus

**Crawford, Jacob**

**Culver, Erin**

**Edelmann, Michelle N.**
Health Data Compass, Office of the Vice Chancellor for Health Affairs, Anschutz Medical Campus

**Fisher, Matthew J.**

**Franklin, Alan W.**

**Frye, Teresa C.**

**George, Hunter**

**Gignoux, Chris R.**
Department of Biomedical Informatics, University of Colorado School of Medicine, Anschutz Medical Campus

**Gilliland, Elizabeth K.**

**Greene, Casey S.**
Department of Biomedical Informatics, University of Colorado School of Medicine, Anschutz Medical Campus

**Hawkes, Brooke**

**Hearst, Emily**
CARES Innovation Center, UCHealth, Anschutz Medical Campus

**Hendricks, Audrey E.**
Department of Biomedical Informatics, University of Colorado School of Medicine, Anschutz Medical Campus; Department of Mathematical and Statistical Sciences, College of Arts and

Sciences, University of Colorado Denver Campus

**Johnson, Randi K.**
Department of Biomedical Informatics, University of Colorado School of Medicine, Anschutz Medical Campus; Department of Epidemiology, Colorado School of Public Health, Anschutz Medical Campus

**Julian, Colleen G.**
Department of Biomedical Informatics, University of Colorado School of Medicine, Anschutz Medical Campus

**Kao, Dave**
Division of Cardiology, Department of Medicine, University of Colorado School of Medicine, Anschutz Medical Campus; CARE Innovation Center, UCHealth, Anschutz Medical Campus

**Konigsberg, Iain**
Department of Biomedical Informatics, University of Colorado School of Medicine, Anschutz Medical Campus

**Ku, Lisa**
Hereditary Cancer Clinic, UCHealth, Anschutz Medical Campus

**Kudron, Elizabeth L.**
Department of Biomedical Informatics, University of Colorado School of Medicine, Anschutz Medical Campus; Department of Pediatrics, University of Colorado School of Medicine, Anschutz Medical Campus

**Lacy, Rashawnda**
Health Data Compass, Office of the Vice Chancellor for Health Affairs, Anschutz Medical Campus

**Lange, Ethan M.**
Department of Biomedical Informatics, University of Colorado School of Medicine, Anschutz Medical Campus

**Lee, Yee Ming**
Department of Clinical Pharmacy, University of Colorado Skaggs School of Pharmacy and Pharmaceutical Sciences, Anschutz Medical Campus

**Lesny, Joe A.**

**Lin, Meng**
Department of Biomedical Informatics, University of Colorado School of Medicine, Anschutz Medical Campus

**Lowery, Jan T.**

**Vargas, Luciana B.**
Department of Biomedical Informatics, University of Colorado School of Medicine, Anschutz Medical Campus

**Maldonado, Betzaida L.**
Department of Biomedical Informatics, University of Colorado School of Medicine, Anschutz Medical Campus

**Marceau, Darcy**

**Martin, James L.**

**Gates, Brianna L.**

**Mayer, David**
Department of Biomedical Informatics, University of Colorado School of Medicine, Anschutz Medical Campus

**McDaniel, Nicole L.**
Department of Clinical Pharmacy, University of Colorado Skaggs School of Pharmacy and Pharmaceutical Sciences, Anschutz Medical Campus

**Monte, Andrew**
University of Colorado School of Medicine; University of Colorado School of Pharmacy & Pharmaceutical Sciences; Rocky Mountain Poison & Drug Safety, Denver Health

**Moore, Ethan**

**Nadrash, Ann**

Ambulatory Pharmacy Health Outcomes Pharmacy Department, UCHealth, Anschutz Medical Campus

**Pattee, Jack**
Department of Biostatistics and Informatics, Center for Innovative Design and Analysis, Anschutz Medical Campus

**Pozdeyev, Nikita**
Department of Biomedical Informatics, University of Colorado School of Medicine, Anschutz Medical Campus; Division of Endocrinology, University of Colorado School of Medicine, Anschutz Medical Campus

**Radwan, Alaa**
Department of Clinical Pharmacy, University of Colorado Skaggs School of Pharmacy and Pharmaceutical Sciences, Anschutz Medical Campus

**Rafaels, Nick**

**Raghavan, Sridharan**

**Rasouli, Neda**
Division of Endocrinology, Department of Medicine, University of Colorado School of Medicine, Anschutz Medical Campus

**Shalowitz, Elise L.**

**Sherif, Hoda**

**Shortt, Johnathan A.**
Department of Biomedical Informatics, University of Colorado School of Medicine, Anschutz Medical Campus

**Stewart, Adrian M.**

**Sutton, Kristen J.**
Department of Biomedical Informatics, University of Colorado School of Medicine, Anschutz Medical Campus

**Swartz, Carolyn T.**
UCHealth Epic IT Department

**Tanaka, Anna**
CARES Innovation Center, UCHealth, Anschutz Medical Campus

**Taylor, Matthew R.G.**

**Teague, Candace**

**Todd, Emily B.**
Department of Biomedical Informatics, University of Colorado School of Medicine, Anschutz Medical Campus

**Trinkley, Katy E.**
University of Colorado Department of Family Medicine, University of Colorado School of Medicine, Anschutz Medical Campus

**Wiley, Laura K** Department of Biomedical Informatics, University of Colorado School of Medicine, Anschutz Medical Campus

### The BioBank Japan Project

Koichi Matsuda[1,2], Yuji Yamanashi[3], Yoichi Furukawa[4], Takayuki Morisaki[5], Yukinori Okada[6–10], Yoshinori Murakami[11], Yoichiro Kamatani[12], Kaori Muto[13], Akiko Nagai[2], Yusuke Nakamura[14], Wataru Obara[15], Ken Yamaji[16], Kazuhisa Takahashi[17], Satoshi Asai[18,19], Yasuo Takahashi[19], Shinichi Higashiue[20], Shuzo Kobayashi[20], Hiroki Yamaguchi[21], Yasunobu Nagata[21], Satoshi Wakita[21], Chikako Nito[22], Yu-ki Iwasaki[23], Shigeo Murayama[24], Kozo Yoshimori[25], Yoshio Miki[26], Daisuke Obata[27], Masahiko Higashiyama[28], Akihide Masumoto[29], Yoshinobu Koga[29], and Yukihiro Koretsune[30]

[1]Laboratory of Genome Technology, Human Genome Center, Institute of Medical Science, The University of Tokyo, Tokyo, Japan.

[2]Laboratory of Clinical Genome Sequencing, Graduate School of Frontier Sciences, The University of Tokyo, Tokyo, Japan.

[3]Division of Genetics, The Institute of Medical Science, The University of Tokyo, Tokyo, Japan.

[4]Division of Clinical Genome Research, Institute of Medical Science, The University of Tokyo, Tokyo, Japan.

[5]Department of Computational Biology and Medical Sciences, Graduate School of Frontier Sciences, BioBank Japan, Institute of Medical Science, The University of Tokyo, Tokyo, Japan.

[6]Department of Statistical Genetics, Osaka University Graduate School of Medicine, Suita, Japan.

[7]Laboratory for Systems Genetics, RIKEN Center for Integrative Medical Sciences, Yokohama, Japan.

[8]Department of Genome Informatics, Graduate School of Medicine, the University of Tokyo, Tokyo, Japan.

[9]Laboratory of Statistical Immunology, Immunology Frontier Research Center (WPI-IFReC), Osaka University, Suita, Japan.
[10]Premium Research Institute for Human Metaverse Medicine (WPI-PRIMe), Osaka University, Osaka, Japan.
[11]Department of Cancer Biology, Institute of Medical Science, The University of Tokyo, Tokyo, Japan.
[12]Laboratory of Complex Trait Genomics, Graduate School of Frontier Sciences, The University of Tokyo, Tokyo, Japan.
[13]Department of Public Policy, Institute of Medical Science, The University of Tokyo, Tokyo, Japan.
[14]The Institute of Medical Science, The University of Tokyo, Tokyo, Japan.
[15]Department of Urology, Iwate Medical University, Iwate, Japan.
[16]Department of Internal Medicine and Rheumatology, Juntendo University Graduate School of Medicine, Tokyo, Japan.
[17]Department of Respiratory Medicine, Juntendo University Graduate School of Medicine, Tokyo, Japan.
[18]Division of Pharmacology, Department of Biomedical Science, Nihon University School of Medicine, Tokyo, Japan.
[19]Division of Genomic Epidemiology and Clinical Trials, Clinical Trials Research Center, Nihon University. School of Medicine, Tokyo, Japan.
[20]Tokushukai Group, Tokyo, Japan.
[21]Departmentof Hematology, Nippon Medical School, Tokyo, Japan.
[22]Laboratory for Clinical Research, Collaborative Research Center, Nippon Medical School, Tokyo, Japan.
[23]Department of Cardiovascular Medicine, Nippon Medical School, Tokyo, Japan.
[24]Tokyo Metropolitan Geriatric Hospital and Institute of Gerontology, Tokyo, Japan.

[25]Fukujuji Hospital, Japan Anti-Tuberculosis Association, Tokyo, Japan.

[26]The Cancer Institute Hospital of the Japanese Foundation for Cancer Research, Tokyo, Japan.

[27]Center for Clinical Research and Advanced Medicine, Shiga University of Medical Science, Shiga, Japan.

[28]Department of General Thoracic Surgery, Osaka International Cancer Institute, Osaka, Japan.

[29]Iizuka Hospital, Fukuoka, Japan.

[30]National Hospital Organization Osaka National Hospital, Osaka, Japan.

**Genes & Health Research Team**

https://docs.google.com/spreadsheets/d/1D9HLbc_m0KdOUN-gS0hymTLewJ36e8ETSEb8Tu_CqWY/edit?gid=0#gid=0
